# The Stress-Pain Connection in Chronic Primary Pain: A Systematic Review and Meta-Analysis of Physiological Stress Markers in Relation to Experimental Pain Responses

**DOI:** 10.1101/2025.09.24.25336431

**Authors:** Joren Vyverman, Robrecht De Baere, Inge Timmers, Iris Coppieters, Jessica Van Oosterwijck, Matthijs Moerkerke

**Author notes:** Corresponding author: Jessica Van Oosterwijck, Spine, Head and Pain Research Unit Ghent, Department of Rehabilitation Sciences, Ghent University, Campus UZ Ghent, Corneel Heymanslaan 10, B3, 9000 Ghent, Belgium. These authors contributed equally to this work (Joren Vyverman, Robrecht De Baere). www.paininmotion.be.

## Abstract

Dysfunctioning of stress systems, i.e., the autonomic nervous system (ANS) and hypothalamic-pituitary-adrenal (HPA) axis, has been implicated in chronic pain. However, the exact interplay between (re)activity and recovery of stress and pain systems in chronic pain remains unclear. A systematic review and meta-analysis was pre-registered on PROSPERO (CRD42024495934). Six databases were searched to identify relevant literature. Risk of bias (RoB) was evaluated with the Newcastle-Ottawa Scale, and certainty of evidence (CoE) with GRADE. Clusters of interactions between physiological markers of stress and experimental outcomes of pain were formed based on the timing of the stress measurements. Fifty-two studies (5 cross-sectional, 47 case-control; *n* = 2,657) were included and scored on average 9/12 (range: 2-11) on RoB. Overall CoE was very low to moderate. Qualitative analyses showed significant correlations between lower mean arterial pressure and higher pain sensitivity at baseline in individuals with chronic primary pain, which was supported by a meta-analysis. Furthermore, meta-analyses showed that higher pain sensitivity was associated with higher cortisol levels at baseline, lower high-frequency heart rate variability during recovery, and higher heart rate at multiple timepoints of the stress system response. Other associations did not yield significance. Taken together, these findings suggest that dysfunction of the ANS and HPA axis is linked to heightened pain sensitivity in chronic primary pain populations. However, the level of evidence remains low due to methodological heterogeneity, highlighting the need for studies combining stress markers and pain measures to provide insights into underlying stress-pain mechanisms.

**HIGHLIGHTS:**

– Sympathetic cardiovascular dominance appears to be associated with enhanced pain sensitivity in chronic primary pain
– A dysregulation in baroreflex activity might be present in chronic primary pain
– HPA axis dysfunctioning seems to be related to enhanced pain sensitivity in chronic primary pain
– Need for more standardized and comprehensive mapping of stress-pain interactions to unravel underlying mechanisms

## 1. INTRODUCTION

Stress and pain are connected through a complex and bidirectional interaction that involves several underlying mechanisms (e.g., Abdallah & Geha, 2017; Vachon-Presseau, 2018). This interaction has already been demonstrated in pain-free individuals: on the one hand, acute stress can both increase and decrease sensitivity to experimentally induced pain (Crettaz, et al., 2013; Gera, et al., 2025; Loffler, et al., 2023; Timmers, et al., 2018), while on the other hand, pain itself can also be a potent stressor, as demonstrated by the use of experimental painful stimuli to experimentally induce stress (Lovallo, 1975; Smeets, et al., 2012). Although the exact mechanisms remain unclear, the stress-pain relationship appears even more complex in individuals with chronic primary pain (CPP), who experience pain lasting over three months which is accompanied by significant emotional distress or disability and cannot be explained better by another medical condition (Nicholas, et al., 2019). Since individuals with CPP often exhibit different responses to both experimental pain and stress inductions then pain-free individuals evidencing dysregulated stress functioning and pain processing, gaining a deeper understanding of this relationship is especially relevant for this population.

Standardized noxious stimuli can be experimentally administered to induce pain in order to evaluate the pain system. These procedures, also known as quantitative sensory testing (QST), can be categorized as static QST, which is used to evaluate pain sensitivity by determination of pain thresholds and tolerances, and dynamic QST, which is used to evaluate the functionality of the pain system by assessment of pain modulatory mechanisms. Examples of dynamic QST include conditioned pain modulation (CPM), where one pain stimulus reduces the perception of another; or temporal summation of pain (TSP), which is the progressive increase in pain perception with repeated equal intensity stimuli over time. Individuals with CPP often exhibit heightened pain sensitivity (Amiri, et al., 2021) and dysfunctional pain modulation (Marcuzzi, et al., 2019; O’Brien, et al., 2018).

Besides altered pain processing, individuals with CPP also show altered stress responses and functionality of the stress system, consisting of the autonomic nervous system (ANS) and hypothalamic-pituitary-adrenal (HPA) axis. Typically, increased activity in the sympathetic branch of the ANS in response to a stressor produces a fast stress response by releasing catecholamines (Cardinali, 2018). This results in responses such as an increase in heart rate (HR), blood pressure (BP) and skin conductance (SC), to prepare the body to attend to the stressor. Conversely, activation of the parasympathetic branch of the ANS is characterized by a decrease in these parameters and an increase in heart rate variability (HRV), which reflects the adaptive autonomic responding and self-regulatory capacity of the ANS (Shaffer & Ginsberg, 2017). HRV seems to be reduced in populations experiencing chronic musculoskeletal pain (Rampazo, et al., 2024), and under acute stress, decreased responses and delayed parasympathetic recovery have been observed in CPP conditions (Saka-Kochi, et al., 2024; Ying-Chih, et al., 2020). In addition to the ANS responses, the HPA axis provides a slower and longer lasting response to a stressor through the release of cortisol, a well-known stress hormone (Herman, et al., 2016). However, when stress is prolonged and becomes chronic, the HPA axis can become dysregulated. In CPP, both decreased basal cortisol levels (Riva, et al., 2012; Riva, et al., 2010) as well as increased basal cortisol levels have been described (de Paiva Tosato, et al., 2015; H. I. Kim, et al., 2012; Riva, et al., 2012). Beyond basal activity, stress reactivity patterns also appear inconsistent, with some studies reporting elevated cortisol responses to experimental stressors (Yoshihara, et al., 2005), while others find blunted responses (Coppens, et al., 2018). These divergent findings point to the need for more comprehensive analyses of the full temporal profile of the stress response, rather than isolated time points. Differences in pain extent (localized versus widespread) may contribute to these discrepancies, indicating distinct stress-related mechanisms across subgroups, although this hypothesis remains to be tested (Hannibal & Bishop, 2014; Woda, et al., 2016). It must also be noted that some studies observed the involvement of the ANS in the stress-pain relationship but not the HPA axis (al’Absi & Petersen, 2003), and others found the opposite pattern (Timmers, et al., 2018). Overall, both stress systems appear to be dysregulated in CPP, but comprehensive assessment of their interaction with pain remains limited (Hannibal & Bishop, 2014; Woda, et al., 2016).

From a clinical perspective, stress system dysregulation in CPP is highly relevant, as it contributes not only to pain persistence but also to fatigue, mood disturbances, and sleep problems (Wyns, et al., 2023). Addressing these mechanisms may support more personalized management strategies, for example by integrating stress-regulation techniques with pain therapies, ultimately improving both pain outcomes and overall functioning (Hannibal & Bishop, 2014; Koenig, et al., 2016).

To summarize the evidence on the associations between physiological markers of stress response systems (E) and experimental pain outcomes (O) in CPP conditions (P), a meta-analytic review was performed providing a comprehensive overview of the literature and findings by distinguishing between basal activity, reactivity to a stressor and recovery from a stressor of the HPA axis and ANS, between static and dynamic experimental pain outcome measures, and different (pain) groups (i.e., chronic widespread, chronic localized and pain-free). This overview will provide insight into the mechanisms of the stress-pain relationship and how it has been assessed so far, as well as a critical lens through which to evaluate the employed methodology thus far.

## 2. METHODS

### 2.1 Protocol Registration

This systematic review and meta-analysis was conducted and reported following the Preferred Reporting Items for Systematic Reviews and Meta-analyses (PRISMA) guidelines as reflected in the PRISMA checklist (Supplementary File 1) (Liberati, et al., 2009; Page, et al., 2021). The protocol of the systematic review was registered prospectively on PROSPERO (Registration number: CRD42024495934).

### 2.2 Eligibility Criteria

The Population, Exposure, Comparator and Outcomes (PECO) framework (Morgan, et al., 2018) was used to formulate the following research question: “Is there an association between physiological markers of stress response systems (E) and experimentally induced pain assessments (O) in adults with chronic primary pain (P)? If so, how exactly do these parameters interact?”

Eligibility assessment of the retrieved studies was performed against predefined inclusion and exclusion criteria (Supplementary File 2 - Table S1). Studies performed on human adults (≥ 18 years old) with CPP (i.e., persistent pain lasting more than three months) (P) were included. CPP was defined according to the International Classification of Diseases 11 (ICD-11) for Mortality and Morbidity Statistics (https://icd.who.int/browse11; Nicholas, et al., 2019). Both case-control studies with healthy pain-free controls (C) and cross-sectional studies without controls were included. Studies reporting at least one marker of stress system (re)activity and/or recovery of either the ANS or HPA axis (E), and at least one experimental outcome measure of pain (O) were included. Studies that did not report either outcome were excluded. Only pre-treatment assessments were included from intervention studies.

### 2.3 Information Sources and Search Strategy

Searches were conducted across six electronic databases (MEDLINE (PubMed interface), Embase (Embase.com interface), Web of Science, CINAHL, Scopus, PsycARTICLES) from inception to August 21^th^, 2025. Based on the PECO question, free-text terms in combination with Boolean operators (“AND” and “OR”) were searched on title, abstract and keyword fields where applicable. Table S2 (Supplementary File 3 - Table S2) details the full search strategy for each database. Manual searches of reference lists of review papers and included publications, cited reference searches in Web of Science, and expert consultation supplemented the electronic search to ensure all relevant studies were included.

### 2.4 Selection Process

Following the removal of duplicates using EndNote (www.endnote.com), two reviewers (J.V., R.D.B.) independently screened the titles and abstracts of all identified records. Subsequently, full-text articles were assessed for eligibility using Rayyan (https://www.rayyan.ai/). Disagreements were resolved through discussion between the two reviewers, and if no consensus could be reached, the final decision was made by the last author (M.M.).

### 2.5 Data Items and Collection Process

Two reviewers independently extracted data using a pilot-tested table. Extracted data included study characteristics, participant details, stress and pain measures, and associations (e.g., correlations) between stress and pain. Any disagreements were resolved through discussion between the two reviewers, and if no agreement could be reached, the opinion of the last author (M.M.) was consulted. Authors were contacted to provide missing or additional data (e.g., correlation coefficients). Full data extraction procedures are detailed in Supplementary File 4.

### 2.6 Risk of Bias in Individual Studies

Two reviewers (J.V., R.D.B.) independently assessed the risk of bias using adapted versions of the Newcastle-Ottawa Scale (NOS) for case-control (Dhondt, et al., 2019) and cross-sectional studies (Herzog, et al., 2013). The NOS implements a “star system” to score risk of bias, and following categories were evaluated: selection of the study group, exposure, comparability, and outcome. Total scores range between zero and 12, with higher scores indicating lower risk of bias. The scores were compared in a meeting, and in case of discrepancies in scores, the article was screened again and discussed by both reviewers to resolve the differences. In case no agreement could be reached, the decision of the last author (M.M.) was decisive. Details of the scoring system and category interpretations are provided in Supplementary File 4 and Supplementary File 5 - Table S4.

### 2.7 Certainty of Evidence

The certainty of evidence was assessed using the Grading of Recommendations, Assessment, Development, and Evaluation criteria (GRADE) approach (Balshem, et al., 2011), starting at “low” due to inclusion of non-randomized studies. Downgrading (−1) was considered based on risk of bias, inconsistency, imprecision, and publication bias, while upgrading (+1) was possible for moderate to large effect sizes. The certainty of evidence could range between “very low” to “high”. Full criteria and decision rules are detailed in the Supplementary File 4.

### 2.8 Qualitative and Quantitative Analyses

Only studies reporting data on the association between stress and pain were included in the analyses. If the database was provided by the author upon request, correlation coefficients (*r*) were calculated using R 4.4.1 software in RStudio (R Core Team, 2024). The distribution of the data was checked to decide whether to use the Pearson or Spearman correlation method. Clusters were formed based on the timing of stress and pain measurements (i.e., assessed at baseline, during a stressor and after a stressor/during recovery of a stressor). Meta-analyses were conducted using random-effects models in R (version 4.4.1) when at least two studies reported correlations between similar stress and pain measures. Fisher’s z-transformation was applied to stabilize variances, and heterogeneity was assessed using the *I*² statistic. Where applicable, separate meta-analyses were conducted for the chronic widespread pain group and the chronic localized pain group. For more details on analysis procedures, heterogeneity handling, and publication bias assessment, see Supplementary File 4.

## 3. RESULTS

### 3.1 Study Selection

The database search resulted in a total of 9,740 studies. After deduplication, 6,739 records remained and were screened on title and abstract. A total of 159 studies were included for the screening on full text and were sought for retrieval. Finally, a total of 49 studies were included via the databases search, and three studies were additionally identified and included via other methods (*n* = 2 from the reference list of included studies; *n* = 1 from an expert in the field), resulting in a total of 52 studies. Fifteen studies did not report any associations between stress markers and pain and information could also not be retrieved from the authors. Of 46 contacted authors, 28 authors could not provide the requested additional data. Therefore, thirty-seven studies were included in the qualitative analysis, and 21 studies in the meta-analysis. A detailed overview of the selection process is presented in Figure 1 according to PRISMA flowchart guidelines (Page, et al., 2021).

**Figure 1.**
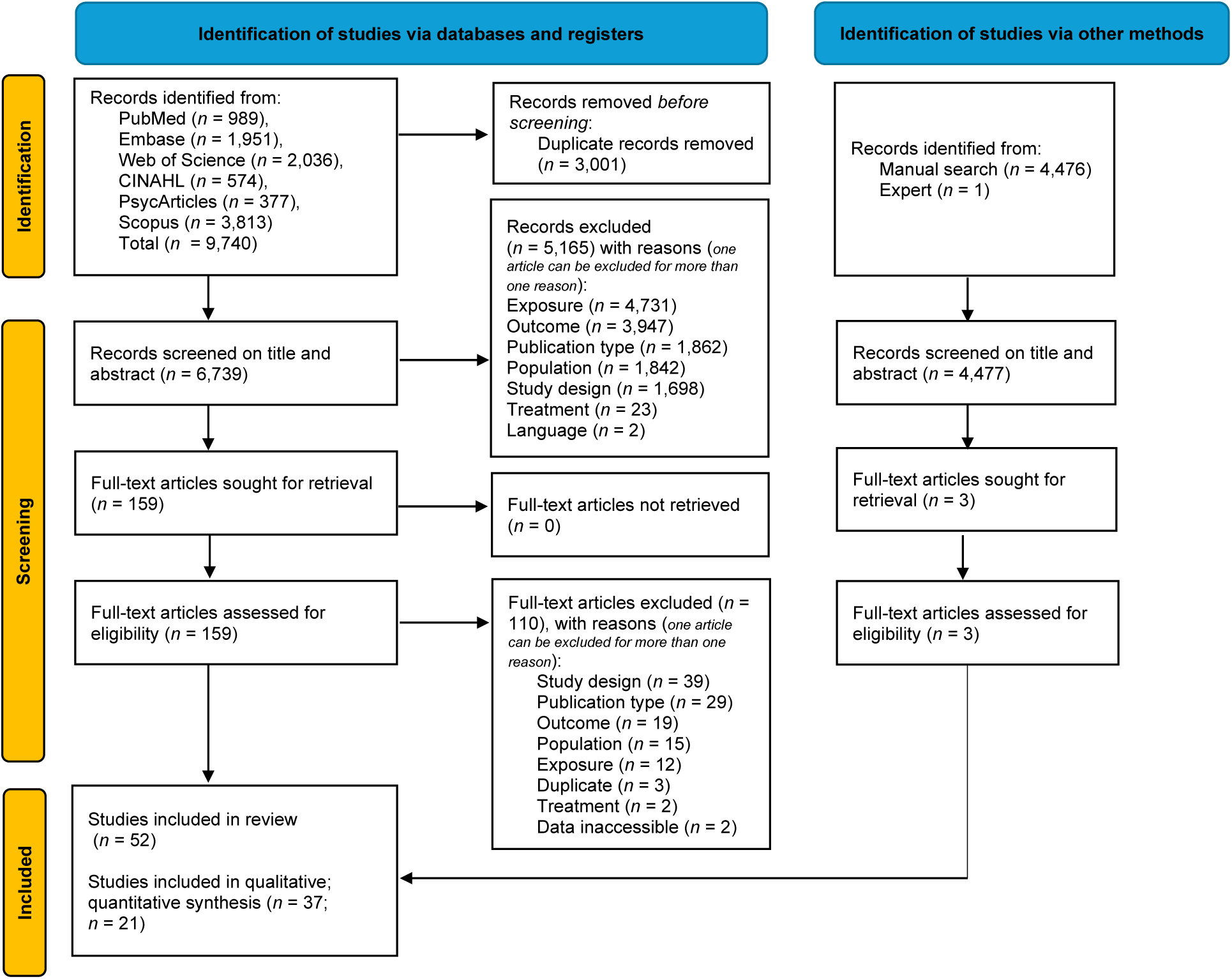
PRISMA Flowchart of study selection. Flowchart depicting the number of records identified, screened, excluded (with reasons), and included in the final qualitative and quantitative synthesis, following the PRISMA 2020 guidelines.

### 3.2 Study Characteristics

The main study characteristics of the 37 studies included in the analyses are summarized below, while a summary of all 52 studies can be found in Supplementary File 10 and an overview of the relevant data of each individual study in Supplementary File 6 - Table S3.

#### 3.2.1 Study Design and Population

Four of the 37 studies had a cross-sectional design, and 33 studies had a case-control design. A total of 1,977 participants were included across the studies (*n* = 840 pain-free controls; *n* = 653 individuals with chronic widespread pain; *n* = 484 individuals with chronic localized pain). Samples sizes ranged from 14 to 59 in the pain-free controls, from eight to 81 in the chronic widespread pain groups, and from 13 to 94 in the chronic localized pain groups. The chronic widespread pain groups only consisted of patients with fibromyalgia and were investigated in 23 studies, except for one study that investigated a mix of patients with chronic widespread and localized back pain (Loffler, et al., 2023). Chronic localized pain groups were investigated in 14 studies and consisted of chronic (low) back pain (Bandeira, et al., 2021; Miyachi, et al., 2025; Muhtz, et al., 2013; Nees, et al., 2019; Tan, et al., 2023; Venezia, et al., 2024), chronic neck pain (Valera-Calero & Varol, 2022), temporomandibular disorders (Maixner, et al., 1997; Mohn, et al., 2008), irritable bowel syndrome (Jarrett, et al., 2016; Jarrett, et al., 2014), chronic pelvic pain (Poli-Neto, et al., 2020), and chronic complex regional pain syndrome (Scheuren, et al., 2023). The mean age ± SD was 42.2 ± 9.3 (range: 27.6-53.3) years for the pain-free controls, 47.1 ± 9.0 (range: 37.3-57.1) years for the chronic widespread pain group, and 40.1 ± 9.9 (range: 27.2-67.9) years for the chronic localized pain groups. Three studies did not report the mean or SD of the participants’ age, or the distribution of sex (Kadetoff & Kosek, 2007, 2010; Reshkova, et al., 2015). In 21 of the 37 studies, only female participants were investigated. All 37 studies used baseline assessments, 14 studies reported data concerning reactivity to a stressor, and 10 studies reported data concerning recovery of the stress systems. Eight studies used a non-painful stressor and seven studies used a painful stressor. One study incorporated both a painful and non-painful stressor (Reyes Del Paso, et al., 2022). In total, 16 studies employed an acute stress induction. Twenty-eight studies only investigated the ANS, eight studies focused solely on the HPA axis, and one study investigated both systems (Kadetoff & Kosek, 2010).

#### 3.2.2 Physiological stress outcome measures

##### Baseline – ANS

Parameters of HRV were assessed in 11 studies, pre-ejection period (PEP) in two studies, HR frequency in 13 studies, BP and cardiac baroreflex sensitivity (BRS) in 16 studies, SC in six studies, and (nor)epinephrine in two studies (Kadetoff & Kosek, 2010; Reshkova, et al., 2015). Parameters of HRV consisted of high frequency (HF) HRV, low frequency (LF) HRV, LF/HF ratio, total power, root mean square of successive differences between normal heartbeats (RMSSD), the mean of standard deviations of the NN intervals (SDNN) and R-R intervals (RRI). The duration of physiological autonomic nervous system (ANS) measurements varied among the studies; however, the majority utilized a standard recording period of five minutes (Bandeira, et al., 2021; Bossenger, et al., 2023; de la Coba, et al., 2018; Jarrett, et al., 2016; Loffler, et al., 2023; Lopez-Lopez, et al., 2021; Venezia, et al., 2024) or ten minutes (Crettaz, et al., 2013; Kadetoff & Kosek, 2010; Reyes Del Paso, et al., 2022; Reyes del Paso, et al., 2011; Van Den Houte, et al., 2018).

##### Baseline – HPA axis

Basal cortisol levels were assessed in nine studies. One study assessed cortisol in hair (Davydov, et al., 2024). Two studies assessed cortisol in serum either in the morning (i.e., 8:00-9:30) or in the early afternoon (de Abreu Freitas, et al., 2012; Kadetoff & Kosek, 2010), and six studies assessed cortisol in saliva at various timepoints: i) throughout the day (Nees, et al., 2019; Wingenfeld, et al., 2010), ii) in the morning (Jarrett, et al., 2014; Valera-Calero & Varol, 2022), and iii) before the start of pain assessments (Meeus, et al., 2008). One study did not specify the timing of the day (Muhtz, et al., 2013). Dehydroepiandrosterone (DHEA) was measured in one study (de Abreu Freitas, et al., 2012).

##### Reactivity – ANS

Both non-painful (i.e., passive visualization task; arithmetic task; Trier Social Stress Test (TSST); personal involvement task; and isometric contraction of the quadriceps muscle) and painful (i.e., cold pressor test, CPT; hot water, mechanical, thermal, and pinprick stimuli) stressors were used to induce an acute stress response. During the stressor, parameters of HRV were examined in five studies, HR in six studies, BP and BRS in eight studies, SC in two studies (Bossenger, et al., 2023; Scheuren, et al., 2023), PEP in two studies (Bossenger, et al., 2023; Reyes Del Paso, et al., 2022), and (nor)epinephrine in one study (Kadetoff & Kosek, 2010).

##### Reactivity – HPA axis

One study measured cortisol at exhaustion of an isometric quadriceps muscle contraction (Kadetoff & Kosek, 2010), and two studies measured cortisol after a CPM procedure with thermal stimuli (Jarrett, et al., 2014; Meeus, et al., 2008).

##### Recovery – ANS

The recovery of HRV and PEP was measured over a five-minute interval immediately after a CPT in two studies (Bandeira, et al., 2021; Reyes Del Paso, et al., 2022), and recovery of HR and BP (sensitivity) ranged from immediately after a stressor for five minutes to 30 minutes after a stressor across five studies (Crettaz, et al., 2013; Kadetoff & Kosek, 2010; Lopez-Lopez, et al., 2021; Reyes del Paso, et al., 2011; Woda, et al., 2013).

##### Recovery – HPA axis

The assessment of recovery of cortisol values was 30 minutes after isometric contractions in one study (Kadetoff & Kosek, 2010), and 45 and 60 minutes after heat stimuli in another study (Muhtz, et al., 2013).

#### 3.2.3 Experimental pain outcome measures

The most reported pain outcome measure was the static QST measure pressure pain threshold (PPTh), assessed at various body sites (in 21/37 studies). Different other static QST methods were used to assess pain sensitivity examined at various locations across the different studies. Cold PThs (CPThs) were assessed in five studies, heat PThs (HPThs) in five studies, electrical PThs (EPThs) in three studies and ischemic PThs in one study. Cold pain tolerances (CPTos) were assessed in two studies, heat PTos in one study, electrical PTos (EPTos) in three studies, and mechanical PTos in 6 studies. Pain intensity ratings to cold stimuli were used in one study, to heat stimuli in two studies, and to mechanical stimuli in one study. Functionality of central pain processing using dynamic QST was evaluated in 13 studies. More specifically, TSP was assessed to evaluate the pain facilitation in three studies, sustained response to evoked pain (SREP) was assessed to evaluate pain facilitation in three studies, CPM was assessed to evaluate the efficacy of descending pain inhibition in six studies and the nociceptive flexion reflex threshold was assessed to evaluate spinal cord responses to noxious input in one study.

All studies measured an experimental pain outcome measure at baseline. In three studies, PPThs were assessed during a stressor (isometric contraction of quadriceps muscle and TSST) (Kadetoff & Kosek, 2007, 2010; Lopez-Lopez, et al., 2021). In five studies, PPThs, PPTos, EPThs or EPTos were assessed after a stressor (which ranged from immediately after a stressor to 30 minutes after a stressor) (Bandeira, et al., 2021; Kadetoff & Kosek, 2007, 2010; Loffler, et al., 2023; Lopez-Lopez, et al., 2021).

### 3.3 Risk of bias within studies

The risk of bias for five studies was assessed using an adapted version of the NOS for cross-sectional studies, while the remaining 47 studies were evaluated with the adapted version of the NOS for case-control studies. The mean number of stars for the case-control studies was 9 stars (of 12) stars, ranging between 5 and 11 stars. The mean number of stars for the cross-sectional studies was 7 (of 12) stars, with one study scoring only 2 stars. Case-control studies scored most of the stars on whether the same methods were used for cases and controls (*n* = 47/47) and lost the most stars on the representativeness of the sample (*n* = 14/47) due to recruiting from one specific setting or directly recruiting. Most studies controlled for the most important primary confounding factors, including 1) age, 2) biological sex and 3) at least one relevant medical condition (*n* = 46/52), and for at least one secondary confounding factor, including a) medication use, b) caffeine use, c) nicotine use, d) alcohol use, e) strenuous activity, f) body mass index, g) hormonal cycle, h) menopause and i) hormonal replacement therapy use (*n* = 46/52). The inter-rater agreement (i.e., Kappa score) between two raters (J.V. and R.D.B.) prior to a consensus meeting was 86.4% (445/515). After comparison, consensus was reached for all criteria. A summary of the risk of bias assessment for each study is presented in Supplementary File 5 - Table S4 (S4a for case-control studies; S4b for cross-sectional studies).

### 3.4 Synthesis of results

Below, results pertaining to the CPP groups are synthesized according to the timing of the stress measurement (i.e., **3.4.1 Baseline stress – pain [Cluster 1]** and **3.4.2 Stress during or after a stressor – pain [Cluster 2a & 2b]**) and based on whether they reflect a change in stress (i.e., **3.4.3 Change in stress – pain [Cluster 3a & 3b]**), a change in pain (**3.4.4 Stress – change in pain [Cluster 4]**) or both (**3.4.5 Change in stress and pain [Cluster 5]**).

A full overview of the associations between physiological stress parameters and experimental pain outcome measures is reported in Supplementary File 7 - Table S5. Results of the quantitative analyses are presented in Table 1. The certainty of evidence is presented in Supplementary File 8 - Table S6 for the qualitative syntheses, and in Supplementary File 9 - Table S7 for the quantitative syntheses. Results relating to pain-free controls can be found in Supplementary File 10.

**Table 1.** Overview of meta-analytic findings.

| Cluster | Stress Marker | Pain Measure | Included Studies + Weight (%) | Population: N | Correlation (r) | 95% CI | Overall Effect (Z-value) | P-value | Heterogeneity ($I^2$) |
| --- | --- | --- | --- | --- | --- | --- | --- | --- | --- |
| 1 | HR at baseline | PPTH at baseline | N = 7<br>Kadetoff et al. (2007; 7.4%); Kadetoff et al. (2010; 6.8%); Lopez-Lopez et al. (2021; 7.9%); Mohn et al. (2008; 11.6%); Poli Neto et al. (2020; 9.5%); Valero-Calero et al. (2022; 47.9%); Zamunér et al. (2016; 28.8%) | CPP: 211 | - .10 | [- 0.27; 0.08] | - 1.07 | .28 | 26.3% |
|  |  |  | N = 4<br>Kadetoff et al. (2007; 23.7%); Kadetoff et al. (2010; 22%); Lopez-Lopez et al. (2021; 25.4%); Zamunér et al. (2016; 28.8%) | CPP – widespread: 71 | - .16 | [- 0.48; 0.2] | - 0.85 | .39 | 49.9% |
|  |  |  | N = 3<br>Mohn et al. (2008; 16.8%); Poli Neto et al. (2020; 13.7%); Valero-Calero et al. (2022; 69.5%) | CPP – localized: 140 | - .06 | [- 0.29; 0.17] | - 0.5 | .61 | 31.8% |
|  |  |  | N = 4<br>Kadetoff et al. (2007; 17.5%); Kadetoff et al. (2010; 16.3%); Lopez-Lopez et al. (2021; 43.8%); Poli Neto et al. (2020; 22.5%) | HC: 92 | - .15 | [- 0.36; 0.07] | - 1.37 | .17 | 0% |
| 1 | HR at baseline | PPTH during stressor | N = 3<br>Kadetoff et al. (2007; 33.3%); Kadetoff et al. (2010; 31%); Lopez-Lopez et al. (2021; 35.7%) | CPP – widespread: 51 | - .43 | [- 0.7, - 0.06] | - 2.25 | **.02** | 42.3% |
|  |  |  | N = 3<br>Kadetoff et al. (2007; 22.6%); Kadetoff et al. (2010; 21%); Lopez-Lopez et al. (2021; 56.5%) | HC: 71 | -.15 | [- 0.38; 0.1] | - 1.21 | .23 | 0% |
| 1 | HR at baseline | PPT<sub>h</sub> after stressor | N = 3<br>Kadetoff et al. (2007; 33.3%); Kadetoff et al. (2010; 31%); Lopez-Lopez et al. (2021; 35.7%) | CPP – widespread: 51 | -.37 | [- 0.67; 0.05] | - 1.74 | .08 | 51.2% |
|  |  |  | N = 3<br>Kadetoff et al. (2007; 22.6%); Kadetoff et al. (2010; 21%); Lopez-Lopez et al. (2021; 56.5%) | HC: 71 | -.21 | [- 0.44; 0.03] | - 1.71 | .09 | 0% |
| 1 | HR at baseline | EPTh at baseline | N = 3<br>Löffler et al. (2023; 19%); Mohn et al. (2008; 22%); Van Middendorp et al. (2013; 59%) | CPP: 109 | -.39 | [- 0.88; 0.52] | - 0.81 | .42 | 95.3% |
|  |  |  | N = 2<br>Löffler et al. (2023; 21.1%); Van Middendorp et al. (2013; 78.9%) | HC: 77 | .08 | [- 0.16; 0.3] | 0.64 | .52 | 0% |
| 1 | HR at baseline | PPT<sub>o</sub> at baseline | N = 2<br>Lopez-Lopez et al. (2020; 31.3%); Mohn et al (2008; 68.8%) | CPP: 38 | -.17 | [- 0.48; 0.17] | - 0.98 | .33 | 0% |
| 1 | HR at baseline | EPTo at baseline | N = 3<br>Löffler et al. (2023; 19%); Mohn et al. (2008; 22%); Van Middendorp et al. (2013; 59%) | CPP: 109 | -.003 | [- 0.22; 0.21] | - 0.03 | .98 | 12.4% |
|  |  |  | N = 2<br>Löffler et al. (2023; 21.1%); Van | HC: 77 | -.04 | [- 0.43; 0.36] | - 0.02 | .84 | 57.2% |
|  |  |  | Middendorp et al. (2013; 78.9%) |  |  |  |  |  |  |
| 1 | SBP at baseline | PPTh at baseline | N = 7<br>De La Coba et al. (2018; 13.9%);<br>Kadetoff et al. (2007; 7.2%); Kadetoff et al. (2010; 6.7%);<br>Lopez-Lopez et al. (2021; 7.7%); Poli Neto et al. (2020; 9.2%); Valero-Calero et al. (2022; 46.7%);<br>Zamunér et al. (2016; 8.7%) | CPP: 216 | - .07 | [- 0.21; 0.07] | - 0.96 | .34 | 0% |
|  |  |  | N = 4<br>Kadetoff et al. (2007; 23.7%); Kadetoff et al. (2010; 22%);<br>Lopez-Lopez et al. (2021; 25.4%);<br>Zamunér et al. (2016; 28.8%) | CPP –<br>widespread: 71 | - .11 | [- 0.35; 0.14] | - 0.84 | .40 | 0% |
|  |  |  | N = 3<br>De La Coba et al. (2018; 19.9%); Poli Neto et al. (2020; 13.2%); Valero-Calero et al. (2022; 66.9%) | CPP –<br>localized: 145 | - .03 | [- 0.31; 0.26] | - 0.19 | .85 | 55.5% |
|  |  |  | N = 5<br>De La Coba et al. (2018; 23.1%);<br>Kadetoff et al. (2007; 13.5%); Kadetoff et al. (2010; 12.5%);<br>Lopez-Lopez et al. (2021; 33.7%); Poli Neto et al. (2020; 17.3%) | HC: 119 | .07 | [- 0.26; 0.39] | 0.42 | .67 | 66.5% |
| 1 | SBP at baseline | PPTH during stressor | N = 3<br>Kadetoff et al. (2007; 33.3%); Kadetoff et al. (2010; 31%); Lopez-Lopez et al. (2021; 35.7%) | CPP –<br>widespread:<br>51 | .008 | [- 0.34;<br>0.36] | 0.04 | .97 | 30.6% |
|  |  |  | N = 3<br>Kadetoff et al. (2007; 22.6%); Kadetoff et al. (2010; 21%); Lopez-Lopez et al. (2021; 56.5%) | HC: 71 | .11 | [- 0.14;<br>0.35] | 0.89 | .38 | 0% |
| 1 | SBP at baseline | PPTH after stressor | N = 3<br>Kadetoff et al. (2007; 33.3%); Kadetoff et al. (2010; 31%); Lopez-Lopez et al. (2021; 35.7%) | CPP –<br>widespread:<br>51 | .009 | [- 0.45;<br>0.46] | 0.03 | .97 | 62.4% |
|  |  |  | N = 3<br>Kadetoff et al. (2007; 22.6%); Kadetoff et al. (2010; 21%); Lopez-Lopez et al. (2021; 56.5%) | HC: 71 | .05 | [- 0.2;<br>0.29] | 0.39 | .70 | 0% |
| 1 | SBP at baseline | CPTH at baseline | N = 2<br>Del Paso et al. (2011; 58.2%); Del Paso et al. (2022; 41.8%) | CPP –<br>widespread:<br>61 | .05 | [- 0.22;<br>0.3] | 0.33 | .74 | 0% |
| 1 | SBP at baseline | PPTo at baseline | N = 2<br>De La Coba et al. (2018; 73%); Lopez-Lopez et al. (2020; 27%) | CPP –<br>widespread:<br>43 | - .05 | [- 0.36;<br>0.27] | - 0.31 | .76 | 0% |
|  |  |  | N = 2<br>De La Coba et al. (2018; 40.7%); Lopez-Lopez et al. (2020; 59.3%) | HC: 65 | .06 | [- 0.2;<br>0.3] | 0.44 | .66 | 0% |
| 1 | SBP at baseline | CPTo at baseline | N = 2<br>Del Paso et al. (2011; 58.2%); Del | CPP –<br>widespread:<br>61 | - .17 | [- 0.41;<br>0.09] | - 1.26 | .21 | 0% |
|  |  |  | Paso et al. (2022; 41.8%) |  |  |  |  |  |  |
| 1 | DBP at baseline | PPTh at baseline | N = 6<br>De La Coba et al. (2018; 15%); Kadetoff et al. (2007; 7.8%); Kadetoff et al. (2010; 7.2%); Poli Neto et al. (2020; 10%); Valero-Calero et al. (2022; 50.6%); Zamunér et al. (2016; 9.4%) | CPP: 198 | - .06 | [- 0.2; 0.09] | - 0.76 | .45 | 0% |
|  |  |  | N = 3<br>Kadetoff et al. (2007; 31.8%); Kadetoff et al. (2010; 29.6%); Zamunér et al. (2016; 38.6%) | CPP – widespread: 53 | - .12 | [- 0.39; 0.18] | - 0.76 | .45 | 0% |
|  |  |  | N = 3<br>De La Coba et al. (2018; 19.9%); Poli Neto et al. (2020; 13.2%); Valero-Calero et al. (2022; 66.9%) | CPP – localized: 145 | - .001 | [- 0.28; 0.28] | - 0.19 | >.99 | 53.6% |
|  |  |  | N = 4<br>De La Coba et al. (2018; 33.3%); Kadetoff et al. (2007; 22%); Kadetoff et al. (2010; 19.8%); Poli Neto et al. (2020; 25.9%) | HC: 81 | - .008 | [- 0.28; 0.27] | - 0.05 | .96 | 28.2% |
| 1 | DBP at baseline | PPTh during stressor | N = 2<br>Kadetoff et al. (2007; 51.9%); Kadetoff et al. (2010; 48.2%) | CPP – widespread: 33 | - .07 | [- 0.42; 0.3] | - 0.36 | .72 | 0% |
|  |  |  | N = 2<br>Kadetoff et al. (2007; 51.9%); Kadetoff et al. (2010; 48.2%) | HC: 33 | - .04 | [- 0.4; 0.33] | - 0.21 | .83 | 0% |
| 1 | DBP at baseline | PPTh after stressor | N = 2<br>Kadetoff et al. (2007; 51.9%); Kadetoff et al. (2010; 48.2%) | CPP –<br>widespread:<br>33 | -.20 | [- 0.52;<br>0.18] | - 1.04 | .30 | 0% |
|  |  |  | N = 2<br>Kadetoff et al. (2007; 51.9%); Kadetoff et al. (2010; 48.2%) | HC: 33 | .05 | [- 0.4;<br>0.32] | - 0.27 | .79 | 0% |
| 1 | DBP at baseline | CPT<sub>h</sub> at baseline | N = 2<br>Del Paso et al. (2011; 58.2%); Del Paso et al. (2022; 41.8%) | CPP –<br>widespread:<br>61 | .01 | [- 0.25;<br>0.27] | 0.09 | .93 | 0% |
| 1 | DBP at baseline | CPT<sub>o</sub> at baseline | N = 2<br>Del Paso et al. (2011; 58.2%); Del Paso et al. (2022; 41.8%) | CPP –<br>widespread:<br>61 | -.20 | [- 0.43;<br>0.07] | - 1.48 | .14 | 0% |
| 1 | MAP at baseline | EPTh at baseline | N = 2<br>Mohn et al. (2008; 27.2%); Van Middendorp et al. (2013; 72.8%) | CPP: 87 | .35 | [0.06;<br>0.6] | 2.33 | **.02** | 42.1% |
| 1 | MAP at baseline | EPT<sub>o</sub> at baseline | N = 2<br>Mohn et al. (2008; 27.2%); Van Middendorp et al. (2013; 72.8%) | CPP: 87 | .30 | [0.02;<br>0.53] | 2.13 | **.03** | 33.5% |
| 1 | BRS at baseline | CPT<sub>h</sub> at baseline | N = 2<br>Del Paso et al. (2011; 58.2%); Del Paso et al. (2022; 41.8%) | CPP –<br>widespread:<br>61 | -.05 | [- 0.3;<br>0.22] | - 0.34 | .74 | 0% |
| 1 | BRS at baseline | CPT<sub>o</sub> at baseline | N = 2<br>Del Paso et al. (2011; 58.2%); Del Paso et al. (2022; 41.8%) | CPP –<br>widespread:<br>61 | .12 | [- 0.15;<br>0.36] | 0.85 | .39 | 0% |
| 1 | HF HRV at baseline | PPTh at baseline | N = 2<br>Bandeira et al. (2021; 72.1%); | CPP: 67 | .007 | [- 0.3;<br>0.32] | 0.04 | .97 | 30.9% |
|  |  |  | Zamunér et al. (2016; 27.9%) |  |  |  |  |  |  |
| 1 | HF HRV at baseline | CPT<sub>h</sub> at baseline | N = 2<br>Del Paso et al. (2011; 58.2%); Del Paso et al. (2022; 41.8%) | CPP – widespread: 61 | -.08 | [- 0.33; 0.18] | - 0.61 | .55 | 0% |
| 1 | HF HRV at baseline | CPT<sub>o</sub> at baseline | N = 2<br>Del Paso et al. (2011; 58.2%); Del Paso et al. (2022; 41.8%) | CPP – widespread: 61 | .10 | [- 0.16; 0.35] | 0.75 | .46 | 0% |
| 1 | LF HRV at baseline | PPT<sub>h</sub> at baseline | N = 2<br>Bandeira et al. (2021; 72.1%); Zamunér et al. (2016; 27.9%) | CPP: 67 | .04 | [- 0.21; 0.28] | 0.29 | .77 | 0% |
| 1 | LF HRV at baseline | CPT<sub>h</sub> at baseline | N = 2<br>Del Paso et al. (2011; 58.2%); Del Paso et al. (2022; 41.8%) | CPP – widespread: 61 | -.09 | [- 0.34; 0.18] | - 0.65 | .52 | 0% |
| 1 | LF HRV at baseline | CPT<sub>o</sub> at baseline | N = 2<br>Del Paso et al. (2011; 58.2%); Del Paso et al. (2022; 41.8%) | CPP – widespread: 61 | .12 | [- 0.15; 0.36] | 0.87 | .46 | 0% |
| 1 | LF/HF HRV at baseline | PPT<sub>h</sub> at baseline | N = 3<br>Bandeira et al. (2021; 42.31%); Miyachi et al. (2025; 41.35%); Zamunér et al. (2016; 16.35%) | CPP: 113 | .261 | [- 0.08; 0.29] | 1.12 | .26 | 0.01% |
|  |  |  | N = 2<br>Bandeira et al. (2021; 50.57%); Miyachi et al. (2025; 49.43%) | CPP – localized: 93 | .17 | [- 0.04; 0.36] | 1.6 | .11 | 0% |
| 1 | RRI at baseline | CPT<sub>h</sub> at baseline | N = 2<br>Del Paso et al. (2011; 58.2%); Del | CPP – widespread: 61 | .12 | [- 0.14; 0.37] | 0.89 | .37 | 0% |
|  |  |  | Paso et al. (2022; 41.8%) |  |  |  |  |  |  |
| 1 | RRI at baseline | CPT<sub>0</sub> at baseline | N = 2<br>Del Paso et al. (2011; 58.2%); Del Paso et al. (2022; 41.8%) | CPP – widespread: 61 | .04 | [- 0.22; 0.29] | 0.28 | .78 | 0% |
| 1 | SC at baseline | PPTh at baseline | N = 2<br>Garcia-Hernandez et al. (2022 ; 66.7%); Tan et al. (2023; 33.3%) | CPP: 45 | .19 | [- 0.12; 0.46] | 1.19 | .23 | 0% |
|  |  |  | N = 2<br>Garcia-Hernandez et al. (2022 ; 66.7%); Tan et al. (2023; 33.3%) | HC: 46 | .01 | [- 0.41; 0.40] | - 0.03 | .98 | 42.68% |
| 1 | Epinephrine at baseline | PPTh at baseline | N = 2<br>Kadetoff et al. (2010; 41.94%); Reshkova et al. (2015; 58.06%) | CPP – widespread: 37 | .01 | [- 0.33; 0.34] | 0.03 | 0.97 | 0% |
| 1 | Norepinephrine at baseline | PPTh at baseline | N = 2<br>Kadetoff et al. (2010; 41.94%); Reshkova et al. (2015; 58.06%) | CPP – widespread: 37 | .06 | [- 0.28; 0.39] | 0.36 | 0.72 | 0% |
| 1 | Salivary cortisol at baseline | Heat pain ratings at baseline | N = 2<br>Meeus et al. (2008; 62.2%); Muhtz et al. (2013; 37.8%) | CPP: 51 | - .05 | [- 0.56; 0.48] | - 0.18 | .86 | 72.6% |
|  |  |  | N = 2<br>Meeus et al. (2008; 48.3%); Muhtz et al. (2013; 51.7%) | HC: 64 | - .06 | [- 0.58; 0.5] | - 0.19 | .86 | 81.9% |
| 1 | Cortisol at baseline | PPTh at baseline | N = 4<br>de Abreu Freitas et al. (2012; 10.2%); Kadetoff et al. (2010; 9.5%); Nees et al. (2019; 13.9%); Valera-Calero et al. (2022; 66.4%) | CPP: 149 | - .22 | [- 0.37; - 0.05] | - 2.59 | **.01** | 0.01% |
|  |  |  | N = 2<br>de Abreu Freitas et al. (2012; 51.9%); Kadetoff et al. (2010; 48.2%) | CPP – widespread: 33 | - .31 | [- 0.6; 0.06] | - 1.66 | .10 | 0% |
|  |  |  | N = 2<br>Nees et al. (2019; 17.3%); Valera-Calero et al. (2022; 82.7%) | CPP – localized: 116 | - .12 | [- 0.46; 0.24] | - 0.64 | .52 | 58.8% |
|  |  |  | N = 3<br>de Abreu Freitas et al. (2012; 28.6%); Kadetoff et al. (2010; 23.2%); Nees et al. (2019; 48.2%) | HC: 65 | .02 | [- 0.24; 0.27] | 0.11 | .91 | 0% |
| 1 | Cortisol AUCg at baseline | HPTh at baseline | N = 2<br>Nees et al. (2019; 51.4%); Wingenfeld et al. (2010; 48.7%) | CPP: 43 | - .10 | [- 0.4; 0.22] | - 0.59 | .56 | 0% |
| 1 | Cortisol AUCg at baseline | PPTH at baseline | N = 2<br>Nees et al. (2019; 51.4%); Wingenfeld et al. (2010; 48.7%) | CPP: 43 | - .10 | [- 0.4; 0.22] | - 0.59 | .56 | 0% |
| 2a | HR during a stressor | PPTH at baseline | N = 3<br>Kadetoff et al. (2007; 33.3%); Kadetoff et al. (2010; 31%); Lopez-Lopez et al. (2021; 35.7%) | CPP – widespread: 51 | - .38 | [- 0.61; - 0.1] | - 2.61 | **.01** | 0% |
|  |  |  | N = 3<br>Kadetoff et al. (2007; 22.6%); Kadetoff et al. (2010; 21%); Lopez-Lopez et al. (2021; 56.5%) | HC: 71 | - .15 | [- 0.38; 0.1] | - 1.19 | .23 | 0% |
| 2b | HR during recovery | PPTH at baseline | N = 5<br>De Bruijn et al. (2011; 18.99%); Kadetoff et al. (2007; 27.85%); Kadetoff et al. (2010; 17.72%); | CPP: 94 | - .40 | [- 0.68; - 0.05] | - 2.2 | **.03** | 67.27% |
|  |  |  | Lopez-Lopez et al. (2021; 16.46%);<br>Mohn et al. (2008; 18.99%) |  |  |  |  |  |  |
|  |  |  | N = 4<br>De Bruijn et al. (2011; 26.32%);<br>Kadetoff et al. (2007; 24.56%); Kadetoff et al. (2010; 22.81%);<br>Lopez-Lopez et al. (2021; 26.32%) | CPP –<br>widespread:<br>69 | - .55 | [- 0.71; - 0.34] | - 4.54 | < .005 | 5.07% |
|  |  |  | N = 3<br>Kadetoff et al. (2007; 22.6%); Kadetoff et al. (2010; 21%);<br>Lopez-Lopez et al. (2021; 56.5%) | HC: 71 | - .10 | [- 0.33; 0.15] | - 0.77 | .44 | 0% |
| 2a | HR during a stressor | PPTh during a stressor | N = 3<br>Kadetoff et al. (2007; 33.3%); Kadetoff et al. (2010; 31%);<br>Lopez-Lopez et al. (2021; 35.7%) | CPP –<br>widespread:<br>51 | - .34 | [- 0.64; - 0.06] | - 1.69 | .09 | 43.7% |
|  |  |  | N = 3<br>Kadetoff et al. (2007; 22.6%); Kadetoff et al. (2010; 21%);<br>Lopez-Lopez et al. (2021; 56.5%) | HC: 71 | - .19 | [- 0.41; 0.06] | - 1.51 | .13 | 0% |
| 2b | HR during recovery | PPTh during a stressor | N = 3<br>Kadetoff et al. (2007; 33.3%); Kadetoff et al. (2010; 31%);<br>Lopez-Lopez et al. (2021; 35.7%) | CPP –<br>widespread:<br>51 | - .53 | [- 0.71; - 0.27] | - 3.78 | < .005 | 0% |
|  |  |  | N = 3<br>Kadetoff et al. (2007; 22.6%); Kadetoff et al. (2010; 21%);<br>Lopez-Lopez et al. (2021; 56.5%) | HC: 71 | - .05 | [- 0.29; 0.19] | - 0.42 | .68 | 0% |
| 2a | HR during a stressor | PPTH after stressor | N = 3<br>Kadetoff et al. (2007; 33.3%); Kadetoff et al. (2010; 31%); Lopez-Lopez et al. (2021; 35.7%) | CPP –<br>widespread:<br>51 | - .36 | [- 0.59; - 0.07] | - 2.44 | .02 | 0% |
|  |  |  | N = 3<br>Kadetoff et al. (2007; 22.6%); Kadetoff et al. (2010; 21%); Lopez-Lopez et al. (2021; 56.5%) | HC: 71 | - .28 | [- 0.49; - 0.03] | - 2.22 | .03 | 0% |
| 2b | HR during recovery | PPTH after stressor | N = 3<br>Kadetoff et al. (2007; 33.3%); Kadetoff et al. (2010; 31%); Lopez-Lopez et al. (2021; 35.7%) | CPP –<br>widespread:<br>51 | - .48 | [- 0.68; - 0.22] | - 3.41 | < .005 | 0% |
|  |  |  | N = 3<br>Kadetoff et al. (2007; 22.6%); Kadetoff et al. (2010; 21%); Lopez-Lopez et al. (2021; 56.5%) | HC: 71 | - .17 | [- 0.4; 0.08] | - 1.34 | .18 | 0% |
| 2a | SBP during a stressor | PPTH at baseline | N = 3<br>Kadetoff et al. (2007; 33.3%); Kadetoff et al. (2010; 31%); Lopez-Lopez et al. (2021; 35.7%) | CPP –<br>widespread:<br>51 | - .20 | [- 0.47; 0.01] | - 1.32 | .19 | 0% |
|  |  |  | N = 3<br>Kadetoff et al. (2007; 22.6%); Kadetoff et al. (2010; 21%); Lopez-Lopez et al. (2021; 56.5%) | HC: 71 | - .04 | [- 0.28; 0.21] | - 0.28 | .78 | 0% |
| 2b | SBP during recovery | PPTH at baseline | N = 3<br>Kadetoff et al. (2007; 33.3%); Kadetoff et al. (2010; 31%); Lopez-Lopez et al. (2021; 35.7%) | CPP –<br>widespread:<br>51 | .10 | [- 0.21; 0.38] | 0.61 | .54 | 0% |
|  |  |  | N = 3<br>Kadetoff et al. (2007; 22.6%); Kadetoff et al. (2010; 21%); Lopez-Lopez et al. (2021; 56.5%) | HC: 71 | -.029 | [- 0.27; 0.22] | - 0.23 | .82 | 0% |
| 2a | SBP during a stressor | PPTH during a stressor | N = 3<br>Kadetoff et al. (2007; 33.3%); Kadetoff et al. (2010; 31%); Lopez-Lopez et al. (2021; 35.7%) | CPP – widespread: 51 | -.12 | [- 0.44; 0.22] | - 0.7 | .48 | 22.4% |
|  |  |  | N = 3<br>Kadetoff et al. (2007; 22.6%); Kadetoff et al. (2010; 21%); Lopez-Lopez et al. (2021; 56.5%) | HC: 71 | .04 | [- 0.21; 0.28] | 0.27 | .78 | 0% |
| 2b | SBP during recovery | PPTH during a stressor | N = 3<br>Kadetoff et al. (2007; 33.3%); Kadetoff et al. (2010; 31%); Lopez-Lopez et al. (2021; 35.7%) | CPP – widespread: 51 | 0 | [- 0.3; 0.29] | - 0.03 | .98 | 0% |
|  |  |  | N = 3<br>Kadetoff et al. (2007; 22.6%); Kadetoff et al. (2010; 21%); Lopez-Lopez et al. (2021; 56.5%) | HC: 71 | .10 | [- 0.14; 0.34] | 0.83 | .41 | 0% |
| 2a | SBP during a stressor | PPTH after stressor | N = 3<br>Kadetoff et al. (2007; 33.3%); Kadetoff et al. (2010; 31%); Lopez-Lopez et al. (2021; 35.7%) | CPP – widespread: 51 | -.18 | [- 0.56; 0.27] | - 0.78 | .44 | 54.5% |
|  |  |  | N = 3<br>Kadetoff et al. (2007; 22.6%); Kadetoff et al. (2010; 21%); Lopez-Lopez et al. (2021; 56.5%) | HC: 71 | .02 | [- 0.23; 0.26] | 0.15 | .88 | 0% |
| 2b | SBP during recovery | PPTH after stressor | N = 3<br>Kadetoff et al. (2007; 33.3%); Kadetoff et al. (2010; 31%); Lopez-Lopez et al. (2021; 35.7%) | CPP –<br>widespread:<br>51 | .05 | [- 0.25;<br>0.34] | 0.33 | .74 | 0% |
|  |  |  | N = 3<br>Kadetoff et al. (2007; 22.6%); Kadetoff et al. (2010; 21%); Lopez-Lopez et al. (2021; 56.5%) | HC: 71 | .05 | [- 0.24;<br>0.327] | 0.32 | .75 | 21.95% |
| 2a | SBP during a stressor | CPTH at baseline | N = 2<br>Del Paso et al. (2011; 58.2%); Del Paso et al. (2022; 41.8%) | CPP –<br>widespread:<br>61 | .16 | [- 0.1;<br>0.4] | 1.21 | .23 | 0% |
| 2b | SBP during recovery | CPTH at baseline | N = 2<br>Del Paso et al. (2011; 58.2%); Del Paso et al. (2022; 41.8%) | CPP –<br>widespread:<br>61 | .09 | [- 0.17;<br>0.34] | 0.7 | .49 | 0% |
| 2a | SBP during a stressor | CPTo at baseline | N = 2<br>Del Paso et al. (2011; 58.2%); Del Paso et al. (2022; 41.8%) | CPP –<br>widespread:<br>61 | .07 | [- 0.2;<br>0.32] | 0.48 | .63 | 0% |
| 2b | SBP during recovery | CPTo at baseline | N = 2<br>Del Paso et al. (2011; 58.2%); Del Paso et al. (2022; 41.8%) | CPP –<br>widespread:<br>61 | .17 | [- 0.1;<br>0.41] | 1.25 | .21 | 0% |
| 2a | DBP during a stressor | PPTH at baseline | N = 2<br>Kadetoff et al. (2007; 51.9%); Kadetoff et al. (2010; 48.2%) | CPP –<br>widespread:<br>33 | - .10 | [- 0.45;<br>0.27] | - 0.53 | .6 | 0% |
|  |  |  | N = 2<br>Kadetoff et al. (2007; 51.9%); Kadetoff et al. (2010; 48.2%) | HC: 33 | - .34 | [- 0.63;<br>0.02] | - 1.86 | .06 | 0% |
| 2b | DBP during recovery | PPT<sub>h</sub> at baseline | N = 2<br>Kadetoff et al. (2007; 51.9%); Kadetoff et al. (2010; 48.2%) | CPP –<br>widespread:<br>33 | .03 | [- 0.34;<br>0.39] | 0.15 | .88 | 0% |
|  |  |  | N = 2<br>Kadetoff et al. (2007; 51.9%); Kadetoff et al. (2010; 48.2%) | HC: 33 | - .23 | [- 0.55;<br>0.14] | - 1.21 | .23 | 0% |
| 2a | DBP during a stressor | PPT<sub>h</sub> during a stressor | N = 2<br>Kadetoff et al. (2007; 51.9%); Kadetoff et al. (2010; 48.2%) | CPP –<br>widespread:<br>33 | - .01 | [- 0.37;<br>0.35] | - 0.05 | .96 | 0% |
|  |  |  | N = 2<br>Kadetoff et al. (2007; 51.9%); Kadetoff et al. (2010; 48.2%) | HC: 33 | .23 | [- 0.54;<br>0.14] | - 1.21 | .23 | 0% |
| 2b | DBP during recovery | PPT<sub>h</sub> during a stressor | N = 2<br>Kadetoff et al. (2007; 51.9%); Kadetoff et al. (2010; 48.2%) | CPP –<br>widespread:<br>33 | - .04 | [- 0.4;<br>0.32] | - 0.22 | .83 | 0% |
|  |  |  | N = 2<br>Kadetoff et al. (2007; 51.9%); Kadetoff et al. (2010; 48.2%) | HC: 33 | - .19 | [- 0.52;<br>0.18] | - 1 | .32 | 0% |
| 2a | DBP during a stressor | PPT<sub>h</sub> after stressor | N = 2<br>Kadetoff et al. (2007; 51.9%); Kadetoff et al. (2010; 48.2%) | CPP –<br>widespread:<br>33 | - .30 | [- 0.6;<br>0.07] | - 1.62 | .1 | 0% |
|  |  |  | N = 2<br>Kadetoff et al. (2007; 51.9%); Kadetoff et al. (2010; 48.2%) | HC: 33 | - .22 | [- 0.54;<br>0.15] | - 1.18 | .24 | 0% |
| 2b | DBP during recovery | PPT<sub>h</sub> after stressor | N = 2<br>Kadetoff et al. (2007; 51.9%); Kadetoff et al. (2010; 48.2%) | CPP –<br>widespread:<br>33 | - .17 | [- 0.5;<br>0.21] | - 0.86 | .39 | 0% |
|  |  |  | N = 2<br>Kadetoff et al. (2007; 51.9%); Kadetoff et al. (2010; 48.2%) | HC: 33 | - .11 | [- 0.45;<br>0.26] | - 0.55 | .58 | 0% |
| 2a | DBP during a stressor | CPT<sub>H</sub> at baseline | N = 2<br>Del Paso et al. (2011; 58.2%); Del Paso et al. (2022; 41.8%) | CPP – widespread: 61 | .11 | [- 0.16; 0.36] | 0.79 | .43 | 0% |
| 2b | DBP during recovery | CPT<sub>H</sub> at baseline | N = 2<br>Del Paso et al. (2011; 58.2%); Del Paso et al. (2022; 41.8%) | CPP – widespread: 61 | .07 | [- 0.19; 0.32] | 0.53 | .6 | 0% |
| 2a | DBP during a stressor | CPT<sub>O</sub> at baseline | N = 2<br>Del Paso et al. (2011; 58.2%); Del Paso et al. (2022; 41.8%) | CPP – widespread: 61 | - .10 | [- 0.38; 0.2] | - 0.67 | .5 | 22% |
| 2b | DBP during recovery | CPT<sub>O</sub> at baseline | N = 2<br>Del Paso et al. (2011; 58.2%); Del Paso et al. (2022; 41.8%) | CPP – widespread: 61 | .20 | [- 0.06; 0.44] | 1.49 | .14 | 0% |
| 2a | BRS during a stressor | CPT<sub>H</sub> at baseline | N = 2<br>Del Paso et al. (2011; 58.2%); Del Paso et al. (2022; 41.8%) | CPP – widespread: 61 | - .06 | [- 0.31; 0.2] | - 0.45 | .65 | 0% |
| 2b | BRS during recovery | CPT<sub>H</sub> at baseline | N = 2<br>Del Paso et al. (2011; 58.2%); Del Paso et al. (2022; 41.8%) | CPP – widespread: 61 | .08 | [- 0.18; 0.33] | 0.59 | .56 | 0% |
| 2a | BRS during a stressor | CPT<sub>O</sub> at baseline | N = 2<br>Del Paso et al. (2011; 58.2%); Del Paso et al. (2022; 41.8%) | CPP – widespread: 61 | .03 | [- 0.23; 0.29] | 0.24 | .81 | 0% |
| 2b | BRS during recovery | CPT<sub>O</sub> at baseline | N = 2<br>Del Paso et al. (2011; 58.2%); Del Paso et al. (2022; 41.8%) | CPP – widespread: 61 | .13 | [- 0.14; 0.37] | 0.94 | .35 | 0% |
| 2a | HF HRV during a stressor | CPT<sub>h</sub> at baseline | N = 2<br>Del Paso et al. (2011; 58.2%); Del Paso et al. (2022; 41.8%) | CPP – widespread: 61 | .04 | [- 0.22; 0.3] | 0.33 | .74 | 0% |
| 2b | HF HRV during recovery | CPT<sub>h</sub> at baseline | N = 2<br>Del Paso et al. (2011; 58.2%); Del Paso et al. (2022; 41.8%) | CPP – widespread: 61 | .09 | [- 0.17; 0.34] | 0.66 | .51 | 0% |
| 2a | HF HRV during a stressor | CPT<sub>o</sub> at baseline | N = 2<br>Del Paso et al. (2011; 58.2%); Del Paso et al. (2022; 41.8%) | CPP – widespread: 61 | .28 | [0; 0.51] | 1.96 | .05 | 12.2% |
| 2b | HF HRV during recovery | CPT<sub>o</sub> at baseline | N = 2<br>Del Paso et al. (2011; 58.2%); Del Paso et al. (2022; 41.8%) | CPP – widespread: 61 | .31 | [0.06; 0.53] | 2.36 | **.02** | 0.5% |
| 2a | LF HRV during a stressor | CPT<sub>h</sub> at baseline | N = 2<br>Del Paso et al. (2011; 58.2%); Del Paso et al. (2022; 41.8%) | CPP – widespread: 61 | .14 | [- 0.12; 0.385] | 1.05 | .29 | 0% |
| 2b | LF HRV during recovery | CPT<sub>h</sub> at baseline | N = 2<br>Del Paso et al. (2011; 58.2%); Del Paso et al. (2022; 41.8%) | CPP – widespread: 61 | .13 | [- 0.14; 0.37] | 0.93 | .35 | 0% |
| 2a | LF HRV during a stressor | CPT<sub>o</sub> at baseline | N = 2<br>Del Paso et al. (2011; 58.2%); Del Paso et al. (2022; 41.8%) | CPP – widespread: 61 | .20 | [- 0.06; 0.44] | 1.51 | .13 | 0% |
| 2b | LF HRV during recovery | CPT<sub>o</sub> at baseline | N = 2<br>Del Paso et al. (2011; 58.2%); Del Paso et al. (2022; 41.8%) | CPP – widespread: 61 | .12 | [- 0.14; 0.37] | 0.92 | .36 | 0% |
| 2a | RRI during a stressor | CPT<sub>h</sub> at baseline | N = 2<br>Del Paso et al. (2011; 58.2%); Del Paso et al. (2022; 41.8%) | CPP – widespread: 61 | .07 | [- 0.19; 0.32] | 0.5 | .62 | 0% |
| 2b | RRI during recovery | CPT<sub>h</sub> at baseline | N = 2<br>Del Paso et al. (2011; 58.2%); Del Paso et al. (2022; 41.8%) | CPP – widespread: 61 | .13 | [- 0.13; 0.38] | 0.99 | .32 | 0% |
| 2a | RRI during a stressor | CPT<sub>o</sub> at baseline | N = 2<br>Del Paso et al. (2011; 58.2%); Del Paso et al. (2022; 41.8%) | CPP – widespread: 61 | - .04 | [- 0.29; 0.22] | - 0.28 | .78 | 0% |
| 2b | RRI during recovery | CPT<sub>o</sub> at baseline | N = 2<br>Del Paso et al. (2011; 58.2%); Del Paso et al. (2022; 41.8%) | CPP – widespread: 61 | .03 | [- 0.23; 0.28] | 0.19 | .85 | 0% |
| 3a | ΔHR (during stressor – at baseline) | PPT<sub>h</sub> at baseline | N = 2<br>Lopez-Lopez et al. (2021; 31.25%); Mohn et al. (2008; 68.75%) | CPP: 38 | - .01 | [- 0.6; 0.59] | - 0.04 | .97 | 70.5% |
| 3a | ΔHR (during stressor – at baseline) | PPT<sub>o</sub> at baseline | N = 2<br>Lopez-Lopez et al. (2021; 31.25%); Mohn et al. (2008; 68.75%) | CPP: 38 | .27 | [- 0.07; 0.56] | 1.59 | .11 | 0% |
| 3a | ΔHR (during stressor – at baseline) | EPT<sub>h</sub> at baseline | N = 2<br>Löffler et al. (2023; 46.34%); Mohn et al. (2008; 53.66%) | CPP – localized: 47 | .11 | [- 0.19; 0.4] | 0.72 | .47 | 0% |
|  |  |  | N = 2<br>Löffler et al. (2023; 40.54%); Mohn et al. (2008; 59.46%) | HC: 43 | .23 | [- 0.33; 0.67] | 0.79 | .43 | 67.82% |
| 3a | ΔHR (during stressor – at baseline) | EPT<sub>o</sub> at baseline | N = 2<br>Löffler et al. (2023; 46.34%); Mohn et al. (2008; 53.66%) | CPP – localized: 47 | .19 | [- 0.11; 0.46] | 1.23 | .22 | 0% |
|  |  |  | N = 2<br>Löffler et al. (2023; 40.54%); Mohn et al. (2008; 59.46%) | HC: 43 | .29 | [- 0.1; 0.61] | 1.47 | .14 | 34.77% |

Abbreviations. AUCg: Area Under the Curve (ground), BRS: Baroreflex Sensitivity, CPP: Chronic Primary Pain, CPT<sub>h</sub>: Cold Pain Threshold, CPT<sub>o</sub>: Cold Pain Tolerance, DBP: Diastolic Blood Pressure, EP<sub>Th</sub>: Electrical Pain Threshold, EP<sub>To</sub>: Electrical Pain Tolerance, HC: Healthy Controls, HF: High Frequency, HP<sub>Th</sub>: Heat Pain Threshold, HR: Heart Rate, HRV: Heart Rate Variability, LF: Low Frequency, MAP: Mean Arterial Pressure, PP<sub>Th</sub>: Pressure Pain Threshold, PP<sub>To</sub>: Pressure Pain Tolerance, RRI: R-R Interval, SBP: Systolic Blood Pressure, SC: Skin Conductance, $\Delta$ : change in.

#### 3.4.1 Baseline stress – pain (Cluster 1)

##### ANS: HR – Pain

Data of 13 studies was available concerning the association between HR and pain outcomes. One study reported a significant positive effect size for pain intensity during a CPT (*b* = .43; *p* = .04) (Chalaye, et al., 2014), one study reported a significant negative association with PPTh measured during a stressor (*r* = *-*.68*, p* = .01) (Lopez-Lopez, et al., 2021) and two studies reported a significant negative association with PPTh measured after a stressor (*r* = −.51; *r* = −.56, all *p* < .05) (Kadetoff & Kosek, 2007; Lopez-Lopez, et al., 2021). All other associations were non-significant (all *p* > .05).

**Meta-analyses** concerning the associations between HR and PPThs (at baseline, during a stressor, and after a stressor) (Kadetoff & Kosek, 2007, 2010; Lopez-Lopez, et al., 2021; Mohn, et al., 2008; Poli-Neto, et al., 2020; Valera-Calero & Varol, 2022; Zamunér, et al., 2016), EPThs at baseline (Loffler, et al., 2023; Mohn, et al., 2008; van Middendorp, et al., 2013), PPTos at baseline (Lopez-Lopez, et al., 2021; Mohn, et al., 2008) and EPTos at baseline (Loffler, et al., 2023; Mohn, et al., 2008; van Middendorp, et al., 2013) showed no significant results. When looking at the widespread (Kadetoff & Kosek, 2007, 2010; Lopez-Lopez, et al., 2021) and localized (Mohn, et al., 2008; Poli-Neto, et al., 2020; Valera-Calero & Varol, 2022) pain groups separately, there were also no significant associations between HR at baseline and PPTh at baseline. However, there was a significant negative association between HR at baseline and PPTh during a stressor (*r* = −.43; 95 % CI = [−0.7; - 0.06]) in widespread CPP (Kadetoff & Kosek, 2007, 2010; Lopez-Lopez, et al., 2021), indicating that higher HR at baseline was associated with lower PPTh during a stressor.

##### ANS: SBP – Pain

Eleven studies evaluated the association between Systolic BP (SBP) and static pain outcomes, and found no significant results (all *p* > .05) (de la Coba, et al., 2018; Kadetoff & Kosek, 2007, 2010; Loffler, et al., 2023; Lopez-Lopez, et al., 2021; Maixner, et al., 1997; Poli-Neto, et al., 2020; Reyes Del Paso, et al., 2022; Reyes del Paso, et al., 2011; Valera-Calero & Varol, 2022; Zamunér, et al., 2016). One study measured the interaction between SBP and SREP at baseline and found a significant negative association (*r* = −.55, *p* < .01) (de la Coba, et al., 2018).

**Meta-analyses** on the associations between SBP and PPTh (at baseline, during a stressor, and after a stressor) (de la Coba, et al., 2018; Kadetoff & Kosek, 2007, 2010; Lopez-Lopez, et al., 2021; Poli-Neto, et al., 2020; Valera-Calero & Varol, 2022; Zamunér, et al., 2016), cold PTh (CPTh) at baseline (Reyes Del Paso, et al., 2022; Reyes del Paso, et al., 2011), PPTo at baseline (de la Coba, et al., 2018; Lopez-Lopez, et al., 2021) and cold PTo (CPTo) at baseline (Reyes Del Paso, et al., 2022; Reyes del Paso, et al., 2011) all yielded no significant results. Zooming in on widespread (Kadetoff & Kosek, 2007, 2010; Lopez-Lopez, et al., 2021; Zamunér, et al., 2016) and localized (de la Coba, et al., 2018; Poli-Neto, et al., 2020; Valera-Calero & Varol, 2022) pain separately for the association with PPTh also did not yield any significant findings.

##### ANS: DBP – Pain

For diastolic BP (DBP), most studies showed no significant associations (de la Coba, et al., 2018; Kadetoff & Kosek, 2007, 2010; Loffler, et al., 2023; Poli-Neto, et al., 2020; Reyes Del Paso, et al., 2022; Reyes del Paso, et al., 2011; Valera-Calero & Varol, 2022; Zamunér, et al., 2016). One study analysed the association between DBP and SREP at baseline and found a negative significant association (*r* = −.46, *p* < .01) (de la Coba, et al., 2018).

**Meta-analyses** on the associations between DBP and PPTh (at baseline, during a stressor, and after a stressor) (de la Coba, et al., 2018; Kadetoff & Kosek, 2007, 2010; Poli-Neto, et al., 2020; Valera-Calero & Varol, 2022; Zamunér, et al., 2016), CPTh at baseline (Reyes Del Paso, et al., 2022; Reyes del Paso, et al., 2011) and CPTo at baseline (Reyes Del Paso, et al., 2022; Reyes del Paso, et al., 2011) also produced no significant results. Again, associations with PPTh in widespread (Kadetoff & Kosek, 2010; Zamunér, et al., 2016) or localized (de la Coba, et al., 2018; Poli-Neto, et al., 2020; Valera-Calero & Varol, 2022) pain specifically were also not significant.

##### ANS: MAP – Pain

When observing the interaction between mean arterial pressure (MAP, i.e. the mean arterial blood pressure during one cardiac cycle; MAP = DBP + 1/3(SBP – DBP)) and PPTh, EPTh, PPTo, EPTo and nociceptive flexion reflex threshold at baseline, two of the three studies found a significant positive correlation (*r* = .42 - .88, all *p* < .05) (Mohn, et al., 2008; Umeda, et al., 2013; van Middendorp, et al., 2013). **Meta-analyses** confirmed that there was a significant positive association between MAP and EPTh at baseline (*r* = .35; 95 % CI = [0.06; 0.6]) (Mohn, et al., 2008; van Middendorp, et al., 2013) and EPTo at baseline (*r* = .30; 95 % CI = [0.02; 0.53]) (Mohn, et al., 2008; van Middendorp, et al., 2013), indicating that lower MAP was associated with lower EPTh and EPTo. Subgroup analyses were not possible.

##### ANS: BRS – Pain

Three studies examining the interaction between BRS and CPTh, CPTo and CPM at baseline found no significant association (all *p* > .05) (Reyes Del Paso, et al., 2022; Reyes del Paso, et al., 2011; Venezia, et al., 2024).

**Meta-analyses** on associations between cardiac BRS and CPTh and CPTo at baseline also yielded no significant results (Reyes Del Paso, et al., 2022; Reyes del Paso, et al., 2011). Subgroup analyses were not possible.

##### ANS: HRV and PEP – Pain

Concerning parameters of HRV and PEP, no significant associations were found with both static and dynamic pain outcomes at baseline and after a stressor (all *p* > .05) across eight studies (Bandeira, et al., 2021; Cohen, et al., 2000; Jarrett, et al., 2016; Miyachi, et al., 2025; Reyes Del Paso, et al., 2022; Reyes del Paso, et al., 2011; Van Den Houte, et al., 2018; Zamunér, et al., 2016).

**Meta-analyses** on the association between HF and PPTh (Bandeira, et al., 2021; Zamunér, et al., 2016), CPTh and CPTo at baseline (Reyes Del Paso, et al., 2022; Reyes del Paso, et al., 2011) yielded no significant results. Meta-analyses on the association between LF and PPTh (Bandeira, et al., 2021; Zamunér, et al., 2016), CPTh, and CPTo at baseline (Reyes Del Paso, et al., 2022; Reyes del Paso, et al., 2011) also did not deliver any significant results. The meta-analysis of the association between LF/HF and PPTh at baseline also did not demonstrate a significant relationship (Bandeira, et al., 2021; Miyachi, et al., 2025; Zamunér, et al., 2016). Looking at localized pain specifically in this interaction also did not yield a significant result (Bandeira, et al., 2021; Miyachi, et al., 2025). Finally, meta-analyses were conducted on the association between RRI and CPTh and CPTo at baseline (Reyes Del Paso, et al., 2022; Reyes del Paso, et al., 2011), which both yielded no significant results.

##### ANS: EDA – Pain

For EDA, one study indicated a positive association between sympathetic skin response (SSR) latencies and PPThs at baseline (*r* = .46, p = .01) (Ozgocmen, et al., 2006) and one study indicated a significant negative association between SC and SREP at baseline (*r* = −.46, *p* < .05) (Garcia-Hernandez, et al., 2022). Two studies found no significant associations between SC and PPTh at baseline (Garcia-Hernandez, et al., 2022; Tan, et al., 2023), one study found no significant association with PPTo at baseline (Garcia-Hernandez, et al., 2022), and another study showed no significant association with CPM at baseline (*p* > .05) (Pickering, et al., 2020).

**Meta-analysis** on the association between SC at baseline and PPTh at baseline revealed no significant result (Garcia-Hernandez, et al., 2022; Tan, et al., 2023), and no other meta-analyses concerning EDA could be conducted.

##### ANS: (Nor)epinephrine – Pain

For both epinephrine and norepinephrine, one of two studies showed a significant positive correlation with PPTh (*r* = .13-.15, *p* < .05) (Reshkova, et al., 2015).

The **meta-analysis** on the association between epinephrine or norepinephrine at baseline and PPTh at baseline did not yield significant results.

##### HPA – Pain

Eight studies did not show a significant association between cortisol at baseline and pain measures at baseline, during a stressor or after a stressor (Davydov, et al., 2024; de Abreu Freitas, et al., 2012; Jarrett, et al., 2014; Kadetoff & Kosek, 2010; Meeus, et al., 2008; Muhtz, et al., 2013; Nees, et al., 2019; Wingenfeld, et al., 2010). One study showed a significant negative association between salivary cortisol measured between 10:00-10:30 and PPTh at baseline (*r* = −.26, *p* < .05) (Valera-Calero & Varol, 2022), and one study showed a significant positive association between AUCi of salivary cortisol with PPTh at baseline (*r* = .62, *p* = .02) (Nees, et al., 2019). One study assessed DHEA and showed no significant association with PPTh and PPTo at baseline (de Abreu Freitas, et al., 2012).

**Meta-analysis** on the association between salivary cortisol concentrations and heat pain ratings revealed no significant relationship (Meeus, et al., 2008; Muhtz, et al., 2013). The meta-analysis demonstrated a significant negative association between cortisol concentrations of a single measurement (blood and salivary grouped together; sampling in the morning) and PPTh (*r* = −.22; 95 % CI = [−0.37; - 0.05]) (de Abreu Freitas, et al., 2012; Kadetoff & Kosek, 2010; Nees, et al., 2019; Valera-Calero & Varol, 2022) at baseline, indicating that higher cortisol concentrations were associated with lower pain thresholds. However, when looking at widespread (de Abreu Freitas, et al., 2012; Kadetoff & Kosek, 2010) or localized (Nees, et al., 2019; Valera-Calero & Varol, 2022) pain specifically, the meta-analysis did not show a significant association. Finally, meta-analyses did not demonstrate significant associations between AUCg of salivary cortisol and HPTh or PPTh (Nees, et al., 2019; Wingenfeld, et al., 2010) at baseline.

**In summary**, qualitative analyses showed that lower MAP might be associated with higher pain sensitivity in participants with CPP which is based on very low to low certainty of evidence. Other interactions, also based on very low to low certainty of evidence, showed mixed results. **Quantitative meta-analyses** on the physiological stress markers assessed at baseline demonstrated that higher HR was associated with lower PPTh during a stressor in CPP, supported the qualitative finding that lower MAP was associated with lower EPTh and EPTo, and showed that higher cortisol levels were associated with lower PPThs. All significant associations are based on low certainty of evidence. Other associations did not show significance and were based on very low to low certainty levels of evidence. Meta-analyses yielded no significant associations in pain-free controls.

#### 3.4.2 Stress during or after a stressor – pain (Cluster 2a & 2b)

##### ANS: HR during stressor – Pain

One study showed a significant negative association between HR measured during a stressor (the TSST) and PPThs measured at baseline and during the stressor (*r* = −.58; *r* = .61, all *p* < .04), however, no significant associations were found with PPTos (all *p* > .05) (Lopez-Lopez, et al., 2021). Two other studies did not show significant associations with PPThs at baseline, during a stressor or after a stressor (Kadetoff & Kosek, 2007, 2010).

**Meta-analyses** demonstrated a significant negative association between HR during a stressor and PPTh at baseline (*r* = −.38; 95 % CI = [−0.61; - 0.1]) (Kadetoff & Kosek, 2007, 2010; Lopez-Lopez, et al., 2021), no significant association with PPTh during a stressor, and a significant negative association with PPTh after the stressor (*r* = −.36; 95 % CI = [−0.59; - 0.07]). This indicates that a higher HR during a stressor is associated with lower PPThs at baseline and after a stressor.

##### ANS: SBP, DBP and BRS during stressor – Pain

Two and three studies measured DBP and SBP, respectively, during a stressor (hot water and cold water immersion, and during an arithmetic task) and found no significant association with CPThs and CPTos which were also measured during the CPT (*p* > .05) (Lopez-Lopez, et al., 2021; Reyes Del Paso, et al., 2022; Reyes del Paso, et al., 2011). One of two studies did find a significant negative association between SBP and PPTh, only when pain was measured after the stressor (*r* = −.51, *p* = .04) (Kadetoff & Kosek, 2007, 2010). Cardiac BRS was significantly positively associated with CPTh and CPTo, only when BRS was measured during the arithmetic task (*r* = .30 – .31, all *p* < .03) (Reyes Del Paso, et al., 2022).

**Meta-analyses** did not show a significant association between SBP during a stressor and PPTh (at baseline, during a stressor and during recovery) (Kadetoff & Kosek, 2007, 2010; Lopez-Lopez, et al., 2021), CPTh at baseline and CPTo at baseline (Reyes Del Paso, et al., 2022; Reyes del Paso, et al., 2011). Similarly, meta-analyses on the association between DBP during a stressor and PPTh (at baseline, during a stressor and during recovery) (Kadetoff & Kosek, 2007, 2010; Lopez-Lopez, et al., 2021), CPTh and CPTo at baseline (Reyes Del Paso, et al., 2022; Reyes del Paso, et al., 2011) did not demonstrate a significant relationship.

Additionally, cardiac BRS during a stressor was not significantly associated with either CPTh or CPTo at baseline (Reyes Del Paso, et al., 2022; Reyes del Paso, et al., 2011).

##### ANS: HRV during stressor – Pain

Three studies measured parameters of HRV during a stressor and found no significant associations with CPTh and CPTo at baseline and after the stressor (Bandeira, et al., 2021; Reyes Del Paso, et al., 2022; Reyes del Paso, et al., 2011).

**Meta-analyses** demonstrated no significant association between HF during a stressor and CPTh or CPTo at baseline (Reyes Del Paso, et al., 2022; Reyes del Paso, et al., 2011). Meta-analyses also did not show a significant association between LF during a stressor and CPTh or CPTo at baseline (Reyes Del Paso, et al., 2022; Reyes del Paso, et al., 2011). RRI during a stressor was also not significantly associated with either CPTh or CPTo at baseline (Reyes Del Paso, et al., 2022; Reyes del Paso, et al., 2011).

##### ANS: EDA during stressor – Pain

One study measured SSR elicited by heat stimuli and reported no significant association between SSR and heat pain ratings obtained during the stimulus application (*p* > .05) (Scheuren, et al., 2023).

##### ANS: HR, SBP, DBP, BRS and HRV during recovery – Pain

Most physiological parameters of the ANS assessed during recovery showed no significant association with experimental pain outcomes (*p* > .05) (Lopez-Lopez, et al., 2021; Reyes Del Paso, et al., 2022; Reyes del Paso, et al., 2011), however, two of five studies showed a significant negative association between HR during recovery and PPTh at baseline and during a stressor (*r* = −.67-.70, all *p* < .01) (de Bruijn, et al., 2011; Kadetoff & Kosek, 2007, 2010; Lopez-Lopez, et al., 2021; Mohn, et al., 2008), one study showed a significant positive association between HR during recovery and CPTh at baseline (*p* = .004) (de Bruijn, et al., 2011) and HR during recovery was significantly positively associated with thermal WUR at baseline in one study (*p* = .01) (de Bruijn, et al., 2011).

**Meta-analyses** confirmed that there was a significant negative association between HR during recovery and PPTh (de Bruijn, et al., 2011; Kadetoff & Kosek, 2007, 2010; Lopez-Lopez, et al., 2021; Mohn, et al., 2008). This relationship was observed for PPTh at baseline (*r* = −.40; 95 % CI = [−0.68; - 0.05]), during a stressor (*r* = −.53; 95 % CI = [−0.71; - 0.27]) and during recovery (*r* = −.48; 95 % CI = [−0.68; - 0.22]). This indicates that a higher HR during recovery is associated with lower PPThs before, during and after a stressor. Notably, the correlation values were larger when focusing on widespread pain (de Bruijn, et al., 2011; Kadetoff & Kosek, 2007, 2010; Lopez-Lopez, et al., 2021) for PPTh at baseline (*r* = −.60; 95 % CI = [−0.71; - 0.34]). A sub-analysis for localized CPP was not possible. Additionally, meta-analysis did not demonstrate a significant association between HR at recovery and PPTo.

Meta-analyses on the associations between SBP during recovery and PPTh (either at baseline, during a stressor or during recovery) (Kadetoff & Kosek, 2007, 2010; Lopez-Lopez, et al., 2021), CPTh and CPTo at baseline (Reyes Del Paso, et al., 2022; Reyes del Paso, et al., 2011) yielded no significant results.

Similarly, there were no significant results for meta-analyses on the associations between DBP during recovery and PPTh (either at baseline, during a stressor or after a stressor) (Kadetoff & Kosek, 2007, 2010; Lopez-Lopez, et al., 2021), CPTh and CPTo at baseline (Reyes Del Paso, et al., 2022; Reyes del Paso, et al., 2011) in CPP.

Meta-analyses did not reveal any significant results on the associations between cardiac BRS during recovery and CPTh or CPTo at baseline (Reyes Del Paso, et al., 2022; Reyes del Paso, et al., 2011) in CPP. Meta-analyses were also conducted for LF and HF HRV parameters during recovery. There was a significant positive association between HF during recovery and CPTo at baseline (*r* = .31; 95 % CI = [0.06; 0.53]), indicating that lower HF during recovery was associated with lower CPTo at baseline, but this was not the case for the association with CPTh at baseline (Reyes Del Paso, et al., 2022; Reyes del Paso, et al., 2011). Meta-analyses yielded no significant results for the associations between LF during recovery and CPTh or CPTo at baseline (Reyes Del Paso, et al., 2022; Reyes del Paso, et al., 2011) in CPP.

Finally, meta-analyses on the associations between RRI during recovery and CPTh and CPTo at baseline (Reyes Del Paso, et al., 2022; Reyes del Paso, et al., 2011) did not demonstrate any significant results.

##### HPA during stressor – Pain

No significant association was found between cortisol levels during a stressor and PPTh at baseline, during a stressor or after a stressor, heat pain ratings at baseline and CPM with heat stimuli at baseline (all *p* > .05) (Jarrett, et al., 2014; Kadetoff & Kosek, 2010; Meeus, et al., 2008).

##### HPA during recovery – Pain

Only one study evaluated cortisol levels during recovery in relation to PPTh at baseline, during a stressor or after a stressor, and found no significant results (all *p* > .05) (Kadetoff & Kosek, 2010).

**In summary**, qualitative analyses showed no conclusive associations between stress and pain which were based on very low to low certainty of evidence. **Quantitative meta-analyses** on physiological stress markers assessed in response to a stressor showed that higher HR measured during a stressor was associated with lower PPThs at baseline (in widespread CPP) and after a stressor (in CPP and HC). It also showed that higher HR during recovery was associated with lower PPTh at baseline, during a stressor and after a stressor in CPP, and that lower HF-HRV measured during recovery was associated with lower CPTo at baseline in participants with CPP. These significant relationships are based on moderate certainty of evidence. Other interactions did not show significance and were based on very low to low levels of evidence.

#### 3.4.3 Change in stress – pain (Cluster 3a & 3b)

##### ANS reactivity: HR, SBP and DBP – pain

Across three studies, reactivity of HR and SBP (change from baseline to during stressor) did not significantly correlate with PPThs, EPThs, PPTos and EPTos at baseline, during a stressor or after a stressor (Loffler, et al., 2023; Lopez-Lopez, et al., 2021; Mohn, et al., 2008). Reactivity of DBP (change from baseline to during stressor), based on one study, showed a significant positive association with EPTo at baseline (*r* = .44, *p* = .04) (Loffler, et al., 2023).

**Meta-analyses** on the association between the change in HR from baseline to during a stressor and PPTh at baseline (Lopez-Lopez, et al., 2021; Mohn, et al., 2008), PPTo at baseline (Lopez-Lopez, et al., 2021; Mohn, et al., 2008), EPTh (Loffler, et al., 2023; Mohn, et al., 2008) or EPTo at baseline (Loffler, et al., 2023; Mohn, et al., 2008) yielded no significant results.

##### ANS reactivity: MAP – pain

Reactivity of MAP (change from baseline to during stressor) only showed a significant positive association with PPTo (*r* = .10, *p* < .05), but not with PPTh, EPTh, or EPTo (all *p* > .05) (Mohn, et al., 2008).

##### ANS reactivity: HRV – pain

Reactivity of HF-HRV (change from baseline to during stressor) is the only HRV outcome to show a significant negative association with a pain outcome, more specifically with TSP during cuff pain (*r* = −.50, *p* < .01) (J. Kim, et al., 2015). Associations between HF-HRV and PPTh showed no significant associations (all *p* > .05) (Bandeira, et al., 2021).

##### ANS recovery: HR, SBP, DBP and HRV – pain

No significant associations were found between recovery in HR, SBP, DBP, HRV (change from baseline to zero to five minutes, or five to 10 minutes recording after a stressor) and PPTh, EPTh, PPTo or EPTo at baseline, during a stressor or after a stressor (all *p* > .50) (Bandeira, et al., 2021; Lopez-Lopez, et al., 2021).

##### HPA reactivity – pain

A significant positive association was found between cortisol reactivity (change from baseline to immediately after CPM procedure) and heat pain ratings during a CPM procedure, only for a descending protocol (*r* = .67, *p* = 0.006) and not for the ascending protocol (*p* > .05) (Meeus, et al., 2008).

**In summary**, qualitative analyses showed no conclusive associations between markers of stress reactivity and recovery, and pain, which were based on very low to low certainty of evidence. Only few **quantitative meta-analyses** were possible, and showed no significant associations based on very low certainty of evidence. Meta-analyses also yielded no significant associations in pain-free controls.

#### 3.4.4 Stress – change in pain (Cluster 4)

##### ANS: HR, SBP, DBP and HRV – change in pain

From three studies, data was gathered concerning the association between physiological parameters of the ANS at baseline, reactivity or recovery, and changes in pain outcomes from baseline to during or after a stressor (Bandeira, et al., 2021; Loffler, et al., 2023; Lopez-Lopez, et al., 2021). None of the stress parameters, including HR, SBP, DBP, LF, HF and LF/HF was significantly associated with PPThs, EPThs, PPTos and EPTos (all *p* > .05).

**In summary**, qualitative analyses showed no conclusive associations between stress and changes in pain which were based on very low to low certainty of evidence. No meta-analyses were possible.

#### 3.4.5 Change in stress and pain (Cluster 5)

##### ANS reactivity: HR, SBP, DBP, HRV, PEP and EDA – change in pain

Of four studies, associations were gathered concerning reactivity of HR, SBP, DBP, LF, HF, LF/HF, PEP and EDA (change from baseline to during stressor) and PPTh, PPTo, EPTh, EPTo (change from baseline to during stressor, or change from baseline to after stressor) (Bandeira, et al., 2021; Bossenger, et al., 2023; Loffler, et al., 2023; Lopez-Lopez, et al., 2021). Only changes in DBP from baseline to during a stressor showed a significant negative association with changes in EPTo from baseline to after a stressor (*r* = −.58, *p* = .01), but not with EPTh (*p* > .05) in a singular study (Loffler, et al., 2023).

##### ANS recovery: HR, SBP and HRV – change in pain

Based on two studies, no significant associations were found between HR, SBP, LF, HF and LF/HF (change from baseline to zero to five minutes, or five to 10 minutes recording after a stressor), and PPTh and PPTo (change from baseline to during stressor or after stressor) (all *p* > .05) (Bandeira, et al., 2021; Lopez-Lopez, et al., 2021).

**In summary**, qualitative analyses showed no conclusive associations between changes in both stress and pain outcomes which were based on very low to low certainty of evidence. No meta-analyses were possible.

## 4. DISCUSSION

This systematic review and meta-analysis investigated associations between physiological markers of stress response system functioning and static and dynamic experimental outcome measures of pain in patients with CPP to gain insight into the stress-pain relationship and the employed methodology to study the topic thus far. Stress response system functioning and its interaction with pain were investigated considering the different timings in assessments (at rest or baseline, during a stressor and during recovery from a stressor). A subdivision between chronic localized and chronic widespread pain groups, and comparisons with pain-free controls (see Supplementary File 10), were made where possible. Key findings are that: a) The vast majority of studies (**Cluster 1**) reported associations between stress markers measured *at baseline* and pain assessments *at baseline* (i.e., prior to or in absence of any stress induction), and of these studies only lower MAP *at baseline* was associated with higher pain sensitivity (lower EPTh and EPTo) *at baseline* in participants with CPP (observed in both the qualitative and quantitative analyses); b) A higher HR *at baseline* was associated with higher pain sensitivity (lower PPTh) measured *during a stressor*, and higher cortisol levels *at baseline* were associated with higher pain sensitivity (lower PPTh) *at baseline* in individuals with CPP (observed in the quantitative analyses of **Cluster 1**); c) For stress markers assessed in response to a stressor (i.e., *during or after* stress induction) (**Cluster 2a & 2b**), qualitative analyses showed no conclusive associations between stress and pain. However, quantitative analyses showed that higher HR measured *during and after a stressor* was associated with higher pain sensitivity (lower PPTh) measured *at baseline, during, and following stress induction* in participants with CPP; and lower HF-HRV measured *during recovery* was associated between with higher pain sensitivity (lower CPTo) measured *at baseline* in participants with CPP; d) Only few studies examined associations between changes in stress markers and/or changes in pain assessments (**Clusters 3, 4 & 5**), and of these none of the quantitative analyses yielded significant associations; e) Meta-analyses in pain-free controls only demonstrated one significant association between higher HR measured *during a stressor* and higher pain sensitivity (lower PPTh) measured *following stress induction*, similar as in CPP participants, while no other associations were observed; f) Subgroup analyses on the basis of pain extent could seldom be conducted and revealed no significant differences between widespread and localized pain groups.

Together, these findings show that there is clear evidence of associations between physiological stress markers and experimental pain outcomes in individuals with CPP, implicating both the ANS—or more specifically the cardiovascular system—as well as the HPA axis. However, more research is still needed to clarify the role of pain extent and to strengthen comparisons with pain-free controls.

### 4.1 Cardiovascular functioning and its association with pain

A total of 92 meta-analyses were conducted in CPP populations, of which eleven yielded significant associations and hence the vast majority was indicative of a lack of associations (or a lack of power). It was noticeable that most of these associations were with cardiovascular parameters (i.e., related to HR or BP), which can be explained through multiple frameworks.

First, the observed association between lower MAP and higher pain sensitivity in participants with CPP could indicate a role for baroreceptor regulation in the body’s ability to modulate pain. Typically, under acute stress, an increase in blood pressure activates arterial baroreceptors, which send signals to the brainstem to lower the pressure by decreasing HR and triggering vasodilation (Duschek, et al., 2013). This in turn can trigger spinal inhibitory neurons via the periaqueductal gray (PAG) and rostral ventromedial medulla (RVM), leading to hypoalgesia (i.e., reduced pain sensitivity). In pain-free individuals, this BP-related hypoalgesia is well-documented (Chalaye, et al., 2013; Makovac, et al., 2020) and it is hypothesized that this relationship occurs as an adaptive homeostatic feedback mechanism in acute stress or pain conditions to control the nociceptive response (Bruehl & Chung, 2004; Sacco, et al., 2013). Our finding is in line with previous studies showing that this mechanism is diminished in patients with chronic pain (Duschek, et al., 2009; Olsen, et al., 2013; Sacco, et al., 2013), with a low baroreceptor activity being accompanied by a reduction in central pain inhibition (Bruehl & Chung, 2004). Although this finding could underscore the importance of baroreflex functioning in chronic pain, it is worth noting that MAP—a composite measure derived from both SBP and DBP—showed associations with experimental pain, whereas meta-analyses did not reveal significant associations for SBP or DBP considered separately.

Second, the association between cardiovascular stress parameters and experimental pain outcomes can be conceptualized through dysregulation of the ANS. In this study, higher HR at baseline, during a stressor and following a stressor (during recovery) was associated with higher pain sensitivity (lower PPTh) measured during a stressor and at baseline in CPP. Furthermore, lower HF-HRV measured over a five-minute recovery period was associated with higher pain sensitivity (lower CPTo), which may also indicate that there is a relationship between higher pain sensitivity and reduced parasympathetic reactivation following stress induction (Tiwari, et al., 2021).

Together, these cardiac findings in CPP could be indicative of higher pain sensitivity being related to an overall increased activity in the sympathetic nervous system and/or reduced parasympathetic tone. This pattern of sympathetic dominance—captured by reduced HRV (Tracy, et al., 2016) and other ANS markers (Raison, 2009)—has been systematically observed in chronic pain, and our findings extend this by also showing its relevance in increased pain sensitivity. The causal mechanisms remain insufficiently understood, although overlapping neural networks have been hypothesized to mediate the sympathetic-nociceptor interactions (Benarroch, 2006). Again, this explanation must be nuanced by the absence of significant associations between pain sensitivity and other markers of sympathetic dominance in this meta-analysis (e.g., SBP & DBP).

### 4.2 HPA axis functioning and its association with pain

Although basal cortisol levels can be either elevated or reduced in CPP (Woda, et al., 2016), when increased, they appear to be associated with greater pain sensitivity (lower PPThs), reflecting dysregulation of the HPA axis. Indeed, a previous study demonstrated that baseline cortisol levels were associated with momentary pain ratings in patients with fibromyalgia (McLean, et al., 2005). However, this association was only present during the early morning and not throughout the rest of the day, while the included studies in this meta-analytic finding sampled from early morning to early afternoon and thus expanded on the previous research. Even though stress-induced analgesia (i.e., reduction in pain sensitivity) through cortisol reactivity has been observed in pain-free populations (Timmers, et al., 2018), there were no studies available in this systematic review to shed light on this phenomenon in CPP. In fact, there were virtually no studies examining how changes in physiological stress markers were related to pain outcomes, nor how changes in pain outcomes were related to physiological stress markers, illustrating a clear gap in the existing literature. Therefore, more research is needed on HPA axis functioning in CPP in relation to experimental pain outcomes during and after a stressor, instead of merely at baseline.

### 4.3 Other considerations

Our review specifically focused on synthesizing the evidence on associations between physiological stress markers and experimental pain outcomes. Besides physiological aspects of stress-pain interactions, though, neural and psychosocial factors also contribute to (inter)individual differences in stress and pain responses (Chida & Hamer, 2008; Hirsh, et al., 2008; Simon, et al., 2022), and hence these should also be considered. For instance, evidence suggests that interactions between frontal cortex activity, descending pain inhibition, and ANS responses may underlie symptoms in chronic pain (Makovac, et al., 2021), while ANS activity has also been shown to influence brain connectivity during pain, linking LF-HRV with functional coupling between the dorsal anterior cingulate cortex and the PAG (Hohenschurz-Schmidt, et al., 2020). At the psychosocial level, pain appraisal (i.e., how individuals interpret and evaluate the threat value of pain) has been found to mediate the pain-induced autonomic response (Mischkowski, et al., 2018). Interestingly, in contrast to the findings in CPP, pain-free individuals showed only one association between stress and pain (Kadetoff & Kosek, 2007, 2010; Lopez-Lopez, et al., 2021). One explanation may be that, in healthy populations, stress and pain appraisals are more mixed, with stress sometimes perceived as challenging and other times as threatening, which could lead to more variable stress–pain associations. In CPP, by contrast, appraisals are often predominantly negative and pain-related threat processing more pronounced, resulting in more consistent associations. Chronic exposure to stress and pain may thus progressively entangle these systems, leading to the dysregulation observed in CPP. Future research should examine this continuum across different durations and stages of pain to clarify when and how such dysregulation develops, and doing so will require incorporating both neural and psychosocial characteristics of stress–pain interactions to achieve a more comprehensive understanding of how ANS and HPA axis functioning relate to pain.

### 4.4 Strengths, limitations and future perspectives

This review provides the first comprehensive overview of associations between physiological stress system functioning and experimentally induced pain, revealing a clear link between the two in CPP. An extensive search strategy was employed, and authors were contacted to ensure that all relevant data could be included. The findings should, however, be considered in light of the existing limitations. For instance, it was not possible to conduct quantitative analyses for every potential interaction due to limited data availability. Specifically, associations with dynamic pain outcomes or stress-induced changes were lacking. Based on the current findings, it was also not possible to determine directionality or establish whether altered stress responses drive changes in pain sensitivity or are a consequence of chronic pain. Furthermore, a wide range of measuring methods and hardware, pain assessments of different modalities (stimulating distinct pathways) and at various body sites, different analytical methods (Beiner, et al., 2023) and a variety of different stressors (Brunyé, et al., 2025) were used, complicating interpretation of our findings. A large variety of instructions (e.g., medication use) and eligibility criteria (e.g., psychological conditions) were employed, which are representative of the clinical reality, but also make it difficult to exclude external influences. Furthermore, not every author could provide data, which limited the qualitative analyses and the possibility to perform quantitative analyses. Finally, the overall risk of bias was fairly low, but the level of evidence was low (ranging from very low to moderate), and hence the results should be interpreted with caution.

This study highlights the need for more comprehensive, holistic and methodologically standardized approaches to study the relationship between physiological markers of stress and experimental pain outcomes. Future studies should explore how stress system dysregulation emerges and progresses across different stages and durations of pain (e.g., comparing chronic widespread and chronic localized pain groups), while also addressing questions of directionality to clarify whether dysregulation precedes or follows the persistence of pain. Beyond physiological factors, research should also incorporate neural and psychosocial characteristics of stress–pain interactions to achieve a more complete understanding of how ANS and HPA axis functioning relate to pain. By advancing in these directions, the level of evidence can be increased and a variety of hypotheses on stress system dysregulations can be tested, ultimately informing therapeutic guidelines for CPP.

## 5. CONCLUSION

This systematic review and meta-analysis of the literature on the association between physiological markers of stress response system functioning and experimental pain outcomes in CPP populations provides support for relationships between higher pain sensitivity and higher MAP, higher HR, lower HF-HRV and higher cortisol levels. These findings suggest that dysfunction of the ANS and HPA axis contributes to the heightened pain sensitivity, or vice versa, observed in chronic pain. However, the level of evidence remains low due to methodological heterogeneity and comprehensive approaches are recommended, including studies that integrate diverse physiological stress markers and pain measurements to provide insights into underlying pain mechanisms at different timepoints and across the pain continuum.

## Supporting information

Supplementary 1 - PRISMA_checklist

Supplementary 10 - Results

## AUTHOR CONTRIBUTIONS

Conceptualization: Inge Timmers, Iris Coppieters, Jessica Van Oosterwijck, Joren Vyverman, Matthijs Moerkerke, Robrecht De Baere

Methodology: Jessica Van Oosterwijck, Joren Vyverman, Matthijs Moerkerke, Robrecht De Baere. Data curation: Joren Vyverman, Matthijs Moerkerke, Robrecht De Baere.

Investigation: Joren Vyverman, Matthijs Moerkerke, Robrecht De Baere.

Supervision: Inge Timmers, Iris Coppieters, Jessica Van Oosterwijck, Matthijs Moerkerke. Writing – original draft: Joren Vyverman, Robrecht De Baere.

Writing – Review & editing: Inge Timmers, Iris Coppieters, Jessica Van Oosterwijck, Joren Vyverman, Matthijs Moerkerke, Robrecht De Baere.

All authors contributed to and have approved the final version of the manuscript.

## APPENDICES. Supplementary materials

Supplementary Files 1-10 are available.

## Data Availability

All data produced in the present work are contained in the manuscript

---

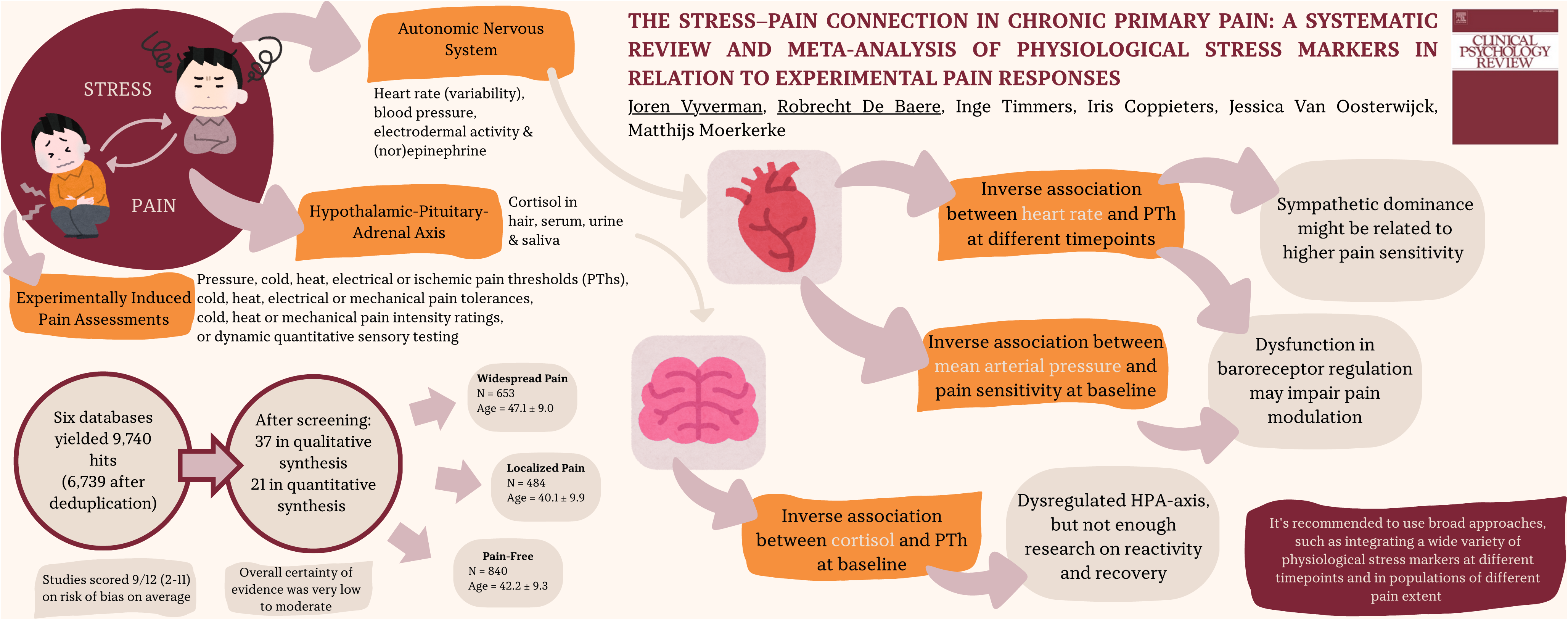

## REFERENCES

Abdallah, C. G., & Geha, P. (2017). Chronic Pain and Chronic Stress: Two Sides of the Same Coin? Chronic Stress 1.

al’Absi, M., & Petersen, K. L. (2003). Blood pressure but not cortisol mediates stress effects on subsequent pain perception in healthy men and women. Pain, 106, 285–95.

Amiri, M., Alavinia, M., Singh, M., & Kumbhare, D. (2021). Pressure Pain Threshold in Patients With Chronic Pain. American Journal of Physical Medicine & Rehabilitation, 100, 656–74.

Balshem, H., Helfand, M., Schünemann, H. J., Oxman, A. D., Kunz, R., Brozek, J., Vist, G. E., Falck-Ytter, Y., Meerpohl, J., & Norris, S. (2011). GRADE guidelines: 3. Rating the quality of evidence. Journal of Clinical Epidemiology, 64, 401–06.

Bandeira, P. M., Reis, F. J. J., Muniz, F. D. N., Chaves, A. C. S., Fernandes, O., Jr., & Arruda-Sanchez, T. (2021). Heart Rate Variability and Pain Sensitivity in Chronic Low Back Pain Patients Exposed to Passive Viewing of Photographs of Daily Activities. The Clinical Journal of Pain 37, 591–97.

Beiner, E., Lucas, V., Reichert, J., Buhai, D.V., Jesinghaus, M., Vock, S., Drusko, A., Baumeister, D., Eich, W., & Friederich, H.C. (2023). Stress biomarkers in individuals with fibromyalgia syndrome: a systematic review with meta-analysis. Pain, 164, 1416–27.

Benarroch, E. (2006). Pain-autonomic interactions. Neurological sciences, 27, s130–s33.

Bossenger, N. R., Lewis, G. N., Rice, D. A., & Shepherd, D. (2023). The autonomic and nociceptive response to acute exercise is impaired in people with knee osteoarthritis. Neurobiology of Pain 13, 100118.

Bruehl, S., & Chung, O. Y. (2004). Interactions between the cardiovascular and pain regulatory systems: an updated review of mechanisms and possible alterations in chronic pain. Neuroscience & Biobehavioral Reviews, 28, 395–414.

Brunyé, T. T., Goring, S. A., Navarro, E., Hart-Pomerantz, H., Grekin, S., McKinlay, A. M., & Plessow, F. (2025). Identifying the most effective acute stress induction methods for producing SAM-and HPA-related physiological responses: a meta-analysis. *Anxiety, Stress*, & Coping, 1–23.

Cardinali, D. P. (2018). Autonomic nervous system. Springer International Publishing, 10, 978–3.

Chalaye, P., Devoize, L., Lafrenaye, S., Dallel, R., & Marchand, S. (2013). Cardiovascular influences on conditioned pain modulation. Pain, 154, 1377–82.

Chalaye, P., Lafrenaye, S., Goffaux, P., & Marchand, S. (2014). The role of cardiovascular activity in fibromyalgia and conditioned pain modulation. Pain, 155, 1064–69.

Chida, Y., & Hamer, M. (2008). Chronic psychosocial factors and acute physiological responses to laboratory-induced stress in healthy populations: a quantitative review of 30 years of investigations. Psychological Bulletin 134, 829.

Cohen, H., Neumann, L., Shore, M., Amir, M., Cassuto, Y., & Buskila, D. (2000). Autonomic dysfunction in patients with fibromyalgia: application of power spectral analysis of heart rate variability. Seminars in Arthritis & Rheumatism, 29, 217–27.

Coppens, E., Kempke, S., Van Wambeke, P., Claes, S., Morlion, B., Luyten, P., & Van Oudenhove, L. (2018). Cortisol and subjective stress responses to acute psychosocial stress in fibromyalgia patients and control participants. Psychosomatic Medicine, 80, 317–26.

Crettaz, B., Marziniak, M., Willeke, P., Young, P., Hellhammer, D., Stumpf, A., & Burgmer, M. (2013). Stress-induced allodynia--evidence of increased pain sensitivity in healthy humans and patients with chronic pain after experimentally induced psychosocial stress. PLoS One, 8, e69460.

Davydov, D. M., de la Coba, P., Contreras-Merino, A. M., & Reyes Del Paso, G. A. (2024). Impact of homeostatic body hydration status, evaluated by hemodynamic measures, on different pain sensitization paths to a chronic pain syndrome. Scientific Reports 14, 1908.

de Abreu Freitas, R. P., Lemos, T. M. A. M., Spyrides, M. H. C., & de Sousa, M. B. C. (2012). Influence of cortisol and DHEA-S on pain and other symptoms in post menopausal women with fibromyalgia. Journal of Back and Musculoskeletal Rehabilitation, 25, 245–52.

de Bruijn, S. T., van Wijck, A. J., Geenen, R., Snijders, T. J., van der Meulen, W. J., Jacobs, J. W., & Veldhuijzen, D. S. (2011). Relevance of physical fitness levels and exercise-related beliefs for self-reported and experimental pain in fibromyalgia: an explorative study. JCR: Journal of Clinical Rheumatology 17, 295–301.

de la Coba, P., Bruehl, S., Duschek, S., & Reyes Del Paso, G. A. (2018). Blood pressure-related pain modulation in fibromyalgia: Differentiating between static versus dynamic pain indicators. International Journal of Psychophysiology 134, 79–85.

de Paiva Tosato, J., Caria, P. H. F., de Paula Gomes, C. A. F., Berzin, F., Politti, F., de Oliveira Gonzalez, T., & Biasotto-Gonzalez, D. A. (2015). Correlation of stress and muscle activity of patients with different degrees of temporomandibular disorder. Journal of physical therapy science, 27, 1227–31.

Dhondt, E., Van Oosterwijck, S., Coppieters, I., Danneels, L., & Van Oosterwijck, J. (2019). Effects of conditioned pain modulation on the nociceptive flexion reflex in healthy people: A systematic review. The Clinical journal of pain, 35, 794–807.

Duschek, S., Dietel, A., Schandry, R., & del Paso, G. A. R. (2009). Increased sensitivity to heat pain in chronic low blood pressure. European Journal of Pain, 13, 28–34.

Duschek, S., Werner, N. S., & Reyes Del Paso, G. A. (2013). The behavioral impact of baroreflex function: a review. Psychophysiology, 50, 1183–93.

Garcia-Hernandez, A., de la Coba, P., & del Paso, G. A. R. (2022). Central sensitisation to pain and autonomic deficiencies in fibromyalgia. Clinical and Experimental Rheumatology 40, 1202–09.

Gera, O., Ginzburg, K., Gur, N., & Defrin, R. (2025). Effects of acute stress exposure on pain sensitivity: the role of individual stress responsiveness and orientation to pain and stress. Pain, 10.1097.

Hannibal, K. E., & Bishop, M. D. (2014). Chronic stress, cortisol dysfunction, and pain: a psychoneuroendocrine rationale for stress management in pain rehabilitation. Physical Therapy 94, 1816–25.

Herman, J. P., McKlveen, J. M., Ghosal, S., Kopp, B., Wulsin, A., Makinson, R., Scheimann, J., & Myers, B. (2016). Regulation of the Hypothalamic-Pituitary-Adrenocortical Stress Response. Comprehensive Physiology 6, 603–21.

Herzog, R., Álvarez-Pasquin, M. J., Díaz, C., Del Barrio, J. L., Estrada, J. M., & Gil, Á. (2013). Are healthcare workers’ intentions to vaccinate related to their knowledge, beliefs and attitudes? A systematic review. BMC public health, 13, 1–17.

Hirsh, A. T., George, S. Z., Bialosky, J. E., & Robinson, M. E. (2008). Fear of pain, pain catastrophizing, and acute pain perception: relative prediction and timing of assessment. The Journal of Pain 9, 806–12.

Hohenschurz-Schmidt, D. J., Calcagnini, G., Dipasquale, O., Jackson, J. B., Medina, S., O’Daly, O., O’Muircheartaigh, J., de Lara Rubio, A., Williams, S. C. R., McMahon, S. B., Makovac, E., & Howard, M. A. (2020). Linking Pain Sensation to the Autonomic Nervous System: The Role of the Anterior Cingulate and Periaqueductal Gray Resting-State Networks. Frontiers in Neuroscience 14, 147.

Jarrett, M. E., Han, C. J., Cain, K. C., Burr, R. L., Shulman, R. J., Barney, P. G., Naliboff, B. D., Zia, J., & Heitkemper, M. M. (2016). Relationships of abdominal pain, reports to visceral and temperature pain sensitivity, conditioned pain modulation, and heart rate variability in irritable bowel syndrome. Neurogastroenterology & Motility 28, 1094–103.

Jarrett, M. E., Shulman, R. J., Cain, K. C., Deechakawan, W., Smith, L. T., Richebé, P., Eugenio, M., & Heitkemper, M. M. (2014). Conditioned Pain Modulation in Women With Irritable Bowel Syndrome. Biological Research for Nursing, 16, 368–77.

Kadetoff, D., & Kosek, E. (2007). The effects of static muscular contraction on blood pressure, heart rate, pain ratings and pressure pain thresholds in healthy individuals and patients with fibromyalgia. European Journal of Pain, 11, 39–47.

Kadetoff, D., & Kosek, E. (2010). Evidence of reduced sympatho-adrenal and hypothalamic-pituitary activity during static muscular work in patients with fibromyalgia. Journal of Rehabilitation Medicine 42, 765–72.

Kim, H. I., Kim, Y. Y., Chang, J. Y., Ko, J. Y., & Kho, H. S. (2012). Salivary cortisol, 17β-estradiol, progesterone, dehydroepiandrosterone, and α-amylase in patients with burning mouth syndrome. Oral Diseases, 18, 613–20.

Kim, J., Loggia, M. L., Cahalan, C. M., Harris, R. E., Beissner, F. D. P. N., Garcia, R. G., Kim, H., Wasan, A. D., Edwards, R. R., & Napadow, V. (2015). The somatosensory link in fibromyalgia: functional connectivity of the primary somatosensory cortex is altered by sustained pain and is associated with clinical/autonomic dysfunction. Arthritis & Rheumatology 67, 1395–405.

Koenig, J., Loerbroks, A., Jarczok, M. N., Fischer, J. E., & Thayer, J. F. (2016). Chronic pain and heart rate variability in a cross-sectional occupational sample: evidence for impaired vagal control. The Clinical journal of pain, 32, 218–25.

Liberati, A., Altman, D. G., Tetzlaff, J., Mulrow, C., Gøtzsche, P. C., Ioannidis, J. P., Clarke, M., Devereaux, P. J., Kleijnen, J., & Moher, D. (2009). The PRISMA statement for reporting systematic reviews and meta-analyses of studies that evaluate health care interventions: explanation and elaboration. Journal of Clinical Epidemiology 62, e1–34.

Loffler, M., Schneider, P., Schuh-Hofer, S., Kamping, S., Usai, K., Treede, R. D., Nees, F., & Flor, H. (2023). Stress-induced hyperalgesia instead of analgesia in patients with chronic musculoskeletal pain. Neurobiology of Pain 13, 100110.

Lopez-Lopez, A., Matias-Pompa, B., Fernandez-Carnero, J., Gil-Martinez, A., Alonso-Fernandez, M., Alonso Perez, J. L., & Gonzalez Gutierrez, J. L. (2021). Blunted Pain Modulation Response to Induced Stress in Women with Fibromyalgia with and without Posttraumatic Stress Disorder Comorbidity: New Evidence of Hypo-Reactivity to Stress in Fibromyalgia? Behavioral Medicine 47, 311–23.

Lovallo, W. (1975). The cold pressor test and autonomic function: a review and integration. Psychophysiology, 12, 268–82.

Maixner, W., Fillingim, R., Kincaid, S., Sigurdsson, A., & Harris, M. B. (1997). Relationship between pain sensitivity and resting arterial blood pressure in patients with painful temporomandibular disorders. Psychosomatic Medicine, 59, 503–11.

Makovac, E., Porciello, G., Palomba, D., Basile, B., & Ottaviani, C. (2020). Blood pressure-related hypoalgesia: a systematic review and meta-analysis. Journal of Hypertension 38, 1420–35.

Makovac, E., Venezia, A., Hohenschurz-Schmidt, D., Dipasquale, O., Jackson, J. B., Medina, S., O’Daly, O., Williams, S. C. R., McMahon, S. B., & Howard, M. A. (2021). The association between pain-induced autonomic reactivity and descending pain control is mediated by the periaqueductal grey. Journal of Physiology 599, 5243–60.

Marcuzzi, A., Chakiath, R. J., Siddall, P. J., Kellow, J. E., Hush, J. M., Jones, M. P., Costa, D. S., & Wrigley, P. J. (2019). Conditioned pain modulation (CPM) is reduced in irritable bowel syndrome: a systematic review and meta-analysis of CPM and the role of psychological factors. Journal of Clinical Gastroenterology 53, 399–408.

McLean, S. A., Williams, D. A., Harris, R. E., Kop, W. J., Groner, K. H., Ambrose, K., Lyden, A. K., Gracely, R. H., Crofford, L. J., & Geisser, M. E. (2005). Momentary relationship between cortisol secretion and symptoms in patients with fibromyalgia. Arthritis & rheumatism, 52, 3660–69.

Meeus, M., Nijs, J., Van de Wauwer, N., Toeback, L., & Truijen, S. (2008). Diffuse noxious inhibitory control is delayed in chronic fatigue syndrome: an experimental study. Pain, 139, 439–48.

Mischkowski, D., Palacios-Barrios, E. E., Banker, L., Dildine, T. C., & Atlas, L. Y. (2018). Pain or nociception? Subjective experience mediates the effects of acute noxious heat on autonomic responses. Pain, 159, 699–711.

Miyachi, R., Nishimura, T., Noguchi, M., Goda, A., Takeda, H., Takeshima, E., Kanazawa, Y., Imai, T., & Tanaka, W. (2025). Subgroup Characteristics of Middle-Aged and Older Women with Chronic Low Back Pain by Multiple Factors: A Hierarchical Cluster Analysis. Journal of Functional Morphology and Kinesiology, 10, 30.

Mohn, C., Vassend, O., & Knardahl, S. (2008). Experimental pain sensitivity in women with temporomandibular disorders and pain-free controls: The relationship to orofacial muscular contraction and cardiovascular responses. The Clinical Journal of Pain 24, 343–52.

Morgan, R. L., Whaley, P., Thayer, K. A., & Schünemann, H. J. (2018). Identifying the PECO: a framework for formulating good questions to explore the association of environmental and other exposures with health outcomes. Environment International 121, 1027.

Muhtz, C., Rodriguez-Raecke, R., Hinkelmann, K., Moeller-Bertram, T., Kiefer, F., Wiedemann, K., May, A., & Otte, C. (2013). Cortisol response to experimental pain in patients with chronic low back pain and patients with major depression. Pain Medicine 14, 498–503.

Nees, F., Loffler, M., Usai, K., & Flor, H. (2019). Hypothalamic-pituitary-adrenal axis feedback sensitivity in different states of back pain. Psychoneuroendocrinology, 101, 60–66.

Nicholas, M., Vlaeyen, J. W. S., Rief, W., Barke, A., Aziz, Q., Benoliel, R., Cohen, M., Evers, S., Giamberardino, M. A., Goebel, A., Korwisi, B., Perrot, S., Svensson, P., Wang, S. J., & Treede, R. D. (2019). The IASP classification of chronic pain for ICD-11: Chronic primary pain. Pain, 160, 28–37.

O’Brien, A. T., Deitos, A., Pego, Y. T., Fregni, F., & Carrillo-de-la-Peña, M. T. (2018). Defective endogenous pain modulation in fibromyalgia: a meta-analysis of temporal summation and conditioned pain modulation paradigms. The Journal of Pain, 19, 819–36.

Olsen, R. B., Bruehl, S., Nielsen, C. S., Rosseland, L. A., Eggen, A. E., & Stubhaug, A. (2013). Hypertension prevalence and diminished blood pressure–related hypoalgesia in individuals reporting chronic pain in a general population: The Tromsø Study. Pain, 154, 257–62.

Ozgocmen, S., Yoldas, T., Yigiter, R., Kaya, A., & Ardicoglu, O. (2006). R-R interval variation and sympathetic skin response in fibromyalgia. Archives of Medical Research 37, 630–4.

Page, M. J., McKenzie, J. E., Bossuyt, P. M., Boutron, I., Hoffmann, T. C., Mulrow, C. D., Shamseer, L., Tetzlaff, J. M., Akl, E. A., Brennan, S. E., Chou, R., Glanville, J., Grimshaw, J. M., Hróbjartsson, A., Lalu, M. M., Li, T., Loder, E. W., Mayo-Wilson, E., McDonald, S., McGuinness, L. A., Stewart, L. A., Thomas, J., Tricco, A. C., Welch, V. A., Whiting, P., & Moher, D. (2021). The PRISMA 2020 statement: an updated guideline for reporting systematic reviews. BMJ 372, n71.

Pickering, G., Achard, A., Corriger, A., Sickout-Arondo, S., Macian, N., Leray, V., Lucchini, C., Cardot, J. M., & Pereira, B. (2020). Electrochemical Skin Conductance and Quantitative Sensory Testing on Fibromyalgia. Pain Practice 20, 348–56.

Poli-Neto, O. B., Oliveira, A. M. Z., Salata, M. C., Cesar Rosa, E. S. J., Machado, D. R. L., Candido-Dos-Reis, F. J., & Nogueira, A. A. (2020). Strength Exercise Has Different Effects on Pressure Pain Thresholds in Women with Endometriosis-Related Symptoms and Healthy Controls: A Quasi-experimental Study. Pain Medicine 21, 2280–87.

Raison, V. (2009). Neurobiology of depression, fibromyalgia and neuropathic pain. Front Biosci, 14, 5291–338.

Rampazo, É. P., Rehder - Santos, P., Catai, A. M., & Liebano, R. E. (2024). Heart rate variability in adults with chronic musculoskeletal pain: A systematic review. Pain Practice, 24, 211–30.

Reshkova, V., Kalinova, D., & Milanov, I. (2015). Evaluation of antiviral antibodies against Epstein-Barr Virus and neurotransmitters in patients with fibromyalgia. J Neurol Neurosci, 6, 35.

Reyes Del Paso, G. A., Contreras-Merino, A. M., & Duschek, S. (2022). The Role of Depressive Disorders in Autonomic Cardiovascular Dysregulation in Fibromyalgia. Psychosomatic Medicine 84, 793–802.

Reyes del Paso, G. A., Garrido, S., Pulgar, A., & Duschek, S. (2011). Autonomic cardiovascular control and responses to experimental pain stimulation in fibromyalgia syndrome. Journal of Psychosomatic Research 70, 125–34.

Riva, R., Mork, P. J., Westgaard, R. H., & Lundberg, U. (2012). Comparison of the cortisol awakening response in women with shoulder and neck pain and women with fibromyalgia. Psychoneuroendocrinology, 37, 299–306.

Riva, R., Mork, P. J., Westgaard, R. H., Rø, M., & Lundberg, U. (2010). Fibromyalgia syndrome is associated with hypocortisolism. International journal of behavioral medicine, 17, 223–33.

Sacco, M., Meschi, M., Regolisti, G., Detrenis, S., Bianchi, L., Bertorelli, M., Pioli, S., Magnano, A., Spagnoli, F., & Giuri, P. G. (2013). The relationship between blood pressure and pain. The Journal of Clinical Hypertension 15, 600–05.

Saka-Kochi, Y., Kanbara, K., Yoshida, K., Kato, F., Kawashima, S., Abe, T., & Hasuo, H. (2024). Stress Response Pattern of Heart Rate Variability in Patients with Functional Somatic Syndromes. Applied Psychophysiology and Biofeedback, 49, 145–55.

Scheuren, P. S., De Schoenmacker, I., Rosner, J., Brunner, F., Curt, A., & Hubli, M. (2023). Pain-autonomic measures reveal nociceptive sensitization in complex regional pain syndrome. European Journal of Pain 27, 72–85.

Shaffer, F., & Ginsberg, J. P. (2017). An Overview of Heart Rate Variability Metrics and Norms. Frontiers in Public Health, 5.

Simon, E., Zsidó, A. N., Birkás, B., & Csathó, Á. (2022). Pain catastrophizing, pain sensitivity and fear of pain are associated with early life environmental unpredictability: a path model approach. BMC Psychology 10, 97.

Smeets, T., Cornelisse, S., Quaedflieg, C. W., Meyer, T., Jelicic, M., & Merckelbach, H. (2012). Introducing the Maastricht Acute Stress Test (MAST): a quick and non-invasive approach to elicit robust autonomic and glucocorticoid stress responses. Psychoneuroendocrinology, 37, 1998–2008.

Tan, H., Tumilty, S., Chapple, C., Liu, L., Othman, R., & Baxter, G. D. (2023). Acupoints sensitization in people with and without chronic low back pain: a matched-sample cross-sectional study. Journal of back and musculoskeletal rehabilitation, 36, 137–46.

Timmers, I., Kaas, A. L., Quaedflieg, C., Biggs, E. E., Smeets, T., & de Jong, J. R. (2018). Fear of pain and cortisol reactivity predict the strength of stress-induced hypoalgesia. European Journal of Pain 22, 1291–303.

Tiwari, R., Kumar, R., Malik, S., Raj, T., & Kumar, P. (2021). Analysis of heart rate variability and implication of different factors on heart rate variability. Current cardiology reviews, 17, 74–83.

Tracy, L. M., Ioannou, L., Baker, K. S., Gibson, S. J., Georgiou-Karistianis, N., & Giummarra, M. J. (2016). Meta-analytic evidence for decreased heart rate variability in chronic pain implicating parasympathetic nervous system dysregulation. Pain, 157, 7–29.

Umeda, M., Corbin, L. W., & Maluf, K. S. (2013). Preliminary investigation of absent nociceptive flexion reflex responses among more symptomatic women with fibromyalgia syndrome. Rheumatology International 33, 2365–72.

Vachon-Presseau, E. (2018). Effects of stress on the corticolimbic system: implications for chronic pain. Progress in Neuro-Psychopharmacology and Biological Psychiatry, 87, 216–23.

Valera-Calero, J. A., & Varol, U. (2022). Correlation among Routinary Physical Activity, Salivary Cortisol, and Chronic Neck Pain Severity in Office Workers: A Cross-Sectional Study. Biomedicines, 10.

Van Den Houte, M., Van Oudenhove, L., Van Diest, I., Bogaerts, K., Persoons, P., De Bie, J., & Van den Bergh, O. (2018). Negative Affectivity, Depression, and Resting Heart Rate Variability (HRV) as Possible Moderators of Endogenous Pain Modulation in Functional Somatic Syndromes. Frontiers in Psychology 9, 275.

van Middendorp, H., Lumley, M. A., Houtveen, J. H., Jacobs, J. W., Bijlsma, J. W., & Geenen, R. (2013). The impact of emotion-related autonomic nervous system responsiveness on pain sensitivity in female patients with fibromyalgia. Psychosomatic Medicine 75, 765–73.

Venezia, A., Fawsitt - Jones, H., Hohenschurz - Schmidt, D., Mancini, M., Howard, M., & Makovac, E. (2024). Investigating the effects of artificial baroreflex stimulation on pain perception: A comparative study in no - pain and chronic low back pain individuals. The Journal of Physiology, 602, 6941–57.

Wingenfeld, K., Nutzinger, D., Kauth, J., Hellhammer, D. H., & Lautenbacher, S. (2010). Salivary Cortisol Release and Hypothalamic Pituitary Adrenal Axis Feedback Sensitivity in Fibromyalgia Is Associated With Depression But Not With Pain. The Journal of Pain, 11, 1195–202.

Woda, A., L’Heveder, G., Ouchchane, L., & Bodéré, C. (2013). Effect of experimental stress in 2 different pain conditions affecting the facial muscles. The Journal of Pain 14, 455–66.

Woda, A., Picard, P., & Dutheil, F. (2016). Dysfunctional stress responses in chronic pain. Psychoneuroendocrinology, 71, 127–35.

Wyns, A., Hendrix, J., Lahousse, A., De Bruyne, E., Nijs, J., Godderis, L., & Polli, A. (2023). The biology of stress intolerance in patients with chronic pain—state of the art and future directions. Journal of clinical medicine, 12, 2245.

Ying-Chih, C., Yu-Chen, H., & Wei-Lieh, H. (2020). Heart rate variability in patients with somatic symptom disorders and functional somatic syndromes: A systematic review and meta-analysis. Neuroscience and Biobehavioral Reviews, 112, 336–44.

Yoshihara, T., Shigeta, K., Hasegawa, H., Ishitani, N., Masumoto, Y., & Yamasaki, Y. (2005). Neuroendocrine responses to psychological stress in patients with myofascial pain. Journal of Orofacial Pain, 19.

Zamunér, A. R., Forti, M., Andrade, C. P., Avila, M. A., & da Silva, E. (2016). Respiratory Sinus Arrhythmia and its Association with Pain in Women with Fibromyalgia Syndrome. Pain Practice 16, 704–11.

