## Supplementary 2 - Table S1 for "The Stress-Pain Connection in Chronic Primary Pain: A Systematic Review and Meta-Analysis of Physiological Stress Markers in Relation to Experimental Pain Responses"

**Supplementary File 2 - Table S1.** Eligibility criteria

|  | **Inclusion** | **Exclusion** |
| --- | --- | --- |
| **Population** | Humans  Adults (≥ 18y of age)  Chronic primary pain according to ICD-11 (including chronic primary visceral pain, chronic widespread pain, chronic primary musculoskeletal pain, chronic primary headache or orofacial pain, complex regional pain syndrome, painful bruising syndrome) | Animals  Children and adolescents (< 18y of age)  Other than chronic primary pain according to ICD-11 (including chronic cancer related pain, chronic postsurgical or post traumatic pain, chronic secondary pain)  (Sub)acute pain |
| **Exposure** | At least one marker of either ANS or HPA axis functionality | Psychological stress or self-reported stress |
| **Outcome** | At least one experimental outcome measure of pain (e.g. pain threshold, pain tolerance, exercise-induced analgesia, nociceptive flexion reflex, conditioned pain modulation, temporal summation of pain, offset analgesia, spatial summation) | Non experimentally-induced pain |
| **Design** | Analytical studies   Full-text reports  Articles in English, Dutch, French or German | Descriptive studies such as systematic reviews, letters to author / editor, editorials, etc.   Abstracts, congress, proceedings, research protocols, posters, etc.  Articles in other languages |

Abbreviations. ANS: Autonomic Nervous System, ICD: International Classification of Diseases, HPA: Hypothalamic-Pituitary-Adrenal, y: years.
