## Supplementary 3 - Table S2 for "The Stress-Pain Connection in Chronic Primary Pain: A Systematic Review and Meta-Analysis of Physiological Stress Markers in Relation to Experimental Pain Responses"

**Supplementary File 3 - Table S2.** Search strategies for each database

| **Database** | **Search string** |
| --- | --- |
| **PubMed** | (“chronic primary pain”[TIAB] OR "chronic visceral pain"[TIAB] OR "chronic chest pain"[TIAB] OR "chronic retrosternal pain"[TIAB] OR "chronic epigastric pain"[TIAB] OR "chronic bladder pain"[TIAB] OR "chronic pelvic pain"[TIAB] OR "chronic testicular pain"[TIAB] OR "chronic abdominal pain"[TIAB] OR "chronic functional abdominal pain"[TIAB] OR "chronic musculoskeletal pain"[TIAB] OR "chronic back pain"[TIAB] OR "chronic postural low back pain"[TIAB] OR "chronic low back pain"[TIAB] OR "chronic lumbar pain"[TIAB] OR "chronic low back ache"[TIAB] OR "chronic lower back ache"[TIAB] OR "chronic lower back pain"[TIAB] OR "chronic lumbar spine pain"[TIAB] OR "chronic lumbar vertebrae pain"[TIAB] OR "chronic lower spine pain"[TIAB] OR "chronic lumbar region pain"[TIAB] OR "chronic lumbar region ache"[TIAB] OR "chronic neck pain"[TIAB] OR "chronic cervical pain"[TIAB] OR "chronic thoracic pain"[TIAB] OR "chronic limb pain"[TIAB] OR "chronic widespread pain"[TIAB] OR "chronic orofacial pain"[TIAB] OR “reflux hypersensitivity”[TIAB] OR “epigastric pain syndrome”[TIAB] OR “functional dyspepsia”[TIAB] OR “painful bladder syndrome”[TIAB] OR “bladder pain syndrome”[TIAB] OR “interstitial cystiti*”[TIAB] OR “anal spasm”[TIAB] OR “chronic proctalgia”[TIAB] OR “pelvic pain syndrome”[TIAB] OR “abdominal pain syndrome”[TIAB] OR "gallbladder dyskinesia*"[TIAB] OR “biliary dyskinesia*”[TIAB] OR “penoscrotodynia”[TIAB] OR “vulvodynia”[TIAB] OR “vestibulodynia”[TIAB] OR “irritable bowel syndrome”[TIAB] OR “IBS”[TIAB] OR “mucous coliti*”[TIAB] OR “functional gallbladder disorder*”[TIAB] OR “functional sphincter of Oddi disorder*”[TIAB] OR "fibromyalgia"[TIAB] OR “fibrosit*”[TIAB] OR “fibromyositis”[TIAB] OR “muscular rheumatism”[TIAB] OR  "myofascial pain syndrome"[TIAB] OR “chronic headache*”[TIAB] OR “chronic temporomandibular disorder*”[TIAB] OR “chronic migraine”[TIAB] OR “burning mouth syndrome”[TIAB] OR “chronic burning mouth”[TIAB] OR “orodynia”[TIAB] OR “oral dysaesthesia”[TIAB] OR “chronic tension type headache*”[TIAB] OR “trigeminal autonomic cephalalgia*”[TIAB] OR "ciliary neuralgia*"[TIAB] OR “chronic cluster headache*”[TIAB] OR “hemicrania continua”[TIAB] OR “chronic paroxysmal hemicrania”[TIAB] OR “unilateral neuralgiform headache*”[TIAB] OR “complex regional pain syndrome”[TIAB] OR “CRPS”[TIAB] OR “reflex sympathetic dystrophy”[TIAB] OR “causalgia”[TIAB] OR “painful bruising syndrome”[TIAB] OR “gardner-diamond syndrome”[TIAB] OR “autoerythrocyte sensitization”[TIAB] OR “psychogenic purpura”[TIAB]) AND ("stress*"[TIAB] OR "hypothalamus pituitary adrenal"[TIAB] OR “hypothalamic pituitary adrenal”[TIAB] OR “hypothalamus pituitary”[TIAB] OR “hypothalamic pituitary”[TIAB] OR “pituitary adrenal”[TIAB] OR “hypophysis adrenal”[TIAB] OR “HPA axis”[TIAB] OR "cortisol"[TIAB] OR "glucocorticoid*"[TIAB] OR "catecholamine*"[TIAB] OR "adrenaline"[TIAB] OR “adrenergic”[TIAB] OR “noradrenaline”[TIAB] OR “noradrenergic”[TIAB] OR "epinephrine"[TIAB] OR “norepinephrine”[TIAB] OR “autonomic”[TIAB] OR “ANS”[TIAB] OR “sympathetic”[TIAB] OR “parasympathetic”[TIAB] OR “vagus”[TIAB] OR “vagal”[TIAB] OR "sympatho adrenal"[TIAB] OR “sympathoadrenal”[TIAB] OR “sympatho adrenergic”[TIAB] OR “sympathoadrenergic”[TIAB] OR “sympatho vagal”[TIAB] OR “sympathovagal”[TIAB] OR "blood pressure"[TIAB] OR “intravascular pressure”[TIAB] OR “venous pressure”[TIAB] OR “arterial pressure”[TIAB] OR "respiration rate*"[TIAB] OR “respiratory rate*”[TIAB] OR “respiratory sinus arrhythmia”[TIAB] OR "breathing rate*"[TIAB] OR “breathing frequency”[TIAB] OR “ventilation volume”[TIAB] OR “heart period*”[TIAB] OR “heart rhythm*”[TIAB] OR "cardiac rate*"[TIAB] OR "cardiac rhythm*"[TIAB] OR "heartbeat*"[TIAB] OR "heart beat*"[TIAB] OR "heart rate*"[TIAB] OR “heartrate*”[TIAB] OR “heart frequency”[TIAB] OR “HRV”[TIAB] OR "pulse rate*"[TIAB] OR “blood volume pulse”[TIAB] OR “preejection period”[TIAB] OR “pre ejection period”[TIAB] OR “body temperature”[TIAB] OR "skin temperature"[TIAB] OR "skin conduct*"[TIAB] OR “skin electric*”[TIAB] OR “electrodermal respon*”[TIAB] OR “psychogalvanic reflex”[TIAB] OR “electrodermal activity”[TIAB] OR “skin response”[TIAB] OR "salivary alpha amylas*"[TIAB] OR "blood oxygen saturation"[TIAB] OR “cardiorespiratory monitoring”[TIAB] OR “Electrocardiogra*”[TIAB] OR “ECG”[TIAB] OR “EKG”[TIAB] OR “baroreflex sensitivity”[TIAB] OR “baroreceptor sensitivity”[TIAB] OR “pupil diameter”[TIAB] OR “pupillary diameter”[TIAB] OR “pupil respons*”[TIAB] OR “pupillary respons*”[TIAB] OR “pupil size”[TIAB] OR “pupillary size”[TIAB] OR “muscle sympathetic nerve activity”[TIAB]) AND ("pain threshold*"[TIAB] OR "pain tolerance"[TIAB] OR "pain sensitivity"[TIAB] OR “hypoalgesia”[TIAB] OR “hyperalgesia”[TIAB] OR “analgesia”[TIAB] OR ((nociceptive[TIAB] OR nociception[TIAB]) AND ("flexion reflex"[TIAB] OR "flexor reflex"[TIAB] OR "withdrawal reflex"[TIAB] OR "RIII reflex"[TIAB] OR “R3 reflex”[TIAB])) OR "quantitative sensory test*"[TIAB] OR "diffuse noxious inhibitory control*"[TIAB] OR “heterotopic noxious conditioning stimul*”[TIAB] OR "endogenous modulation"[TIAB] OR "descending modulation"[TIAB] OR "pain modulation"[TIAB] OR “modulation of pain”[TIAB] OR "pain inhibition"[TIAB] OR "endogenous analgesia"[TIAB] OR “counterirritation”[TIAB] OR “counter irritation”[TIAB] OR "counterstimul*"[TIAB] OR "counter stimul*"[TIAB] OR "conditioning stimul*"[TIAB] OR "spatial summation"[TIAB] OR "temporal summation"[TIAB] OR "wind up"[TIAB] OR “windup”[TIAB] OR "pain facilitation"[TIAB]) |
| **Embase** | (‘chronic primary pain’:ti,ab,kw OR ‘chronic visceral pain’:ti,ab,kw OR ‘chronic chest pain’:ti,ab,kw OR ‘chronic retrosternal pain’:ti,ab,kw OR ‘chronic epigastric pain’:ti,ab,kw OR ‘chronic bladder pain’:ti,ab,kw OR ‘chronic pelvic pain’:ti,ab,kw OR ‘chronic testicular pain’:ti,ab,kw OR ‘chronic abdominal pain’:ti,ab,kw OR ‘chronic functional abdominal pain’:ti,ab,kw OR ‘chronic musculoskeletal pain’:ti,ab,kw OR ‘chronic back pain’:ti,ab,kw OR ‘chronic postural low back pain’:ti,ab,kw OR ‘chronic low back pain’:ti,ab,kw OR ‘chronic lumbar pain’:ti,ab,kw OR ‘chronic low back ache’:ti,ab,kw OR ‘chronic lower back ache’:ti,ab,kw OR ‘chronic lower back pain’:ti,ab,kw OR ‘chronic lumbar spine pain’:ti,ab,kw OR ‘chronic lumbar vertebrae pain’:ti,ab,kw OR ‘chronic lower spine pain’:ti,ab,kw OR ‘chronic lumbar region pain’:ti,ab,kw OR ‘chronic lumbar region ache’:ti,ab,kw OR ‘chronic neck pain’:ti,ab,kw OR ‘chronic cervical pain’:ti,ab,kw OR ‘chronic thoracic pain’:ti,ab,kw OR ‘chronic limb pain’:ti,ab,kw OR ‘chronic widespread pain’:ti,ab,kw OR ‘chronic orofacial pain’:ti,ab,kw OR ‘reflux hypersensitivity’:ti,ab,kw OR ‘epigastric pain syndrome’:ti,ab,kw OR ‘functional dyspepsia’:ti,ab,kw OR ‘painful bladder syndrome’:ti,ab,kw OR ‘bladder pain syndrome’:ti,ab,kw OR ‘interstitial cystiti*’:ti,ab,kw OR ‘anal spasm’:ti,ab,kw OR ‘chronic proctalgia’:ti,ab,kw OR ‘pelvic pain syndrome’:ti,ab,kw OR ‘abdominal pain syndrome’:ti,ab,kw OR ‘gallbladder dyskinesia*’:ti,ab,kw OR ‘biliary dyskinesia*’:ti,ab,kw OR ‘penoscrotodynia’:ti,ab,kw OR ‘vulvodynia’:ti,ab,kw OR ‘vestibulodynia’:ti,ab,kw OR ‘irritable bowel syndrome’:ti,ab,kw OR ‘IBS’:ti,ab,kw OR ‘mucous coliti*’:ti,ab,kw OR ‘functional gallbladder disorder*’:ti,ab,kw OR ‘functional sphincter of Oddi disorder*’:ti,ab,kw OR ‘fibromyalgia’:ti,ab,kw OR ‘fibrosit*’:ti,ab,kw OR ‘fibromyositis’:ti,ab,kw OR ‘muscular rheumatism’:ti,ab,kw OR  ‘myofascial pain syndrome’:ti,ab,kw OR ‘chronic headache*’:ti,ab,kw OR ‘chronic temporomandibular disorder*’:ti,ab,kw OR ‘chronic migraine’:ti,ab,kw OR ‘burning mouth syndrome’:ti,ab,kw OR ‘chronic burning mouth’:ti,ab,kw OR ‘orodynia’:ti,ab,kw OR ‘oral dysaesthesia’:ti,ab,kw OR ‘chronic tension type headache*’:ti,ab,kw OR ‘trigeminal autonomic cephalalgia*’:ti,ab,kw OR ‘ciliary neuralgia*’:ti,ab,kw OR ‘chronic cluster headache*’:ti,ab,kw OR ‘hemicrania continua’:ti,ab,kw OR ‘chronic paroxysmal hemicrania’:ti,ab,kw OR ‘unilateral neuralgiform headache*’:ti,ab,kw OR ‘complex regional pain syndrome’:ti,ab,kw OR ‘CRPS’:ti,ab,kw OR ‘reflex sympathetic dystrophy’:ti,ab,kw OR ‘causalgia’:ti,ab,kw OR ‘painful bruising syndrome’:ti,ab,kw OR ‘gardner-diamond syndrome’:ti,ab,kw OR ‘autoerythrocyte sensitization’:ti,ab,kw OR ‘psychogenic purpura’:ti,ab,kw) AND (‘stress*’:ti,ab,kw OR ‘hypothalamus pituitary adrenal’:ti,ab,kw OR ‘hypothalamic pituitary adrenal’:ti,ab,kw OR ‘hypothalamus pituitary’:ti,ab,kw OR ‘hypothalamic pituitary’:ti,ab,kw OR ‘pituitary adrenal’:ti,ab,kw OR ‘hypophysis adrenal’:ti,ab,kw OR ‘HPA axis’:ti,ab,kw OR ‘cortisol’:ti,ab,kw OR ‘glucocorticoid*’:ti,ab,kw OR ‘catecholamine*’:ti,ab,kw OR ‘adrenaline’:ti,ab,kw OR ‘adrenergic’:ti,ab,kw OR ‘noradrenaline’:ti,ab,kw OR ‘noradrenergic’:ti,ab,kw OR ‘epinephrine’:ti,ab,kw OR ‘norepinephrine’:ti,ab,kw OR ‘autonomic’:ti,ab,kw OR ‘ANS’:ti,ab,kw OR ‘sympathetic’:ti,ab,kw OR ‘parasympathetic’:ti,ab,kw OR ‘vagus’:ti,ab,kw OR ‘vagal’:ti,ab,kw OR ‘sympatho adrenal’:ti,ab,kw OR ‘sympathoadrenal’:ti,ab,kw OR ‘sympatho adrenergic’:ti,ab,kw OR ‘sympathoadrenergic’:ti,ab,kw OR ‘sympatho vagal’:ti,ab,kw OR ‘sympathovagal’:ti,ab,kw OR ‘blood pressure’:ti,ab,kw OR ‘intravascular pressure’:ti,ab,kw OR ‘venous pressure’:ti,ab,kw OR ‘arterial pressure’:ti,ab,kw OR ‘respiration rate*’:ti,ab,kw OR ‘respiratory rate*’:ti,ab,kw OR ‘respiratory sinus arrhythmia’:ti,ab,kw OR ‘breathing rate*’:ti,ab,kw OR ‘breathing frequency’:ti,ab,kw OR ‘ventilation volume’:ti,ab,kw OR ‘heart period*’:ti,ab,kw OR ‘heart rhythm*’:ti,ab,kw OR ‘cardiac rate*’:ti,ab,kw OR ‘cardiac rhythm*’:ti,ab,kw OR ‘heartbeat*’:ti,ab,kw OR ‘heart beat*’:ti,ab,kw OR ‘heart rate*’:ti,ab,kw OR ‘heartrate*’:ti,ab,kw OR ‘heart frequency’:ti,ab,kw OR ‘HRV’:ti,ab,kw OR ‘pulse rate*’:ti,ab,kw OR ‘blood volume pulse’:ti,ab,kw OR ‘preejection period’:ti,ab,kw OR ‘pre ejection period’:ti,ab,kw OR ‘body temperature’:ti,ab,kw OR ‘skin temperature’:ti,ab,kw OR ‘skin conduct*’:ti,ab,kw OR ‘skin electric*’:ti,ab,kw OR ‘electrodermal respon*’:ti,ab,kw OR ‘psychogalvanic reflex’:ti,ab,kw OR ‘electrodermal activity’:ti,ab,kw OR ‘skin response’:ti,ab,kw OR ‘salivary alpha amylas*’:ti,ab,kw OR ‘blood oxygen saturation’:ti,ab,kw OR ‘cardiorespiratory monitoring’:ti,ab,kw OR ‘Electrocardiogra*’:ti,ab,kw OR ‘ECG’:ti,ab,kw OR ‘EKG’:ti,ab,kw OR ‘baroreflex sensitivity’:ti,ab,kw OR ‘baroreceptor sensitivity’:ti,ab,kw OR ‘pupil diameter’:ti,ab,kw OR ‘pupillary diameter’:ti,ab,kw OR ‘pupil respons*’:ti,ab,kw OR ‘pupillary respons*’:ti,ab,kw OR ‘pupil size’:ti,ab,kw OR ‘pupillary size’:ti,ab,kw OR ‘muscle sympathetic nerve activity’:ti,ab,kw) AND (‘pain threshold*’:ti,ab,kw OR ‘pain tolerance’:ti,ab,kw OR ‘pain sensitivity’:ti,ab,kw OR ‘hypoalgesia’:ti,ab,kw OR ‘hyperalgesia’:ti,ab,kw OR ‘analgesia’:ti,ab,kw OR ((nociceptive:ti,ab,kw OR nociception:ti,ab,kw) AND (‘flexion reflex’:ti,ab,kw OR ‘flexor reflex’:ti,ab,kw OR ‘withdrawal reflex’:ti,ab,kw OR ‘RIII reflex’:ti,ab,kw OR ‘R3 reflex’:ti,ab,kw)) OR ‘quantitative sensory test*’:ti,ab,kw OR ‘diffuse noxious inhibitory control*’:ti,ab,kw OR ‘heterotopic noxious conditioning stimul*’:ti,ab,kw OR ‘endogenous modulation’:ti,ab,kw OR ‘descending modulation’:ti,ab,kw OR ‘pain modulation’:ti,ab,kw OR ‘modulation of pain’:ti,ab,kw OR ‘pain inhibition’:ti,ab,kw OR ‘endogenous analgesia’:ti,ab,kw OR ‘counterirritation’:ti,ab,kw OR ‘counter irritation’:ti,ab,kw OR ‘counterstimul*’:ti,ab,kw OR ‘counter stimul*’:ti,ab,kw OR ‘conditioning stimul*’:ti,ab,kw OR ‘spatial summation’:ti,ab,kw OR ‘temporal summation’:ti,ab,kw OR ‘wind up’:ti,ab,kw OR ‘windup’:ti,ab,kw OR ‘pain facilitation’:ti,ab,kw) |
| **Web of Science** | (“chronic primary pain” OR "chronic visceral pain" OR "chronic chest pain" OR "chronic retrosternal pain" OR "chronic epigastric pain" OR "chronic bladder pain" OR "chronic pelvic pain" OR "chronic testicular pain" OR "chronic abdominal pain" OR "chronic functional abdominal pain" OR "chronic musculoskeletal pain" OR "chronic back pain" OR "chronic postural low back pain" OR "chronic low back pain" OR "chronic lumbar pain" OR "chronic low back ache" OR "chronic lower back ache" OR "chronic lower back pain" OR "chronic lumbar spine pain" OR "chronic lumbar vertebrae pain" OR "chronic lower spine pain" OR "chronic lumbar region pain" OR "chronic lumbar region ache" OR "chronic neck pain" OR "chronic cervical pain" OR "chronic thoracic pain" OR "chronic limb pain" OR "chronic widespread pain" OR "chronic orofacial pain" OR “reflux hypersensitivity” OR “epigastric pain syndrome” OR “functional dyspepsia” OR “painful bladder syndrome” OR “bladder pain syndrome” OR “interstitial cystiti*” OR “anal spasm” OR “chronic proctalgia” OR “pelvic pain syndrome” OR “abdominal pain syndrome” OR "gallbladder dyskinesia*" OR “biliary dyskinesia*” OR “penoscrotodynia” OR “vulvodynia” OR “vestibulodynia” OR “irritable bowel syndrome” OR “IBS” OR “mucous coliti*” OR “functional gallbladder disorder*” OR “functional sphincter of Oddi disorder*” OR "fibromyalgia" OR “fibrosit*” OR “fibromyositis” OR “muscular rheumatism” OR  "myofascial pain syndrome" OR “chronic headache*” OR “chronic temporomandibular disorder*” OR “chronic migraine” OR “burning mouth syndrome” OR “chronic burning mouth” OR “orodynia” OR “oral dysaesthesia” OR “chronic tension type headache*” OR “trigeminal autonomic cephalalgia*” OR "ciliary neuralgia*" OR “chronic cluster headache*” OR “hemicrania continua” OR “chronic paroxysmal hemicrania” OR “unilateral neuralgiform headache*” OR “complex regional pain syndrome” OR “CRPS” OR “reflex sympathetic dystrophy” OR “causalgia” OR “painful bruising syndrome” OR “gardner-diamond syndrome” OR “autoerythrocyte sensitization” OR “psychogenic purpura”) AND ("stress*" OR "hypothalamus pituitary adrenal" OR “hypothalamic pituitary adrenal” OR “hypothalamus pituitary” OR “hypothalamic pituitary” OR “pituitary adrenal” OR “hypophysis adrenal” OR “HPA axis” OR "cortisol" OR "glucocorticoid*" OR "catecholamine*" OR "adrenaline" OR “adrenergic” OR “noradrenaline” OR “noradrenergic” OR "epinephrine" OR “norepinephrine” OR “autonomic” OR “ANS” OR “sympathetic” OR “parasympathetic” OR “vagus” OR “vagal” OR "sympatho adrenal" OR “sympathoadrenal” OR “sympatho adrenergic” OR “sympathoadrenergic” OR “sympatho vagal” OR “sympathovagal” OR "blood pressure" OR “intravascular pressure” OR “venous pressure” OR “arterial pressure” OR "respiration rate*" OR “respiratory rate*” OR “respiratory sinus arrhythmia” OR "breathing rate*" OR “breathing frequency” OR “ventilation volume” OR “heart period*” OR “heart rhythm*” OR "cardiac rate*" OR "cardiac rhythm*" OR "heartbeat*" OR "heart beat*" OR "heart rate*" OR “heartrate*” OR “heart frequency” OR “HRV” OR "pulse rate*" OR “blood volume pulse” OR “preejection period” OR “pre ejection period” OR “body temperature” OR "skin temperature" OR "skin conduct*" OR “skin electric*” OR “electrodermal respon*” OR “psychogalvanic reflex” OR “electrodermal activity” OR “skin response” OR "salivary alpha amylas*" OR "blood oxygen saturation" OR “cardiorespiratory monitoring” OR “Electrocardiogra*” OR “ECG” OR “EKG” OR “baroreflex sensitivity” OR “baroreceptor sensitivity” OR “pupil diameter” OR “pupillary diameter” OR “pupil respons*” OR “pupillary respons*” OR “pupil size” OR “pupillary size” OR “muscle sympathetic nerve activity”) AND ("pain threshold*" OR "pain tolerance" OR "pain sensitivity" OR “hypoalgesia” OR “hyperalgesia” OR “analgesia” OR ((nociceptive OR nociception) AND ("flexion reflex" OR "flexor reflex" OR "withdrawal reflex" OR "RIII reflex" OR “R3 reflex”)) OR "quantitative sensory test*" OR "diffuse noxious inhibitory control*" OR "endogenous modulation" OR "descending modulation" OR "pain modulation" OR “modulation of pain” OR "pain inhibition" OR "endogenous analgesia" OR “counterirritation” OR “counter irritation” OR "counterstimul*" OR "counter stimul*" OR "conditioning stimul*" OR "spatial summation" OR "temporal summation" OR "wind up" OR “windup” OR "pain facilitation") |
| **CINAHL** | (“chronic primary pain” OR "chronic visceral pain" OR "chronic chest pain" OR "chronic retrosternal pain" OR "chronic epigastric pain" OR "chronic bladder pain" OR "chronic pelvic pain" OR "chronic testicular pain" OR "chronic abdominal pain" OR "chronic functional abdominal pain" OR "chronic musculoskeletal pain" OR "chronic back pain" OR "chronic postural low back pain" OR "chronic low back pain" OR "chronic lumbar pain" OR "chronic low back ache" OR "chronic lower back ache" OR "chronic lower back pain" OR "chronic lumbar spine pain" OR "chronic lumbar vertebrae pain" OR "chronic lower spine pain" OR "chronic lumbar region pain" OR "chronic lumbar region ache" OR "chronic neck pain" OR "chronic cervical pain" OR "chronic thoracic pain" OR "chronic limb pain" OR "chronic widespread pain" OR "chronic orofacial pain" OR “reflux hypersensitivity” OR “epigastric pain syndrome” OR “functional dyspepsia” OR “painful bladder syndrome” OR “bladder pain syndrome” OR “interstitial cystiti*” OR “anal spasm” OR “chronic proctalgia” OR “pelvic pain syndrome” OR “abdominal pain syndrome” OR "gallbladder dyskinesia*" OR “biliary dyskinesia*” OR “penoscrotodynia” OR “vulvodynia” OR “vestibulodynia” OR “irritable bowel syndrome” OR “IBS” OR “mucous coliti*” OR “functional gallbladder disorder*” OR “functional sphincter of Oddi disorder*” OR "fibromyalgia" OR “fibrosit*” OR “fibromyositis” OR “muscular rheumatism” OR  "myofascial pain syndrome" OR “chronic headache*” OR “chronic temporomandibular disorder*” OR “chronic migraine” OR “burning mouth syndrome” OR “chronic burning mouth” OR “orodynia” OR “oral dysaesthesia” OR “chronic tension type headache*” OR “trigeminal autonomic cephalalgia*” OR "ciliary neuralgia*" OR “chronic cluster headache*” OR “hemicrania continua” OR “chronic paroxysmal hemicrania” OR “unilateral neuralgiform headache*” OR “complex regional pain syndrome” OR “CRPS” OR “reflex sympathetic dystrophy” OR “causalgia” OR “painful bruising syndrome” OR “gardner-diamond syndrome” OR “autoerythrocyte sensitization” OR “psychogenic purpura”) AND ("stress*" OR "hypothalamus pituitary adrenal" OR “hypothalamic pituitary adrenal” OR “hypothalamus pituitary” OR “hypothalamic pituitary” OR “pituitary adrenal” OR “hypophysis adrenal” OR “HPA axis” OR "cortisol" OR "glucocorticoid*" OR "catecholamine*" OR "adrenaline" OR “adrenergic” OR “noradrenaline” OR “noradrenergic” OR "epinephrine" OR “norepinephrine” OR “autonomic” OR “ANS” OR “sympathetic” OR “parasympathetic” OR “vagus” OR “vagal” OR "sympatho adrenal" OR “sympathoadrenal” OR “sympatho adrenergic” OR “sympathoadrenergic” OR “sympatho vagal” OR “sympathovagal” OR "blood pressure" OR “intravascular pressure” OR “venous pressure” OR “arterial pressure” OR "respiration rate*" OR “respiratory rate*” OR “respiratory sinus arrhythmia” OR "breathing rate*" OR “breathing frequency” OR “ventilation volume” OR “heart period*” OR “heart rhythm*” OR "cardiac rate*" OR "cardiac rhythm*" OR "heartbeat*" OR "heart beat*" OR "heart rate*" OR “heartrate*” OR “heart frequency” OR “HRV” OR "pulse rate*" OR “blood volume pulse” OR “preejection period” OR “pre ejection period” OR “body temperature” OR "skin temperature" OR "skin conduct*" OR “skin electric*” OR “electrodermal respon*” OR “psychogalvanic reflex” OR “electrodermal activity” OR “skin response” OR "salivary alpha amylas*" OR "blood oxygen saturation" OR “cardiorespiratory monitoring” OR “Electrocardiogra*” OR “ECG” OR “EKG” OR “baroreflex sensitivity” OR “baroreceptor sensitivity” OR “pupil diameter” OR “pupillary diameter” OR “pupil respons*” OR “pupillary respons*” OR “pupil size” OR “pupillary size” OR “muscle sympathetic nerve activity”) AND ("pain threshold*" OR "pain tolerance" OR "pain sensitivity" OR “hypoalgesia” OR “hyperalgesia” OR “analgesia” OR ((nociceptive OR nociception) AND ("flexion reflex" OR "flexor reflex" OR "withdrawal reflex" OR "RIII reflex" OR “R3 reflex”)) OR "quantitative sensory test*" OR "diffuse noxious inhibitory control*" OR "endogenous modulation" OR "descending modulation" OR "pain modulation" OR “modulation of pain” OR "pain inhibition" OR "endogenous analgesia" OR “counterirritation” OR “counter irritation” OR "counterstimul*" OR "counter stimul*" OR "conditioning stimul*" OR "spatial summation" OR "temporal summation" OR "wind up" OR “windup” OR "pain facilitation") |
| **Psych-ARTICLES** | (“chronic primary pain” OR "chronic visceral pain" OR "chronic chest pain" OR "chronic retrosternal pain" OR "chronic epigastric pain" OR "chronic bladder pain" OR "chronic pelvic pain" OR "chronic testicular pain" OR "chronic abdominal pain" OR "chronic functional abdominal pain" OR "chronic musculoskeletal pain" OR "chronic back pain" OR "chronic postural low back pain" OR "chronic low back pain" OR "chronic lumbar pain" OR "chronic low back ache" OR "chronic lower back ache" OR "chronic lower back pain" OR "chronic lumbar spine pain" OR "chronic lumbar vertebrae pain" OR "chronic lower spine pain" OR "chronic lumbar region pain" OR "chronic lumbar region ache" OR "chronic neck pain" OR "chronic cervical pain" OR "chronic thoracic pain" OR "chronic limb pain" OR "chronic widespread pain" OR "chronic orofacial pain" OR “reflux hypersensitivity” OR “epigastric pain syndrome” OR “functional dyspepsia” OR “painful bladder syndrome” OR “bladder pain syndrome” OR “interstitial cystiti*” OR “anal spasm” OR “chronic proctalgia” OR “pelvic pain syndrome” OR “abdominal pain syndrome” OR "gallbladder dyskinesia*" OR “biliary dyskinesia*” OR “penoscrotodynia” OR “vulvodynia” OR “vestibulodynia” OR “irritable bowel syndrome” OR “IBS” OR “mucous coliti*” OR “functional gallbladder disorder*” OR “functional sphincter of Oddi disorder*” OR "fibromyalgia" OR “fibrosit*” OR “fibromyositis” OR “muscular rheumatism” OR  "myofascial pain syndrome" OR “chronic headache*” OR “chronic temporomandibular disorder*” OR “chronic migraine” OR “burning mouth syndrome” OR “chronic burning mouth” OR “orodynia” OR “oral dysaesthesia” OR “chronic tension type headache*” OR “trigeminal autonomic cephalalgia*” OR "ciliary neuralgia*" OR “chronic cluster headache*” OR “hemicrania continua” OR “chronic paroxysmal hemicrania” OR “unilateral neuralgiform headache*” OR “complex regional pain syndrome” OR “CRPS” OR “reflex sympathetic dystrophy” OR “causalgia” OR “painful bruising syndrome” OR “gardner-diamond syndrome” OR “autoerythrocyte sensitization” OR “psychogenic purpura”) AND ("stress*" OR "hypothalamus pituitary adrenal" OR “hypothalamic pituitary adrenal” OR “hypothalamus pituitary” OR “hypothalamic pituitary” OR “pituitary adrenal” OR “hypophysis adrenal” OR “HPA axis” OR "cortisol" OR "glucocorticoid*" OR "catecholamine*" OR "adrenaline" OR “adrenergic” OR “noradrenaline” OR “noradrenergic” OR "epinephrine" OR “norepinephrine” OR “autonomic” OR “ANS” OR “sympathetic” OR “parasympathetic” OR “vagus” OR “vagal” OR "sympatho adrenal" OR “sympathoadrenal” OR “sympatho adrenergic” OR “sympathoadrenergic” OR “sympatho vagal” OR “sympathovagal” OR "blood pressure" OR “intravascular pressure” OR “venous pressure” OR “arterial pressure” OR "respiration rate*" OR “respiratory rate*” OR “respiratory sinus arrhythmia” OR "breathing rate*" OR “breathing frequency” OR “ventilation volume” OR “heart period*” OR “heart rhythm*” OR "cardiac rate*" OR "cardiac rhythm*" OR "heartbeat*" OR "heart beat*" OR "heart rate*" OR “heartrate*” OR “heart frequency” OR “HRV” OR "pulse rate*" OR “blood volume pulse” OR “preejection period” OR “pre ejection period” OR “body temperature” OR "skin temperature" OR "skin conduct*" OR “skin electric*” OR “electrodermal respon*” OR “psychogalvanic reflex” OR “electrodermal activity” OR “skin response” OR "salivary alpha amylas*" OR "blood oxygen saturation" OR “cardiorespiratory monitoring” OR “Electrocardiogra*” OR “ECG” OR “EKG” OR “baroreflex sensitivity” OR “baroreceptor sensitivity” OR “pupil diameter” OR “pupillary diameter” OR “pupil respons*” OR “pupillary respons*” OR “pupil size” OR “pupillary size” OR “muscle sympathetic nerve activity”) AND ("pain threshold*" OR "pain tolerance" OR "pain sensitivity" OR “hypoalgesia” OR “hyperalgesia” OR “analgesia” OR ((nociceptive OR nociception) AND ("flexion reflex" OR "flexor reflex" OR "withdrawal reflex" OR "RIII reflex" OR “R3 reflex”)) OR "quantitative sensory test*" OR "diffuse noxious inhibitory control*" OR "endogenous modulation" OR "descending modulation" OR "pain modulation" OR “modulation of pain” OR "pain inhibition" OR "endogenous analgesia" OR “counterirritation” OR “counter irritation” OR "counterstimul*" OR "counter stimul*" OR "conditioning stimul*" OR "spatial summation" OR "temporal summation" OR "wind up" OR “windup” OR "pain facilitation") |
| **Scopus** | (“chronic primary pain” OR "chronic visceral pain" OR "chronic chest pain" OR "chronic retrosternal pain" OR "chronic epigastric pain" OR "chronic bladder pain" OR "chronic pelvic pain" OR "chronic testicular pain" OR "chronic abdominal pain" OR "chronic functional abdominal pain" OR "chronic musculoskeletal pain" OR "chronic back pain" OR "chronic postural low back pain" OR "chronic low back pain" OR "chronic lumbar pain" OR "chronic low back ache" OR "chronic lower back ache" OR "chronic lower back pain" OR "chronic lumbar spine pain" OR "chronic lumbar vertebrae pain" OR "chronic lower spine pain" OR "chronic lumbar region pain" OR "chronic lumbar region ache" OR "chronic neck pain" OR "chronic cervical pain" OR "chronic thoracic pain" OR "chronic limb pain" OR "chronic widespread pain" OR "chronic orofacial pain" OR “reflux hypersensitivity” OR “epigastric pain syndrome” OR “functional dyspepsia” OR “painful bladder syndrome” OR “bladder pain syndrome” OR “interstitial cystiti*” OR “anal spasm” OR “chronic proctalgia” OR “pelvic pain syndrome” OR “abdominal pain syndrome” OR "gallbladder dyskinesia*" OR “biliary dyskinesia*” OR “penoscrotodynia” OR “vulvodynia” OR “vestibulodynia” OR “irritable bowel syndrome” OR “IBS” OR “mucous coliti*” OR “functional gallbladder disorder*” OR “functional sphincter of Oddi disorder*” OR "fibromyalgia" OR “fibrosit*” OR “fibromyositis” OR “muscular rheumatism” OR  "myofascial pain syndrome" OR “chronic headache*” OR “chronic temporomandibular disorder*” OR “chronic migraine” OR “burning mouth syndrome” OR “chronic burning mouth” OR “orodynia” OR “oral dysaesthesia” OR “chronic tension type headache*” OR “trigeminal autonomic cephalalgia*” OR "ciliary neuralgia*" OR “chronic cluster headache*” OR “hemicrania continua” OR “chronic paroxysmal hemicrania” OR “unilateral neuralgiform headache*” OR “complex regional pain syndrome” OR “CRPS” OR “reflex sympathetic dystrophy” OR “causalgia” OR “painful bruising syndrome” OR “gardner-diamond syndrome” OR “autoerythrocyte sensitization” OR “psychogenic purpura”) AND ("stress*" OR "hypothalamus pituitary adrenal" OR “hypothalamic pituitary adrenal” OR “hypothalamus pituitary” OR “hypothalamic pituitary” OR “pituitary adrenal” OR “hypophysis adrenal” OR “HPA axis” OR "cortisol" OR "glucocorticoid*" OR "catecholamine*" OR "adrenaline" OR “adrenergic” OR “noradrenaline” OR “noradrenergic” OR "epinephrine" OR “norepinephrine” OR “autonomic” OR “ANS” OR “sympathetic” OR “parasympathetic” OR “vagus” OR “vagal” OR "sympatho adrenal" OR “sympathoadrenal” OR “sympatho adrenergic” OR “sympathoadrenergic” OR “sympatho vagal” OR “sympathovagal” OR "blood pressure" OR “intravascular pressure” OR “venous pressure” OR “arterial pressure” OR "respiration rate*" OR “respiratory rate*” OR “respiratory sinus arrhythmia” OR "breathing rate*" OR “breathing frequency” OR “ventilation volume” OR “heart period*” OR “heart rhythm*” OR "cardiac rate*" OR "cardiac rhythm*" OR "heartbeat*" OR "heart beat*" OR "heart rate*" OR “heartrate*” OR “heart frequency” OR “HRV” OR "pulse rate*" OR “blood volume pulse” OR “preejection period” OR “pre ejection period” OR “body temperature” OR "skin temperature" OR "skin conduct*" OR “skin electric*” OR “electrodermal respon*” OR “psychogalvanic reflex” OR “electrodermal activity” OR “skin response” OR "salivary alpha amylas*" OR "blood oxygen saturation" OR “cardiorespiratory monitoring” OR “Electrocardiogra*” OR “ECG” OR “EKG” OR “baroreflex sensitivity” OR “baroreceptor sensitivity” OR “pupil diameter” OR “pupillary diameter” OR “pupil respons*” OR “pupillary respons*” OR “pupil size” OR “pupillary size” OR “muscle sympathetic nerve activity”) AND ("pain threshold*" OR "pain tolerance" OR "pain sensitivity" OR “hypoalgesia” OR “hyperalgesia” OR “analgesia” OR ((nociceptive OR nociception) AND ("flexion reflex" OR "flexor reflex" OR "withdrawal reflex" OR "RIII reflex" OR “R3 reflex”)) OR "quantitative sensory test*" OR "diffuse noxious inhibitory control*" OR "endogenous modulation" OR "descending modulation" OR "pain modulation" OR “modulation of pain” OR "pain inhibition" OR "endogenous analgesia" OR “counterirritation” OR “counter irritation” OR "counterstimul*" OR "counter stimul*" OR "conditioning stimul*" OR "spatial summation" OR "temporal summation" OR "wind up" OR “windup” OR "pain facilitation") |
