## Supplementary 4 - Methods for "The Stress-Pain Connection in Chronic Primary Pain: A Systematic Review and Meta-Analysis of Physiological Stress Markers in Relation to Experimental Pain Responses"

**Supplementary File** **4.** Methods

#### **Information Sources and Search Strategy**

Articles were searched on title, abstract and keywords, except in CINAHL and PsycARTICLES due to limitations regarding search options. In Scopus, this was achieved through using the ‘Article Title, Abstract and Keywords’ search option. In Web of Science, the used search option was ‘Topic’.

#### **Data Items and Collection Process**

The following data were extracted from the included studies: (i) authors and year, (ii) study design (i.e., case-control or cross-sectional) and timing of the stress and pain measurements, (iii) participant characterization (sample size, biological sex, mean and standard deviation of age in years) for the chronic primary pain group (subcategorized as widespread or localized pain), (iv) identical study sample characteristics of the pain-free control group (if applicable), (v) the type of physiological marker of the stress systems (either ANS or HPA axis), (vi) the experimental pain measure, (vii) the significance levels for each physiological marker of stress (per group and timepoint, if applicable), (viii) the significance levels for each experimental pain outcome (per group and timepoint, if applicable), and (ix) the data concerning the interaction (e.g., correlation coefficients) between the physiological markers of stress and pain outcome measures. Relevant information presented in graphs was extracted using Web Plot Digitizer (<https://plotdigitizer.com/>) when data was otherwise not accessible. Forty authors were contacted to provide additional data (e.g., correlation coefficients for the stress-pain interaction) that was not accessible from the paper. Reminders were sent after two weeks and four weeks.

#### **Risk of Bias in Individual Studies**

The NOS implements a “star system” and consists of four categories. For cross-sectional studies, selection of the study group is the first category (maximum three stars), exposure is the second category (maximum two stars), comparability of the participants is the third category (maximum two stars), and outcome is the fourth category (maximum three stars). For case-control studies, the same categories were used except the outcome and exposure are considered as one category, and the included sections differ slightly. Therefore, the maximum number of stars for the selection of the study group was four, for the comparability two, and for the exposure and outcome four. An additional self-constructed criterion (maximum two stars) was developed for the outcome in both designs. This criterion evaluates the ascertainment of the experimental pain outcome measure since only the ascertainment of the exposure (physiological markers of stress) is implemented in the NOS. Per criterion of each category, a star could be awarded, indicating low risk of bias. The total score for the cross-sectional studies and case-control studies is 12 stars (including the additional self-constructed criterion). A summary of each category with its predefined interpretation is presented in Supplementary File 5 - Table S4.

#### **Certainty of Evidence**

Certainty of evidence was assessed in accordance with the Grading of Recommendations, Assessment, Development, and Evaluations (GRADE) approach (Balshem, et al., 2011). Body of evidence for each outcome started at “low” because only non-randomized studies were included. Five factors may lead to downgrading the certainty of evidence by one or two levels when there are serious or very serious concerns, respectively: risk of bias, inconsistency, indirectness, imprecision, or publication bias. The certainty of the evidence was not downgraded for indirectness, as all results directly addressed the research question. Additionally, one factor, large magnitude of an effect, could lead to an upgrade of the certainty of evidence of the quantitative syntheses. The certainty of evidence was upgraded (+1) if the effect size was moderate to large (pooled correlation coefficient ≥ .30) (Cohen, 2013) and can eventually vary from very low (⨁◯◯◯) to high (⨁⨁⨁⨁) (G. Guyatt, et al., 2011; Prasad, 2024).

#### **Qualitative and Quantitative Analyses**

Studies not reporting, or authors not providing data concerning an interaction between stress and pain were excluded for the qualitative and quantitative syntheses. Clusters were established based on the timing of the stress measurements and pain assessments (i.e., assessed at baseline, during a stressor and after a stressor/during recovery of a stressor). Quantitative analyses were executed with R 4.4.1 using the RStudio interface (R Core Team, 2024). Correlation coefficients (*r*) between physiological stress parameters and experimental pain outcome measures were gathered, along with the corresponding sample size. Meta-analyses were conducted when at least two studies provided data on the interaction between the same physiological stress parameter and experimental outcome, measured at the same time point (e.g., baseline). First, a Fisher’s z-transformation to each correlation coefficient was performed to stabilize variances. Afterwards, meta-analyses were performed using random-effect model (with inverse-variance method), implemented through the “metaphor” (version 4.6-0) package. Random-effects were chosen due to the expected heterogeneity of the studies concerning study populations and the measurements of stress and pain. The between-study variance (*τ*²) was estimated using the Restricted Maximum Likelihood (REML) method. Wald-type confidence intervals were calculated for the pooled effect sizes and afterwards back-transformed to the correlation scale for interpretation. Positive values indicate a positive correlation whereas negative values indicate a negative correlation. Heterogeneity among studies was assessed using the *I*² statistic. The *I*² statistic quantified the percentage of total variation in effect sizes attributable to heterogeneity rather than random chance (Higgins & Thompson, 2002). *I*² values above 25% were considered as mild heterogeneity (- 1) and values above 50% were considered as strong heterogeneity (- 2). In case of substantial heterogeneity (> 50%), subgroup analyses and sensitivity analyses based on the risk of bias were performed to investigate important sources of heterogeneity where possible. Additionally, if a minimum of ten studies were included in a meta-analysis, potential asymmetry was visually inspected with funnel plots to help judging the publication bias. In case of publication bias, the trim-and-fill method was used to adjust for funnel plot asymmetry (G. H. Guyatt, et al., 2011).

### **REFERENCES**

Balshem, H., Helfand, M., Schünemann, H. J., Oxman, A. D., Kunz, R., Brozek, J., Vist, G. E., Falck-Ytter, Y., Meerpohl, J., Norris, S., & Guyatt, G. H. (2011). GRADE guidelines: 3. Rating the quality of evidence. *Journal of Clinical Epidemiology, 64*, 401-6.

Cohen, J. (2013). *Statistical power analysis for the behavioral sciences*: routledge.

Guyatt, G., Oxman, A. D., Akl, E. A., Kunz, R., Vist, G., Brozek, J., Norris, S., Falck-Ytter, Y., Glasziou, P., DeBeer, H., Jaeschke, R., Rind, D., Meerpohl, J., Dahm, P., & Schünemann, H. J. (2011). GRADE guidelines: 1. Introduction-GRADE evidence profiles and summary of findings tables. *Journal of Clinical Epidemiology 64*, 383-94.

Guyatt, G. H., Oxman, A. D., Montori, V., Vist, G., Kunz, R., Brozek, J., Alonso-Coello, P., Djulbegovic, B., Atkins, D., Falck-Ytter, Y., Williams, J. W., Jr., Meerpohl, J., Norris, S. L., Akl, E. A., & Schünemann, H. J. (2011). GRADE guidelines: 5. Rating the quality of evidence--publication bias. *Journal of Clinical Epidemiology 64*, 1277-82.

Higgins, J. P., & Thompson, S. G. (2002). Quantifying heterogeneity in a meta-analysis. *Statistics in Medicine 21*, 1539-58.

Prasad, M. (2024). Introduction to the GRADE tool for rating certainty in evidence and recommendations. *Clinical Epidemiology and Global Health, 25*, 101484.
