## Supplementary 5 - Table S4 for "The Stress-Pain Connection in Chronic Primary Pain: A Systematic Review and Meta-Analysis of Physiological Stress Markers in Relation to Experimental Pain Responses"

**Supplementary File 5**

**Table S4a.** Risk of bias results for case-control studies

|  | **Selection** | | | | **Comparability** | | **Exposure and Outcome** | | | |  |
| --- | --- | --- | --- | --- | --- | --- | --- | --- | --- | --- | --- |
| **Study** | **1** | **2** | **3** | **4** | **1a** | **1b** | **1** | **2** | **3** | ***4*** | **Total stars (/12)** |
| Bandeira et al. (2021) | a* | d | c | a* | yes* | yes* | a** | a* | a* | *c** | 9 |
| Bossenger et al. (2023) | a* | d | c | a* | yes* | yes* | b** | a* | a* | *b*** | 10 |
| Chalaye et al. (2012) | a* | c | a* | a* | yes* | yes* | b** | a* | c | *b*** | 10 |
| Chalaye et al. (2014) | c | d | c | b | yes* | yes* | b** | a* | c | *b*** | 7 |
| Cohen et al. (2000) | a* | c | b | a* | yes* | yes* | c* | a* | a* | *c** | 8 |
| Davydov et al. (2024) | a* | c | c | a* | yes* | yes* | b** | a* | a* | *b*** | 10 |
| De Abreu Freitas et al. (2012) | a* | c | b | b | yes* | yes* | c* | a* | a* | *a*** | 8 |
| de la Coba et al. (2018) | a* | c | a* | a* | yes* | yes* | c* | a* | a* | *c** | 9 |
| del Paso et al. (2011) | a* | c | a* | a* | yes* | yes* | c* | a* | a* | *c** | 9 |
| del Paso et al. (2022) | a* | c | a* | a* | yes* | yes* | c* | a* | a* | *b*** | 10 |
| Farmer et al. (2014) | a* | c | a* | a* | yes* | yes* | a** | a* | a* | *b*** | 11 |
| Flor et al. (2004) | b | d | c | b | yes* | yes* | b** | a* | a* | *b*** | 8 |
| Galeazzi et al. (2001) | a* | c | c | b | no | no | d | a* | a* | *b*** | 5 |
| Garcia-Hernandez et al. (2022) | a* | b* | c | a* | no | yes* | c* | a* | a* | *c** | 8 |
| Geiss et al. (2012) | a* | c | a* | a* | yes* | yes* | b** | a* | a* | *c** | 10 |
| Granot et al. (2002) | a* | c | b | b | yes* | yes* | c* | a* | a* | *c** | 7 |
| Jarrett et al. (2014) | a* | b* | a* | a* | yes* | yes* | c* | a* | b | *c** | 9 |
| Jarrett et al. (2016) | a* | b* | a* | a* | yes* | yes* | b** | a* | b | *b*** | 11 |
| Jonsson et al. (2025) | a* | a* | b | a* | yes* | yes* | a** | a* | a* | *c** | 10 |
| Kadetoff et al. (2007) | a* | c | c | b | yes* | yes* | c* | a* | a* | *c** | 7 |
| Kadetoff et al. (2010) | a* | c | b | b | yes* | yes* | c* | a* | a* | *c** | 7 |
| Kim et al. (2015) | a* | d | c | a* | yes* | yes* | c* | a* | c | *b*** | 8 |
| Larsson et al. (2008) | b | c | a* | b | yes* | yes* | c* | a* | a* | *b*** | 8 |
| Löffler et al. (2023) | a* | a* | a* | a* | yes* | yes* | c* | a* | a* | *c** | 10 |
| Lòpez-Lòpez et al. (2021) | a* | b* | a* | a* | yes* | yes* | c* | a* | a* | *b*** | 11 |
| Maixner et al. (1997) | b | c | a* | a* | no | yes* | a** | a* | a* | *b*** | 9 |
| Meeus et al. (2008) | a* | c | a* | a* | yes* | yes* | a** | a* | a* | *b*** | 11 |
| Mohn et al. (2008) | a* | c | b | a* | yes* | yes* | c* | a* | a* | *c** | 8 |
| Muhtz et al. (2013) | a* | c | c | a* | yes* | no | c* | a* | a* | *c** | 7 |
| Murray et al. (2004) | a* | c | c | a* | yes* | yes* | b** | a* | a* | *a*** | 10 |
| Nees et al. (2019) | b | b* | a* | b | yes* | yes* | c* | a* | c | *a*** | 8 |
| Ozgocmen et al. (2006) | b | d | c | b | yes* | yes* | b** | a* | a* | *b*** | 8 |
| Pardo et al. (2019) | b | b* | a* | a* | no | yes* | c* | a* | a* | *c** | 8 |
| Pickering et al. (2019) | b | d | c | b | yes* | no | a** | a* | c | *b*** | 6 |
| Poli-Neto et al. (2020) | b | c | b | b | yes* | yes* | a** | a* | a* | *c** | 7 |
| Quartana et al. (2010) | a* | c | b | a* | no | yes* | b** | a* | a* | *b*** | 8 |
| Rampazo et al. (2024) | a* | b* | a* | a* | yes* | yes* | a** | a* | c | *b*** | 11 |
| Scheuren et al. (2023) | a* | c | c | a* | yes* | no | b** | a* | a* | *a*** | 9 |
| Tan et al. (2023) | a* | c | a* | a* | yes* | yes* | b** | a* | a* | *a*** | 11 |
| Thieme et al. (2022) | a* | c | c | b | yes* | yes* | a** | a* | a* | *c** | 8 |
| Umeda et al. (2013) | a* | b* | a* | a* | yes* | yes* | c* | a* | b | *b*** | 10 |
| Van Den Houte et al. (2018) | a* | b* | a* | a* | yes* | yes* | b** | a* | c | *b*** | 10 |
| Van Middendorp et al. (2013) | a* | b* | a* | b | yes* | yes* | c* | a* | a* | *b*** | 10 |
| Venezia et al. (2024) | a* | b* | a* | a* | yes* | yes* | b** | a* | c | *b*** | 11 |
| Wingenfeld et al. (2010) | a* | c | a* | b | yes* | yes* | b** | a* | a* | *c** | 9 |
| Woda et al. (2013) | a* | c | b | b | yes* | yes* | c* | a* | a* | *a*** | 8 |
| Zamunér et al. (2016) | a* | a* | a* | b | yes* | yes* | b** | a* | a* | *b*** | 11 |

Results of the risk of bias assessment using an adapted version of the Newcastle-Ottawa scale.

**Selection**: **1.** Is the case definition adequate?: a = yes, with independent validation*, b = yes, e.g. record linkage or based on self-reports, c = no description; **2.** Representativeness of the cases: a = truly representative of the average in the target population*, b = somewhat representative of the average in the target population*, c = selected group of users, d = no description of the sampling strategy; **3.** Selection of controls: a = community controls*, b = hospital controls, c = no description; **4.** Definition of controls: a = no history of disease*, b = no description of source.

**Comparability**: **1.** Comparability of cases and controls on the basis of the design or analysis: yes = study controls for the most important factor*, yes = study controls for any additional factor*.

**Exposure and Outcome**: **1.** Ascertainment of the exposure: a = validated measurement tool/method**, b = non-validated measurement tool/method, but an existing protocol which previously has been used**, c = non-validated measurement tool/method, but the tool/method is available or described*, d = no description; **2.** Same method of ascertainment for cases and controls: a = yes*, b = no; **3.** Non-respondents: a = comparability between respondents and non-respondents characteristics is established, and the response rate is satisfactory, or when a response rate is inappropriate*, b = the response rate is unsatisfactory, or the comparability between respondents and non-respondents is unsatisfactory, c = no description. ***4.*** *Ascertainment of the outcome: a = validated measurement too/method**, b = non-validated measurement tool/method, but an existing protocol which previously has been used**, c = non-validated measurement tool/method, but the tool/method is available or described*, d = no description.*

.

**Table S4b.** Risk of bias assessment for cross-sectional studies

|  | **Selection** | | | **Exposure** | **Comparability** | | **Outcome** | | |  |
| --- | --- | --- | --- | --- | --- | --- | --- | --- | --- | --- |
| **Study** | **1** | **2** | **3** | **1** | **1a** | **1b** | **1** | **2** | ***3*** | **Total stars (/12)** |
| Crettaz et al. (2013) | b* | b | c | c* | yes* | yes* | c* | a* | *b*** | 8 |
| De Bruijn et al. (2011) | c | a* | c | c* | yes* | yes* | c* | a* | *b*** | 8 |
| Miyachi et al. (2025) | c | b | a* | c* | yes* | no | c* | a* | *b*** | 7 |
| Reshkova et al. (2015) | c | d | c | c* | no | no | d | a* | *d* | 2 |
| Valera-Calero et al. (2022) | c | a* | c | c* | yes* | yes* | c* | a* | *b*** | 8 |

Results of the risk of bias assessment using an adapted version of the Newcastle-Ottawa scale.

**Selection**: **1.** Representativeness of the sample: a = truly representative of the average in the target population*, b = somewhat representative of the average in the target population*, c = selected group of users, d = no description of the sampling strategy; **2.** Sample size: a = justified and satisfactory*, b = not justified; **3.** Non-respondents: a = comparability between respondents and non-respondents characteristics is established, and the response rate is satisfactory*, b = the response rate is unsatisfactory, or the comparability between respondents and non-respondents is unsatisfactory, c = no description of the response rate or the characteristics of the responders and the non-responders.

**Exposure**: **1.** Ascertainment of the outcome: a = validated measurement too/method**, b = non-validated measurement tool/method, but an existing protocol which previously has been used**, c = non-validated measurement tool/method, but the tool/method is available or described*, d = no description.

**Comparability**: **1.** The participants in different outcome groups are comparable, based on the study design or analysis. Confounding factors are controlled: yes = the study controls for the most important factor*, yes = the study controls for any additional factor*.

**Outcome**: **1.** Assessment of the outcome: a = independent blind assessment**, b = record linkage**, c = self-report*, d = no description; **2.** Statistical test: a = the statistical test used to analyse the data is clearly described and appropriate, and the measurement of the association is presented, including confidence intervals and probability levels (p-value)*, b = the statistical test is not appropriate, not described of incomplete; **3**. *Ascertainment of the outcome: a = validated measurement too/method**, b = non-validated measurement tool/method, but an existing protocol which previously has been used**, c = non-validated measurement tool/method, but the tool/method is available or described*, d = no description.*
