## Supplementary 6 - Table S3 for "The Stress-Pain Connection in Chronic Primary Pain: A Systematic Review and Meta-Analysis of Physiological Stress Markers in Relation to Experimental Pain Responses"

**Supplementary File 6 - Table S3.** Study Characteristics

| **Study** | ***Design* and timing of stress; pain measurement** | **Participants** | | **Stress marker** | **Pain outcome** | **Separate results** | |
| --- | --- | --- | --- | --- | --- | --- | --- |
|  |  | **CPP**  **(loc/ws)** | **HC** |  |  | **STRESS** | **PAIN** |
| Bandeira et al. (2021) | *Case-control*  Stress:  Baseline (5 min.) (1), during (5 min.) (2) and after (5 min.) (3) passive visualization task; Pain: Baseline and after stressor  **Stress reactivity and recovery;**  **Pain after stressor** | CLBP (loc);  *n* = 47 (26F/21M); 46.0 (9.6) y | *n* = 47 (25F/22M); 46.4 (8.68) y | HRV (LF, HF, LF/HF) with Polar H7 Bluetooth portable heart rate monitor fitted below V6 | PPTh at trapezius muscle, elbow, lumbar spine and suprapatellar region with a digital algometer | Group difference compared to baseline:  **LF HRV** CPP>HC  (2) *p* = .002  (3) *p* = .088  **HF HRV** CPP<HC (2) *p* < .001  (3) *p* = .212  **LF/HF** CPP>HC (2) *p* < .001  (3) *p* = .828  Change in LF; HF; LF/HF from baseline to during stress:  CPP: ↑; ↓*; ↑*  HC: ↓; ↑; ↓  Difference between 1-2; 1-3 in CPP:  **LF HRV**  *p* = .008; *p* = .58  **HF HRV**  *p* = .017  **LF/HF**  *p* < .001; *p* = .71  Difference between 1-2; 1-3 in HC:  **LF HRV**  *p* = .71; *p* = > .99  **HF HRV**  *p* = .80; *p* = > .99  **LF/HF**  *p* = .74; *p* = .33  Significant main effects for group:  for LF (*p* < .001); CPP>HC and HF  (*p* < .001); CPP<HC  Post stimulus:  *p* > .05 for diff from baseline  for LF, HF and  LF/HF within-group | Group difference after stressor:  Trapezius R, Trapezius L, Elbow R, Elbow L, Lumbar R, Lumbar L, Suprapatellar R, Suprapatellar L  *p* < .001  Change in PPTh (> for HC; < for CPP) after stressor: Trapezius R CPP: *p* < .001 HC: *p* = .005  Trapezius L CPP: *p* < .001 HC: *p* = .004  Elbow R CPP: *p* < .001 HC: *p* = .004  Elbow L  CPP: *p* < .001 HC: *p* < .001  Lumbar R CPP: *p* < .001 HC: *p* < .001  Lumbar L CPP: *p* < .001 HC: *p* = .97  Suprapatellar R  CPP: *p* < .001 HC: *p* < .001  Suprapatellar L CPP: *p* < .001 HC: *p* < .001 |
| Bossenger et al. (2023) | *Case-control*  Stress: Baseline (5 min.) and during isometric contraction of the quadriceps muscle;  Pain: baseline and during isometric contraction  **Stress reactivity;**  **Pain during stressor** | FM (ws);  *n* = 13 (13F/0M);  47 (14) y | *n* = 15 (11F/4M); 53 (10) y | HF HRV and PEP with ECG on each side of the neck and on each side of the thorax, SCL and SCR with electrodes on index and middle finger | PPTh on 2 cm distal of patella and volar forearm with a handheld algometer | Baseline: **HF HRV**  CPP<HC (*p* = .03)  **PEP** NS group diff (*p* = .43)  **SCL** CPP>HC *(p* = .002)  **SCR** CPP>HC (*p* = .08)  During stressor:  **HF HRV**  NS group diff  **PEP**  NS group diff  **SCL** NS group diff  **SCR** NS group diff  During stressor compared to baseline:  **HF HRV** ↓ in CPP (*p* < .001) and HC (*p* < .001)  **PEP** ↓ in CPP (*p* = .03) and HC (*p* = .009)  **SCL** ↑ in CPP (*p* < .001) and HC (*p* < .001) **SCR** ↑ in CPP (*p* < .001) and HC (*p* < .001)  Changes in ANS:  **HF HRV**  ↓ CPP<HC (*p* = .063)  **PEP**  ↓ NS (*p* = .53)  **SCL**  ↑ NS (*p* = .16)  **SCR**  ↑ NS (*p* = .20) | Baseline:  **PPTh patella**  CPP<HC (*p* = .01)  **PPTh forearm** NS group diff (*p* = .83)  During stressor: **PPTh patella**  ↓ in HC (*p =* .01), NS diff for CPP (*p =* .06) **PPTh forearm** NS group diff (*p =* .18) |
| Chalaye et al. (2012) | *Case-control*  Stress: Baseline (2 min.) and during CPT  **Stress reactivity** | FM (ws);  *n* = 10 (10F/0M); 46.7 (7.1) y  IBS (loc);  *n* = 13 (13F/0M);  37 (15.8) y | *n* = 10 (10F/0M); 41 (8.5) y | HR, HRV (LF, HF) with standard 3-lead montage | CPT: immersion of fingers, wrist, elbow and shoulder in circulating cold water (12°C) for 2 min. | During pain:  **HR**  FM: ↑ (*p* < .02) IBS and HC: ↑ (*p >* .16)  **LF HRV** FM: ↑ (*p =* .143) IBS: ↓ (*p =* .125)  HC: ↓ (*p =* .019)  **HF HRV**  FM: ↓ (*p =* .06)  IBS: NS diff  HC: ↑ (*p =* .06)  Change in ANS:  **LF HRV** CPP<HC (*p =* .017)  **HF HRV**  NS group diff | Pain in FM>HC (*p =* .02) |
| Chalaye et al. (2014) | *Case-control*  Stress: Baseline (2 min.) and during CPT  **Stress reactivity** | FM (ws);  *n* = 23 (23F/0M); 49.3 (2) y | *n* = 25 (25F/0); 48.4 (1.7) y | HR with standard 3-lead montage, BP and BRS noninvasively with Nexfin monitor (finger cuff on middle phalanx of left index or middle finger) | CPM, CPT; HPTh, HPTo: immersion of right arm (up to the elbow) in circulating cold water (12°C) for 2 min. | Baseline:  **HR** CPP*>*HC (*p =* .01) **BP and BRS**  NS group diff (*p >* .05)  During pain: **HR** CPP*>*HC (*p =* .015)  **BP** NS group diff (*p >* .19)  **BRS** NS group diff (*p =* .13)  Change in ANS:  **HR**  NS group diff (*p =* .13)  **BP**  CPP weaker SBP (*p =* .007) and DBP (*p =* .1) incline  **BRS**  NS group diff (*p =* .39) | **HPTh**  CPP<HC (*p =* .002)  **HPTo**  CPP<HC (*p =* .001)  **Pain intensity during CPT** CPP>HC (*p =* .004)  **CPM** **efficiency**  CPP<HC (*p =* .009) |
| Cohen et al. (2000) | *Case-control*  Stress: Baseline (15 min.) | FM (ws);  *n* = 22 (22F/0M); 47 (7.1) y | *n* = 22 (22F/0M); 47(7) y | HR, HRV (LF, HF) with lead II ECG | Tenderness assessment measured with involuntary verbal or facial expressions on 18 tender points by thumb palpation and dolorimeter | **HR**  CPP>HC (*p =* .0057)  **HRV**  CPP<HC (*p =* .0012)  **LF HRV**  CPP>HC (*p* < .0001)  **HF HRV**  CPP<HC (*p* < .0001) | **Tender point counts**  CPP>HC (*p* < .01)  **Mean tenderness**  CPP<HC (*p <* .01) |
| Crettaz et al. (2013) | *Cross-sectional*  Stress: Baseline (10 min.), during and after TSST (immediately & after 30 min., 20 min. after pain); Pain: Baseline and after TSST (immediately for 10 min. and 70 min. after TSST)  **Stress reactivity and recovery; pain after stressor** | FM (ws);  *n* = 13 (13F/0M); 49.85 (10.55) y | *n* = 10 (10F/0M); 27.70 (5.58) | HR, SVB with 2-channel ECG | CPTh and HPTh with a TSA 2001-II, PPTh with a pressure gauge device, MPTh and WUR with pinprick stimuli at medial forearm | During stressor: **HR** ↑ in CPP (*p <* .001) and HC (*p =* .13)  **SVB** NS ↑ in CPP (*p =* .06) and HC (*p =* .12) | Baseline: **CPTh, HPTh, PPTh, MPTh** CPP<HC (*p =* .01; *p =* .007; *p =* .001; *p =* .004 respectively)  After stressor (up to 10’ and 70’):  **HPTh**  ↓ in HC (*p =* .057), mainly after 90 min. and CPP (*p =* .06) **CPTh**  ↓ in HC (*p =* .08) and CPP (*p =* .007) mainly after 10 min.  **PPTh**  ↓ in CPP (*p =* .004) mainly after 10 min., but not in HC **MPTh, WUR** No change |
| Davydov et al. (2024) | *Case-control*  Stress: Baseline | FM (ws);  *n* = 48 (48F/0M); 52.35 (8.95) y | *n* = 37 (37F/0M); 49.84 (6.65) y | Hair cortisol: 5 cm hair in length, ±150 strands with enzyme-linked immunosorbent assay | PPTh & PPTo on fingernail and SREP with a pressure algometer | Baseline:  **Cortisol**  CPP>HC (*p =* .121) | **PPTh**  CPP<HC (*p =* .001)  **PPTo**  CPP<HC (*p =* .001)  **SREP**  CPP>HC (*p =* .001) |
| De Abreu Freitas et al. (2012) | *Case-control*  Stress: Baseline | FM (ws);  *n* = 17 (17F/0M); 53 (7.98) y | *n* = 19 (19F/0M); 53.32 (6.46) y | Serum cortisol and DHEA between 8:00 - 9:30 with Immulite 1000 Immunoassay System | PPTh & PPTo at 18 tender points with a pressure algometer | Baseline:  **Cortisol & DHEA**  NS diff in cortisol (*p =* .325) and  DHEA-S (*p =* .094) between CPP and HC | **PPTh and PPTo**  CPP<HC (*p <* .0001) |
| De Bruijn et al. (2011) | *Cross-sectional*  Stress: Baseline (N.A.) and after ergobicycle test over a 1 min. period;  Pain: Baseline  **Stress recovery** | FM (ws);  *n* = 18 (18F/0M); 37.3 (7.7) y | N.A. | HR(R), SBP, DBP | CPTh and HPTh with a 9-cm^2^ Peltier-based ATS thermode,  PPTh at thenar/foot with an algometer, MPTh with weighted pinpricks,  thermal WUR with a contact heat-evoked potential stimulator | N.A. | N.A. |
| De La Coba et al. (2018) | *Case-control*  Stress: Baseline (5 min.) | FM (ws);  *n* = 30 (30F/0M); 52 (9.57) y | *n* = 27 (27F/0M); 51.4 (9.94) y | SBP, DBP continuously with a Task Force Monitor at the first phalange of the second and third finger of the left hand | PPTh, PPTo SREP on fingernail with a pressure algometer | Baseline:  **SBP** CPP>HC (*p =* .004) **DBP** CPP<HC (*p =* .069) | **PPTh**  CPP<HC (*p =* .001)  **PPTo**  CPP<HC (*p =* .002)  **SREP sensitization** CPP>HC (*p <* .001) |
| Del Paso et al. (2011) | *Case-control*  Stress: Baseline (10 min.), during CPT recovery in 5 min. after CPT  **Stress reactivity and recovery** | FM (ws);  *n* = 35 (32F/3M); 50.5 (6.7) y | *n* = 29 (27F/2M); 49.4 (9.4) y | (very) LF HRV, HF HRV, LF/HF, RRI with ECG, with 4 electrodes (Einthoven I and II), SBP, DBP oscillometrically from right brachial artery and continuously at first phalange of second and third finger of left hand | CPTh, CPTo by immersing hand and forearm in ice-cold water (1-3°C) | Baseline:  **RRI** CPP<HC (*p <* .01)  **Very LF and HF HRV** CPP<HC (*p <* .01) **LF HRV** CPP<HC (*p =* .09)  **LF/HF HRV**  CPP>HC (*p <* .01)  During pain:  **RRI**  ↓ (*p <* .01) **SBP-DBP**  ↑ (*p <* .01)  **LF/HF** **HRV** ↑ In CPP only (*p <* .05)  Recovery:  **RRI**  ↑ (*p <* .01)  **SBP-DBP**  ↓ (p *<* .01)  **HF HRV** ↑ compared to baseline for HC only (*p <* .05)  **LF/HF HRV** ↑ compared to baseline for CPP only (*p <* .05) | **CPTh**  CPP<HC (*p <* .01)  **CPTo** CPP<HC (*p <* .01) |
| Del Paso et al. (2022) | *Case-control*  Stress: Baseline (10 min.), during pain, during mental stress task (Uchida-Kraepelin), recovery over a 5 min. period after stressor  **Stress reactivity and recovery** | FM (without depression; ws);  *n* = 26 (25F/1M); 51.12 (7.29) y | *n* = 29 (27F/2M); 49.34 (9.53) y | HF HRV, IBI, PEP with ECG with 4 electrodes (Einthoven I and II), BRS, BP oscillometrically from left brachial artery, and continuously at first phalange of the second and third finger of the right hand | CPTh, CPTo during CPT immersing hand and forearm (temperature of 1-3°C) | During pain:  **IBI**  CPP & HC ↓ (*p <* .001)  NS group diff (*p =* .26)  **BP** CPP & HC ↑ (*p <* .001)  Change of BP: CPP<HC (*p <* .001)  **PEP**  CPP & HC ↓  (*p <* .001)  NS group diff  **BRS**  CPP & HC ↓ (*p <* .002)  NS group diff (*p =* .30)  During mental task:  **IBI**  CPP & HC ↓ (*p <* .001)  Change of IBI: CPP<HC (*p <* .001)  NS group diff (*p =* .49)  **BP**  CPP & HC ↑ (*p <* .001)  Change of BP: CPP<HC (*p =* .002)  *SBP* CPP<HC (*p =* .013)  *DBP*  CPP<HC (*p =* .005)  **PEP**  CPP & HC ↓  (*p <* .001)  NS group diff  **BRS**  NS diff in BRS  **HF HRV**  HC: ↓ (*p =* .002)  CPP: = (*p =* .13)  NS group diff (*p =* .277) | Baseline:  **CPTh, CPTo**  CPP<HC (*p* < .05) |
| Farmer et al. (2014) | *Case-control*  Stress: Baseline (5 min.) and during (ANS)/ 2 min. after (HPA) pain  **Stress reactivity** | FCP (loc);  *n* = 20 (11F/9M); 38.7 (28-59) y | *n* = 20 (11F/9M, ethnicity-matched); 38.2 (24-49) y | BP with photoplethysmography, SC with electrodes at distal digit of the right index and ring fingers, CVT, HR, BRS with ECG at right and left subclavicular areas and cardiac apex, serum cortisol between 14:00 – 16:00 | Somatic PTo with a strain gauge,  visceral PTo with an oesophageal catheter | Baseline:  **HR and SC**  CPP>HC (*p =* .04)  **CVT**  CPP<HC (*p =* .003)  **BRS**  CPP<HC (*p =* .02)  **BP**  NS group diff  **Cortisol**  CPP>HC (*p <* .01)  Change in ANS parameters During somatic pain:  **HR** ↑ in CPP & HC  CPP<HC (*p =* .01)  **SBP** ↓ in CPP, ↑ in HC  CPP<HC (*p =* .03)  **DBP** ↑in CPP & HC  CPP<HC (*p =* .18)  **CVT** ↑ in CPP, ↓ in HC CPP>HC (*p =* .002)  **BRS**  ↑ in CPP, ↓ in HC  CPP>HC (*p =* .01)  **SC** ↑ in CPP & HC  CPP<HC (*p =* .007)  Change in ANS parameters during visceral pain:  **HR** ↓ in CPP, ↑ in HC  CPP<HC (*p =* .002)  **SBP**  ↓ in CPP, ↑ in HC  CPP<HC (*p =* .008)  **DBP** ↓ in CPP, ↑ in HC  CPP<HC (*p =* .06)  **CVT** ↑ in CPP, ↓ in HC CPP>HC (*p =* .008)  **BRS** ↑ in CPP, ↓ in HC  CPP>HC (*p =* .008)  **SC** ↓ in CPP, ↑ in HC  CPP<HC (*p =* .006)  After pain:  **Cortisol**  CPP>HC (*p <* .01), NS diff in absolute ↑ between patients | **Somatic PTo**  CPP<HC (*p <* .0001)  **Visceral PTo**  CPP<HC (*p =* .009) |
| Flor et al. (2004) | *Case-control*  Stress: Baseline and during pain induction  **Stress reactivity** | CLBP (loc);  *n* = 16 (10F/6M); 42.2 (12.9) y | *n* = 16 (10F/6M); 38.6 (10.2) y | HR with cardiotachometer with electrodes on the right clavicula and the left and right lower part of the rib cage, SC with electrodes at the thenar and hypothenar eminence of the dominant hand | EPTh and EPTo on third digit with a gold electrode | **HR**  NS group diff, increased with increasing intensity (*p <* .05)  **SC** NS | **EPTh**  CLBP<HC (*p <* .05)  **EPTo**  CLBP<CMTH (*p <* .05)  CMTH>HC (*p <* .05) |
| Galeazzi et al. (2001) | *Case-control*  Stress: During pain induction (5 min.)  Pain: Baseline and after CPT  **Stress reactivity; pain after stressor** | UC (loc);  *n* = 10 (N.A.); 25-58 y | *n* = 12 (5F/7M); 22-38 y | HR, BP | Oesophageal PTh with a latex balloon | During pain:  **Change in HR**  CPP<HC (*p =* .07)  **Change in BP**  CPP>HC (*p =* .07) Parameters ↑ during pain in both groups | Baseline:  **Oesophageal PTh**  CPP<HC (*p <* .05)  After stressor:  **Oesophageal PTh**  HC: ↓ (*p <* .05) CPP: NS diff |
| Garcia-Hernandez et al. (2022) | *Case-control*  Stress: Baseline (6 min.) | FM (ws);  *n* = 29 (29F/0M); 50 (10.71) y | *n* = 30 (30F/0M); 50.37 (7.54) y | SC with a MP36 Biopac polygraph with electrodes on the thenar and hypothenar areas of the right hand | PPTh, PPTo and SREP on second finger with a pressure algometer | Baseline:  **SC**  CPP<HC (*p =* .045) | **PPTh**  CPP<HC (*p <* .001)  **PPTo**  CPP<HC (*p =* .012)  **SREP**  CPP>HC (*p <* .001) |
| Geiss et al. (2012) | *Case-control*  Stress: Baseline and recovery (1 – 375 min. after PPTh)  **Stress recovery** | FM (ws);  *n* = 12 (12F/0M); 50 (2.7) y | *n* = 15 (15F/0M); 41 (2.98) y | Salivary cortisol (with resolved fluorescence immunoassay) and norepinephrine (with high-performance liquid chromatography with electrochemical detection) at +15, 30, 45, 60 min. after awakening; 8:00 – 11:00; 16:30 (30 min. before PPTh); 20:00 | PPTh at 4 tender points with a pressure algometer | Baseline:  **Cortisol (CAR-day)**  NS group diff (*p <* .44)  **Norepinephrine**  Baseline (10’ before PPTh):  CPP>HC ([F (1,22) = 6.04, *p <* .02])  Recovery:  **Cortisol**  After pain (10 min.)  CPP>HC (*p <* .01)  After pain (60 min.)  CPP>HC (*p* > .05)  After pain (10 min.):  **Norepinephrine**  CPP>HC  ([F (1,22) = 13.08, *p < .*002]) | **PPTh** CPP<HC (*p < .*001) |
| Granot et al. (2002) | *Case-control*  Stress: Baseline (1 min.), during (1 min.) and after pain induction (1 min.)  **Stress reactivity and recovery** | VVS (loc);  *n* = 44 (44F/0M); 27.1 (7.6) y | *n* = 41 (41F/0M); 25.4 (5.2) y | HR, BP | Heat pain rating with a TSA-2001 (30x30 mm^2^) | During pain:  **Change in SBP** CPP ↑, HC ↓ (*p =* .0026)  **Change in HR** NS group diff | **Heat pain**  CPP>HC (*p =* .006) |
| Jarrett et al. (2014) | *Case-control*  Stress: Baseline and immediately after CPM  **Stress reactivity** | IBS (loc);  *n* = 20 (20F/0M, 75% white); 27.4 (6.6) y | *n* = 20 (20F/0M, 75% white); 27.6 (5.5) y | Salivary cortisol between 8:00 – 10:00 with a horseradish peroxidase-linked immunoassay | CPM using heat stimuli with a Pain & Sensory Evaluation System and for test stimulus, and 12°C water bath for conditioning stimulus | **Cortisol**  Baseline:  NS group diff (*p =* .94)  Immediately after pain induction:  NS group diff (*p =* .38) | **CPM efficiency**  NS group diff (*p =* .46) |
| Jarrett et al. (2016) | *Case-control*  Stress: Baseline (5’ blocks out of 12h) | IBS (loc);  *n* = 54 (54F/0M, 92% white); 28.4 (6.7) y | *n* = 37 (37F/0M, 88% white); 28.6 (6.8) | HRV with 3-channel digital Holter ECG | CPM using heat stimuli on volar forearm with a Pain & Sensory Evaluation System as test stimulus, and 46.5°C hot water bath as conditioning stimulus | Baseline:  **HRV** NS group diff (*p >* .05) | **CPM efficiency**  NS group diff |
| Jonsson et al. (2025) | *Case-control*  Stress: Baseline | FM (ws); *n* = 38 (38F/0M); 44.2 (9.8) y | *n* = 44 (44F/0M); 40.5 (9.8) y | Respiration rate with portable intensive care monitor, SBP, DBP | PPTh at 14 paraspinal locations with a gun-shaped Somedic algometer | **SBP**  NS group diff (*p* = .842)  **DBP**  NS group diff (*p* = .168)  **Respiratory rate**  CPP>HC (*p* = .001) | **PPTh**  CPP<HC (*p* = .001) |
| Kadetoff et al. (2007) | *Case-control*  Stress: Baseline (N.A.), during contraction of max isometric strength test of knee extensor, and recovery (immediately, 5 min., 10 min. and 15 min. following contraction)  Pain: Baseline and between contractions  **Stress reactivity and recovery; pain after stressor** | FM (ws);  *n* = 17 (17F/0M); 38.8 (N.A.) (22-56) y | *n* = 17 (17F/0M); 37.4 (N.A.) (22-53) y | HR, BP with a digital blood pressure monitor UA-767 | PPTh at quadriceps muscle and deltoid muscle with a pressure algometer | Baseline:  **HR, BP** NS group diff (*p =* .28)  During stressor: **SBP- DBP**  ↑ in both groups (*p <* .001) **HR** ↑ in both groups (*p < .*001)  **Change in HR from baseline** CPP>HC (*p < .*02) | Baseline: **PPTh** CPP<HC (*p* < .0001)  Between contractions:  ↑ PPTh in both groups  sign change over time in both groups (*p =* .0001), but no group-time effect |
| Kadetoff et al. (2010) | *Case-control*  Stress: Baseline (10 min.), during contraction of max isometric strength test of knee extensor, and recovery (30 min. after exhaustion)  Pain: Baseline, between contractions, recovery  **Stress reactivity and recovery; pain after stressor** | FM (ws);  *n* = 16 (16F/0M); 38.2 (N.A.) y | *n* = 16 (16F/0M); 38.3 (N.A.) y | HR, BP with digital blood pressure monitor UA-767, serum cortisol either in morning or in afternoon with immunofluorescence | PPTh at quadriceps and deltoid muscle with a pressure algometer | Baseline:  **SBP - DBP** NS group diff for SBP (*p < .*77) or DBP (*p < .*74) **HR** NS group diff (*p <* .34)  **Cortisol** NS group diff (*p >* .05)  During stressor:  **SBP - DBP**  ↑ in both groups (*p < .*001)  **HR** ↑ in both groups  (*p < .*0001)  **Cortisol**  NS ↑ at exhaustion in both groups | Baseline:  **PPTh** CPP<HC at quadriceps (*p < .*0001) and deltoid (*p < .*0001) muscle  After stressor: NS time effect in quadriceps (*p <* .19) or deltoid (*p <* .07) muscle  NS interaction between group/time (quadriceps: *p <* .78, deltoid: *p <* .62) |
| Kim et al. (2015) | *Case-control*  Stress: Baseline (6 min.) and during pain in MRI  **Stress reactivity** | FM (ws);  *n* = 35 (32F/3M); 44.9 (12) y | *n* = 14 (10F/4M); 44.2 (14.3) y | HF HRV with ECG (MRI-compatible Patient Monitoring System) | 40/100 pain intensity, TSP with cuff on gastrocnemius muscle | During pain:  **HF HRV** ↓ in CPP (*p < .*01) and HC (*p =* .15).  More robust ↓ over time in CPP (first 2 min.: *p =* .60; middle 2 min.: *p =* .07; last 2 min.: *p < .*01) | **40/100** NS group diff (*p =* .86)  **TSP** CPP>HC (*p < .*05) |
| Larsson et al. (2008) | *Case-control*  Stress: Baseline | Chronic neck- and shoulder pain (loc);  *n* = 20 (20F/0M); 43.8 (9.8) y | *n* = 20 (20F/0M); 45.2 (11.3) y | Urinary cortisol from the night proceeding the experiment and during the experiment with DelFIA technique | PPTh on trapezius and tibialis anterior muscle with pressure algometer | **Cortisol**  NS group diff | **PPTh on trapezius** CPP<HC examined with microdialysis (*p =* .013)  CPP<HC not examined with microdialysis (*p =* .20)  **PPTh on tibialis anterior** NS group diff (*p =* .80; *p =* .40) |
| Löffler et al. (2023) | *Case-control*  Stress: Baseline (5 min.) and during mental arithmetic task (similar to konzentrations-und leistungstest)  Pain: Baseline and after stressor  **Stress reactivity; pain after stressor** | Chronic widespread or localized back pain (ws/loc);  *n* = 22 (15F/7M); 56.6 (12.4) y | *n* = 18 (15F/3M); 52.9 (11.9) y | HR, BP with Criticare 506 N vital signs monitor | EPTh and EPTo on back with a pair of needle electrodes | Baseline:  NS group diff for HR or BP (*p >* .11)  During stressor:  NS group diff for HR or BP (*p >* .11)  CPP>HC: ↑ in SBP (*p =* .01) and DBP (*p =* .03) was sign ↑, not for HR (*p =* .53)  HR, SBP and DBP ↑ in both groups (*p =* .002; *p < .*001; *p < .*001) | **EPTh** NS (*p =* .12)  **EPTo** CPP<HC (*p =* .04) |
| Lòpez-Lòpez et al. (2021) | *Case-control*  Stress: Baseline (5 min. calculated out of 10 min.), during TSST, recovery  Pain: Baseline, during TSST, recovery (following 10 min. after TSST)  **Stress reactivity and recovery; pain during and after stressor** | FM (ws);  *n* = 18 (18F/0M); 57.1 (6.2) y | *n* = 38 (38F/0M); 48.7 (8.4) y | HR, BP continuously with plethysmography | PPTh and PPTo on right epicondyle with a pressure algometer | Baseline:  Sign group diff (CPP<HC)  During stressor:  Sign group diff (CPP<HC)  ↑ SBP and HR in both groups (*p < .*001)  Sign group-time interaction: CPP<HC (*p =* .029)  Recovery:  Sign group diff  **SBP**  Recovery>baseline in both groups (*p < .*001)  **HR**  Recovery>baseline only in CPP (*p =* .019) | **PPTh**  Only for HC: baseline>during stressor (*p =* .021)  During stressor>recovery (*p =* .012)  **PPTo**  CPP:  sign ↓ between baseline and recovery (*p =* .015) and between stress and recovery (*p =* .004)  HC: sign ↓ between baseline and recovery (*p =* .05), and sign ↑ between stress and recovery (*p =* .032) |
| Maixner et al. (1997) | *Case-control*  Stress: Between 2 pain tasks | TMD (loc);  *n* = 64 (64F/0M, 61 white, 2 black, 1 other minority); 27.2 (1.1) y | *n* = 23 (23F/0M, 21 white, 1 black, 1 other minority); 28.6 (1.5) y | HR, BP with automated blood pressure monitor with a pneumatic cuff around the left ankle | HPTh and HPTo on volar forearm with a 1-cm diameter contact thermode, ischemic PTh and PTo with pressure cuff | **HR, BP** NS group diff | **HPTh, HPTo, ischemic PTh, ischemic PTo**  CPP<HC (*p < .*05) |
| Meeus et al. (2008) | *Case-control*  Stress: Before and immediately after pain induction  **Stress reactivity** | FM (ws);  *n* = 31 (21F/10M); 42.56 (8.61) y for ascending, 47.27 (8.93) y for descending | *n* = 31 (21F/10M); 41.13 (12.7) y for ascending; 46.8 (8.2) for descending | Salivary cortisol before pain induction with radioimmunoassay | CPM with hot water bath (46°C) | Baseline:  **Cortisol**  CPP<HC, NS  After pain:  In both groups (NS difference)  cortisol ↓  The change in cortisol was NS (*p =* .186) | Pain is sign ↑ in CPP compared to HC (*p < .*001) |
| Miyachi et al. (2025) | *Cross-sectional*  Stress: Baseline (1 min.) | CLBP (loc)  *n* = 46 (N.A., mostly women); 67.9 (9.8) y | N.A. | HRV with pulse analyzer (TAS9 VIEW) at index finger | PPTh at lumbar erector spinae muscles with digital algometer (1 cm diameter probe) | N.A. | N.A. |
| Mohn et al. (2008) | *Case-control*  Stress: Baseline (1.5 min.) and after job interview task for 3 min.  Pain: Baseline and after job interview task  **Stress recovery; pain after stressor** | TMD (loc);  *n* = 25 (25F/0M, 100% white); 35.2 (11.9) y | *n* = 25 (25F/0M, 100% white); 33.9 (11.1) y | HR, MAP continuously with Penaz method (Finapres) at the middle phalanx of the third finger of the left hand | EPTh and EPTo with 2 electrodes, PPTh and PPTo with pressure algometer on dorsal part of hand | Baseline:  **MAP**  CPP>HC (*p < .*001)  **HR**  NS  Recovery:  **MAP and HR**  ↑ Sign (*p < .*05) | Baseline & after stressor:  NS group diff (*p >* .05) |
| Muhtz et al. (2013) | *Case-control*  Stress: Baseline and 45 min. and 60 min. after pain testing  **Stress recovery** | CBP (loc), *n* = 20 (12F/8M); 44.9 (14.6) y | *n* = 33 (21F/12M); 33.3 (12) y | Salivary cortisol with radioimmunoassay | Pain rating for heat stimuli on forearm with TSA II (30 x 30 mm thermode) | Baseline:  **AUC cortisol** CPP<HC (*p <* .01)  After pain: **AUC cortisol**  NS ↑ across groups | **Pain rating**  CPP<HC (*p =* .44) |
| Murray et al. (2004) | *Case-control*  Stress and pain: Baseline (N.A.), during stress (CPT and psychological) for 10 min., recovery for 10 min. after stress  **Stress reactivity and recovery** | IBS (loc);  *n* = 24 (20F/4M, 14 white, 7 asian, 3 black); 40 (N.A.) y | *n* = 12 (8F/4M, 6 white, 5 asian, 1 black); 32 (N.A.) y | HR, MAP with a Dinamap | EPTh in anal canal with a 1-cm bipolar electrode mounted on a a14-gauge Foely catheter | Baseline: NS group diff in HR or MAP  During stressor: ↑ sign in HR and MAP during CPT in both groups (*p < .*05)  ↑ sign in HR during psychological stress in both groups (*p < .*05) | Baseline:  NS group diff (*p =* .89)  During stressor:  CPP<HC: ↓ (*p < .*001) |
| Nees et al. (2019) | *Case-control*  Stress: Baseline | CBP (loc);  *n* = 22 (12F/10M); 44 (13.23) y | *n* = 30 (16F/14M); 41.01 (16.21) y | Salivary cortisol throughout the day (+15, 30, 45, 60 min. after awakening; 11:00, 13:00, 15:00, 18:00) with a commercially available immunoassay | CPTh and HPTh with TSA 2001 II (9.0 cm^2^ contact thermode), PPTh on forearm with pressure gauge device,  WUR with pinprick | **Cortisol** NS group effect (*p >* .05) | **CPTh, HPTh, PPTh, WUR** NS group effect (*p >* .05) |
| Ozgocmen et al. (2006) | *Case-control*  Stress: Baseline | FM (ws);  *n* = 29 (29F/0M); 39.6 (8.6) y | *n* = 22 (22F/0M); 34.6 (9.4) y | RRI with equipment from Dantec with 2 electrodes on the chest or alternatively on the dorsum of the hand, SSR with a standard electromyographic electrode to the palm and the sole | PPTh on 18 tender points and 3 control points with mechanical algometer | **RRI, SSR**  NS group effects (*p >* .05) | N.A. |
| Pardo et al. (2019) | *Case-control*  Stress: Baseline, during and after cold room exposure (3.5h)  Pain: Baseline and after cold room exposure  **Stress reactivity; pain during/after stressor** | FM (ws);  *n* = 13 (13F/0M, 100% white); 38 (9.58) y | *n* = 11 (11F/0M, 100% white); 25.73 (7.07) y | HR, BP with Spot Vital Signs Model, serum cortisol using a competitive enzyme immunoassay | PPTh on tender points with a Compact Digital Force Gauge | Baseline:  **BP and HR**  NS group diff (*p >* .05)  During stressor:  **SBP**  ↑ in HC (*p =* .002), not in CPP (*p =* .07)  Recovery:  **HR-SBP-DBP**  NS group diff after 1h  **DBP**  ↑ 1h after stressor in HC (*p =* .004) and CPP (*p = .*0013)  **Cortisol**  NS group diff | Baseline: **PPTh** CPP<HC (*p =* .029)  After stressor: **PPTh**  NS effect of stressor on pain, consistent diff between groups (*p =* .029) |
| Pickering et al. (2019) | *Case-control*  Stress: Baseline | FM (ws);  *n* = 24 (24F/0M); 51 (9) y | *n* = 24 (24F/0M); 51 (10) y | SC | CPTh, HPTh, CPM with advanced thermal stimulator (30 x 30 mm) thermode on forearm as test stimulus and warm water bath (46.5°C) as conditioning stimulus | **SC**  CPP<HC (*p =* .003) | **HPTh**  CPP<HC (*p < .*001)  **CPTh**  CPP<HC (*p < .*001)  **CPM**  Not functional in CPP compared to HC (*p =* .001) |
| Poli Neto et al. (2020) | *Case-control*  Stress: Baseline | Chronic pelvic pain (loc);  *n* = 21 (21F/0M); 28.2 (6.0) y | *n* = 21 (21F/0M); 28.0 (7.3) y | HR, BP with digital device (OMRON) | PPTh on forearm with pressure algometer | **HR**  CPP>HC (*p =* .013)  **SBP** CPP>HC (*p =* .170)  **DBP**  CPP>HC (*p =* .006)  **Average BP** CPP>HC (*p =* .014) | **PPTh** CPP<HC: *p < .*0001 |
| Quartana et al. (2010) | *Case-control*  Stress: Baseline, reactivity to pain induction, recovery of pain induction (20 min. after CPT)  **Stress reactivity and recovery** | TMD (loc);  *n* = 39 (32F/7M, 79.5% white, 7.7% African American, 7/7% Asian American, 5.1% multi-racial); 33.79 (12.00) y | *n* = 22 (21F/1M, 4.9% white, 27.3% African American, 22.7% Asian American, 4.5% multi-racial); 25.91 (5.76) y | Salivary cortisol measured ± 52 min. after awakening with commercially available enzyme immunoassay | PPTh on masseter, forearm and trapezius with somedic algometer, HPTh on ventral forearm with TDA-2001 (30 x 30 mm thermode), cold pain ratings by immersing hand in cold water of 4°C | N.A. | PPTh **Masseter muscle**  CPP<HC (*p < .*05)  **Trapezius muscle**  NS group diff  HPTh  NS group diff  Cold pain  NS group diff |
| Rampazo et al. (2024) | *Case-control*  Stress: during CPM  **Stress reactivity** | CNP (loc); *n* = 25 (16F/9M); 31.9 (11.0) y | *n* = 25 (16F/9M); 29.4 (6.6) y | HRV with Polar 800 CX heart monitor | CPM with PPTh on upper trapezius muscle as test stimulus, cold pressor test as conditioning stimulus | **HR**  NS group diff (*p* > .05) | **CPM**  NS group diff (*p* = .148) |
| Reshkova et al. (2015) | *Cross-sectional*  Stress: Baseline | FM (ws);  *n* = 21  (N.A.) |  | Adrenaline and noradrenaline with enzyme linked immunosorbent assay | PPTh with Fisher dolorimeter in all trigger points | N.A. | N.A. |
| Scheuren et al. (2023) | *Case-control*  Stress: during pain induction  **Stress reactivity** | Chronic CRPS (loc); *n* = 20 (17F/3M); 44.9 (12.8) y | *n* = 16 (14F/2M); 41.8 (13.3) y | SSR with electrodes to the hand palm and the reference electrode to the hand dorsum | Heat stimuli with CHEPs thermode (27 mm diameter), pinprick stimuli, MPTh | **SSR** CRPS>HC (*p* < *.*01) for affected side | **Pinprick**  CPP>HC (affected: *p =* .02), not in controls (*p =* .87)  **MPTh**  NS group diff (affected: *p =* .31; control: *p =* .57)  **HPTh**  NS group diff (affected: *p =* .14; control: *p =* .16) |
| Tan et al. (2023) | *Case-control*  Stress: Baseline | CLBP (loc);  *n* = 16 (8F/8M); 33.31 (13.4) y | *n* = 16 (8F/8M); 32.69 (13.01) y | SC on eight detection points with diode laser THOR LX2 | PPTh with digital algometer | **SC** NS group diff (*p* < .05) | **PPTh**  CPP<HC for all points (*p* < .05) |
| Thieme et al. (2022) | *Case-control*  Stress: Baseline | FM (ws);  *n* = 32 (32F/0M); N.A. | *n* = 30 (30F/0M); N.A. | BP with Finapres attached to the middle finger of the left hand, HRV (TP, HF, LF, Very LF, SSDN, RMSSD) with a LabLinc V modular Instrument (ECG) | EPTh, EPTo with electrodes to the index and ring finger of the right hand | Baseline:  **HRV** (TP, HF, LF, Very LF, SSDN, RMSSD)  CPP<HC (*p < .*003) | **EPTh**  CPP<HC (*p < .*001)  **EPTo**  CPP<HC  (*p < .*001) |
| Umeda et al. (2013) | *Case-control*  Stress: Baseline | FM (ws);  *n* = 8 (8F/0M); 47.63 (16.46) y | *n* = 14 (14F/0M); 41.93 (11.46) y | HR, MAP with monitor (OMRON) | NFR on BF muscle with 2 8-mm Ag-AgCl recording electrodes | **HR**  CPP>HC (*p =* .02)  **MAP**  NS group diff (*p <* .05) | **NFR** **threshold** NS diff between responders (*p =* .35) |
| Valera-Calero et al. (2022) | *Cross-sectional*  Stress: Baseline | CNP (loc);  *n* = 94 (74F/20M); 38.1 (8.7) y | N.A. | Salivary cortisol between 10:00 – 10:30 with enzyme-linked immunosorbent assay | PPTh on zygapophyseal joint with pressure algometer | N.A. | N.A. |
| Van Den Houte et al. (2018) | *Case-control*  Stress: Baseline (10 min.) | FM (ws);  *n* = 81 (71F/10M); 42.28 (10.78) y | *n* = 41 (36F/5M); 40.52 (10.87) y | HRV (RMSSD) with ECG with electrodes below right and left clavicle and at the left lower ribs | CPM with electrical stimuli (test stimulus) on right ankle with 2 8-mm Ag-AgCl electrodes, cold pain (conditioning stimulus) on forearm with a cold pressor arm wrap | **RMSSD** NS group diff (*p =* .90) | **CPM**  NS group diff (*p =* .27) |
| Van Middendorp et al. (2013) | *Case-control*  Stress: Baseline (i.e., neutral recall; most reliable 90 sec. of 120 sec.) | FM (ws);  *n* = 62 (62F/0M); 46.3 (10.8) y | *n* = 59 (59F/0M); 48.9 (11.4) y | HR, MAP, PEP, with Biopac systems MP150 ECG at lead II configuration | EPTh, EPTo on inner side of forearm with a Tursky concentric electrode | **HR**  CPP>HC (*p =* .002)  **MAP**  CPP<HC (*p =* .030) | N.A. |
| Venezia et al. (2024) | *Case-control*  Stress: Baseline (5 min.), reactivity to cold pain stimulation  **Stress reactivity** | CLBP (loc);  *n* = 22 (14F/8M); 39.2 (11.7) y | *n =* 29 (13F/16M); 33.7 (10.8) y | BRS, HR, BP continuously with CareTaker device placed on the right finger of the right hand | PPTh on thumbnail, CPM with pressure stimuli on thumbnails | **BRS**  NS group diff in change in BRS due to cold stimulation | **PPTh**  NS group diff (*p* > .369) |
| Wingenfeld et al. (2010) | *Case-control*  Stress: Baseline | FM (ws);  *n* = 23 (23F/0M); 49.1 (8.7) y | *n* = 26 (26F/0M); 46.5 (6.1) y | Salivary cortisol +15, 30, 45, 60 min. after awakening; 8:00,11:00, 15:00, 20:00 with time-resolved immunoassay with fluorometric detection | PPTh with hand-held dolorimeter and HPTh with pain and thermal sensitivity tester (9 cm^2^ thermode) on 1 tender point (trapezius) and 1 control point (volar forearm) | **AUC cortisol** NS group diff (*p =* .176)  NS diff in slope between groups (*p =* .37) | **HPTh**  NS group diff  **PPTh**  CPP<HC (*p < .*002) |
| Woda et al. (2013) | *Case-control*  Stress: Baseline (N.A.), reactivity and recovery from TSST (10 min. and 30 min. after TSST)  **Stress reactivity and recovery** | FM (ws);  *n* = 31 (31F/0M); 47.39 (1.79) y | *n* = 32 (32F/0M); 36.34 (2.32) y | HR, BP with an automatic blood pressure monitor (OMRON) | Pain rating after palpation on masseter and trapezius muscle | Baseline:  **HR and BP**  NS group diff  During stressor:  **DBP** ↑  Sign time effect (*p =* .006), no group effect  **SBP** ↑  Group but not time had a sign effect (*p =* .0451), CPP>HC  **HR**  No time effect. | **Palpation pain** CPP>HC (sign) |
| Zamunér et al. (2016) | *Case-control*  Stress: Baseline (20 min.) | FM (ws);  *n* = 20 (20F/0M); 48.2 (6.1) | *n* = 20 (20F/0M); 45.8 (7.3) y | HR, BP, HRV (RMSSD, SDNN, LF, HF, HFnu, LF/HF) with a Polar | PPTh on 18 tender points with a digital algometer | **RMSSD, SDNN, LF, HF, HFnu HRV**  CPP<HC (*p < .*05)  **LFnu** **and LF/HF** CPP>HC (*p < .*05)  **HR**, **SBP, DBP**  NS group diff (*p =* .64, *p =* .54, *p =* .06) | **PPTh**  Not measured in HC |

Abbreviations. ANS: Autonomic Nervous System, AUC: Area Under the Curve, BP: Blood Pressure, BRS: Baroreflex Sensitivity, C(L)BP: Chronic (Low) Back Pain, CMTH: Chronic Muscle Tension Headache, CNP: Chronic Neck Pain, CPP: Chronic Primary Pain, CPM: Conditioned Pain Modulation, CPT: Cold Pressor Test, CPTh: Cold Pain Threshold, CPTo: Cold Pain Tolerance, CVT: Cardiac Vagal Tone, DBP: Diastolic Blood Pressure, DHEA: Dehydroepiandrosterone, diff: difference, ECG: Electrocardiography, EPTh: Electrical Pain Threshold, EPTo: Electrical Pain Tolerance, F: Female, F: F-statistic, FCP: Functional Chest Pain, FM: Fibromyalgia, HC: Healthy Controls, HF(nu): High Frequency (normal unit) HRV, HPTh: Heat Pain Threshold, HPTo: Heat Pain Tolerance, HR(R): Heart Rate (Recovery), HRV: Heart Rate Variability, IBI: Inter-Beat-Interval, IBS: Irritable Bowel Syndrome, L: Left, LF(nu): Low Frequency (normal unit) HRV, loc: Localized pain, M: Male, MAP: Mean Arterial Pressure, MBP: Mean Blood Pressure, MPTh: Mechanical Pain Threshold, N.A.: Not Applicable, NFR(T): Nociceptive Flexor Reflex (Threshold), NS: Non-Significant, p: probability value, PEP: Pre-Ejection Period, (P)PTh: Pressure Pain Threshold, (P)PTo: Pressure Pain Tolerance, PSD: Total Power Spectral Density, r: correlation coefficient, R: Right, RMSSD: Root mean square of the differences between successive N-N intervals, RRI: R-R Interval, SBP: Systolic Blood Pressure, SCL: Skin Conductance Level, SC(R): Skin Conductance (Response), SD: Standard Deviation, sign: significant, SREP: Slowly Repeated Evoked Pain, SSDN: Standard Deviation of Normal-to-Normal Intervals, SSR: Sympathetic Skin Response, SVB: Sympatho-Vagal Balance, t: t-statistic, TMD: Temporomandibular Disorder, TP: Total Power, TSP: Temporal Summation of Pain, TSST: Trier Social Stress Test, UC: Ulcerative Colitis, VVS: Vulvar Vestibulitis Syndrome, ws: Widespread pain, WUR: Wind-Up Ratio, y: years, Δ: change in
