## Supplementary 7 - Table S5 for "The Stress-Pain Connection in Chronic Primary Pain: A Systematic Review and Meta-Analysis of Physiological Stress Markers in Relation to Experimental Pain Responses"

**Supplementary File 7 - Table S5.** Characteristics of included studies

| **Study** | **Interactions STRESS-PAIN** | |
| --- | --- | --- |
|  | **CPP** | **Controls** |
| Bandeira et al. (2021) | Stress baseline – Pain baseline  **CPP (loc)**  LF & PPTh trap R: *r = -.*07; *p = .*64  HF & PPTh trap R: *r =* .077; *p = .*60  LF/HF & PPTh trap R: *r = -.*008; *p = .*96  LF & PPTh trap L: *r =* .129; *p = .*382  HF & PPTh trap L: *r = -.*121; *p = .*413  LF/HF & PPTh trap L: *r =* .165; *p = .*257  LF & PPTh elbow R: *r = -.*007; *p = .*96  HF & PPTh elbow R: *r =* .061; *p = .*678  LF/HF & PPTh elbow R: *r = -.*083; *p = .*576  LF & PPTh elbow L: *r =* .076; *p = .*607  HF & PPTh elbow L: *r = -.*119; *p = .*419  LF/HF & PPTh elbow L: *r =* .182; *p = .*211  LF & PPTh lumbar R: *r =* .092; *p = .*535  HF & PPTh lumbar R: *r = -.*112; *p = .*446  LF/HF & PPTh lumbar R: *r =* .189; *p = .*194  LF & PPTh lumbar L: *r =* .118; *p = .*423  HF & PPTh lumbar L: *r = -.*15; *p = .*304  LF/HF & PPTh lumbar L: *r =* .184; *p = .*207  LF & PPTh patellar R: *r =* .064; *p = .*663  HF & PPTh patellar R: *r = -.*074; *p = .*616  LF/HF & PPTh patellar R: *r =* .056; *p = .*701  LF & PPTh patellar L: *r =* .01; *p = .*944  HF & PPTh patellar L: *r = -.*099; *p = .*502  LF/HF & PPTh patellar L: *r =* .073; *p = .*619  Δ Stress (baseline to during) – Pain baseline  **CPP (loc)**  LF & PPTh trap R: *r = -.*032; *p = .*83  HF & PPTh trap R: *r =* .028; *p = .*85  LF/HF & PPTh trap R: *r* = -.065; *p = .*662  LF & PPTh trap L: *r = -.*038; *p = .*798  HF & PPTh trap L: *r =* .041; *p = .*781  LF/HF & PPTh trap L: *r = -.*073; *p = .*625  LF & PPTh elbow R: *r =* .086; *p = .*569  HF & PPTh elbow R: *r = -.*092; *p = .*543  LF/HF & PPTh elbow R: *r =* .091; *p = .*545  LF & PPTh elbow L: *r = -.*066; *p = .*652  HF & PPTh elbow L: *r =* .071; *p = .*628  LF/HF & PPTh elbow L: *r = -.*051; *p = .*724  LF & PPTh lumbar R: *r =* .024; *p = .*871  HF & PPTh lumbar R: *r = -.*019; *p = .*897  LF/HF & PPTh lumbar R: *r =* .035; *p = .*809  LF & PPTh lumbar L: *r =* .102; *p = .*479  HF & PPTh lumbar L: *r = -.*114; *p = .*43  LF/HF & PPTh lumbar L: *r =* .065; *p = .*66  LF & PPTh patellar R: *r =* .087; *p = .*564  HF & PPTh patellar R: *r = -.*073; *p = .*619  LF/HF & PPTh patellar R: *r =* .065; *p = .*658  LF & PPTh patellar L: *r =* .014; *p = .*916  HF & PPTh patellar L: *r =* .005; *p = .*971  LF/HF & PPTh patellar L: *r = -.*031; *p = .*832  Δ Stress (baseline to after) – Pain baseline  **CPP (loc)**  LF & PPTh trap R: *r =* .028; *p = .*852  HF & PPTh trap R: *r = -.*03; *p = .*839  LF/HF & PPTh trap R: r = 0.114; *p = .*439  LF & PPTh trap L: *r = -.*047; *p = .*752  HF & PPTh trap L: *r =* .049; *p = .*738  LF/HF & PPTh trap L: *r = -.*013; *p = .*921  LF & PPTh elbow R: *r =* .035; *p = .*816  HF & PPTh elbow R: *r = -.*041; *p = .*781  LF/HF & PPTh elbow R: *r =* .097; *p = .*513  LF & PPTh elbow L: *r =* .011; *p = .*931  HF & PPTh elbow L: *r =* .025; *p = .*861  LF/HF & PPTh elbow L: *r = -.*067; *p = .*648  LF & PPTh lumbar R: *r = -.*023; *p = .*871  HF & PPTh lumbar R: *r = -.*011; *p = .*932  LF/HF & PPTh lumbar R: *r =* .038; *p = .*80  LF & PPTh lumbar L: *r =* .028; *p = .*847  HF & PPTh lumbar L: *r = -.*037; *p = .*803  LF/HF & PPTh lumbar L: *r =* .067; *p = .*65  LF & PPTh patellar R: *r = -.*061; *p = .*679  HF & PPTh patellar R: *r =* .064; *p = .*664  LF/HF & PPTh patellar R: *r =* .03; *p = .*836  LF & PPTh patellar L: *r =* .065; *p = .*663  HF & PPTh patellar L: *r = -.*029; *p = .*841  LF/HF & PPTh patellar L: *r =* .098; *p = .*51  Stress baseline - Δ Pain (baseline to after):  **CPP (loc)**  LF & PPTh trap R: *r =* .152; *p = .*302  HF & PPTh trap R: *r = -.*177; *p = .*229  LF/HF & PPTh trap R: *r* = .031; *p = .*835  LF & PPTh trap L: *r = -.*133; *p = .*369  HF & PPTh trap L: *r =* .123; *p = .*406  LF/HF & PPTh trap L: *r = -.*029; *p = .*843  LF & PPTh elbow R: *r = -.*04; *p = .*786  HF & PPTh elbow R: *r =* .027; *p = .*853  LF/HF & PPTh elbow R: *r = -.*073; *p = .*621  LF & PPTh elbow L: *r =* .024; *p = .*867  HF & PPTh elbow L: *r = -.*081; *p = .*585  LF/HF & PPTh elbow L: *r =* .064; *p = .*664  LF & PPTh lumbar R: *r = -.*047; *p = .*751  HF & PPTh lumbar R: *r = -.*001; *p = .*996  LF/HF & PPTh lumbar R: *r = -.*002; *p = .*991  LF & PPTh lumbar L: *r =* .097; *p = .*511  HF & PPTh lumbar L: *r =* .061; *p = .*679  LF/HF & PPTh lumbar L: *r =* .047; *p = .*753  LF & PPTh patellar R: *r =* .065; *p = .*661  HF & PPTh patellar R: *r = -.*018; *p = .*896  LF/HF & PPTh patellar R: *r =* .043; *p = .*769  LF & PPTh patellar L: *r = -.*02; *p = .*889  HF & PPTh patellar L: *r =* .065; *p = .*662  LF/HF & PPTh patellar L: *r = -.*024; *p = .*868  Δ Stress (baseline to during) - Δ Pain (baseline to after):  **CPP (loc)**  LF & PPTh trap R: *r =* .065; *p = .*659  HF & PPTh trap R: *r = -.*062; *p = .*677  LF/HF & PPTh trap R: r = -0.065; *p = .*661  LF & PPTh trap L: *r =* .042; *p = .*778  HF & PPTh trap L: *r =* .048; *p = .*745  LF/HF & PPTh trap L: *r = -.*014; *p = .*919  LF & PPTh elbow R: *r =* .011; *p = .*93  HF & PPTh elbow R: *r = -.*028; *p = .*852  LF/HF & PPTh elbow R: *r = -.*041; *p = .*781  LF & PPTh elbow L: *r =* .081; *p = .*579  HF & PPTh elbow L: *r = -.*071; *p = .*623  LF/HF & PPTh elbow L: *r =* .061; *p = .*676  LF & PPTh lumbar R: *r = -.*054; *p = .*723  HF & PPTh lumbar R: *r =* .019; *p = .*897  LF/HF & PPTh lumbar R: *r = -.*067; *p = .*649  LF & PPTh lumbar L: *r =* .065; *p = .*66  HF & PPTh lumbar L: *r = -.*047; *p = .*748  LF/HF & PPTh lumbar L: *r =* .028; *p = .*855  LF & PPTh patellar R: *r =* .021; *p = .*888  HF & PPTh patellar R: *r = -.*011; *p = .*931  LF/HF & PPTh patellar R: *r =* .046; *p = .*752  LF & PPTh patellar L: *r =* .054; *p = .*721  HF & PPTh patellar L: *r = -.*034; *p = .*82  LF/HF & PPTh patellar L: *r =* .032; *p = .*832  Δ Stress (baseline to after) - Δ Pain (baseline to after):  **CPP (loc)**  LF & PPTh trap R: *r =* .016; *p = .*916  HF & PPTh trap R: *r = -.*016; *p = .*913  LF/HF & PPTh trap R: r = -0.137; p = 0.355  LF & PPTh trap L: *r = -.*147; *p = .*32  HF & PPTh trap L: *r =* .15; *p = .*31  LF/HF & PPTh trap L: *r = -.*041; *p = .*784  LF & PPTh elbow R: *r =* .024; *p = .*874  HF & PPTh elbow R: *r = -.*011; *p = .*932  LF/HF & PPTh elbow R: *r =* .048; *p = .*749  LF & PPTh elbow L: *r =* .043; *p = .*774  HF & PPTh elbow L: *r =* .067; *p = .*652  LF/HF & PPTh elbow L: *r =* .012; *p = .*926  LF & PPTh lumbar R: *r = -.*061; *p = .*678  HF & PPTh lumbar R: *r =* .046; *p = .*756  LF/HF & PPTh lumbar R: *r = -.*084; *p = .*576  LF & PPTh lumbar L: *r =* .03; *p = .*839  HF & PPTh lumbar L: *r = -.*057; *p = .*704  LF/HF & PPTh lumbar L: *r =* .065; *p = .*661  LF & PPTh patellar R: *r =* .064; *p = .*666  HF & PPTh patellar R: *r = -.*045; *p = .*76  LF/HF & PPTh patellar R: *r =* .012; *p = .*929  LF & PPTh patellar L: *r =* .024; *p = .*875  HF & PPTh patellar L: *r =* .011; *p = .*933  LF/HF & PPTh patellar L: *r = -.*042; *p = .*779  Stress during - Pain baseline:  **CPP (loc)**  LF & PPTh trap R: *r = -.*113; *p = .*444 HF & PPTh trap R: *r =* .117; *p = .*427 LF/HF & PPTh trap R: *r = -.*085; *p = .*566  LF & PPTh trap L: *r =* .118; *p = .*426 HF & PPTh trap L: *r = -.*105; *p = .*479 LF/HF & PPTh trap L: *r =* .064; *p = .*661  LF & PPTh elbow R: *r =* .024; *p = .*875 HF & PPTh elbow R: *r =* .011; *p = .*932 LF/HF & PPTh elbow R: *r = -.*064; *p = .*665  LF & PPTh elbow L: *r = -.*037; *p = .*805 HF & PPTh elbow L: *r = -.*041; *p = .*784 LF/HF & PPTh elbow L: *r =* .071; *p = .*627  LF & PPTh lumbar R: *r =* .065; *p = .*66 HF & PPTh lumbar R: *r = -.*058; *p = .*695 LF/HF & PPTh lumbar R: *r =* .038; *p = .*803  LF & PPTh lumbar L: *r = -.*049; *p = .*739 HF & PPTh lumbar L: *r =* .051; *p = .*735 LF/HF & PPTh lumbar L: *r =* .092; *p = .*546  LF & PPTh patellar R: *r =* .054; *p = .*721 HF & PPTh patellar R: *r = -.*012; *p = .*931 LF/HF & PPTh patellar R: *r =* .021; *p = .*887  LF & PPTh patellar L: *r = -.*024; *p = .*872 HF & PPTh patellar L: *r =* .065; *p = .*663 LF/HF & PPTh patellar L: *r = -.*029; *p = .*85  Stress after - Pain baseline:  **CPP (loc)**  LF & PPTh trap R: *r = -.*027; *p = .*853 HF & PPTh trap R: *r =* .03; *p = .*837 LF/HF & PPTh trap R: *r =* .109; *p = .*462  LF & PPTh trap L: *r =* .056; *p = .*707 HF & PPTh trap L: *r = -.*045; *p = .*76 LF/HF & PPTh trap L: *r =* .041; *p = .*778  LF & PPTh elbow R: *r = -.*038; *p = .*803 HF & PPTh elbow R: *r =* .064; *p = .*666 LF/HF & PPTh elbow R: *r = -.*027; *p = .*855  LF & PPTh elbow L: *r =* .012; *p = .*93 HF & PPTh elbow L: *r =* .054; *p = .*722 LF/HF & PPTh elbow L: *r =* .047; *p = .*749  LF & PPTh lumbar R: *r = -.*041; *p = .*781 HF & PPTh lumbar R: *r =* .031; *p = .*838 LF/HF & PPTh lumbar R: *r = -.*065; *p = .*663  LF & PPTh lumbar L: *r =* .048; *p = .*748 HF & PPTh lumbar L: *r = -.*058; *p = .*695 LF/HF & PPTh lumbar L: *r =* .011; *p = .*936  LF & PPTh patellar R: *r =* .041; *p = .*781 HF & PPTh patellar R: *r = -.*045; *p = .*758 LF/HF & PPTh patellar R: *r =* .048; *p = .*744  LF & PPTh patellar L: *r = -.*011; *p = .*936 HF & PPTh patellar L: *r =* .031; *p = .*837 LF/HF & PPTh patellar L: *r =* .012; *p = .*929  Stress baseline - Pain after:  **CPP (loc)**  LF & PPTh trap R: *r =* .037; *p = .*802 HF & PPTh trap R: *r = -.*047; *p = .*749 LF/HF & PPTh trap R: *r =* .012; *p = .*933  LF & PPTh trap L: *r = -.*011; *p = .*94 HF & PPTh trap L: *r =* .009; *p = .*951 LF/HF & PPTh trap L: *r = -.*028; *p = .*852  LF & PPTh elbow R: *r =* .031; *p = .*837 HF & PPTh elbow R: *r =* .029; *p = .*849 LF/HF & PPTh elbow R: *r = -.*021; *p = .*888  LF & PPTh elbow L: *r = -.*034; *p = .*822 HF & PPTh elbow L: *r = -.*029; *p = .*849 LF/HF & PPTh elbow L: *r =* .056; *p = .*706  LF & PPTh lumbar R: *r =* .041; *p = .*782 HF & PPTh lumbar R: *r =* .034; *p = .*821 LF/HF & PPTh lumbar R: *r =* .024; *p = .*874  LF & PPTh lumbar L: *r = -.*019; *p = .*898 HF & PPTh lumbar L: *r = -.*012; *p = .*931 LF/HF & PPTh lumbar L: *r =* .031; *p = .*836  LF & PPTh patellar R: *r = -.*048; *p = .*745 HF & PPTh patellar R: *r =* .027; *p = .*855 LF/HF & PPTh patellar R: *r = -.*012; *p = .*929  LF & PPTh patellar L: *r =* .034; *p = .*82 HF & PPTh patellar L: *r = -.*039; *p = .*799 LF/HF & PPTh patellar L: *r =* .048; *p = .*744  Stress during - Pain after:  **CPP (loc)**  LF & PPTh trap R: *r = -.*02; *p = .*895 HF & PPTh trap R: *r =* .002; *p = .*988 LF/HF & PPTh trap R: *r =* .002; *p = .*99  LF & PPTh trap L: *r = -.*008; *p = .*959 HF & PPTh trap L: *r =* .002; *p = .*988 LF/HF & PPTh trap L: *r = -.*027; *p = .*86  LF & PPTh elbow R: *r =* .041; *p = .*781 HF & PPTh elbow R: *r = -.*031; *p = .*837 LF/HF & PPTh elbow R: *r =* .021; *p = .*888  LF & PPTh elbow L: *r =* .034; *p = .*822 HF & PPTh elbow L: *r = -.*029; *p = .*849 LF/HF & PPTh elbow L: *r =* .065; *p = .*66  LF & PPTh lumbar R: *r =* .038; *p = .*803 HF & PPTh lumbar R: *r = -.*051; *p = .*734 LF/HF & PPTh lumbar R: *r = -.*028; *p = .*855  LF & PPTh lumbar L: *r =* .051; *p = .*735 HF & PPTh lumbar L: *r = -.*029; *p = .*85 LF/HF & PPTh lumbar L: *r =* .031; *p = .*837  LF & PPTh patellar R: *r = -.*048; *p = .*745 HF & PPTh patellar R: *r =* .027; *p = .*855 LF/HF & PPTh patellar R: *r = -.*012; *p = .*929  LF & PPTh patellar L: *r =* .048; *p = .*744 HF & PPTh patellar L: *r = -.*021; *p = .*888 LF/HF & PPTh patellar L: *r =* .012; *p = .*93  Stress after - Pain after:  **CPP (loc)**  LF & PPTh trap R: *r =* .049; *p = .*739 HF & PPTh trap R: *r = -.*060; *p = .*685 LF/HF & PPTh trap R: *r =* .188; *p = .*202  LF & PPTh trap L: *r =* .111; *p = .*452 HF & PPTh trap L: *r = -.*113; *p = .*443 LF/HF & PPTh trap L: *r =* .054; *p = .*714  LF & PPTh elbow R: *r = -.*034; *p = .*822 HF & PPTh elbow R: *r =* .029; *p = .*849 LF/HF & PPTh elbow R: *r =* .064; *p = .*664  LF & PPTh elbow L: *r =* .012; *p = .*93 HF & PPTh elbow L: *r =* .054; *p = .*722 LF/HF & PPTh elbow L: *r = -.*031; *p = .*837  LF & PPTh lumbar R: *r =* .041; *p = .*782 HF & PPTh lumbar R: *r = -.*065; *p = .*66 LF/HF & PPTh lumbar R: *r =* .021; *p = .*888  LF & PPTh lumbar L: *r = -.*048; *p = .*748 HF & PPTh lumbar L: *r =* .065; *p = .*661 LF/HF & PPTh lumbar L: *r = -.*031; *p = .*836  LF & PPTh patellar R: *r = -.*048; *p = .*746 HF & PPTh patellar R: *r =* .027; *p = .*855 LF/HF & PPTh patellar R: *r = -.*012; *p = .*93  LF & PPTh patellar L: *r =* .048; *p = .*744 HF & PPTh patellar L: *r = -.*021; *p = .*888 LF/HF & PPTh patellar L: *r =* .012; *p = .*93  Stress during - Δ Pain:  **CPP (loc)**  LF & PPTh trap R: *r = -.*119; *p = .*42 HF & PPTh trap R: *r =* .152; *p = .*302 LF/HF & PPTh trap R: *r = -.*116; *p = .*433  LF & PPTh trap L: *r =* .118; *p = .*423 HF & PPTh trap L: *r = -.*100; *p = .*498 LF/HF & PPTh trap L: *r =* .072; *p = .*622  LF & PPTh elbow R: *r =* .044; *p = .*772 HF & PPTh elbow R: *r = -.*062; *p = .*673 LF/HF & PPTh elbow R: *r =* .034; *p = .*822  LF & PPTh elbow L: *r = -.*028; *p = .*853 HF & PPTh elbow L: *r =* .032; *p = .*834 LF/HF & PPTh elbow L: *r = -.*061; *p = .*678  LF & PPTh lumbar R: *r =* .012; *p = .*927 HF & PPTh lumbar R: *r = -.*021; *p = .*888 LF/HF & PPTh lumbar R: *r =* .031; *p = .*836  LF & PPTh lumbar L: *r =* .031; *p = .*836 HF & PPTh lumbar L: *r =* .047; *p = .*75 LF/HF & PPTh lumbar L: *r =* .024; *p = .*871  LF & PPTh patellar R: *r =* .065; *p = .*661 HF & PPTh patellar R: *r = -.*051; *p = .*733 LF/HF & PPTh patellar R: *r =* .011; *p = .*934  LF & PPTh patellar L: *r = -.*021; *p = .*887 HF & PPTh patellar L: *r =* .027; *p = .*856 LF/HF & PPTh patellar L: *r =* .041; *p = .*78  Stress after - Δ Pain (baseline to after):  **CPP (loc)**  LF & PPTh trap R: *r = -.*115; *p = .*438 HF & PPTh trap R: *r =* .136; *p = .*358 LF/HF & PPTh trap R: *r = -.*153; *p = .*299  LF & PPTh trap L: *r = -.*065; *p = .*663 HF & PPTh trap L: *r =* .076; *p = .*605 LF/HF & PPTh trap L: *r = -.*043; *p = .*772  LF & PPTh elbow R: *r = -.*037; *p = .*805 HF & PPTh elbow R: *r =* .029; *p = .*849 LF/HF & PPTh elbow R: *r =* .048; *p = .*746  LF & PPTh elbow L: *r =* .054; *p = .*721 HF & PPTh elbow L: *r =* .032; *p = .*835 LF/HF & PPTh elbow L: *r = -.*041; *p = .*781  LF & PPTh lumbar R: *r =* .065; *p = .*661 HF & PPTh lumbar R: *r = -.*052; *p = .*733 LF/HF & PPTh lumbar R: *r =* .021; *p = .*888  LF & PPTh lumbar L: *r = -.*031; *p = .*836 HF & PPTh lumbar L: *r =* .031; *p = .*837 LF/HF & PPTh lumbar L: *r = -.*048; *p = .*745  LF & PPTh patellar R: *r =* .065; *p = .*66 HF & PPTh patellar R: *r =* .011; *p = .*935 LF/HF & PPTh patellar R: *r = -.*012; *p = .*93  LF & PPTh patellar L: *r =* .048; *p = .*746 HF & PPTh patellar L: *r =* .027; *p = .*856 LF/HF & PPTh patellar L: *r =* .041; *p = .*78  Δ Stress (baseline to during) - Pain after:  **CPP (loc)**  LF & PPTh trap R: *r = -.*068; *p = .*647 HF & PPTh trap R: *r =* .062; *p = .*674 LF/HF & PPTh trap R: *r = -.*013; *p = .*928  LF & PPTh trap L: *r =* .006; *p = .*967 HF & PPTh trap L: *r = -.*009; *p = .*951 LF/HF & PPTh trap L: *r =* .034; *p = .*822  LF & PPTh elbow R: *r =* .012; *p = .*93 HF & PPTh elbow R: *r =* .054; *p = .*722 LF/HF & PPTh elbow R: *r =* .048; *p = .*746  LF & PPTh elbow L: *r = -.*031; *p = .*837 HF & PPTh elbow L: *r = -.*029; *p = .*85 LF/HF & PPTh elbow L: *r =* .021; *p = .*888  LF & PPTh lumbar R: *r =* .054; *p = .*722 HF & PPTh lumbar R: *r =* .041; *p = .*780 LF/HF & PPTh lumbar R: *r =* .028; *p = .*855  LF & PPTh lumbar L: *r =* .061; *p = .*677 HF & PPTh lumbar L: *r = -.*034; *p = .*822 LF/HF & PPTh lumbar L: *r =* .041; *p = .*78  LF & PPTh patellar R: *r =* .065; *p = .*661 HF & PPTh patellar R: *r = -.*048; *p = .*746 LF/HF & PPTh patellar R: *r =* .031; *p = .*837  LF & PPTh patellar L: *r =* .021; *p = .*888 HF & PPTh patellar L: *r =* .048; *p = .*746 LF/HF & PPTh patellar L: *r =* .029; *p = .*849  Δ Stress (baseline to after) - Pain after:  **CPP (loc)**  LF & PPTh trap R: *r =* .013; *p = .*928 HF & PPTh trap R: *r = -.*015; *p = .*92 LF/HF & PPTh trap R: *r =* .182; *p = .*216  LF & PPTh trap L: *r =* .098; *p = .*508 HF & PPTh trap L: *r = -.*099; *p = .*505 LF/HF & PPTh trap L: *r =* .054; *p = .*721  LF & PPTh elbow R: *r =* .054; *p = .*721 HF & PPTh elbow R: *r = -.*061; *p = .*680 LF/HF & PPTh elbow R: *r =* .045; *p = .*761  LF & PPTh elbow L: *r =* .011; *p = .*937 HF & PPTh elbow L: *r =* .031; *p = .*838 LF/HF & PPTh elbow L: *r =* .019; *p = .*897  LF & PPTh lumbar R: *r = -.*031; *p = .*838 HF & PPTh lumbar R: *r =* .038; *p = .*803 LF/HF & PPTh lumbar R: *r = -.*065; *p = .*661  LF & PPTh lumbar L: *r =* .048; *p = .*746 HF & PPTh lumbar L: *r = -.*052; *p = .*733 LF/HF & PPTh lumbar L: *r =* .021; *p = .*887  LF & PPTh patellar R: *r = -.*048; *p = .*746 HF & PPTh patellar R: *r =* .027; *p = .*856 LF/HF & PPTh patellar R: *r =* .031; *p = .*837  LF & PPTh patellar L: *r = -.*031; *p = .*837 HF & PPTh patellar L: *r =* .031; *p = .*837 LF/HF & PPTh patellar L: *r =* .054; *p = .*721 | Stress baseline – Pain baseline  **HC**  LF & PPTh trap R: *r =* .032; *p = .*827  HF & PPTh trap R: *r = -.*032; *p = .*827  LF/HF & PPTh trap R: *r =* .084; *p = .*568  LF & PPTh trap L: *r = -.*024; *p = .*87  HF & PPTh trap L: *r =* .007; *p = .*961  LF/HF & PPTh trap L: *r = -.*062; *p = .*674  LF & PPTh elbow R: *r =* .111; *p = .*448  HF & PPTh elbow R: *r = -.*017; *p = .*902  LF/HF & PPTh elbow R: *r =* .122; *p = .*405  LF & PPTh elbow L: *r = -.*019; *p = .*892  HF & PPTh elbow L: *r = -.*071; *p = .*632  LF/HF & PPTh elbow L: *r =* .081; *p = .*585  LF & PPTh lumbar R: *r = -.*007; *p = .*963  HF & PPTh lumbar R: *r =* .042; *p = .*775  LF/HF & PPTh lumbar R: *r = -.*041; *p = .*782  LF & PPTh lumbar L: *r =* .025; *p = .*866  HF & PPTh lumbar L: *r =* .021; *p = .*885  LF/HF & PPTh lumbar L: *r =* .004; *p = .*981  LF & PPTh patellar R: *r = -.*071; *p = .*632  HF & PPTh patellar R: *r =* .011; *p = .*935  LF/HF & PPTh patellar R: *r = -.*081; *p = .*586  LF & PPTh patellar L: *r =* .121; *p = .*411  HF & PPTh patellar L: *r = -.*082; *p = .*581  LF/HF & PPTh patellar L: *r =* .173; *p = .*269  Δ Stress (baseline to during) – Pain baseline  **HC**  LF & PPTh trap R: *r =* .029; *p = .*844  HF & PPTh trap R: *r =* .063; *p = .*667  LF/HF & PPTh trap R: *r = -.*056; *p = .*707  LF & PPTh trap L: *r = -.*056; *p = .*706  HF & PPTh trap L: *r =* .015; *p = .*911  LF/HF & PPTh trap L: *r = -.*051; *p = .*732  LF & PPTh elbow R: *r = -.*022; *p = .*883  HF & PPTh elbow R: *r =* .016; *p = .*907  LF/HF & PPTh elbow R: *r = -.*021; *p = .*888  LF & PPTh elbow L: *r =* .066; *p = .*655  HF & PPTh elbow L: *r = -.*057; *p = .*701  LF/HF & PPTh elbow L: *r =* .062; *p = .*671  LF & PPTh lumbar R: *r =* .084; *p = .*57  HF & PPTh lumbar R: *r = -.*088; *p = .*554  LF/HF & PPTh lumbar R: *r =* .094; *p = .*528  LF & PPTh lumbar L: r = -0.027; *p = .*854  HF & PPTh lumbar L: *r =* .021; *p = .*887  LF/HF & PPTh lumbar L: *r = -.*046; *p = .*752  LF & PPTh patellar R: *r =* .026; *p = .*857  HF & PPTh patellar R: *r =* .041; *p = .*779  LF/HF & PPTh patellar R: *r =* .009; *p = .*95  LF & PPTh patellar L: *r =* .012; *p = .*933  HF & PPTh patellar L: *r =* .073; *p = .*617  LF/HF & PPTh patellar L: *r =* .004; *p = .*977  Δ Stress (baseline to after) – Pain baseline  **HC**  LF & PPTh trap R: *r = -.*031; *p = .*839  HF & PPTh trap R: *r =* .049; *p = .*739  LF/HF & PPTh trap R: *r =* .067; *p = .*646  LF & PPTh trap L: *r =* .011; *p = .*931  HF & PPTh trap L: *r = -.*064; *p = .*661  LF/HF & PPTh trap L: *r =* .059; *p = .*684  LF & PPTh elbow R: *r =* .021; *p = .*882  HF & PPTh elbow R: *r = -.*045; *p = .*762  LF/HF & PPTh elbow R: *r =* .011; *p = .*933  LF & PPTh elbow L: *r =* .031; *p = .*836  HF & PPTh elbow L: *r =* .063; *p = .*668  LF/HF & PPTh elbow L: *r = -.*081; *p = .*589  LF & PPTh lumbar R: *r =* .045; *p = .*764  HF & PPTh lumbar R: *r =* .047; *p = .*753  LF/HF & PPTh lumbar R: *r =* .091; *p = .*549  LF & PPTh lumbar L: *r =* .064; *p = .*662  HF & PPTh lumbar L: *r =* .034; *p = .*819  LF/HF & PPTh lumbar L: *r =* .051; *p = .*737  LF & PPTh patellar R: *r = -.*062; *p = .*672  HF & PPTh patellar R: *r =* .011; *p = .*932  LF/HF & PPTh patellar R: *r =* .038; *p = .*80  LF & PPTh patellar L: *r =* .071; *p = .*631  HF & PPTh patellar L: *r = -.*045; *p = .*762  LF/HF & PPTh patellar L: *r =* .048; *p = .*744  Stress baseline - Δ Pain (baseline to after):  **HC**  LF & PPTh trap R: *r =* .088; *p = .*55  HF & PPTh trap R: *r =* .072; *p = .*631  LF/HF & PPTh trap R: *r =* .045; *p = .*758  LF & PPTh trap L: *r =* .03; *p = .*843  HF & PPTh trap L: *r = -.*003; *p = .*982  LF/HF & PPTh trap L: *r =* .011; *p = .*936  LF & PPTh elbow R: *r =* .105; *p = .*474  HF & PPTh elbow R: *r = -.*058; *p = .*688  LF/HF & PPTh elbow R: *r =* .103; *p = .*481  LF & PPTh elbow L: *r =* .054; *p = .*707  HF & PPTh elbow L: *r = -.*084; *p = .*568  LF/HF & PPTh elbow L: *r =* .078; *p = .*593  LF & PPTh lumbar R: *r =* .077; *p = .*599  HF & PPTh lumbar R: *r =* .047; *p = .*749  LF/HF & PPTh lumbar R: *r =* .036; *p = .*803  LF & PPTh lumbar L: *r =* .01; *p = .*943  HF & PPTh lumbar L: *r =* .046; *p = .*752  LF/HF & PPTh lumbar L: *r = -.*014; *p = .*918  LF & PPTh patellar R: *r =* .065; *p = .*661  HF & PPTh patellar R: *r = -.*018; *p = .*896  LF/HF & PPTh patellar R: *r =* .043; *p = .*769  LF & PPTh patellar L: *r =* .088; *p = .*551  HF & PPTh patellar L: *r = -.*047; *p = .*748  LF/HF & PPTh patellar L: *r =* .04; *p = .*783  Δ Stress (baseline to during) - Δ Pain (baseline to after):  **HC**  LF & PPTh trap R: *r = -.*027; *p = .*86  HF & PPTh trap R: *r =* .051; p = 0.732  LF/HF & PPTh trap R: *r = -.*061; *p = .*68  LF & PPTh trap L: *r =* .049; *p = .*743  HF & PPTh trap L: *r =* .041; *p = .*78  LF/HF & PPTh trap L: *r =* .012; *p = .*932  LF & PPTh elbow R: *r = -.*047; *p = .*751  HF & PPTh elbow R: *r =* .071; *p = .*628  LF/HF & PPTh elbow R: *r = -.*061; *p = .*681  LF & PPTh elbow L: *r =* .064; *p = .*667  HF & PPTh elbow L: *r = -.*031; *p = .*837  LF/HF & PPTh elbow L: *r =* .043; *p = .*772  LF & PPTh lumbar R: *r =* .011; *p = .*933  HF & PPTh lumbar R: *r =* .034; *p = .*818  LF/HF & PPTh lumbar R: *r = -.*014; *p = .*916  LF & PPTh lumbar L: *r =* .057; *p = .*70  HF & PPTh lumbar L: *r =* .043; *p = .*771  LF/HF & PPTh lumbar L: *r =* .061; *p = .*676  LF & PPTh patellar R: *r =* .029; *p = .*849  HF & PPTh patellar R: *r =* .051; *p = .*734  LF/HF & PPTh patellar R: r = 0.064; *p = .*663  LF & PPTh patellar L: *r =* .048; *p = .*748  HF & PPTh patellar L: *r = -.*051; *p = .*734  LF/HF & PPTh patellar L: *r =* .045; *p = .*76  Δ Stress (baseline to after) - Δ Pain (baseline to after):  **HC**  LF & PPTh trap R: *r =* .031; *p = .*838  HF & PPTh trap R: *r = -.*029; p = 0.849  LF/HF & PPTh trap R: *r =* .035; *p = .*819  LF & PPTh trap L: *r = -.*014; *p = .*917  HF & PPTh trap L: *r =* .043; *p = .*773  LF/HF & PPTh trap L: *r =* .051; *p = .*735  LF & PPTh elbow R: *r =* .011; *p = .*935  HF & PPTh elbow R: *r =* .031; *p = .*836  LF/HF & PPTh elbow R: *r =* .048; *p = .*746  LF & PPTh elbow L: *r = -.*047; *p = .*752  HF & PPTh elbow L: *r =* .061; *p = .*676  LF/HF & PPTh elbow L: *r = -.*063; *p = .*669  LF & PPTh lumbar R: *r =* .021; *p = .*886  HF & PPTh lumbar R: *r =* .012; *p = .*929  LF/HF & PPTh lumbar R: *r =* .037; *p = .*807  LF & PPTh lumbar L: *r =* .058; *p = .*695  HF & PPTh lumbar L: *r = -.*052; *p = .*733  LF/HF & PPTh lumbar L: *r =* .047; *p = .*751  LF & PPTh patellar R: *r = -.*012; *p = .*93  HF & PPTh patellar R: *r =* .041; *p = .*781  LF/HF & PPTh patellar R: r = -0.029; *p = .*849  LF & PPTh patellar L: *r = -.*017; *p = .*906  HF & PPTh patellar L: *r =* .051; *p = .*734  LF/HF & PPTh patellar L: *r =* .036; *p = .*813  Stress during - Pain baseline:  **HC**  LF & PPTh trap R: *r = -.*032; *p = .*834 HF & PPTh trap R: *r =* .063; *p = .*666 LF/HF & PPTh trap R: *r =* .038; *p = .*804  LF & PPTh trap L: *r =* .041; *p = .*784 HF & PPTh trap L: *r = -.*039; *p = .*797 LF/HF & PPTh trap L: *r =* .031; *p = .*836  LF & PPTh elbow R: *r = -.*011; *p = .*932 HF & PPTh elbow R: *r =* .043; *p = .*776 LF/HF & PPTh elbow R: *r = -.*034; *p = .*822  LF & PPTh elbow L: *r =* .064; *p = .*665 HF & PPTh elbow L: *r = -.*061; *p = .*677 LF/HF & PPTh elbow L: *r =* .037; *p = .*804  LF & PPTh lumbar R: *r = -.*047; *p = .*752 HF & PPTh lumbar R: *r =* .048; *p = .*748 LF/HF & PPTh lumbar R: *r =* .065; *p = .*66  LF & PPTh lumbar L: *r = -.*029; *p = .*85 HF & PPTh lumbar L: *r =* .057; *p = .*705 LF/HF & PPTh lumbar L: *r = -.*051; *p = .*736  LF & PPTh patellar R: *r =* .071; *p = .*625 HF & PPTh patellar R: *r = -.*042; *p = .*779 LF/HF & PPTh patellar R: *r =* .021; *p = .*886  LF & PPTh patellar L: *r =* .012; *p = .*93 HF & PPTh patellar L: *r = -.*029; *p = .*848 LF/HF & PPTh patellar L: *r =* .051; *p = .*734  Stress after - Pain baseline:  **HC**  LF & PPTh trap R: *r =* .024; *p = .*872 HF & PPTh trap R: *r = -.*034; *p = .*82 LF/HF & PPTh trap R: *r =* .064; *p = .*664  LF & PPTh trap L: *r =* .061; *p = .*679 HF & PPTh trap L: *r = -.*054; *p = .*722 LF/HF & PPTh trap L: *r =* .047; *p = .*752  LF & PPTh elbow R: *r = -.*037; *p = .*805 HF & PPTh elbow R: *r =* .044; *p = .*771 LF/HF & PPTh elbow R: *r = -.*067; *p = .*649  LF & PPTh elbow L: *r =* .021; *p = .*889 HF & PPTh elbow L: *r = -.*045; *p = .*758 LF/HF & PPTh elbow L: *r =* .031; *p = .*837  LF & PPTh lumbar R: *r =* .034; *p = .*819 HF & PPTh lumbar R: *r =* .051; *p = .*735 LF/HF & PPTh lumbar R: *r =* .029; *p = .*85  LF & PPTh lumbar L: *r = -.*019; *p = .*897 HF & PPTh lumbar L: *r =* .065; *p = .*662 LF/HF & PPTh lumbar L: *r = -.*043; *p = .*772  LF & PPTh patellar R: *r =* .021; *p = .*889 HF & PPTh patellar R: *r =* .061; *p = .*678 LF/HF & PPTh patellar R: *r =* .054; *p = .*722  LF & PPTh patellar L: *r = -.*051; *p = .*733 HF & PPTh patellar L: *r =* .048; *p = .*748 LF/HF & PPTh patellar L: *r =* .065; *p = .*663  Stress baseline - Pain after:  **HC**  LF & PPTh trap R: *r =* .029; *p = .*85 HF & PPTh trap R: *r = -.*038; *p = .*804 LF/HF & PPTh trap R: *r =* .065; *p = .*661  LF & PPTh trap L: *r = -.*057; *p = .*704 HF & PPTh trap L: *r =* .048; *p = .*748 LF/HF & PPTh trap L: *r =* .041; *p = .*781  LF & PPTh elbow R: *r =* .047; *p = .*751 HF & PPTh elbow R: *r = -.*037; *p = .*806 LF/HF & PPTh elbow R: *r =* .064; *p = .*665  LF & PPTh elbow L: *r =* .014; *p = .*917 HF & PPTh elbow L: *r = -.*029; *p = .*850 LF/HF & PPTh elbow L: *r =* .012; *p = .*932  LF & PPTh lumbar R: *r =* .031; *p = .*837 HF & PPTh lumbar R: *r = -.*011; *p = .*932 LF/HF & PPTh lumbar R: *r =* .021; *p = .*888  LF & PPTh lumbar L: *r =* .061; *p = .*677 HF & PPTh lumbar L: *r =* .034; *p = .*822 LF/HF & PPTh lumbar L: *r =* .051; *p = .*734  LF & PPTh patellar R: *r =* .048; *p = .*746 HF & PPTh patellar R: *r =* .027; *p = .*855 LF/HF & PPTh patellar R: *r = -.*012; *p = .*93  LF & PPTh patellar L: *r = -.*021; *p = .*888 HF & PPTh patellar L: *r =* .065; *p = .*663 LF/HF & PPTh patellar L: *r =* .031; *p = .*837  Stress during - Pain after:  **HC**  LF & PPTh trap R: *r =* .031; *p = .*837 HF & PPTh trap R: *r =* .043; *p = .*773 LF/HF & PPTh trap R: *r =* .064; *p = .*665  LF & PPTh trap L: *r =* .057; *p = .*704 HF & PPTh trap L: *r =* .048; *p = .*748 LF/HF & PPTh trap L: *r =* .041; *p = .*781  LF & PPTh elbow R: *r =* .047; *p = .*751 HF & PPTh elbow R: *r = -.*037; *p = .*806 LF/HF & PPTh elbow R: *r =* .064; *p = .*665  LF & PPTh elbow L: *r =* .014; *p = .*917 HF & PPTh elbow L: *r = -.*029; *p = .*85 LF/HF & PPTh elbow L: *r =* .012; *p = .*932  LF & PPTh lumbar R: *r =* .031; *p = .*837 HF & PPTh lumbar R: *r = -.*011; *p = .*932 LF/HF & PPTh lumbar R: *r =* .021; *p = .*888  LF & PPTh lumbar L: *r =* .061; *p = .*677 HF & PPTh lumbar L: *r =* .034; *p = .*822 LF/HF & PPTh lumbar L: *r =* .051; *p = .*734  LF & PPTh patellar R: *r =* .048; *p = .*746 HF & PPTh patellar R: *r =* .027; *p = .*855 LF/HF & PPTh patellar R: *r = -.*012; *p = .*930  LF & PPTh patellar L: *r = -.*021; *p = .*888 HF & PPTh patellar L: *r =* .065; *p = .*663 LF/HF & PPTh patellar L: *r =* .031; *p = .*837  Stress after - Pain after:  **HC**  LF & PPTh trap R: *r =* .031; *p = .*837 HF & PPTh trap R: *r =* .043; *p = .*773 LF/HF & PPTh trap R: *r =* .064; *p = .*665  LF & PPTh trap L: *r =* .057; *p = .*704 HF & PPTh trap L: *r =* .048; *p = .*748 LF/HF & PPTh trap L: *r =* .041; *p = .*781  LF & PPTh elbow R: *r =* .047; *p = .*751 HF & PPTh elbow R: *r = -.*037; *p = .*806 LF/HF & PPTh elbow R: *r =* .064; *p = .*665  LF & PPTh elbow L: *r =* .014; *p = .*917 HF & PPTh elbow L: *r = -.*029; *p = .*85 LF/HF & PPTh elbow L: *r =* .012; *p = .*932  LF & PPTh lumbar R: *r =* .031; *p = .*837 HF & PPTh lumbar R: *r = -.*011; *p = .*932 LF/HF & PPTh lumbar R: *r =* .021; *p = .*888  LF & PPTh lumbar L: *r =* .061; *p = .*677 HF & PPTh lumbar L: *r =* .034; *p = .*822 LF/HF & PPTh lumbar L: *r =* .051; *p = .*734  LF & PPTh patellar R: *r =* .048; *p = .*746 HF & PPTh patellar R: *r =* .027; *p = .*855 LF/HF & PPTh patellar R: *r = -.*012; *p = .*93  LF & PPTh patellar L: *r = -.*021; *p = .*888 HF & PPTh patellar L: *r =* .065; *p = .*663 LF/HF & PPTh patellar L: *r =* .031; *p = .*837  Stress during - Δ Pain:  **HC**  LF & PPTh trap R: *r =* .062; *p = .*673 HF & PPTh trap R: *r =* .058; *p = .*694 LF/HF & PPTh trap R: *r =* .041; *p = .*781  LF & PPTh trap L: *r =* .031; *p = .*836 HF & PPTh trap L: *r = -.*029; *p = .*849 LF/HF & PPTh trap L: *r =* .034; *p = .*822  LF & PPTh elbow R: *r =* .048; *p = .*748 HF & PPTh elbow R: *r = -.*031; *p = .*837 LF/HF & PPTh elbow R: *r =* .043; *p = .*774  LF & PPTh elbow L: *r =* .064; *p = .*667 HF & PPTh elbow L: *r = -.*029; *p = .*849 LF/HF & PPTh elbow L: *r =* .012; *p = .*932  LF & PPTh lumbar R: *r =* .014; *p = .*916 HF & PPTh lumbar R: *r =* .021; *p = .*887 LF/HF & PPTh lumbar R: *r =* .011; *p = .*935  LF & PPTh lumbar L: *r =* .065; *p = .*661 HF & PPTh lumbar L: *r = -.*052; *p = .*733 LF/HF & PPTh lumbar L: *r =* .021; *p = .*888  LF & PPTh patellar R: *r =* .054; *p = .*721 HF & PPTh patellar R: *r =* .011; *p = .*935 LF/HF & PPTh patellar R: *r =* .065; *p = .*66  LF & PPTh patellar L: *r = -.*021; *p = .*887 HF & PPTh patellar L: *r =* .031; *p = .*837 LF/HF & PPTh patellar L: *r =* .048; *p = .*745  Stress after - Δ Pain (baseline to after):  **HC**  LF & PPTh trap R: *r =* .031; *p = .*837 HF & PPTh trap R: *r =* .043; *p = .*773 LF/HF & PPTh trap R: *r =* .064; *p = .*665  LF & PPTh trap L: *r =* .057; *p = .*704 HF & PPTh trap L: *r =* .048; *p = .*748 LF/HF & PPTh trap L: *r =* .041; *p = .*781  LF & PPTh elbow R: *r =* .047; *p = .*751 HF & PPTh elbow R: *r = -.*037; *p = .*806 LF/HF & PPTh elbow R: *r =* .064; *p = .*665  LF & PPTh elbow L: *r =* .014; *p = .*917 HF & PPTh elbow L: *r = -.*029; *p = .*850 LF/HF & PPTh elbow L: *r =* .012; *p = .*932  LF & PPTh lumbar R: *r =* .031; *p = .*837 HF & PPTh lumbar R: *r = -.*011; *p = .*932 LF/HF & PPTh lumbar R: *r =* .021; *p = .*888  LF & PPTh lumbar L: *r =* .061; *p = .*677 HF & PPTh lumbar L: *r =* .034; *p = .*822 LF/HF & PPTh lumbar L: *r =* .051; *p = .*734  LF & PPTh patellar R: *r =* .048; *p = .*746 HF & PPTh patellar R: *r =* .027; *p = .*856 LF/HF & PPTh patellar R: *r = -.*012; *p = .*93  LF & PPTh patellar L: *r = -.*021; *p = .*888 HF & PPTh patellar L: *r =* .065; *p = .*663 LF/HF & PPTh patellar L: *r =* .031; *p = .*837  Δ Stress (baseline to during) - Pain after:  **HC**  LF & PPTh trap R: *r =* .011; *p = .*933 HF & PPTh trap R: *r =* .031; *p = .*837 LF/HF & PPTh trap R: *r =* .048; *p = .*745  LF & PPTh trap L: *r =* .057; *p = .*704 HF & PPTh trap L: *r = -.*048; *p = .*748 LF/HF & PPTh trap L: *r =* .041; *p = .*781  LF & PPTh elbow R: *r =* .047; *p = .*751 HF & PPTh elbow R: *r = -.*037; *p = .*806 LF/HF & PPTh elbow R: *r =* .064; *p = .*665  LF & PPTh elbow L: *r =* .014; *p = .*917 HF & PPTh elbow L: *r = -.*029; *p = .*85 LF/HF & PPTh elbow L: *r =* .012; *p = .*932  LF & PPTh lumbar R: *r =* .031; *p = .*837 HF & PPTh lumbar R: *r = -.*011; *p = .*932 LF/HF & PPTh lumbar R: *r =* .021; *p = .*888  LF & PPTh lumbar L: *r =* .061; *p = .*677 HF & PPTh lumbar L: *r =* .034; *p = .*822 LF/HF & PPTh lumbar L: *r =* .051; *p = .*734  LF & PPTh patellar R: *r =* .048; *p = .*746 HF & PPTh patellar R: *r =* .027; *p = .*855 LF/HF & PPTh patellar R: *r = -.*012; *p = .*93  LF & PPTh patellar L: *r = -.*021; *p = .*888 HF & PPTh patellar L: *r =* .065; *p = .*663 LF/HF & PPTh patellar L: *r =* .031; *p = .*837  Δ Stress (baseline to after) - Pain after:  **HC**  LF & PPTh trap R: *r =* .014; *p = .*92 HF & PPTh trap R: *r = -.*021; *p = .*887 LF/HF & PPTh trap R: *r =* .031; *p = .*836  LF & PPTh trap L: *r =* .031; *p = .*837 HF & PPTh trap L: *r = -.*012; *p = .*929 LF/HF & PPTh trap L: *r =* .048; *p = .*746  LF & PPTh elbow R: *r =* .048; *p = .*746 HF & PPTh elbow R: *r =* .027; *p = .*856 LF/HF & PPTh elbow R: *r = -.*012; *p = .*929  LF & PPTh elbow L: *r =* .048; *p = .*746 HF & PPTh elbow L: *r =* .027; *p = .*856 LF/HF & PPTh elbow L: *r = -.*021; *p = .*887  LF & PPTh lumbar R: *r = -.*048; *p = .*746 HF & PPTh lumbar R: *r =* .027; *p = .*856 LF/HF & PPTh lumbar R: *r =* .031; *p = .*837  LF & PPTh lumbar L: *r =* .048; *p = .*746 HF & PPTh lumbar L: *r = -.*021; *p = .*887 LF/HF & PPTh lumbar L: *r =* .031; *p = .*837  LF & PPTh patellar R: *r = -.*048; *p = .*746 HF & PPTh patellar R: *r =* .027; *p = .*856 LF/HF & PPTh patellar R: *r =* .031; *p = .*837  LF & PPTh patellar L: *r = -.*021; *p = .*887 HF & PPTh patellar L: *r =* .031; *p = .*837 LF/HF & PPTh patellar L: *r =* .048; *p = .*746 |
| Bossenger et al. (2023) | Δ Stress – Δ Pain:  **CPP (ws)**  HF HRV during stressor & PPTh after stressor: *p* < .05  Other ANS measures: *r = -.*20; *p = .*52 | Δ Stress – Δ Pain:  **HC**  HF HRV during stressor & PPTh after stressor: *p* < .05  Other ANS measures: *r =* .24; *p = .*43 |
| Chalaye et al. (2012) | N.A. | N.A. |
| Chalaye et al. (2014) | Δ Stress – Pain baseline (regression):  **CPP (ws)**  Increase in SBP during CPT & CPM effectiveness: **b = 0.44; t = 2.2; *p = .*04**  **Entire sample**  Increase in SBP during CPT & CPM effectiveness: **b = 0.34; t = 2.4; *p = .*019**  No sign association between other ANS measures and CPM (*p* > .05)  Stress baseline – Pain baseline (regression):  **CPP (ws)**  HR & pain intensity during CPT: **b = 0.43; t = 2.2; *p = .*043**  **Entire sample**  HR & pain intensity during CPT: **b = 0.31; t = 2.2; *p = .*036**  Stress during – Pain Baseline (regression):  **CPP (ws)**  HR during CPT & pain intensity during CPT: **b = 0.51; t = 2.6; *p = .*015**  **Entire sample**  HR during CPT & pain intensity during CPT: **b = 0.34; t = 2.4; *p = .*02** | N.A. |
| Cohen et al. (2000) | Stress baseline – Pain baseline:  **CPP**  LF HRV & tender/control: *r = -.*01/ .04; *p* > .05  HF & tender/control: *r =* .01/ -.04; *p* > .05  LF/HF & tender/control: *r = -.*12/ -.15; *p* > .05  HRV & tender/control: *r = -.*38/ -.30; *p* > .05  HR & tender/control: *r =* .08/ .02; *p* > .05 | N.A. |
| Crettaz et al. (2013) | N.A. | N.A. |
| Davydov et al. (2024) | Stress baseline – Pain baseline  **CPP (ws)**  Hair cortisol & PPTh: *r =* .092; *p = .*54  Hair cortisol & PPTo: *r =* .054; *p = .*72  Hair cortisol & SREP: *r =* .08; *p = .*59 | Stress baseline – Pain baseline  **HC**  Hair cortisol & PPTh: *r =* .031; *p = .*86  Hair cortisol & PPTo: *r = -.*18; *p = .*30  Hair cortisol & SREP: *r =* .093; *p = .*59 |
| De Abreu Freitas et al. (2012) | Stress baseline – Pain baseline  **CPP**  Cortisol Day 1 & PPTh: ***r = -.*606; *p = .*01** Cortisol Day 1 & PPTo: *r = -.*439; *p = .*078 Cortisol Day 2 & PPTh: *r = -.*372; *p = .*141 Cortisol Day 2 & PPTo: *r = -.*028; *p = .*915 Cortisol Day 3 & PPTh: *r = -.*286; *p = .*266 Cortisol Day 3 & PPTo: *r = .*02; *p = .*938 Cortisol Mean & PPTh: *r = -.*419; *p = .*094 Cortisol Mean & PPTo: *r = -.*049; *p = .*851  DHEA Day 1 & PPTh: *r = .*412; *p = .*10 DHEA Day 1 & PPTo: *r = .*348; *p = .*171 DHEA Day 2 & PPTh: *r = .*256; *p = .*321 DHEA Day 2 & PPTo: *r = .*144; *p = .*582 DHEA Day 3 & PPTh: *r = .*245; *p = .*343 DHEA Day 3 & PPTo: *r = .*243; *p = .*347 DHEA Mean & PPTh: *r = .*295; *p = .*251 DHEA Mean & PPTo: *r = .*22; *p = .*396 | Stress baseline – Pain baseline  **HC**  Cortisol Day 1 & PPTh: *r = .*162; *p = .*508 Cortisol Day 1 & PPTo: *r = .*09; *p = .*713 Cortisol Day 2 & PPTh: *r = -.*154; *p = .*529 Cortisol Day 2 & PPTo: *r = -.*136; *p = .*579 Cortisol Day 3 & PPTh: *r = -.*044; *p = .*86 Cortisol Day 3 & PPTo: *r = -.*017; *p = .*946 Cortisol Mean & PPTh: *r = .*076; *p = .*758 Cortisol Mean & PPTo: *r = .*055; *p = .*822  DHEA Day 1 & PPTh: ***r = .*697; *p = .*001** DHEA Day 1 & PPTo: ***r = .*525; *p = .*021** DHEA Day 2 & PPTh: ***r = .*71; *p = .*001** DHEA Day 2 & PPTo: ***r = .*597; *p = .*007** DHEA Day 3 & PPTh: ***r = .*595; *p = .*007** DHEA Day 3 & PPTo: *r = .*443; *p = .*057 DHEA Mean & PPTh: ***r = .*693; *p = .*001** DHEA Mean & PPTo: ***r = .*54; *p = .*017** |
| De Bruijn et al. (2011) | Stress after – Pain baseline  **CPP (ws)**  HR & CPTh: ***r =* .70; *p = .*004**  HR & PPTh: ***r = -.*70; *p = .*004**  HR & WUR: ***r =* .66; *p = .*01** | N.A. |
| De La Coba et al. (2018) | Stress after – Pain baseline  **CPP (ws)**  SBP & SREP: ***r = -.*55; *p* ≤ .01**  DBP & SREP: ***r = -.*46; *p* ≤ .01**  SBP & PPTo: *r = -.*14; *p > .*05  DBP & PPTo: *r = -.*18; *p* > .05  SBP & PPTh: *r = -.*27; *p* > .05  DBP & PPTh: *r = -.*19; *p* > .05 | Stress after – Pain baseline  **HC**  SBP & SREP: *r = -.*02; *p* > .05  DBP & SREP: *r = -.*06; *p* > .05  SBP & PPTo: ***r =* .38; *p* ≤ .05**  DBP & PPTo: *r =* .12; *p* > .05  SBP & PPTh: ***r =* .52; *p* ≤ .05**  DBP & PPTh: ***r =* .28; *p* ≤ .05** |
| Del Paso et al. (2011) | Stress baseline – Pain baseline  **CPP (ws)**  LF HRV & CPTh: *r = -.*055; *p = .*756 LF HRV & CPTo: *r =* .203; *p = .*249 HF HRV & CPTh: *r = -.*131; *p = .*483 HF HRV & CPTo: *r =* .147; *p = .*431 LF/HF HRV & CPTh: *r = -.*19; *p = .*282 LF/HF HRV & CPTo: *r = -.*014; *p = .*936 SBP & CPTh: *r =* .151; *p = .*387 SBP & CPTo: *r = -.*163; *p = .*348 DBP & CPTh: *r =* .079; *p = .*652 DBP & CPTo: *r = -.*229; *p = .*186 RRI & CPTh: *r =* .132; *p = .*451 RRI & CPTo: *r =* .071; *p = .*684 BRS & CPTh: *r = -.*072; *p = .*681 BRS & CPTo: *r =* .14; *p = .*423  Stress during – Pain Baseline  **CPP (ws)**  LF HRV during hot water & CPTh: *r =* .156; *p = .*41 LF HRV during hot water & CPTo**: *r =* .479; *p = .*007** LF HRV during cold water & CPTh: *r =* .101; *p = .*596 LF HRV during cold water & CPTo: *r =* .257; *p = .*17 HF HRV during hot water & CPTh: *r = -.*033; *p = .*859 HF HRV during hot water & CPTo: ***r =* .639; *p* ≤ .001** HF HRV during cold water & CPTh: *r = -.*005; *p = .*979 HF HRV during cold water & CPTo: ***r =* .387; *p = .*026** LF/HF HRV during hot water & CPTh: *r = -.*208; *p = .*246 LF/HF HRV during hot water & CPTo: *r = -.*187; *p = .*296 LF/HF HRV during cold water & CPTh: *r =* .083; *p = .*64 LF/HF HRV during cold water & CPTo: *r =* .197; *p = .*264 SBP during hot water & CPTh: *r =* .079; *p = .*651 SBP during hot water & CPTo: *r = -.*190; *p = .*274 SBP during cold water & CPTh: *r =* .203; *p = .*243 SBP during cold water & CPTo: *r =* .023; *p = .*897 DBP during hot water & CPTh: *r = -.*048; *p = .*785 DBP during hot water & CPTo: ***r = -.*334; *p = .*05** DBP during cold water & CPTh: *r =* .065; *p = .*711 DBP during cold water & CPTo: *r = -.*234; *p = .*176 RRI during hot water & CPTh: *r =* .106; *p = .*545 RRI during hot water & CPTo: *r =* .065; *p = .*712 RRI during cold water & CPTh: *r =* .096; *p = .*584 RRI during cold water & CPTo: *r = -.*011; *p = .*951 BRS during hot water & CPTh: *r = -.*078; *p = .*656 BRS during hot water & CPTo: *r =* .169; *p = .*33 BRS during cold water & CPTh: *r = -.*137; *p = .*431 BRS during cold water & CPTo: *r =* .026; *p = .*883  Stress after – Pain Baseline  **CPP (ws)**  LF HRV at recovery & CPTh: *r =* .035; *p = .*85 LF HRV at recovery & CPTo: *r =* .094; *p = .*609 HF HRV at recovery & CPTh: *r =* .018; *p = .*92 HF HRV at recovery & CPTo: ***r =* .409; *p = .*018** LF/HF HRV at recovery & CPTh: *r = -.*019; *p = .*916 LF/HF HRV at recovery & CPTo: *r = -.*069; *p = .*695 SBP at recovery & CPTh: *r =* .119; *p = .*496 SBP at recovery & CPTo: *r = -.*215; *p = .*216 DBP at recovery & CPTh: *r =* .06; *p = .*733 DBP at recovery & CPTo: *r = -.*284; *p = .*099 RRI at recovery & CPTh: *r =* .136; *p = .*436 RRI at recovery & CPTo: *r =* .04; *p = .*819 BRS at recovery & CPTh: *r = -.*109; *p = .*534 BRS at recovery & CPTo: *r =* .107; *p = .*539 | Stress baseline – Pain baseline  **HC**  LF HRV & CPTh: *r = -.*01; *p = .*959 LF HRV & CPTo: *r = -.*093; *p = .*637 HF HRV & CPTh: *r = -.*106; *p = .*597 HF HRV & CPTo: *r = -.*083; *p = .*679 LF/HF HRV & CPTh: *r = -.*017; *p = .*932 LF/HF HRV & CPTo: *r = -.*114; *p = .*556 SBP & CPTh: *r =* .262; *p = .*17 SBP & CPTo: *r =* .02; *p = .*917 DBP & CPTh: *r =* .246; *p = .*199 DBP & CPTo: *r =* .065; *p = .*739 RRI & CPTh: *r = -.*216; *p = .*261  RRI & CPTo: *r = -.*318; *p = .*093  BRS & CPTh: *r = -.*039; *p = .*843 BRS & CPTo: *r = -.*119; *p = .*54  Stress during – Pain Baseline  **HC**  LF HRV during hot water & CPTh: *r = -.*015; *p = .*941 LF HRV during hot water & CPTo: *r = -.*095; *p = .*63 LF HRV during cold water & CPTh: *r = -.*177; *p = .*377 LF HRV during cold water & CPTo: *r = -.*027; *p = .*895 HF HRV during hot water & CPTh: *r =* .087; *p = .*662 HF HRV during hot water & CPTo: *r =* .024; *p = .*904 HF HRV during cold water & CPTh: *r =* .121; *p = .*546 HF HRV during cold water & CPTo: *r =* .31; *p = .*116 LF/HF HRV during hot water & CPTh: *r = -.*105; *p = .*587 LF/HF HRV during hot water & CPTo: *r = -.*23; *p = .*23 LF/HF HRV during cold water & CPTh: *r = -.*109; *p = .*588  LF/HF HRV during cold water & CPTo: *r = -.*263; *p = .*184  SBP during hot water & CPTh: *r =* .322; *p = .*089 SBP during hot water & CPTo: *r =* .011; *p = .*955 SBP during cold water & CPTh: *r =* .276; *p = .*147 SBP during cold water & CPTo: *r =* .044; *p = .*82 DBP during hot water & CPTh: *r =* .224; *p = .*243 DBP during hot water & CPTo: *r = -.*007; *p = .*97 DBP during cold water & CPTh: *r =* .226; *p = .*238 DBP during cold water & CPTo: *r =* .038; *p = .*843 RRI during hot water & CPTh: *r = -.*212; *p = .*27 RRI during hot water & CPTo: *r = -.*264; *p = .*166 RRI during cold water & CPTh: *r = -.*148; *p = .*444 RRI during cold water & CPTo: *r = -.*253; *p = .*186 BRS during hot water & CPTh: *r = -.*003; *p = .*988 BRS during hot water & CPTo: *r = -.*09; *p = .*644 BRS during cold water & CPTh: *r = -.*191; *p = .*32 BRS during cold water & CPTo: *r = -.*121; *p = .*531  Stress after – Pain Baseline  **HC**  LF HRV at recovery & CPTh: *r = -.*052; *p = .*794 LF HRV at recovery & CPTo: *r = -.*048; *p = .*807 HF HRV at recovery & CPTh: *r =* .003; *p = .*988 HF HRV at recovery & CPTo: *r =* .089; *p = .*651 LF/HF HRV at recovery & CPTh: *r = -.*035; *p = .*855 LF/HF HRV at recovery & CPTo: *r = -.*233; *p = .*223 SBP at recovery & CPTh: *r =* .248; *p = .*194 SBP at recovery & CPTo: *r = -.*106; *p = .*585 DBP at recovery & CPTh: *r =* .243; *p = .*204 DBP at recovery & CPTo: *r = -.*083; *p = .*67 RRI at recovery & CPTh: *r = -.*178; *p = .*356 RRI at recovery & CPTo: *r = -.*279; *p = .*142 BRS at recovery & CPTh: *r = -.*048; *p = .*803 BRS at recovery & CPTo: *r = -.*119; *p = .*539 |
| Del Paso et al. (2022) | Stress baseline – Pain Baseline  **CPP**  LF HRV & CPTh: *r = -.*131; *p = .*353 LF HRV & CPTo: *r = -.*005; *p = .*974 LF HRV (baseline 2) & CPTh: *r = -.*122; *p = .*397 LF HRV (baseline 2) & CPTo: *r =* .076; *p = .*599  HF HRV & CPTh: *r = -.*012; *p = .*933 HF HRV & CPTo: *r =* .035; *p = .*811 HF HRV (baseline 2) & CPTh: *r =* .091; *p = .*534 HF HRV (baseline 2) & CPTo: *r =* .219; *p = .*13  SBP & CPTh: *r = -.*104; *p = .*458 SBP & CPTo: *r = -.*175; *p = .*21 SBP (baseline 2) & CPTh: *r =* .024; *p = .*862 SBP (baseline 2) & CPTo: *r = -.*053; *p = .*708  DBP & CPTh: *r = -.*08; *p = .*57 DBP & CPTo: *r = -.*151; *p = .*282 DBP (baseline 2) & CPTh: *r =* .082; *p = .*557 DBP (baseline 2) & CPTo: *r = -.*096; *p = .*494  RRI & CPTh: *r =* .102; *p = .*467 RRI & CPTo: *r = -.*01; *p = .*943 RRI (baseline 2) & CPTh: *r =* .111; *p = .*427 RRI (baseline 2) & CPTo: *r =* .039; *p = .*779  BRS & CPTh: *r = -.*008; *p = .*953 BRS & CPTo: *r =* .079; *p = .*572 BRS (baseline 2) & CPTh: *r =* .114; *p = .*416 BRS (baseline 2) & CPTo: *r =* .182; *p = .*192  Stress during – Pain Baseline  **CPP**  LF HRV during arithmetic & CPTh: *r =* .134; *p = .*342 LF HRV during arithmetic & CPTo: ***r =* .320; *p = .*021** LF HRV during hot water & CPTh: *r =* .178; *p = .*225 LF HRV during hot water & CPTo: *r =* .145; *p = .*325 LF HRV during cold water & CPTh: *r =* .196; *p = .*181 LF HRV during cold water & CPTo: *r =* .12; *p = .*415  HF HRV during arithmetic & CPTh: *r =* .111; *p = .*439 HF HRV during arithmetic & CPTo: *r =* .186; *p = .*192 HF HRV during hot water & CPTh: *r =* .171; *p = .*236 HF HRV during hot water & CPTo: *r =* .136; *p = .*345 HF HRV during cold water & CPTh: *r =* .112; *p = .*433 HF HRV during cold water & CPTo: *r =* .116; *p = .*417  SBP during arithmetic & CPTh: *r = -.*044; *p = .*756 SBP during arithmetic & CPTo: *r =* .004; *p = .*979 SBP during hot water & CPTh: *r =* .012; *p = .*933 SBP during hot water & CPTo: *r = -.*108; *p = .*441 SBP during cold water & CPTh: *r =* .102; *p = .*466 SBP during cold water & CPTo: *r =* .122; *p = .*383  DBP during arithmetic & CPTh: *r =* .034; *p = .*808 DBP during arithmetic & CPTo: *r = -.*029; *p = .*836 DBP during hot water & CPTh: *r =* .051; *p = .*716 DBP during hot water & CPTo: *r = -.*144; *p = .*304 DBP during cold water & CPTh: *r =* .164; *p = .*24 DBP during cold water & CPTo: *r =* .071; *p = .*614  RRI during arithmetic & CPTh: *r =* .15; *p = .*283 RRI during arithmetic & CPTo: *r =* .034; *p = .*811 RRI during hot water & CPTh: *r =* .031; *p = .*826 RRI during hot water & CPTo: *r = -.*042; *p = .*765 RRI during cold water & CPTh: *r =* .028; *p = .*841 RRI during cold water & CPTo: *r = -.*076; *p = .*589  BRS during arithmetic & CPTh: ***r =* .314; *p = .*022** BRS during arithmetic & CPTo: ***r =* .301; *p = .*028** BRS during hot water & CPTh: *r = -.*017; *p = .*905 BRS during hot water & CPTo: *r =* .118; *p = .*40 BRS during cold water & CPTh: *r =* .047; *p = .*736 BRS during cold water & CPTo: *r =* .042; *p = .*765  Stress after – Pain Baseline  **CPP**  LF HRV after arithmetic & CPTh: *r =* .016; *p = .*91 LF HRV after arithmetic & CPTo: *r =* .138; *p = .*328 LF HRV after water & CPTh: *r =* .247; *p = .*084 LF HRV after water & CPTo: *r =* .165; *p = .*253  HF HRV after arithmetic & CPTh: *r =* .054; *p = .*706 HF HRV after arithmetic & CPTo: *r =* .23; *p = .*105 HF HRV after water & CPTh: *r =* .186; *p = .*191 HF HRV after water & CPTo: *r =* .159; *p = .*265  SBP after arithmetic & CPTh: *r = -.*005; *p = .*971 SBP after arithmetic & CPTo: *r = -.*063; *p = .*653 SBP after water & CPTh: *r =* .058; *p = .*68 SBP after water & CPTo: *r = -.*10; *p = .*478  DBP after arithmetic & CPTh: *r =* .057; *p = .*687 DBP after arithmetic & CPTo: *r = -.*20; *p = .*151 DBP after water & CPTh: *r =* .086; *p = .*542 DBP after water & CPTo: *r = -.*075; *p = .*592  RRI after arithmetic & CPTh: *r =* .119; *p = .*398 RRI after arithmetic & CPTo: *r =* .029; *p = .*835 RRI after water & CPTh: *r =* .129; *p = .*358 RRI after water & CPTo: *r =* .006; *p = .*964  BRS after arithmetic & CPTh: *r =* .225; *p = .*105 BRS after arithmetic & CPTo: *r =* .227; *p = .*102 BRS after water & CPTh: *r =* .038; *p = .*785 BRS after water & CPTo: *r =* .151; *p = .*281 | Stress baseline – Pain Baseline  **HC**  LF HRV & CPTh: *r = -.*01; *p = .*957 LF HRV & CPTo: *r = -.*093; *p = .*63 LF HRV (baseline 2) & CPTh: *r = -.*071; *p = .*721 LF HRV (baseline 2) & CPTo: *r = -.*188; *p = .*337  HF HRV & CPTh: *r = -.*106; *p = .*593 HF HRV & CPTo: *r = -.*085; *p = .*669 HF HRV (baseline 2) & CPTh: *r =* .062; *p = .*748 HF HRV (baseline 2) & CPTo: *r = -.*011; *p = .*955  SBP & CPTh: *r =* .261; *p = .*164 SBP & CPTo: *r =* .02; *p = .*916 SBP (baseline 2) & CPTh: ***r =* .363; *p = .*049** SBP (baseline 2) & CPTo: *r =* .121; *p = .*524  DBP & CPTh: *r =* .245; *p = .*192 DBP & CPTo: *r =* .064; *p = .*735 DBP (baseline 2) & CPTh: *r =* .314; *p = .*091 DBP (baseline 2) & CPTo: *r =* .266; *p = .*155  RRI & CPTh: *r = -.*215; *p = .*254 RRI & CPTo: *r = -.*316; *p = .*089 RRI (baseline 2) & CPTh: *r = -.*172; *p = .*363 RRI (baseline 2) & CPTo: *r = -.*258; *p = .*169  BRS & CPTh: *r = -.*037; *p = .*848 BRS & CPTo: *r = -.*113; *p = .*554 BRS (baseline 2) & CPTh: *r = -.*043; *p = .*822 BRS (baseline 2) & CPTo: *r = -.*177; *p = .*349  Stress during – Pain Baseline  **HC**  LF HRV during arithmetic & CPTh: *r =* .074; *p = .*72 LF HRV during arithmetic & CPTo: *r = -.*003; *p = .*989 LF HRV during hot water & CPTh: *r = -.*015; *p = .*939 LF HRV during hot water & CPTo: *r = -.*095; *p = .*623 LF HRV during cold water & CPTh: *r = -.*177; *p = .*368 LF HRV during cold water & CPTo: *r = -.*027; *p = .*893  HF HRV during arithmetic & CPTh: *r =* .051; *p = .*804 HF HRV during arithmetic & CPTo: *r =* .07; *p = .*734 HF HRV during hot water & CPTh: *r =* .087; *p = .*655 HF HRV during hot water & CPTo: *r =* .024; *p = .*902 HF HRV during cold water & CPTh: *r =* .121; *p = .*539 HF HRV during cold water & CPTo: *r =* .309; *p = .*11  SBP during arithmetic & CPTh: *r =* .319; *p = .*086 SBP during arithmetic & CPTo: *r =* .189; *p = .*318 SBP during hot water & CPTh: *r =* .313; *p = .*093 SBP during hot water & CPTo: *r =* .011; *p = .*956 SBP during cold water & CPTh: *r =* .273; *p = .*144 SBP during cold water & CPTo: *r =* .044; *p = .*818  DBP during arithmetic & CPTh: *r =* .301; *p = .*106 DBP during arithmetic & CPTo: ***r =* .475; *p = .*008** DBP during hot water & CPTh: *r =* .222; *p = .*239 DBP during hot water & CPTo: *r = -.*007; *p = .*97 DBP during cold water & CPTh: *r =* .225; *p = .*232 DBP during cold water & CPTo: *r =* .038; *p = .*841  RRI during arithmetic & CPTh: *r = -.*12; *p = .*529 RRI during arithmetic & CPTo: *r = -.*252; *p = .*179 RRI during hot water & CPTh: *r = -.*21; *p = .*266 RRI during hot water & CPTo: *r = -.*262; *p = .*163 RRI during cold water & CPTh: *r = -.*142; *p = .*454 RRI during cold water & CPTo: *r = -.*242; *p = .*197  BRS during arithmetic & CPTh: *r =* .008; *p = .*968 BRS during arithmetic & CPTo: *r = -.*16; *p = .*398 BRS during hot water & CPTh: *r = -.*003; *p = .*988 BRS during hot water & CPTo: *r = -.*089; *p = .*64 BRS during cold water & CPTh: *r = -.*185; *p = .*327 BRS during cold water & CPTo: *r = -.*117; *p = .*537  Stress after – Pain Baseline  **HC**  LF HRV after arithmetic & CPTh: *r = -.*16; *p = .*408 LF HRV after arithmetic & CPTo: *r = -.*253; *p = .*185 LF HRV after water & CPTh: *r = -.*051; *p = .*791 LF HRV after water & CPTo: *r = -.*048; *p = .*805  HF HRV after arithmetic & CPTh: *r = -.*16; *p = .*406 HF HRV after arithmetic & CPTo: *r = -.*258; *p = .*176 HF HRV after water & CPTh: *r =* .003; *p = .*987 HF HRV after water & CPTo: *r =* .09; *p = .*644  SBP after arithmetic & CPTh: *r =* .296; *p = .*113 SBP after arithmetic & CPTo: *r =* .14; *p = .*46 SBP after water & CPTh: *r =* .245; *p = .*191 SBP after water & CPTo: *r = -.*105; *p = .*582  DBP after arithmetic & CPTh: ***r =* .381; *p = .*038** DBP after arithmetic & CPTo: *r =* .241; *p = .*199 DBP after water & CPTh: *r =* .242; *p = .*198 DBP after water & CPTo: *r = -.*082; *p = .*666  RRI after arithmetic & CPTh: *r = -.*21; *p = .*265 RRI after arithmetic & CPTo: ***r = -.*382; *p = .*037** RRI after water & CPTh: *r = -.*176; *p = .*351 RRI after water & CPTo: *r = -.*277; *p = .*138  BRS after arithmetic & CPTh: *r = -.*099; *p = .*601 BRS after arithmetic & CPTo: *r = -.*155; *p = .*412  BRS after water & CPTh: *r = -.*048; *p = .*801 BRS after water & CPTo: *r = -.*118; *p = .*534 |
| Farmer et al. (2014) | N.A. | N.A. |
| Flor et al. (2004) | N.A. | N.A. |
| Galeazzi et al. (2001) | N.A. | N.A. |
| Garcia-Hernandez et al. (2022) | Stress baseline – Pain baseline  **CPP** (**ws,** without anti-depressants)  SC & PPTh: *r =* .107; *p = .*58  SC & PPTo: *r =* .208; *p = .*279  **CPP (ws,** with + without anti-depressants)  SC & SREP: ***r = -.*46; *p < .*05** | Stress baseline – Pain baseline  **HC**  SC & PPTh: *r = -.*182; *p = .*335  SC & PPTo: *r = -.*227; *p = .*228  SC & SREP: *r = -.*024; *p = .*898 |
| Geiss et al. (2012) | N.A. | N.A. |
| Granot et al. (2002) | N.A. | N.A. |
| Jarrett et al. (2014) | Stress baseline – Pain baseline  **CPP (loc)**  Cortisol & CPM: *r =* .03; *p* > .40  Stress after – Pain baseline  **CPP (loc)**  Cortisol & CPM: *r =* .04; *p* > .40 | Stress baseline – Pain baseline  **HC**  Cortisol & CPM: *r = -.*19; *p* > .40  Stress after – Pain baseline  **HC**  Cortisol & CPM: *r =* .19; *p* > .40 |
| Jarrett et al. (2016) | Stress baseline – Pain baseline  **CPP (loc)**  HF HRV & CPM: *r =* .24; *p* > .05  LF HRV & CPM: *r =* .08; *p > .*05  TP HRV & CPM: r = .11; *p > .*05  LF/HF HRV & CPM: *r = -.*13; *p > .*05  HR & CPM: *r = -.*08; *p > .*05 | Stress baseline – Pain baseline  **HC**  HF HRV & CPM: *r = -.*09; *p > .*05  LF HRV & CPM: *r = -.*17; *p > .*05  TP HRV & CPM: *r = -.*11; *p > .*05  LF/HF HRV & CPM: *r =* .11; *p > .*05  HR & CPM: *r =* .15; *p > .*05 |
| Kadetoff et al. (2007) | Stress baseline – Pain baseline  **CPP (ws)**  HR & PPTh: *r =* -.29; *p* > .05  SBP & PPTh: *r =* -.04; *p* > .05  DBP & PPTh: *r =* .06; *p* > .05  Stress baseline – Pain during  **CPP (ws)**  HR & PPTh: *r =* -.35; *p* > .05  SBP & PPTh: *r =* -.23; *p* > .05  DBP & PPTh: *r =* -.11; *p* > .05  Stress baseline – Pain after  **CPP (ws)**  HR & PPTh: ***r =* -.51; *p* < .05**  SBP & PPTh: *r =* -.24; *p* > .05  DBP & PPTh: *r =* -.24; *p* > .05  Stress during – Pain baseline  **CPP (ws)**  HR & PPTh: *r =* -.32; *p* > .05  SBP & PPTh: *r =* -.33; *p* > .05  DBP & PPTh: *r =* -.06; *p* > .05  Stress during – Pain during  **CPP (ws)**  HR & PPTh: *r =* -.30; *p* > .05  SBP & PPTh: *r =* -.30; *p* > .05  DBP & PPTh: *r =* -.03; *p* > .05  Stress during – Pain after  **CPP (ws)**  HR & PPTh: *r =* -.24; *p* > .05  SBP & PPTh: *r =* -.22; *p* > .05  DBP & PPTh: *r =* -.33; *p* > .05  Stress after – Pain baseline  **CPP (ws)**  HR & PPTh: *r =* -.35; *p* > .05  SBP & PPTh: *r =* .10; *p* > .05  DBP & PPTh: *r =* .04; *p* > .05  Stress after – Pain during  **CPP (ws)**  HR & PPTh: *r =* -.39; *p* > .05  SBP & PPTh: *r =* -.18; *p* > .05  DBP & PPTh: *r =* -.21; *p* > .05  Stress after – Pain after  **CPP (ws)**  HR & PPTh: *r =* -.56; *p* > .05  SBP & PPTh: *r =* -.04; *p* > .05  DBP & PPTh: *r =* -.24; *p* > .05 | Stress baseline – Pain baseline  **HC**  HR & PPTh: *r =* -.34; *p* > .05  SBP & PPTh: *r =* .31; *p* > .05  DBP & PPTh: *r =* -.24; *p* > .05  Stress baseline – Pain during  **HC**  HR & PPTh: *r =* -.27; *p* > .05  SBP & PPTh: *r =* -.11; *p* > .05  DBP & PPTh: *r =* -.17; *p* > .05  Stress baseline – Pain after  **HC**  HR & PPTh: *r =* -.41; *p* > .05  SBP & PPTh: *r =* .06; *p* > .05  DBP & PPTh: *r =* -.21; *p* > .05  Stress during – Pain baseline  **HC**  HR & PPTh: *r =* -.28; *p* > .05  SBP & PPTh: *r =* -.23; *p* > .05  DBP & PPTh: *r =* -.47; *p* > .05  Stress during – Pain during  **HC**  HR & PPTh: *r =* -.39; *p* > .05  SBP & PPTh: *r =* -.17; *p* > .05  DBP & PPTh: *r =* -.21; *p* > .05  Stress during – Pain after  **HC**  HR & PPTh: ***r =* -.56; *p* < .05**  SBP & PPTh: *r =* -.27; *p* > .05  DBP & PPTh: *r =* -.30; *p* > .05  Stress after – Pain baseline  **HC**  HR & PPTh: *r =* -.32; *p* > .05  SBP & PPTh: *r =* .04; *p* > .05  DBP & PPTh: *r =* -.26; *p* > .05  Stress after – Pain during  **HC**  HR & PPTh: *r =* -.11; *p* > .05  SBP & PPTh: *r =* .08; *p* > .05  DBP & PPTh: *r =* -.31; *p* > .05  Stress after – Pain after  **HC**  HR & PPTh: *r =* -.28; *p* > .05  SBP & PPTh: *r =* -.21; *p* > .05  DBP & PPTh: *r =* -.09; *p* > .05 |
| Kadetoff et al. (2010) | Stress baseline – Pain baseline **CPP (ws)** HR & PPTh: *r =* .12; *p* > .05 SBP & PPTh: *r =* -.16; *p* > .05 DBP & PPTh: *r =* -.14; *p* > .05 Cortisol & PPTh: *r =* -.18; *p* > .05 Stress baseline – Pain during **CPP (ws)** HR & PPTh: *r =* -.15; *p* > .05 SBP & PPTh: *r =* -.12; *p* > .05 DBP & PPTh: *r =* -.03; *p* > .05 Cortisol & PPTh: *r =* -.16; *p* > .05 Stress baseline – Pain after **CPP (ws)** HR & PPTh: *r =* .07; *p* > .05 SBP & PPTh: *r =* -.24; *p* > .05 DBP & PPTh: *r =* -.16; *p* > .05 Cortisol & PPTh: *r =* -.12; *p* > .05 Stress during – Pain baseline **CPP (ws)** HR & PPTh: *r =* -.17; *p* > .05 SBP & PPTh: *r =* -.34; *p* > .05 DBP & PPTh: *r =* -.14; *p* > .05 Cortisol & PPTh: *r =* -.20; *p* > .05 Stress during – Pain during **CPP (ws)** HR & PPTh: *r* < .01; *p* > .05 SBP & PPTh: *r =* -.29; *p* > .05 DBP & PPTh: *r =* .01; *p* > .05 Cortisol & PPTh: *r =* -.16; *p* > .05 Stress during – Pain after **CPP (ws)** HR & PPTh: *r =* -.29; *p* > .05 SBP & PPTh: ***r =* -.51; *p* < .05** DBP & PPTh: *r =* -.28; *p* > .05 Cortisol & PPTh: *r =* -.14; *p* > .05 Stress after – Pain baseline **CPP (ws)** HR & PPTh: *r =* -.36; *p* > .05 SBP & PPTh: *r =* .02; *p* > .05 DBP & PPTh: *r =* .02; *p* > .05 Cortisol & PPTh: *r =* -.06; *p* > .05 Stress after – Pain during **CPP (ws)** HR & PPTh: *r =* -.46; *p* > .05 SBP & PPTh: *r =* -.08; *p* > .05 DBP & PPTh: *r =* .14; *p* > .05 Cortisol & PPTh: *r =* -.09; *p* > .05 Stress after – Pain after **CPP (ws)** HR & PPTh: *r =* -.35; *p* > .05 SBP & PPTh: *r =* -.04; *p* > .05 DBP & PPTh: *r =* -.08; *p* > .05 Cortisol & PPTh: *r =* -.06; *p* > .05 | Stress baseline – Pain baseline **HC** HR & PPTh: *r =* -.07; *p* > .05 SBP & PPTh: *r =* -.11; *p* > .05 DBP & PPTh: *r =* .08; *p* > .05 Cortisol & PPTh: *r =* .28; *p* > .05 Stress baseline – Pain during **HC** HR & PPTh: *r =* .04; *p* > .05 SBP & PPTh: *r =* -.07; *p* > .05 DBP & PPTh: *r =* .10; *p* > .05 Cortisol & PPTh: *r =* .51; *p* > .05 Stress baseline – Pain after **HC** HR & PPTh: *r =* .02; *p* > .05 SBP & PPTh: *r =* -.07; *p* > .05 DBP & PPTh: *r =* .12; *p* > .05 Cortisol & PPTh: *r =* .42; *p* > .05 Stress during – Pain baseline **HC** HR & PPTh: *r =* -.29; *p* > .05 SBP & PPTh: *r =* .22; *p* > .05 DBP & PPTh: *r =* -.19; *p* > .05 Cortisol & PPTh: *r =* .23; *p* > .05 Stress during – Pain during **HC** HR & PPTh: *r =* -.09; *p* > .05 SBP & PPTh: *r =* .18; *p* > .05 DBP & PPTh: ***r =* -.25; *p* < .05** Cortisol & PPTh: *r =* .44; *p* > .05 Stress during – Pain after **HC** HR & PPTh: *r =* -.13; *p* > .05 SBP & PPTh: *r =* .06; *p* > .05 DBP & PPTh: *r =* -.14; *p* > .05 Cortisol & PPTh: *r =* .39; *p* > .05 Stress after – Pain baseline **HC** HR & PPTh: *r =* -.05; *p* > .05 SBP & PPTh: *r =* -.05; *p* > .05 DBP & PPTh: *r =* -.19; *p* > .05 Cortisol & PPTh: *r =* .39; *p* > .05 Stress after – Pain during **HC** HR & PPTh: *r =* .05; *p* > .05 SBP & PPTh: *r =* -.06; *p* > .05 DBP & PPTh: *r =* -.06; *p* > .05 Cortisol & PPTh: *r =* .54; *p* > .05 Stress after – Pain after **HC** HR & PPTh: *r =* -.09; *p* > .05 SBP & PPTh: *r =* -.06; *p* > .05 DBP & PPTh: *r =* -.12; *p* > .05 Cortisol & PPTh: ***r =* .55; *p* < .05** |
| Kim et al. (2015) | Δ Stress (baseline to during) – Pain baseline:  **CPP (ws)**  HF HRV & TSP: ***r = -.*50; *p* < .01** | N.A. |
| Larsson et al. (2008) | N.A. | N.A. |
| Löffler et al. (2023) | Stress baseline – Pain baseline  **CPP (ws/loc)**  DBP & EPTh: *r =* .14; *p = .*54 DBP & EPTo: *r = -.*02; *p = .*93 SBP & EPTh: *r =* .33; *p = .*14 SBP & EPTo: *r =* .12; *p = .*58 HR & EPTh: *r =* .22; *p = .*32 HR & EPTo: *r =* .17; *p = .*44  Stress baseline – Δ Pain (baseline to after)  **CPP (ws/loc)**  DBP & EPTh: *r = -.*21; *p = .*34 DBP & EPTo: *r = -.*07; *p = .*76 SBP & EPTh: *r = -.*28; *p = .*21 SBP & EPTo: *r =* .02; *p = .*94 HR & EPTh: *r =* .23; *p = .*31 HR & EPTo: *r =* .18; *p = .*41  Δ Stress (baseline to during) – Pain baseline  **CPP (ws/loc)**  DBP & EPTh: *r =* .02; *p = .*94 DBP & EPTo: ***r =* .44; *p = .*04** SBP & EPTh: *r = -.*08; *p = .*74 SBP & EPTo: *r =* .24; *p = .*28 HR & EPTh: *r = -.*01; *p = .*98 HR & EPTo: *r =* .13; *p = .*56  Δ Stress (baseline to during) - Δ Pain (baseline to after)  **CPP (ws/loc)**  DBP & EPTh: *r = -.*21; *p = .*34 DBP & EPTo: ***r = -.*58; *p = .*005** SBP & EPTh: *r =* .06; *p = .*79 SBP & EPTo: *r = -.*41; *p = .*06 HR & EPTh: *r = -.*20; *p = .*37 HR & EPTo: *r = -.*22; *p = .*33 | Stress baseline – Pain baseline  **HC**  DBP & EPTh: *r =* .12; *p = .*64 DBP & EPTo: *r =* .41; *p = .*09 SBP & EPTh: *r =* .31; *p = .*21 SBP & EPTo: ***r =* .69; *p* < .001** HR & EPTh: *r =* .13; *p = .*61 HR & EPTo: *r = -.*31; *p = .*21  Stress baseline – Δ Pain (baseline to after)  **HC**  DBP & EPTh: *r =* .23; *p = .*35 DBP & EPTo: *r = -.*25; *p = .*31 SBP & EPTh: *r =* .16; *p = .*52 SBP & EPTo: ***r = -.*51; *p = .*03** HR & EPTh: *r = -.*09; *p = .*73 HR & EPTo: *r =* .26; *p = .*29  Δ Stress (baseline to during) – Pain baseline  **HC**  DBP & EPTh: *r = -.*03; *p = .*91 DBP & EPTo: *r = -.*42; *p = .*08  SBP & EPTh: *r =* .16; *p = .*52 SBP & EPTo: *r = -.*26; *p = .*29 HR & EPTh: *r = -.*08; *p = .*74 HR & EPTo: *r =* .07; *p = .*77  Δ Stress (baseline to during) - Δ Pain (baseline to after)  **HC**  DBP & EPTh: *r = -.*12; *p = .*64 DBP & EPTo: *r =* .42; *p = .*08 SBP & EPTh: *r = -.*03; *p = .*91 SBP & EPTo: *r =* .39; *p = .*11 HR & EPTh: *r =* .07; *p = .*78 HR & EPTo: *r = -.*11; *p = .*66 |
| Lòpez-Lòpez et al. (2021) | Stress baseline – Pain baseline  **CPP (ws)**  HR & PPTh: *r = -.*546; *p = .*054  SBP & PPTh: *r =* .091; *p = .*768  HR & PPTo: *r = -.*381; *p = .*20  SBP & PPTo: *r =* .189; *p = .*537  ∆ Stress (baseline to during) – Pain baseline  **CPP (ws)**  HR & PPTh: *r = -.*383; *p = .*119  SBP & PPTh: *r = -.*016; *p = .*958  HR & PPTo: *r =* .259; *p = .*394  SBP & PPTo: *r =* .085; *p = .*782  ∆ Stress (baseline to after) – Pain baseline  **CPP (ws)**  HR & PPTh: ***r = -.*561; *p = .*046**  SBP & PPTh: *r =* .158; *p = .*607  HR & PPTo: *r = -.*013; *p = .*966  SBP & PPTo: *r = -.*324; *p = .*281  Stress baseline - ∆ Pain (baseline to during)  **CPP (ws)**  HR & PPTh: *r =* .088; *p = .*775  SBP & PPTh: *r = -.*361; *p = .*225  HR & PPTo: *r =* .098; *p = .*749  SBP & PPTo: *r = -.*135; *p = .*66  Stress baseline - ∆ Pain (baseline to after)  **CPP (ws)**  HR & PPTh: *r = -.*10; *p = .*746  SBP & PPTh: *r = -.*388; *p = .*19  HR & PPTo: *r = -.*045; *p = .*741  SBP & PPTo: *r = -.*009; *p = .*858  ∆ Stress (baseline to during) - ∆ Pain (baseline to during)  **CPP (ws)**  HR & PPTh: *r = -.*230; *p = .*101  SBP & PPTh: *r =* .062; *p = .*842  HR & PPTo: *r =* .561; *p = .*837  SBP & PPTo: *r =* .034; *p = .*913  ∆ Stress (baseline to after) - ∆ Pain (baseline to during)  **CPP (ws)**  HR & PPTh: *r = -.*437; *p = .*136  SBP & PPTh: *r =* .334; *p = .*264  HR & PPTo: *r*  = .116; *p = .*706  SBP & PPTo: *r = -.*176; *p = .*565  ∆ Stress (baseline to during) - ∆ Pain (baseline to after)  **CPP (ws)**  HR & PPTh: *r = -.*225; *p = .*055  SBP & PPTh: *r =* .159; *p = .*603  HR & PPTo: *r =* .619; *p = .*782  SBP & PPTo: *r =* .098; *p = .*908  ∆ Stress (baseline to after) - ∆ Pain (baseline to after)  **CPP (ws)**  HR & PPTh: *r = -.*511; *p = .*074  SBP & PPTh: *r =* .421; *p = .*152  HR & PPTo: *r =* .287; *p* > .999  SBP & PPTo: *r = -.*274; *p = .*858  Stress during – Pain baseline  **CPP (ws)**  HR & PPTh: ***r = -.*584; *p = .*036**  SBP & PPTh: *r =* .054; *p = .*86  HR & PPTo: *r = -.*083; *p = .*786  SBP & PPTo: *r =* .191; *p = .*533  Stress after – Pain baseline  **CPP (ws)**  HR & PPTh: ***r = -.*671; *p = .*012**  SBP & PPTh: *r =* .147; *p = .*633  HR & PPTo: *r = -.*289; *p = .*338  SBP & PPTo: *r = -.*042; *p = .*892  Stress baseline – Pain during  **CPP (ws)**  HR & PPTh: ***r = -.*682: *p = .*01**  SBP & PPTh: *r =* .341; *p = .*254  HR & PPTo: *r = -.*398; *p = .*178  SBP & PPTo: *r =* .244; *p = .*422  Stress baseline – Pain after  **CPP (ws)**  HR & PPTh: ***r = -.*561; *p = .*046**  SBP & PPTh: *r =* .462; *p = .*112  HR & PPTo: *r = -.*332; *p = .*268  SBP & PPTo: *r =* .183; *p = .*55  Stress during – Pain during  **CPP (ws)**  HR & PPTh: ***r = -.*611; *p = .*027**  SBP & PPTh: *r =* .206; *p = .*50  HR & PPTo: *r = -.*297; *p = .*325  SBP & PPTo: *r =* .213; *p = .*484  Stress after – Pain during  **CPP (ws)**  HR & PPTh: ***r = -.*674; *p = .*012**  SBP & PPTh: *r =* .215; *p = .*481  HR & PPTo: *r = -.*331; *p = .*269  SBP & PPTo: *r =* .063; *p = .*837  Stress during – Pain after  **CPP (ws)**  HR & PPTh: *r = -.*514; *p = .*073  SBP & PPTh: *r =* .223; *p = .*464  HR & PPTo: *r = -.*288; *p = .*34  SBP & PPTo: *r =* .145; *p = .*635  Stress after – Pain after  **CPP (ws)**  HR & PPTh: *r = -.*514; *p = .*072  SBP & PPTh: *r =* .215; *p = .*48  HR & PPTo: *r = -.*333; *p = .*266  SBP & PPTo: *r =* .051; *p = .*869  Stress during - ∆ Pain (baseline to during)  **CPP (ws)**  HR & PPTh: *r = -.*085; *p = .*782  SBP & PPTh: *r = -.*218; *p = .*474  HR & PPTo: *r =* .408; *p = .*166  SBP & PPTo: *r = -.*075; *p = .*809  Stress after - ∆ Pain (baseline to during)  **CPP (ws)**  HR & PPTh: *r = -.*141; *p = .*645  SBP & PPTh: *r = -.*072; *p = .*816  HR & PPTo: *r =* .128; *p = .*676  SBP & PPTo: *r = -.*187; *p = .*541  Stress during - ∆ Pain (baseline to after)  **CPP (ws)**  HR & PPTh: *r = -.*202; *p = .*509  SBP & PPTh: *r = -.*174; *p = .*57  HR & PPTo: *r =* .352; *p = .*865  SBP & PPTo: *r =* .058; *p = .*748  Stress after - ∆ Pain (baseline to after)  **CPP (ws)**  HR & PPTh: *r = -.*316; *p = .*293  SBP & PPTh: *r = -.*044; *p = .*886  HR & PPTo: *r =* .102; *p = .*894  SBP & PPTo: *r = -.*152; *p = .*727  ∆ Stress (baseline to during) – Pain during  **CPP (ws)**  HR & PPTh: *r = -.*286; *p = .*196  SBP & PPTh: *r = -.*059; *p = .*847  HR & PPTo: *r = -.*069; *p = .*993  SBP & PPTo: *r =* .059; *p = .*848  ∆ Stress (baseline to during) – Pain after  **CPP (ws)**  HR & PPTh: *r = -.*252; *p = .*554  SBP & PPTh: *r = -.*165; *p = .*591  HR & PPTo: *r = -.*123; *p = .*906  SBP & PPTo: *r =* .022; *p = .*943  ∆ Stress (baseline to after) – Pain during  **CPP (ws)**  HR & PPTh: *r = -.*353; *p = .*237  SBP & PPTh: *r = -.*04; *p = .*896  HR & PPTo: *r = -.*075; *p = .*808  SBP & PPTo: *r = -.*198; *p = .*517  ∆ Stress (baseline to after) – Pain after  **CPP (ws)**  HR & PPTh: *r = -.*204; *p = .*503  SBP & PPTh: *r = -.*196; *p = .*522  HR & PPTo: *r = -.*183; *p = .*55  SBP & PPTo: *r = -.*142; *p = .*643 | Stress baseline – Pain baseline  **HC**  HR & PPTh: *r = -.*079; *p = .*653  SBP & PPTh: *r = -.*05; *p = .*744  HR & PPTo: *r = -.*189; *p = .*277  SBP & PPTo: *r =* .11; *p = .*531  ∆ Stress (baseline to during) – Pain baseline  **HC**  HR & PPTh: *r =* .031; *p = .*861  SBP & PPTh: *r = -.*021; *p = .*905  HR & PPTo: *r = -.*186; *p = .*285  SBP & PPTo: *r =* .038; *p = .*828  ∆ Stress (baseline to after) – Pain baseline  **HC**  HR & PPTh: *r =* .15; *p = .*389  SBP & PPTh: *r = -.*003; *p = .*612  HR & PPTo: *r = -.*066; *p = .*705  SBP & PPTo: *r =* .076; *p = .*147  Stress baseline - ∆ Pain (baseline to during)  **HC**  HR & PPTh: *r =* .179; *p = .*304  SBP & PPTh: ***r = -.*40; *p = .*017**  HR & PPTo: *r = -.*091; *p = .*604  SBP & PPTo: *r = -.*027; *p = .*879  Stress baseline - ∆ Pain (baseline to after)  **HC**  HR & PPTh: *r =* .207; *p = .*234  SBP & PPTh: *r = -.*180; *p = .*30  HR & PPTo: *r = -.*012; *p = .*945  SBP & PPTo: *r = -.*101; *p = .*564  ∆ Stress (baseline to during) - ∆ Pain (baseline to during)  **HC**  HR & PPTh: *r =* .048; *p = .*784  SBP & PPTh: *r =* .182; *p = .*294  HR & PPTo: *r =* .095; *p = .*589  SBP & PPTo: *r =* .115; *p = .*512  ∆ Stress (baseline to after) - ∆ Pain (baseline to during)  **HC**  HR & PPTh: *r = -.*241; *p = .*162  SBP & PPTh: *r = -.*019; *p = .*606  HR & PPTo: *r* = -.061; *p = .*729  SBP & PPTo: *r =* .077; *p = .*724  ∆ Stress (baseline to during) - ∆ Pain (baseline to after)  **HC**  HR & PPTh: *r =* .09; *p = .*606  SBP & PPTh: *r = -.*141; *p = .*418  HR & PPTo: *r =* .078; *p = .*654  SBP & PPTo: *r = -.*179; *p = .*305  ∆ Stress (baseline to after) - ∆ Pain (baseline to after)  **HC**  HR & PPTh: *r = -.*033; *p = .*85  SBP & PPTh: *r = -.*301; *p = .*155  HR & PPTo: *r = -.*034; *p = .*848  SBP & PPTo: *r = -.*149; *p = .*417  Stress during – Pain baseline  **HC**  HR & PPTh: *r = -.*042; *p = .*809  SBP & PPTh: *r = -.*052; *p = .*765  HR & PPTo: *r = -.*281; *p = .*103  SBP & PPTo: *r =* .109; *p = .*532  Stress after – Pain baseline  **HC**  HR & PPTh: *r = -.*021; *p = .*906  SBP & PPTh: *r = -.*049; *p = .*839  HR & PPTo: *r = -.*213; *p = .*219  SBP & PPTo: *r =* .155; *p = .*426  Stress baseline – Pain during  **HC**  HR & PPTh: *r = -.*177: *p = .*309  SBP & PPTh: *r =* .181; *p = .*297  HR & PPTo: *r = -.*121; *p = .*489  SBP & PPTo: *r =* .112; *p = .*523  Stress baseline – Pain after  **HC**  HR & PPTh: *r = -.*213; *p = .*219  SBP & PPTh: *r =* .086; *p = .*623  HR & PPTo: *r = -.*16; *p = .*357  SBP & PPTo: *r =* .16; *p = .*358  Stress during – Pain during  **HC**  HR & PPTh: *r = -.*142; *p = .*417  SBP & PPTh: *r =* .061; *p = .*73  HR & PPTo: *r = -.*246; *p = .*155  SBP & PPTo: *r =* .07; *p = .*691  Stress after – Pain during  **HC**  HR & PPTh: *r = -.*069; *p = .*695  SBP & PPTh: *r =* .173; *p = .*351  HR & PPTo: *r = -.*131; *p = .*454  SBP & PPTo: *r =* .122; *p = .*638  Stress during – Pain after  **HC**  HR & PPTh: *r = -.*198; *p = .*253  SBP & PPTh: *r =* .12; *p = .*492  HR & PPTo: *r = -.*277; *p = .*107  SBP & PPTo: *r =* .217; *p = .*21  Stress after – Pain after  **HC**  HR & PPTh: *r = -.*155; *p = .*375  SBP & PPTh: *r =* .228; *p = .*392  HR & PPTo: *r = -.*174; *p = .*317  SBP & PPTo: *r =* .26; *p = .*257  Stress during - ∆ Pain (baseline to during)  **HC**  HR & PPTh: *r =* ,177; *p = .*308  SBP & PPTh: *r = -.*192; *p = .*269  HR & PPTo: *r = -.*008; *p = .*964  SBP & PPTo: *r =* .053; *p = .*763  Stress after - ∆ Pain (baseline to during)  **HC**  HR & PPTh: *r =* .086; *p = .*625  SBP & PPTh: *r = -.*382; *p = .*088  HR & PPTo: *r = -.*114; *p = .*516  SBP & PPTo: *r =* .03; *p = .*546  Stress during - ∆ Pain (baseline to after)  **HC**  HR & PPTh: *r =* .229; *p = .*186  SBP & PPTh: *r = -.*23; *p = .*183  HR & PPTo: *r =* .044; *p = .*801  SBP & PPTo: *r = -.*193; *p = .*267  Stress after - ∆ Pain (baseline to after)  **HC**  HR & PPTh: *r =* .193; *p = .*267  SBP & PPTh: *r = -.*378; *p = .*181  HR & PPTo: *r = -.*025; *p = .*886  SBP & PPTo: *r = -.*197; *p = .*574  ∆ Stress (baseline to during) – Pain during  **HC**  HR & PPTh: *r =* .001; *p = .*993  SBP & PPTh: *r = -.*124; *p = .*477  HR & PPTo: *r = -.*215; *p = .*215  SBP & PPTo: *r = -.*026; *p = .*882  ∆ Stress (baseline to during) – Pain after  **HC**  HR & PPTh: *r = -.*039; *p = .*825  SBP & PPTh: *r =* .083; *p = .*635  HR & PPTo: *r = -.*214; *p = .*217  SBP & PPTo: *r =* .145; *p = .*407  ∆ Stress (baseline to after) – Pain during  **HC**  HR & PPTh: *r =* .281; *p = .*103  SBP & PPTh: *r =* .008; *p = .*96  HR & PPTo: *r = -.*027; *p = .*877  SBP & PPTo: *r =* .027; *p = .*198  ∆ Stress (baseline to after) – Pain after  **HC**  HR & PPTh: *r =* .149; *p = .*392  SBP & PPTh: *r =* .212; *p = .*188  HR & PPTo: *r = -.*038; *p = .*829  SBP & PPTo: *r =* .16; *p = .*238 |
| Maixner et al. (1997) | N.A. | Stress baseline – Pain baseline (ANOVA)  **HC (not in CPP)**  High BP subgroup & HPTo: **F(1,42) = 13.49; *p* < .001**  High BP subgroup & ischemic PTh: **F(1,43) = 8.54; *p* < .01**  High BP subgroup & ischemic PTo: **F(1,43) = 7.23; *p = .*01** |
| Meeus et al. (2008) | Stress baseline – Pain baseline (ascending) **CPP (ws)** Cortisol & mean pain intensity finger: *r =* .004; *p >* .999  Stress immediately after pain – Pain baseline (ascending) **CPP (ws)** Cortisol & mean pain intensity finger: *r = -.*107; *p = .*704  Δ Stress (baseline to immediately after pain) – Pain baseline (ascending)  **CPP (ws)** Cortisol & mean pain intensity finger: *r =* .131; *p = .*642  Stress baseline – Pain baseline (descending)  **CPP (ws)** Cortisol & mean pain intensity shoulder: *r =* .218; *p = .*434  Stress immediately after pain – Pain baseline (descending)  **CPP (ws)** Cortisol & mean pain intensity shoulder: *r = -.*512; *p = .*051  Δ Stress (baseline to immediately after pain) – Pain baseline (descending) **CPP (ws)** Cortisol & mean pain intensity shoulder: ***r =* .675; *p = .*006** | Stress baseline – Pain baseline (ascending)  **HC**  Cortisol & mean pain intensity finger: *r =* .003; *p = .*398  Stress immediately after pain – Pain baseline (ascending)  **HC** Cortisol & mean pain intensity finger: *r =* .226; *p = .*433  Δ Stress (baseline to immediately after pain) – Pain baseline (ascending)  **HC** Cortisol & mean pain intensity finger: *r = -.*181; *p = .*646  Stress baseline – Pain baseline (descending)  **HC** Cortisol & mean pain intensity shoulder: *r =* .186; p = .803  Stress immediately after pain – Pain baseline (descending)  **HC** Cortisol & mean pain intensity shoulder: *r = -.*071; *p = .*545  Δ Stress (baseline to immediately after pain) – Pain baseline (descending)  **HC** Cortisol & mean pain intensity shoulder: *r =* .261; *p = .*987 |
| Miyachi et al. (2025) | Stress Baseline – Pain Baseline  **CPP (loc)**  LF/HF & PPTh: *r* = .15; *p* = .33 | N.A. |
| Mohn et al. (2008) | Stress Baseline – Pain Baseline  **CPP (loc)** MAP & EPTh: ***r =* .52; *p* < .01** MAP & EPTo: ***r =* .47; *p* < .05** MAP & PPTh for Masseter muscle: *r =* .33; *p > .*05 MAP & PPTh for sternum: *r =* .35; *p > .*05 MAP & PPTo for Masseter muscle: *r =* .35; *p > .*05 MAP & PPTo for sternum: ***r =* .42; *p* < .05** HR & EPTh: *r = -.*03; *p > .*05 HR & EPTo: *r =* .13; *p > .*05 HR & PPTh for Masseter muscle: *r =* .29; *p > .*05 HR & PPTh for sternum: *r = -.*01; *p > .*05 HR & PPTo for Masseter muscle: *r =* .21; *p > .*05 HR & PPTo for sternum: *r = -.*07; *p > .*05  Stress After – Pain Baseline  **CPP (loc)** MAP & EPTh: ***r =* .59; *p* < .01** MAP & EPTo: ***r =* .55; *p* < .01** MAP & PPTh for Masseter muscle: *r =* .36; *p > .*05 MAP & PPTh for sternum: *r =* .40; *p > .*05 MAP & PPTo for Masseter muscle: *r =* .33; *p > .*05 MAP & PPTo for sternum: ***r =* .45; *p* < .05** HR & EPTh: *r =* .10; *p > .*05 HR & EPTo: *r =* .25; *p > .*05 HR & PPTh for Masseter muscle: *r =* .33; *p > .*05 HR & PPTh for sternum: *r =* .17; *p > .*05 HR & PPTo for Masseter muscle: *r =* .31; *p > .*05 HR & PPTo for sternum: *r =* .11; *p > .*05  Δ Stress (baseline to during) – Pain baseline  **CPP (loc)** MAP & EPTh: *r =* .20; *p > .*05 MAP & EPTo: *r =* .21; *p > .*05 MAP & PPTh for Masseter muscle: *r =* .09; *p > .*05 MAP & PPTh for sternum: *r =* .11; *p > .*05 MAP & PPTo for Masseter muscle: *r =* .02; *p > .*05 MAP & PPTo for sternum: ***r =* .10; *p* < .05** HR & EPTh: *r =* .21; *p > .*05 HR & EPTo: *r =* .24; *p > .*05 HR & PPTh for Masseter muscle: *r =* .15; *p > .*05 HR & PPTh for sternum: *r =* .29; *p > .*05 HR & PPTo for Masseter muscle: *r =* .23; *p > .*05 HR & PPTo for sternum: *r =* .28; *p > .*05 | Δ Stress (baseline to during) – Pain Baseline  **HC**  HR & EPTh: ***r =* .47; *p* < .05**  HR & EPTo: ***r =* .45; *p* < .05** |
| Muhtz et al. (2013) | Stress baseline – Pain baseline  **Across groups**  Cortisol & heat stimuli intensity: *r = -.*35; *p = .*75 | N.A. |
| Murray et al. (2004) | N.A. | N.A. |
| Nees et al. (2019) | Stress baseline – Pain baseline  **CPP (loc)**  Cortisol CARg & CPTh: *r = -.*178; *p = .*60  Cortisol CARg & HPTh: *r = -.*265; *p = .*431  Cortisol CARg & PPTh: *r =* .331; *p = .*269  Cortisol CARg & WUR: *r =* .273; *p = .*417  Cortisol CARi & CPTh: *r = -.*378; *p = .*252  Cortisol CARi & HPTh: *r =* .198; *p = .*559  Cortisol CARi & PPTh: *r =* .644; *p = .*017  Cortisol CARi & WUR: *r =* .326; *p = .*328  Cortisol CAR & CPTh: *r = -.*151; *p = .*659  Cortisol CAR & HPTh: *r = -.*245; *p = .*467  Cortisol CAR & PPTh: *r =* .148; *p = .*63  Cortisol CAR & WUR: *r =* .235; *p = .*487  Cortisol 11am & CPTh: *r =* .102; *p = .*766  Cortisol 11am & HPTh: *r = -.*048; *p = .*888  Cortisol 11am & PPTh: *r =* .127; *p = .*679  Cortisol 11am & WUR: *r = -.*151; *p = .*657  Cortisol 1pm & CPTh: *r = -.*068; *p = .*842  Cortisol 1pm & HPTh: *r =* .159; *p = .*64  Cortisol 1pm & PPTh: *r =* .531; *p = .*062  Cortisol 1pm & WUR: *r =* .076; *p = .*824  Cortisol 3pm & CPTh: *r = -.*282; *p = .*401  Cortisol 3pm & HPTh: *r =* .209; *p = .*537  Cortisol 3pm & PPTh: *r =* .115; *p = .*708  Cortisol 3pm & WUR: *r =* .223; *p = .*509  Cortisol 6pm & CPTh: *r =* .476; *p = .*138  Cortisol 6pm & HPTh: *r = -.*598; *p = .*052  Cortisol 6pm & PPTh: *r = -.*494; *p = .*086  Cortisol 6pm & WUR: *r =* .214; *p = .*527  Cortisol AUCg & CPTh: *r = -.*191; *p = .*574  Cortisol AUCg & HPTh: *r = -.*196; *p = .*563  Cortisol AUCg & PPTh: *r =* .357; *p = .*232  Cortisol AUCg & WUR: *r =* .298; *p = .*374  Cortisol AUCi & CPTh: *r = -.*408; *p = .*212  Cortisol AUCi & HPTh: *r =* .231; *p = .*494  Cortisol AUCi & PPTh: ***r =* .618; *p = .*024**  Cortisol AUCi & WUR: *r =* .403; *p = .*219 | Stress baseline – Pain baseline  **HC**  Cortisol CARg & CPTh: *r = -.*193; *p = .*415  Cortisol CARg & HPTh: *r =* .064; *p = .*795  Cortisol CARg & PPTh: *r =* .278; *p = .*235  Cortisol CARg & WUR: *r = -.*191; *p = .*42  Cortisol CARi & CPTh: *r = -.*393; *p = .*087  Cortisol CARi & HPTh: *r =* .192; *p = .*431  Cortisol CARi & PPTh: *r =* .232; *p = .*326  Cortisol CARi & WUR: *r = -.*147; *p = .*537  Cortisol CAR & CPTh: *r = -.*031; *p = .*896  Cortisol CAR & HPTh: *r = -.*099; *p = .*688  Cortisol CAR & PPTh: *r =* .237; *p = .*315  Cortisol CAR & WUR: *r = -.*157; *p = .*51  Cortisol 11am & CPTh: *r =* .297; *p = .*203  Cortisol 11am & HPTh: *r = -.*118; *p = .*631  Cortisol 11am & PPTh: *r = -.*152; *p = .*522  Cortisol 11am & WUR: *r = -.*221; *p = .*35  Cortisol 1pm & CPTh: *r =* .23; *p = .*344  Cortisol 1pm & HPTh: ***r = -.*463; *p = .*046**  Cortisol 1pm & PPTh: *r = -.*255; *p = .*291  Cortisol 1pm & WUR: *r = -.*285; *p = .*238  Cortisol 3pm & CPTh: *r = -.*308; *p = .*20  Cortisol 3pm & HPTh: *r = -.*111; *p = .*66  Cortisol 3pm & PPTh: *r = -.*024; *p = .*923  Cortisol 3pm & WUR: *r = -.*246; *p = .*31  Cortisol 6pm & CPTh: *r = -.*323; *p = .*177  Cortisol 6pm & HPTh: *r =* .038; *p = .*878  Cortisol 6pm & PPTh: *r =* .242; *p = .*319  Cortisol 6pm & WUR: *r = -.*271; *p = .*262  Cortisol AUCg & CPTh: *r = -.*203; *p = .*391  Cortisol AUCg & HPTh: *r = -.*023; *p = .*925  Cortisol AUCg & PPTh: *r =* .232; *p = .*326  Cortisol AUCg & WUR: *r = -.*267; *p = .*255  Cortisol AUCi & CPTh: ***r = -.*455; *p =* .044**  Cortisol AUCi & HPTh: *r =* .126; *p = .*608  Cortisol AUCi & PPTh: *r =* .229; *p = .*331  Cortisol AUCi & WUR: *r = -.*204; *p = .*388 |
| Ozgocmen et al. (2006) | Stress baseline – Pain baseline  **CPP (ws; not in HC)**  SSR latencies (hands) & PPTh: ***r =* .46; *p = .*012** | N.A. |
| Pardo et al. (2019) | N.A. | N.A. |
| Pickering et al. (2019) | Stress baseline – Pain baseline  SC & CPM: *r =* .17; *p = .*63  Or other measured parameters (*r* ranging from -.18 to .20, with *p >* .05) | N.A. |
| Poli Neto et al. (2020) | Stress baseline – Pain baseline  **CPP (loc)**  HR & PPTh: *r =* .19, *p = .*40  SBP & PPTh: *r =* .34; *p = .*13  DBP & PPTh: *r =* .38; *p = .*09  MBP & PPTh: *r =* .40; *p* = .075 | Stress baseline – Pain baseline  **HC**  HR & PPTh: *r = -.*20; *p = .*39  SBP & PPTh: *r = -.*38; *p = .*092  DBP & PPTh: *r = -.*23; *p = .*32  MBP & PPTh: *r = -.*36; *p = .*11 |
| Quartana et al. (2010) | N.A. | N.A. |
| Reshkova et al. (2015) | Stress baseline – Pain baseline  Noradrenaline & PPTh: *r* = .15; *p* < .05  Adrenaline & PPTh: *r* = .13; *p* < .05 | N.A. |
| Scheuren et al. (2023) | Stress during – Pain baseline  **CPP (loc)**  SSR & heat pain ratings: *r =* .13; *p = .*45 | Stress during – Pain baseline  **HC**  SSR & heat pain ratings: ***r =* .50; *p = .*004** |
| Tan et al. (2023) | Stress baseline – Pain baseline  **CPP (loc)**  SC & PPTh at acupoint 23 L: *r =* -.382; *p* = .294  SC & PPTh at acupoint 25 L: *r* = -.024; *p* = .931  SC & PPTh at acupoint 40 L: *r* = .343; *p* = .553  SC & PPTh at non-acupoint L: *r* = .275; *p* = .512  SC & PPTh at acupoint 23 R: *r* = -.115; *p* = .43  SC & PPTh at acupoint 25 R: *r* = .10; *p* = .505  SC & PPTh at acupoint 40 R: *r* = .106; *p* = .506  SC & PPTh at non-acupoint R: *r* = .18; *p* = .14 | Stress baseline – Pain baseline  **HC**  SC & PPTh at acupoint 23 L: *r =* .019; *p* = .681  SC & PPTh at acupoint 25 L: *r* = -.004; *p* = .73  SC & PPTh at acupoint 40 L: *r* = .256; *p* = .891  SC & PPTh at non-acupoint L: *r* = .18; *p* = .788  SC & PPTh at acupoint 23 R: *r* = -.023; *p* = .839  SC & PPTh at acupoint 25 R: *r* = -.012; *p* = .165  SC & PPTh at acupoint 40 R: *r* = -.031; *p* = 805  SC & PPTh at non-acupoint R: *r* = .088; *p* = .31 |
| Thieme et al. (2022) | N.A. | N.A. |
| Umeda et al. (2013) | Stress baseline – Pain baseline  **NFRT-responders**  MAP & NFRT: ***r =* .88; *p* < .01**  NS correlation between NFRT and other autonomic measure | Stress baseline – Pain baseline  **HC**  MAP & NFRT: *r =* .35; *p = .*23 |
| Valera-Calero et al. (2022) | Stress baseline – Pain baseline  **CPP (loc)**  Cortisol & PPTh: ***r = -.*259; *p* < .05**  SBP & PPTh: *r = -.*064; *p = .*54  DBP & PPTh: *r = -.*078, *p = .*455  HR & PPTh: *r = -.*191; *p = .*065 | N.A. |
| Van Den Houte et al. (2018) | Stress baseline – Pain baseline  **CPP (ws)**  RMSSD & CPM: *r =* .042; *p = .*72 | Stress baseline – Pain baseline  **HC**  RMSSD & CPM: r =0.18; *p = .*33 |
| Van Middendorp et al. (2013) | Stress baseline – Pain baseline  **CPP (ws)**  HR & EPTh: *r = -.*88; *p = .*495  MAP & EPTh: *r =* .243; *p = .*077  IBI & EPTh: *r = -.*001; = 0.994  PEP & EPTh: *r = -.*047; *p = .*722  HR & EPTo: *r = -.*128; *p = .*201  MAP & EPTo: *r =* .201; *p = .*144  IBI & EPTo: *r = -.*086; *p = .*514  PEP & EPTo: *r =* .027; *p = .*839 | Stress baseline – Pain baseline  **HC**  HR & EPTh: *r =* .061; *p = .*661  MAP & EPTh: *r =* .213; *p = .*17  IBI & EPTh: *r = -.*111; *p = .*427  PEP & EPTh: *r =* .085; *p = .*549  HR & EPTo: *r =* .123; *p = .*374  MAP & EPTo: *r =* .263; *p = .*089  IBI & EPTo: *r = -.*091; *p = .*517  PEP & EPTo: *r =* .117; *p = .*408 |
| Venezia et al. (2024) | Stress baseline – Pain baseline  **CPP (loc) + HC**  BRS & CPM(sham): *r* = -.21, *p* = .13 | |
| Wingenfeld et al. (2010) | Stress baseline – Pain baseline  Cortisol AUC & HPTh control: *r =* .01  Cortisol AUC & HPTh tender: *r = -.*07  Cortisol AUC & PPTh control: *r =* .05  Cortisol AUC & PPTh tender: *r = -.*03 | N.A. |
| Woda et al. (2013) | N.A. | N.A. |
| Zamunér et al. (2016) | Stress baseline – Pain baseline  **CPP (ws)**  RMSSD & PPTh: *r =* .38; *p = .*09  PSD & PPTh: *r = -.*17; *p = .*47  LF/HF & PPTh: *r = -.*20; *p = .*39  SBP & PPTh: *r = -.*30; *p = .*20  DBP & PPTh: *r = -.*23; *p = .*33  HR & PPTh: *r =* .15, *p = .*52  SSDN & PPTh: *r = -.*17, *p = .*47  LF & PPTh: *r = -.*10, *p = .*66  HF & PPTh: *r =* .23, *p = .*34  LFnu & PPTh: *r = -.*20, *p = .*39  HFnu & PPTh: *r =* .20, *p = .*39 | N.A. |

Abbreviations. ANS: Autonomic Nervous System, AUC: Area Under the Curve, BP: Blood Pressure, BRS: Baroreflex Sensitivity, CPM: Conditioned Pain Modulation, CPP: Chronic Primary Pain, CPT: Cold Pressor Test, CPTh: Cold Pain Threshold, CPTo: Cold Pain Tolerance, DBP: Diastolic Blood Pressure, DHEA: Dehydroepiandrosterone, EPTh: Electrical Pain Threshold, EPTo: Electrical Pain Tolerance, HC: Healthy Controls, HF(nu): High Frequency (normal unit) HRV, HPTh: Heat Pain Threshold, HPTo: Heat Pain Tolerance, HR(R): Heart Rate (Recovery), HRV: Heart Rate Variability, IBI: Inter-Beat-Interval, L: Left, LF(nu): Low Frequency (normal unit) HRV, MAP: Mean Arterial Pressure, MBP: Mean Blood Pressure, N.A.: Not Applicable, NFR(T): Nociceptive Flexor Reflex (Threshold), (P)PTh: Pressure Pain Threshold, (P)PTo: Pressure Pain Tolerance, PEP: Pre-Ejection Period, PSD: Total Power Spectral Density, R: Right, RMSSD: Root mean square of the differences between successive N-N intervals, RRI: R-R Interval, SBP: Systolic Blood Pressure, SC(R): Skin Conductance (Response), SCL: Skin Conductance Level, SREP: Slowly Repeated Evoked Pain, SSDN: Standard Deviation of Normal-to-Normal Intervals, SSR: Sympathetic Skin Response, TP: Total Power, trap: Trapezius muscle, TSP: Temporal Summation of Pain, WUR: Wind-Up Ratio, b: regression coefficient, loc: Localized pain, p: probability value, r: correlation coefficient, ws: Widespread pain, Δ: change in.
