## Supplementary 8 - Table S6 for "The Stress-Pain Connection in Chronic Primary Pain: A Systematic Review and Meta-Analysis of Physiological Stress Markers in Relation to Experimental Pain Responses"

**Supplementary File 8 - Table S6.** Certainty of evidence with GRADE of the qualitative analyses

| **Cluster** | **Outcome:**  **interaction**  **stress-pain** | **Study limitations**  **(0, -1)** | **Imprecision**  **(0, -1)** | **Inconsistency of results**  **(0, -1)** | **Indirectness of evidence**  **(0, -1)** | **Publication bias**  **(0, -1)** | **Certainty of the evidence** |
| --- | --- | --- | --- | --- | --- | --- | --- |
| 1 | HR-pain during CPT | -1 | 0 | 0 | 0 | 0 | ⨁◯◯◯ |
| 1 | HR-tenderness | 0 | 0 | 0 | 0 | 0 | ⨁⨁◯◯ |
| 1 | HR-PPTh | 0 | 0 | 0 | 0 | 0 | ⨁⨁◯◯ |
| 1 | HR-EPTh | 0 | 0 | -1 | 0 | 0 | ⨁◯◯◯ |
| 1 | HR-PPTo | 0 | 0 | 0 | 0 | 0 | ⨁⨁◯◯ |
| 1 | HR-EPTo | 0 | 0 | 0 | 0 | 0 | ⨁⨁◯◯ |
| 1 | HR-NFRT | 0 | 0 | 0 | 0 | 0 | ⨁⨁◯◯ |
| 1 | HR-CPM | 0 | 0 | 0 | 0 | 0 | ⨁⨁◯◯ |
| 1 | SBP-PPTh | -1 | 0 | 0 | 0 | 0 | ⨁◯◯◯ |
| 1 | SBP-CPTh | 0 | 0 | 0 | 0 | 0 | ⨁⨁◯◯ |
| 1 | SBP-EPTh | 0 | 0 | 0 | 0 | 0 | ⨁⨁◯◯ |
| 1 | SBP-Isch PTh | 0 | 0 | 0 | 0 | 0 | ⨁⨁◯◯ |
| 1 | SBP-PPTo | 0 | 0 | 0 | 0 | 0 | ⨁⨁◯◯ |
| 1 | SBP-CPTo | 0 | 0 | 0 | 0 | 0 | ⨁⨁◯◯ |
| 1 | SBP-EPTo | 0 | 0 | 0 | 0 | 0 | ⨁⨁◯◯ |
| 1 | SBP-Isch PTo | 0 | 0 | 0 | 0 | 0 | ⨁⨁◯◯ |
| 1 | SBP-HPTo | 0 | 0 | 0 | 0 | 0 | ⨁⨁◯◯ |
| 1 | SBP-SREP | 0 | 0 | 0 | 0 | 0 | ⨁⨁◯◯ |
| 1 | DBP-PPTh | 0 | 0 | 0 | 0 | 0 | ⨁⨁◯◯ |
| 1 | DBP-CPTh | 0 | 0 | 0 | 0 | 0 | ⨁⨁◯◯ |
| 1 | DBP-EPTh | 0 | 0 | 0 | 0 | 0 | ⨁⨁◯◯ |
| 1 | DBP-PPTo | 0 | 0 | 0 | 0 | 0 | ⨁⨁◯◯ |
| 1 | DBP-CPTo | 0 | 0 | 0 | 0 | 0 | ⨁⨁◯◯ |
| 1 | DBP-EPTo | 0 | 0 | 0 | 0 | 0 | ⨁⨁◯◯ |
| 1 | DBP-SREP | 0 | 0 | 0 | 0 | 0 | ⨁⨁◯◯ |
| 1 | MAP-PPTh | -1 | 0 | 0 | 0 | 0 | ⨁◯◯◯ |
| 1 | MAP-EPTh | 0 | 0 | 0 | 0 | 0 | ⨁⨁◯◯ |
| 1 | MAP-PPTo | 0 | 0 | 0 | 0 | 0 | ⨁⨁◯◯ |
| 1 | MAP-EPTo | 0 | 0 | 0 | 0 | 0 | ⨁⨁◯◯ |
| 1 | MAP-NFRT | 0 | 0 | 0 | 0 | 0 | ⨁⨁◯◯ |
| 1 | BRS-CPTh | 0 | 0 | 0 | 0 | 0 | ⨁⨁◯◯ |
| 1 | BRS-CPTo | 0 | 0 | 0 | 0 | 0 | ⨁⨁◯◯ |
| 1 | LF-tenderness | 0 | 0 | 0 | 0 | 0 | ⨁⨁◯◯ |
| 1 | LF-PPTh | 0 | 0 | 0 | 0 | 0 | ⨁⨁◯◯ |
| 1 | LF-CPTh | 0 | 0 | 0 | 0 | 0 | ⨁⨁◯◯ |
| 1 | LF-CPTo | 0 | 0 | 0 | 0 | 0 | ⨁⨁◯◯ |
| 1 | LF-CPM | 0 | 0 | 0 | 0 | 0 | ⨁⨁◯◯ |
| 1 | HF-trenderness | 0 | 0 | 0 | 0 | 0 | ⨁⨁◯◯ |
| 1 | HF-PPTh | 0 | 0 | 0 | 0 | 0 | ⨁⨁◯◯ |
| 1 | HF-CPTh | 0 | 0 | 0 | 0 | 0 | ⨁⨁◯◯ |
| 1 | HF-CPTo | 0 | 0 | 0 | 0 | 0 | ⨁⨁◯◯ |
| 1 | HF-CPM | 0 | 0 | 0 | 0 | 0 | ⨁⨁◯◯ |
| 1 | LF/HF-tenderness | 0 | 0 | 0 | 0 | 0 | ⨁⨁◯◯ |
| 1 | LF/HF-PPTh | 0 | 0 | 0 | 0 | 0 | ⨁⨁◯◯ |
| 1 | LF/HF-CPTh | 0 | 0 | 0 | 0 | 0 | ⨁⨁◯◯ |
| 1 | LF/HF-CPTo | 0 | 0 | 0 | 0 | 0 | ⨁⨁◯◯ |
| 1 | LF/HF-CPM | 0 | 0 | 0 | 0 | 0 | ⨁⨁◯◯ |
| 1 | PSD-PPTh | 0 | 0 | 0 | 0 | 0 | ⨁⨁◯◯ |
| 1 | SDNN-PPTh | 0 | 0 | 0 | 0 | 0 | ⨁⨁◯◯ |
| 1 | RMSSD-PPTh | 0 | 0 | 0 | 0 | 0 | ⨁⨁◯◯ |
| 1 | RMSSD-CPM | 0 | 0 | 0 | 0 | 0 | ⨁⨁◯◯ |
| 1 | TP-CPM | 0 | 0 | 0 | 0 | 0 | ⨁⨁◯◯ |
| 1 | IBI-EPTh | 0 | 0 | 0 | 0 | 0 | ⨁⨁◯◯ |
| 1 | IBI-EPTo | 0 | 0 | 0 | 0 | 0 | ⨁⨁◯◯ |
| 1 | PEP-EPTh | 0 | 0 | 0 | 0 | 0 | ⨁⨁◯◯ |
| 1 | PEP-EPTo | 0 | 0 | 0 | 0 | 0 | ⨁⨁◯◯ |
| 1 | RRI-CPTh | 0 | 0 | 0 | 0 | 0 | ⨁⨁◯◯ |
| 1 | RRI-CPTo | 0 | 0 | 0 | 0 | 0 | ⨁⨁◯◯ |
| 1 | HRV-tenderness | 0 | 0 | 0 | 0 | 0 | ⨁⨁◯◯ |
| 1 | SC-PPTh | 0 | 0 | 0 | 0 | 0 | ⨁⨁◯◯ |
| 1 | SSR-PPThs | 0 | 0 | 0 | 0 | 0 | ⨁⨁◯◯ |
| 1 | SC-PPTo | 0 | 0 | 0 | 0 | 0 | ⨁⨁◯◯ |
| 1 | SC-SREP | 0 |  | 0 | 0 | 0 | ⨁⨁◯◯ |
| 1 | SC-CPM | -1 | 0 | 0 | 0 | 0 | ⨁◯◯◯ |
| 1 | Cortisol-heat pain rating | 0 | 0 | -1 | 0 | 0 | ⨁⨁◯◯ |
| 1 | Cortisol-PPTh | 0 | 0 | 0 | 0 | 0 | ⨁⨁◯◯ |
| 1 | Cortisol-CPTh | 0 | 0 | 0 | 0 | 0 | ⨁⨁◯◯ |
| 1 | Cortisol-HPTh | 0 | 0 | 0 | 0 | 0 | ⨁⨁◯◯ |
| 1 | Cortisol-PPTo | 0 | 0 | 0 | 0 | 0 | ⨁⨁◯◯ |
| 1 | Cortisol-CPM | 0 | 0 | 0 | 0 | 0 | ⨁⨁◯◯ |
| 1 | Cortisol-WUR | 0 | 0 | 0 | 0 | 0 | ⨁⨁◯◯ |
| 1 | CortisolAUC-PPTh | 0 | 0 | 0 | 0 | 0 | ⨁⨁◯◯ |
| 1 | CortisolAUC-CPTh | 0 | 0 | 0 | 0 | 0 | ⨁⨁◯◯ |
| 1 | CortisolAUC-HPTh | 0 | 0 | 0 | 0 | 0 | ⨁⨁◯◯ |
| 1 | CortisolAUC-WUR | 0 | 0 | 0 | 0 | 0 | ⨁⨁◯◯ |
| 1 | CortisolCAR-PPTh | 0 | 0 | 0 | 0 | 0 | ⨁⨁◯◯ |
| 1 | CortisolCAR-CPTh | 0 | 0 | 0 | 0 | 0 | ⨁⨁◯◯ |
| 1 | CortisolCAR-HPTh | 0 | 0 | 0 | 0 | 0 | ⨁⨁◯◯ |
| 1 | CortisolCAR-WUR | 0 | 0 | 0 | 0 | 0 | ⨁⨁◯◯ |
| 1 | Hair cortisol-PPTh | 0 | 0 | 0 | 0 | 0 | ⨁⨁◯◯ |
| 1 | Hair cortisol-PPTo | 0 | 0 | 0 | 0 | 0 | ⨁⨁◯◯ |
| 1 | Hair cortisol-SREP | 0 | 0 | 0 | 0 | 0 | ⨁⨁◯◯ |
| 1 | DHEA-PPTh | 0 | 0 | 0 | 0 | 0 | ⨁⨁◯◯ |
| 1 | DHEA-PPTo | 0 | 0 | 0 | 0 | 0 | ⨁⨁◯◯ |
| 1 | Epinephrine-PPTh | -1 | -1 | 0 | 0 | 0 | ⨁◯◯◯ |
| 1 | Norepinephrine-PPTh | -1 | -1 | 0 | 0 | 0 | ⨁◯◯◯ |
| 2 a | HR-pain during CPT | -1 | 0 | 0 | 0 | 0 | ⨁◯◯◯ |
| 2 a | HR-PPTh | -1 | 0 | 0 | 0 | 0 | ⨁◯◯◯ |
| 2 a | HR-PPTo | 0 | 0 | 0 | 0 | 0 | ⨁⨁◯◯ |
| 2 a | SBP-PPTh | -1 | 0 | 0 | 0 | 0 | ⨁◯◯◯ |
| 2 a | SBP-CPTh | 0 | 0 | 0 | 0 | 0 | ⨁⨁◯◯ |
| 2 a | SBP-PPTo | 0 | 0 | 0 | 0 | 0 | ⨁⨁◯◯ |
| 2 a | SBP-CPTo | 0 | 0 | 0 | 0 | 0 | ⨁⨁◯◯ |
| 2 a | DBP-PPTh | -1 | 0 | 0 | 0 | 0 | ⨁◯◯◯ |
| 2 a | DBP-CPTh | 0 | 0 | 0 | 0 | 0 | ⨁⨁◯◯ |
| 2 a | DBP-CPTo | 0 | 0 | 0 | 0 | 0 | ⨁⨁◯◯ |
| 2 a | BRS-CPTh | 0 | 0 | 0 | 0 | 0 | ⨁⨁◯◯ |
| 2 a | BRS-CPTo | 0 | 0 | 0 | 0 | 0 | ⨁⨁◯◯ |
| 2 a | LF-PPTh | 0 | 0 | 0 | 0 | 0 | ⨁⨁◯◯ |
| 2 a | LF-CPTh | 0 | 0 | 0 | 0 | 0 | ⨁⨁◯◯ |
| 2 a | LF-CPTo | 0 | 0 | 0 | 0 | 0 | ⨁⨁◯◯ |
| 2 a | HF-PPTh | 0 | 0 | 0 | 0 | 0 | ⨁⨁◯◯ |
| 2 a | HF-CPTh | 0 | 0 | 0 | 0 | 0 | ⨁⨁◯◯ |
| 2 a | HF-CPTo | 0 | 0 | 0 | 0 | 0 | ⨁⨁◯◯ |
| 2 a | LF/HF-PPTh | 0 | 0 | 0 | 0 | 0 | ⨁⨁◯◯ |
| 2 a | LF/HF-CPTh | 0 | 0 | 0 | 0 | 0 | ⨁⨁◯◯ |
| 2 a | LF/HF-CPTo | 0 | 0 | 0 | 0 | 0 | ⨁⨁◯◯ |
| 2 a | RRI-CPTh | 0 | 0 | 0 | 0 | 0 | ⨁⨁◯◯ |
| 2 a | RRI-CPTo | 0 | 0 | 0 | 0 | 0 | ⨁⨁◯◯ |
| 2 a | SSR-heat pain rating | 0 | 0 | 0 | 0 | 0 | ⨁⨁◯◯ |
| 2 a | Cortisol-PPTh | -1 | 0 | 0 | 0 | 0 | ⨁◯◯◯ |
| 2 a | Cortisol-heat pain rating | 0 | 0 | 0 | 0 | 0 | ⨁⨁◯◯ |
| 2 a | Cortisol-CPM | 0 | 0 | 0 | 0 | 0 | ⨁⨁◯◯ |
| 2 b | HR-PPTh | 0 | 0 | 0 | 0 | 0 | ⨁⨁◯◯ |
| 2 b | HR-CPTh | 0 | 0 | 0 | 0 | 0 | ⨁⨁◯◯ |
| 2 b | HR-EPTh | 0 | 0 | 0 | 0 | 0 | ⨁⨁◯◯ |
| 2 b | HR-PPTo | 0 | 0 | 0 | 0 | 0 | ⨁⨁◯◯ |
| 2 b | HR-EPTo | 0 | 0 | 0 | 0 | 0 | ⨁⨁◯◯ |
| 2 b | HR-WUR | 0 | 0 | 0 | 0 | 0 | ⨁⨁◯◯ |
| 2 b | SBP-PPTh | -1 | 0 | 0 | 0 | 0 | ⨁◯◯◯ |
| 2 b | SBP-CPTh | 0 | 0 | 0 | 0 | 0 | ⨁⨁◯◯ |
| 2 b | SBP-PPTo | 0 | 0 | 0 | 0 | 0 | ⨁⨁◯◯ |
| 2 b | SBP-CPTo | 0 | 0 | 0 | 0 | 0 | ⨁⨁◯◯ |
| 2 b | DBP-PPTh | -1 | 0 | 0 | 0 | 0 | ⨁◯◯◯ |
| 2 b | DBP-CPTh | 0 | 0 | 0 | 0 | 0 | ⨁⨁◯◯ |
| 2 b | DBP-CPTo | 0 | 0 | 0 | 0 | 0 | ⨁⨁◯◯ |
| 2 b | MAP-PPTh | 0 | 0 | 0 | 0 | 0 | ⨁⨁◯◯ |
| 2 b | MAP-EPTh | 0 | 0 | 0 | 0 | 0 | ⨁⨁◯◯ |
| 2 b | MAP-PPTo | 0 | 0 | 0 | 0 | 0 | ⨁⨁◯◯ |
| 2 b | MAP-EPTo | 0 | 0 | 0 | 0 | 0 | ⨁⨁◯◯ |
| 2 b | BRS-CPTh | 0 | 0 | 0 | 0 | 0 | ⨁⨁◯◯ |
| 2 b | BRS-CPTo | 0 | 0 | 0 | 0 | 0 | ⨁⨁◯◯ |
| 2 b | LF-PPTh | 0 | 0 | 0 | 0 | 0 | ⨁⨁◯◯ |
| 2 b | LF-CPTh | 0 | 0 | 0 | 0 | 0 | ⨁⨁◯◯ |
| 2 b | LF-CPTo | 0 | 0 | 0 | 0 | 0 | ⨁⨁◯◯ |
| 2 b | HF-PPTh | 0 | 0 | 0 | 0 | 0 | ⨁⨁◯◯ |
| 2 b | HF-CPTh | 0 | 0 | 0 | 0 | 0 | ⨁⨁◯◯ |
| 2 b | HF-CPTo | 0 | 0 | 0 | 0 | 0 | ⨁⨁◯◯ |
| 2 b | LF/HF-PPTh | 0 | 0 | 0 | 0 | 0 | ⨁⨁◯◯ |
| 2 b | LF/HF-CPTh | 0 | 0 | 0 | 0 | 0 | ⨁⨁◯◯ |
| 2 b | LF/HF-CPTo | 0 | 0 | 0 | 0 | 0 | ⨁⨁◯◯ |
| 2 b | RRI-CPTh | 0 | 0 | 0 | 0 | 0 | ⨁⨁◯◯ |
| 2 b | RRI-CPTo | 0 | 0 | 0 | 0 | 0 | ⨁⨁◯◯ |
| 2 b | Cortisol-PPTh | -1 | 0 | 0 | 0 | 0 | ⨁◯◯◯ |
| 3 a | HR-PPTh | 0 | 0 | 0 | 0 | 0 | ⨁⨁◯◯ |
| 3 a | HR-EPTh | 0 | 0 | 0 | 0 | 0 | ⨁⨁◯◯ |
| 3 a | HR-PPTo | 0 | 0 | 0 | 0 | 0 | ⨁⨁◯◯ |
| 3 a | HR-EPTo | 0 | 0 | 0 | 0 | 0 | ⨁⨁◯◯ |
| 3 a | SBP-PPTh | 0 | 0 | 0 | 0 | 0 | ⨁⨁◯◯ |
| 3 a | SBP-EPTh | 0 | 0 | 0 | 0 | 0 | ⨁⨁◯◯ |
| 3 a | SBP-PPTo | 0 | 0 | 0 | 0 | 0 | ⨁⨁◯◯ |
| 3 a | SBP-EPTo | 0 | 0 | 0 | 0 | 0 | ⨁⨁◯◯ |
| 3 a | SBP-CPM | -1 | 0 | 0 | 0 | 0 | ⨁◯◯◯ |
| 3 a | DBP-EPTh | 0 | 0 | 0 | 0 | 0 | ⨁⨁◯◯ |
| 3 a | DBP-EPTo | 0 | 0 | 0 | 0 | 0 | ⨁⨁◯◯ |
| 3 a | MAP-PPTh | 0 | 0 | 0 | 0 | 0 | ⨁⨁◯◯ |
| 3 a | MAP-EPTh | 0 | 0 | 0 | 0 | 0 | ⨁⨁◯◯ |
| 3 a | MAP-PPTo | 0 | 0 | 0 | 0 | 0 | ⨁⨁◯◯ |
| 3 a | MAP-EPTo | 0 | 0 | 0 | 0 | 0 | ⨁⨁◯◯ |
| 3 a | LF-PPTh | 0 | 0 | 0 | 0 | 0 | ⨁⨁◯◯ |
| 3 a | HF-PPTh | 0 | 0 | 0 | 0 | 0 | ⨁⨁◯◯ |
| 3 a | HF-TSP | 0 | 0 | 0 | 0 | 0 | ⨁⨁◯◯ |
| 3 a | LF/HF-PPTh | 0 | 0 | 0 | 0 | 0 | ⨁⨁◯◯ |
| 3 a | Cortisol-heat pain rating | 0 | 0 | 0 | 0 | 0 | ⨁⨁◯◯ |
| 3 b | HR-PPTh | 0 | 0 | 0 | 0 | 0 | ⨁⨁◯◯ |
| 3 b | HR-PPTo | 0 | 0 | 0 | 0 | 0 | ⨁⨁◯◯ |
| 3 b | SBP-PPTh | 0 | 0 | 0 | 0 | 0 | ⨁⨁◯◯ |
| 3 b | SBP-PPTo | 0 | 0 | 0 | 0 | 0 | ⨁⨁◯◯ |
| 3 b | LF-PPTh | 0 | 0 | 0 | 0 | 0 | ⨁⨁◯◯ |
| 3 b | HF-PPTh | 0 | 0 | 0 | 0 | 0 | ⨁⨁◯◯ |
| 3 b | LF/HF-PPTh | 0 | 0 | 0 | 0 | 0 | ⨁⨁◯◯ |
| 4 | HR-PPTh | 0 | 0 | 0 | 0 | 0 | ⨁⨁◯◯ |
| 4 | HR-EPTh | 0 | 0 | 0 | 0 | 0 | ⨁⨁◯◯ |
| 4 | HR-PPTo | 0 | 0 | 0 | 0 | 0 | ⨁⨁◯◯ |
| 4 | HR-EPTo | 0 | 0 | 0 | 0 | 0 | ⨁⨁◯◯ |
| 4 | SBP-PPTh | 0 | 0 | 0 | 0 | 0 | ⨁⨁◯◯ |
| 4 | SBP-EPTh | 0 | 0 | 0 | 0 | 0 | ⨁⨁◯◯ |
| 4 | SBP-PPTo | 0 | 0 | 0 | 0 | 0 | ⨁⨁◯◯ |
| 4 | SBP-EPTo | 0 | 0 | 0 | 0 | 0 | ⨁⨁◯◯ |
| 4 | DBP-EPTh | 0 | 0 | 0 | 0 | 0 | ⨁⨁◯◯ |
| 4 | DBP-EPTo | 0 | 0 | 0 | 0 | 0 | ⨁⨁◯◯ |
| 4 | LF-PPTh | 0 | 0 | 0 | 0 | 0 | ⨁⨁◯◯ |
| 4 | HF-PPTh | 0 | 0 | 0 | 0 | 0 | ⨁⨁◯◯ |
| 4 | LF/HF-PPTh | 0 | 0 | 0 | 0 | 0 | ⨁⨁◯◯ |
| 5 a | HR-PPTh | 0 | 0 | 0 | 0 | 0 | ⨁⨁◯◯ |
| 5 a | HR-EPTh | 0 | 0 | 0 | 0 | 0 | ⨁⨁◯◯ |
| 5 a | HR-PPTo | 0 | 0 | 0 | 0 | 0 | ⨁⨁◯◯ |
| 5 a | HR-EPTo | 0 | 0 | 0 | 0 | 0 | ⨁⨁◯◯ |
| 5 a | SBP-PPTh | 0 | 0 | 0 | 0 | 0 | ⨁⨁◯◯ |
| 5 a | SBP-EPTh | 0 | 0 | 0 | 0 | 0 | ⨁⨁◯◯ |
| 5 a | SBP-PPTo | 0 | 0 | 0 | 0 | 0 | ⨁⨁◯◯ |
| 5 a | SBP-EPTo | 0 | 0 | 0 | 0 | 0 | ⨁⨁◯◯ |
| 5 a | DBP-EPTh | 0 | 0 | 0 | 0 | 0 | ⨁⨁◯◯ |
| 5 a | DBP-EPTo | 0 | 0 | 0 | 0 | 0 | ⨁⨁◯◯ |
| 5 a | LF-PPTh | 0 | 0 | 0 | 0 | 0 | ⨁⨁◯◯ |
| 5 a | HF-PPTh | 0 | 0 | 0 | 0 | 0 | ⨁⨁◯◯ |
| 5 a | LF/HF-PPTh | 0 | 0 | 0 | 0 | 0 | ⨁⨁◯◯ |
| 5 a | PEP-PPTh | 0 | 0 | 0 | 0 | 0 | ⨁⨁◯◯ |
| 5 a | SC-PPTh | 0 | 0 | 0 | 0 | 0 | ⨁⨁◯◯ |
| 5 a | SSR-PPTh | 0 | 0 | 0 | 0 | 0 | ⨁⨁◯◯ |
| 5 b | HR-PPTh | 0 | 0 | 0 | 0 | 0 | ⨁⨁◯◯ |
| 5 b | HR-PPTo | 0 | 0 | 0 | 0 | 0 | ⨁⨁◯◯ |
| 5 b | SBP-PPTh | 0 | 0 | 0 | 0 | 0 | ⨁⨁◯◯ |
| 5 b | SBP-PPTo | 0 | 0 | 0 | 0 | 0 | ⨁⨁◯◯ |
| 5 b | LF-PPTh | 0 | 0 | 0 | 0 | 0 | ⨁⨁◯◯ |
| 5 b | HF-PPTh | 0 | 0 | 0 | 0 | 0 | ⨁⨁◯◯ |
| 5 b | LF/HF-PPTh | 0 | 0 | 0 | 0 | 0 | ⨁⨁◯◯ |

Abbreviations. AUC: Aa Under the Curve, BRS: Baroreflex Sensitivity, CAR: Cortisol Awakening Response, CPM: Conditioned Pain Modulation, CPT: Cold Pressor Task, CPTh: Cold Pain Threshold, CPTo: Cold Pain Tolerance, DBP: Diastolic Blood Pressure, DHEA: Dehydroepiandrosterone, EPTh: Electrical Pain Threshold, EPTo: Electrical Pain Tolerance, GRADE: Grading of Recommendations, Assessments, Development and Evaluation, HF: High Frequency Heart Rate Variability, HPA: Hypothalamus-Pituitary-Adrenal axis, HPTh: Heat Pain Threshold, HPTo: Heat Pain Tolerance, HR: Heart Rate, HRV: Heart Rate Variability, IBI: Inter Beat Interval, Isch. PTh: Ischemic Pain Threshold, Isch. PTo: Ischemic Pain Tolerance, LF: Low Frequency Heart Rate Variability, LF/HF: Low Frequency/High Frequency Heart Rate Variability Ratio, MAP: Mean Arterial Pressure, NFRT: Nociceptive Flexor Reflex Threshold, PEP: Pre Ejection Period, PPTh: Pressure Pain Threshold, PPTo: Pressure Pain Tolerance, PSD: Power Spectral Density, RMSSD: Root Mean Square of Successive Differences, RRI: R-R Interval, SBP: Systolic Blood Pressure, SC: Skin Conductance, SDNN: Standard Deviation of NN Intervals, SREP: Slowly Repeated Evoked Pain, SSR: Sympathetic Skin Response, TP: Total Power, TSP: Temporal Summation of Pain, WUR: Wind-Up Ratio.
