## Supplementary 9 - Table S7 for "The Stress-Pain Connection in Chronic Primary Pain: A Systematic Review and Meta-Analysis of Physiological Stress Markers in Relation to Experimental Pain Responses"

**Supplementary File 9 - Table S8.** Certainty of evidence with GRADE of the quantitative analyses

| **Cluster** | **Outcome** | | **Population** | **Study limitations**  **(0, -1)** | **Imprecision**  **(0, -1)** | **Inconsistency of results**  **(0, -1, -2)** | **Indirectness of evidence**  **(0, -1)** | **Publication bias**  **(0, -1)** | **Large effect size (0, +1)** | **Certainty of the evidence** |
| --- | --- | --- | --- | --- | --- | --- | --- | --- | --- | --- |
|  | **Stress** | **Pain** |  |  |  |  |  |  |  |  |
| 1 | HR at baseline | PPTh at baseline | CPP | 0 | - 1 | - 1 | 0 | 0 | 0 | ⨁◯◯◯ |
|  |  |  | CPP – widespread | 0 | - 1 | - 1 | 0 | 0 | 0 | ⨁◯◯◯ |
|  |  |  | CPP – localized | 0 | - 1 | - 1 | 0 | 0 | 0 | ⨁◯◯◯ |
|  |  |  | HC | 0 | - 1 | 0 | 0 | 0 | 0 | ⨁◯◯◯ |
| 1 | HR at baseline | PPTh during stressor | CPP – widespread | 0 | 0 | - 1 | 0 | 0 | + 1 | ⨁⨁◯◯ |
|  |  |  | HC | 0 | - 1 | 0 | 0 | 0 | 0 | ⨁◯◯◯ |
| 1 | HR at baseline | PPTh during recovery | CPP – widespread | 0 | - 1 | - 2 | 0 | 0 | + 1 | ⨁◯◯◯ |
|  |  |  | HC | 0 | - 1 | 0 | 0 | 0 | 0 | ⨁◯◯◯ |
| 1 | HR at baseline | EPTh at baseline | CPP | 0 | - 1 | - 2 | 0 | 0 | + 1 | ⨁◯◯◯ |
|  |  |  | HC | 0 | - 1 | 0 | 0 | 0 | 0 | ⨁◯◯◯ |
| 1 | HR at baseline | PPTo at baseline | CPP | 0 | - 1 | 0 | 0 | 0 | 0 | ⨁◯◯◯ |
| 1 | HR at baseline | EPTo at baseline | CPP | 0 | - 1 | 0 | 0 | 0 | 0 | ⨁◯◯◯ |
|  |  |  | HC | 0 | - 1 | - 2 | 0 | 0 | 0 | ⨁◯◯◯ |
| 1 | SBP at baseline | PPTh at baseline | CPP | 0 | - 1 | 0 | 0 | 0 | 0 | ⨁◯◯◯ |
|  |  |  | CPP – widespread | 0 | - 1 | 0 | 0 | 0 | 0 | ⨁◯◯◯ |
|  |  |  | CPP – localized | 0 | - 1 | - 2 | 0 | 0 | 0 | ⨁◯◯◯ |
|  |  |  | HC | 0 | - 1 | - 2 | 0 | 0 | 0 | ⨁◯◯◯ |
| 1 | SBP at baseline | PPTh during stressor | CPP – widespread | 0 | - 1 | - 1 | 0 | 0 | 0 | ⨁◯◯◯ |
|  |  |  | HC | 0 | - 1 | 0 | 0 | 0 | 0 | ⨁◯◯◯ |
| 1 | SBP at baseline | PPTh during recovery | CPP – widespread | 0 | - 1 | - 2 | 0 | 0 | 0 | ⨁◯◯◯ |
|  |  |  | HC | 0 | - 1 | 0 | 0 | 0 | 0 | ⨁◯◯◯ |
| 1 | SBP at baseline | CPTh at baseline | CPP – widespread | 0 | - 1 | 0 | 0 | 0 | 0 | ⨁◯◯◯ |
| 1 | SBP at baseline | PPTo at baseline | CPP – widespread | 0 | - 1 | 0 | 0 | 0 | 0 | ⨁◯◯◯ |
|  |  |  | HC | 0 | - 1 | 0 | 0 | 0 | 0 | ⨁◯◯◯ |
| 1 | SBP at baseline | CPTo at baseline | CPP – widespread | 0 | - 1 | 0 | 0 | 0 | 0 | ⨁◯◯◯ |
| 1 | DBP at baseline | PPTh at baseline | CPP | 0 | - 1 | 0 | 0 | 0 | 0 | ⨁◯◯◯ |
|  |  |  | CPP – widespread | 0 | - 1 | 0 | 0 | 0 | 0 | ⨁◯◯◯ |
|  |  |  | CPP – localized | 0 | - 1 | - 2 | 0 | 0 | 0 | ⨁◯◯◯ |
|  |  |  | HC | 0 | - 1 | - 1 | 0 | 0 | 0 | ⨁◯◯◯ |
| 1 | DBP at baseline | PPTh during stressor | CPP – widespread | - 1 | - 1 | 0 | 0 | 0 | 0 | ⨁◯◯◯ |
|  |  |  | HC | - 1 | - 1 | 0 | 0 | 0 | 0 | ⨁◯◯◯ |
| 1 | DBP at baseline | PPTh during recovery | CPP – widespread | - 1 | - 1 | 0 | 0 | 0 | 0 | ⨁◯◯◯ |
|  |  |  | HC | - 1 | - 1 | 0 | 0 | 0 | 0 | ⨁◯◯◯ |
| 1 | DBP at baseline | CPTh at baseline | CPP – widespread | 0 | - 1 | 0 | 0 | 0 | 0 | ⨁◯◯◯ |
| 1 | DBP at baseline | CPTo at baseline | CPP – widespread | 0 | - 1 | 0 | 0 | 0 | 0 | ⨁◯◯◯ |
| 1 | MAP at baseline | EPTh at baseline | CPP | 0 | 0 | - 1 | 0 | 0 | + 1 | ⨁⨁◯◯ |
| 1 | MAP at baseline | EPTo at baseline | CPP | 0 | 0 | - 1 | 0 | 0 | 0 | ⨁◯◯◯ |
| 1 | BRS at baseline | CPTh at baseline | CPP – widespread | 0 | - 1 | 0 | 0 | 0 | 0 | ⨁◯◯◯ |
| 1 | BRS at baseline | CPTo at baseline | CPP – widespread | 0 | - 1 | 0 | 0 | 0 | 0 | ⨁◯◯◯ |
| 1 | HF HRV at baseline | PPTh at baseline | CPP | 0 | - 1 | - 1 | 0 | 0 | 0 | ⨁◯◯◯ |
| 1 | HF HRV at baseline | CPTh at baseline | CPP – widespread | 0 | - 1 | 0 | 0 | 0 | 0 | ⨁◯◯◯ |
| 1 | HF HRV at baseline | CPTo at baseline | CPP – widespread | 0 | - 1 | 0 | 0 | 0 | 0 | ⨁◯◯◯ |
| 1 | LF HRV at baseline | PPTh at baseline | CPP | 0 | - 1 | 0 | 0 | 0 | 0 | ⨁◯◯◯ |
| 1 | LF HRV at baseline | CPTh at baseline | CPP – widespread | 0 | - 1 | 0 | 0 | 0 | 0 | ⨁◯◯◯ |
| 1 | LF HRV at baseline | CPTo at baseline | CPP – widespread | 0 | - 1 | 0 | 0 | 0 | 0 | ⨁◯◯◯ |
| 1 | LF/HF HRV at baseline | PPTh at baseline | CPP | 0 | - 1 | 0 | 0 | 0 | 0 | ⨁◯◯◯ |
|  |  |  | CPP – localized | 0 | - 1 | 0 | 0 | 0 | 0 | ⨁◯◯◯ |
| 1 | RRI at baseline | CPTh at baseline | CPP – widespread | 0 | - 1 | 0 | 0 | 0 | 0 | ⨁◯◯◯ |
| 1 | RRI at baseline | CPTo at baseline | CPP – widespread | 0 | - 1 | 0 | 0 | 0 | 0 | ⨁◯◯◯ |
| 1 | SC at baseline | PPTh at baseline | CPP | 0 | - 1 | 0 | 0 | 0 | 0 | ⨁◯◯◯ |
|  |  |  | HC | 0 | - 1 | - 1 | 0 | 0 | 0 | ⨁◯◯◯ |
| 1 | Epinephrine at baseline | PPTh at baseline | CPP – widespread | - 1 | - 1 | 0 | 0 | 0 | 0 | ⨁◯◯◯ |
| 1 | Norepinephrine at baseline | PPTh at baseline | CPP – widespread | - 1 | - 1 | 0 | 0 | 0 | 0 | ⨁◯◯◯ |
| 1 | Salivary cortisol at baseline | Heat pain ratings at baseline | CPP | 0 | - 1 | - 2 | 0 | 0 | 0 | ⨁◯◯◯ |
|  |  |  | HC | 0 | - 1 | - 2 | 0 | 0 | 0 | ⨁◯◯◯ |
| 1 | Cortisol at baseline | PPTh at baseline | CPP | 0 | 0 | 0 | 0 | 0 | 0 | ⨁⨁◯◯ |
|  |  |  | CPP – widespread | 0 | - 1 | 0 | 0 | 0 | + 1 | ⨁⨁◯◯ |
|  |  |  | CPP – localized | 0 | - 1 | - 2 | 0 | 0 | 0 | ⨁◯◯◯ |
|  |  |  | HC | 0 | - 1 | 0 | 0 | 0 | 0 | ⨁◯◯◯ |
| 1 | Cortisol AUCg at baseline | HPTh at baseline | CPP | 0 | - 1 | 0 | 0 | 0 | 0 | ⨁◯◯◯ |
| 1 | Cortisol AUCg at baseline | PPTh at baseline | CPP | 0 | - 1 | 0 | 0 | 0 | 0 | ⨁◯◯◯ |
| 2a | HR during a stressor | PPTh at baseline | CPP – widespread | 0 | 0 | 0 | 0 | 0 | + 1 | ⨁⨁⨁◯ |
|  |  |  | HC | 0 | - 1 | 0 | 0 | 0 | 0 | ⨁◯◯◯ |
| 2b | HR during recovery | PPTh at baseline | CPP | 0 | 0 | - 2 | 0 | 0 | + 1 | ⨁◯◯◯ |
|  |  |  | CPP – widespread | 0 | 0 | 0 | 0 | 0 | + 1 | ⨁⨁⨁◯ |
|  |  |  | HC | 0 | - 1 | 0 | 0 | 0 | 0 | ⨁◯◯◯ |
| 2a | HR during a stressor | PPTh during a stressor | CPP – widespread | 0 | - 1 | - 1 | 0 | 0 | + 1 | ⨁◯◯◯ |
|  |  |  | HC | 0 | - 1 | 0 | 0 | 0 | 0 | ⨁◯◯◯ |
| 2b | HR during recovery | PPTh during a stressor | CPP – widespread | 0 | 0 | 0 | 0 | 0 | + 1 | ⨁⨁⨁◯ |
|  |  |  | HC | 0 | - 1 | 0 | 0 | 0 | 0 | ⨁◯◯◯ |
| 2a | HR during a stressor | PPTh during recovery | CPP – widespread | 0 | 0 | 0 | 0 | 0 | + 1 | ⨁⨁⨁◯ |
|  |  |  | HC | 0 | 0 | 0 | 0 | 0 | 0 | ⨁⨁◯◯ |
| 2b | HR during recovery | PPTh during recovery | CPP – widespread | 0 | 0 | 0 | 0 | 0 | + 1 | ⨁⨁⨁◯ |
|  |  |  | HC: 71 | 0 | - 1 | 0 | 0 | 0 | 0 | ⨁◯◯◯ |
| 2a | SBP during a stressor | PPTh at baseline | CPP – widespread | 0 | - 1 | 0 | 0 | 0 | 0 | ⨁◯◯◯ |
|  |  |  | HC | 0 | - 1 | 0 | 0 | 0 | 0 | ⨁◯◯◯ |
| 2b | SBP during recovery | PPTh at baseline | CPP – widespread | 0 | - 1 | 0 | 0 | 0 | 0 | ⨁◯◯◯ |
|  |  |  | HC | 0 | - 1 | 0 | 0 | 0 | 0 | ⨁◯◯◯ |
| 2a | SBP during a stressor | PPTh during a stressor | CPP – widespread | 0 | - 1 | 0 | 0 | 0 | 0 | ⨁◯◯◯ |
|  |  |  | HC | 0 | - 1 | 0 | 0 | 0 | 0 | ⨁◯◯◯ |
| 2b | SBP during recovery | PPTh during a stressor | CPP – widespread | 0 | - 1 | 0 | 0 | 0 | 0 | ⨁◯◯◯ |
|  |  |  | HC | 0 | - 1 | 0 | 0 | 0 | 0 | ⨁◯◯◯ |
| 2a | SBP during a stressor | PPTh during recovery | CPP – widespread | 0 | - 1 | - 2 | 0 | 0 | 0 | ⨁◯◯◯ |
|  |  |  | HC | 0 | - 1 | 0 | 0 | 0 | 0 | ⨁◯◯◯ |
| 2b | SBP during recovery | PPTh during recovery | CPP – widespread | 0 | - 1 | 0 | 0 | 0 | 0 | ⨁◯◯◯ |
|  |  |  | HC | 0 | - 1 | 0 | 0 | 0 | 0 | ⨁◯◯◯ |
| 2a | SBP during a stressor | CPTh at baseline | CPP – widespread | 0 | - 1 | 0 | 0 | 0 | 0 | ⨁◯◯◯ |
| 2b | SBP during recovery | CPTh at baseline | CPP – widespread | 0 | - 1 | 0 | 0 | 0 | 0 | ⨁◯◯◯ |
| 2a | SBP during a stressor | CPTo at baseline | CPP – widespread | 0 | - 1 | 0 | 0 | 0 | 0 | ⨁◯◯◯ |
| 2b | SBP during recovery | CPTo at baseline | CPP – widespread | 0 | - 1 | 0 | 0 | 0 | 0 | ⨁◯◯◯ |
| 2a | DBP during a stressor | PPTh at baseline | CPP – widespread | - 1 | - 1 | 0 | 0 | 0 | 0 | ⨁◯◯◯ |
|  |  |  | HC | - 1 | - 1 | 0 | 0 | 0 | + 1 | ⨁◯◯◯ |
| 2b | DBP during recovery | PPTh at baseline | CPP – widespread | - 1 | - 1 | 0 | 0 | 0 | 0 | ⨁◯◯◯ |
|  |  |  | HC | - 1 | - 1 | 0 | 0 | 0 | 0 | ⨁◯◯◯ |
| 2a | DBP during a stressor | PPTh during a stressor | CPP – widespread | - 1 | - 1 | 0 | 0 | 0 | 0 | ⨁◯◯◯ |
|  |  |  | HC | - 1 | - 1 | 0 | 0 | 0 | 0 | ⨁◯◯◯ |
| 2b | DBP during recovery | PPTh during a stressor | CPP – widespread | - 1 | - 1 | 0 | 0 | 0 | 0 | ⨁◯◯◯ |
|  |  |  | HC | - 1 | - 1 | 0 | 0 | 0 | 0 | ⨁◯◯◯ |
| 2a | DBP during a stressor | PPTh during recovery | CPP – widespread | - 1 | - 1 | 0 | 0 | 0 | 0 | ⨁◯◯◯ |
|  |  |  | HC | - 1 | - 1 | 0 | 0 | 0 | 0 | ⨁◯◯◯ |
| 2b | DBP during recovery | PPTh during recovery | CPP – widespread | - 1 | - 1 | 0 | 0 | 0 | 0 | ⨁◯◯◯ |
|  |  |  | HC: 33 | - 1 | - 1 | 0 | 0 | 0 | 0 | ⨁◯◯◯ |
| 2a | DBP during a stressor | CPTh at baseline | CPP – widespread | 0 | - 1 | 0 | 0 | 0 | 0 | ⨁◯◯◯ |
| 2b | DBP during recovery | CPTh at baseline | CPP – widespread | 0 | - 1 | 0 | 0 | 0 | 0 | ⨁◯◯◯ |
| 2a | DBP during a stressor | CPTo at baseline | CPP – widespread | 0 | - 1 | 0 | 0 | 0 | 0 | ⨁◯◯◯ |
| 2b | DBP during recovery | CPTo at baseline | CPP – widespread | 0 | - 1 | 0 | 0 | 0 | 0 | ⨁◯◯◯ |
| 2a | BRS during a stressor | CPTh at baseline | CPP – widespread | 0 | - 1 | 0 | 0 | 0 | 0 | ⨁◯◯◯ |
| 2b | BRS during recovery | CPTh at baseline | CPP – widespread | 0 | - 1 | 0 | 0 | 0 | 0 | ⨁◯◯◯ |
| 2a | BRS during a stressor | CPTo at baseline | CPP – widespread | 0 | - 1 | 0 | 0 | 0 | 0 | ⨁◯◯◯ |
| 2b | BRS during recovery | CPTo at baseline | CPP – widespread | 0 | - 1 | 0 | 0 | 0 | 0 | ⨁◯◯◯ |
| 2a | HF HRV during a stressor | CPTh at baseline | CPP – widespread | 0 | - 1 | 0 | 0 | 0 | 0 | ⨁◯◯◯ |
| 2b | HF HRV during recovery | CPTh at baseline | CPP – widespread | 0 | - 1 | 0 | 0 | 0 | 0 | ⨁◯◯◯ |
| 2a | HF HRV during a stressor | CPTo at baseline | CPP – widespread | 0 | 0 | 0 | 0 | 0 | 0 | ⨁⨁◯◯ |
| 2b | HF HRV during recovery | CPTo at baseline | CPP – widespread | 0 | 0 | 0 | 0 | 0 | + 1 | ⨁⨁⨁◯ |
| 2a | LF HRV during a stressor | CPTh at baseline | CPP – widespread | 0 | - 1 | 0 | 0 | 0 | 0 | ⨁◯◯◯ |
| 2b | LF HRV during recovery | CPTh at baseline | CPP – widespread | 0 | - 1 | 0 | 0 | 0 | 0 | ⨁◯◯◯ |
| 2a | LF HRV during a stressor | CPTo at baseline | CPP – widespread | 0 | - 1 | 0 | 0 | 0 | 0 | ⨁◯◯◯ |
| 2b | LF HRV during recovery | CPTo at baseline | CPP – widespread | 0 | - 1 | 0 | 0 | 0 | 0 | ⨁◯◯◯ |
| 2a | RRI during a stressor | CPTh at baseline | CPP – widespread | 0 | - 1 | 0 | 0 | 0 | 0 | ⨁◯◯◯ |
| 2b | RRI during recovery | CPTh at baseline | CPP – widespread | 0 | - 1 | 0 | 0 | 0 | 0 | ⨁◯◯◯ |
| 2a | RRI during a stressor | CPTo at baseline | CPP – widespread | 0 | - 1 | 0 | 0 | 0 | 0 | ⨁◯◯◯ |
| 2b | RRI during recovery | CPTo at baseline | CPP – widespread | 0 | - 1 | 0 | 0 | 0 | 0 | ⨁◯◯◯ |
| 3a | ∆HR (during stressor – at baseline) | PPTh at baseline | CPP | 0 | - 1 | - 2 | 0 | 0 | 0 | ⨁◯◯◯ |
| 3a | ∆HR (during stressor – at baseline) | PPTo at baseline | CPP | 0 | - 1 | 0 | 0 | 0 | 0 | ⨁◯◯◯ |
| 3a | ∆HR (during stressor – at baseline) | EPTh at baseline | CPP – localized | 0 | - 1 | 0 | 0 | 0 | 0 | ⨁◯◯◯ |
|  |  |  | HC: 43 | 0 | - 1 | - 2 | 0 | 0 | 0 | ⨁◯◯◯ |
| 3a | ∆HR (during stressor – at baseline) | EPTo at baseline | CPP – localized | 0 | - 1 | 0 | 0 | 0 | 0 | ⨁◯◯◯ |
|  |  |  | HC | 0 | - 1 | - 1 | 0 | 0 | 0 | ⨁◯◯◯ |

Abbreviations. AUCg: Area Under the Curve (ground), BRS: Baroreflex Sensitivity, CPP: Chronic Primary Pain, CPTh: Cold Pain Threshold, CPTo: Cold Pain Tolerance, DBP: Diastolic Blood Pressure, EPTh: Electrical Pain Threshold, EPTo: Electrical Pain Tolerance, HC: Healthy Controls, HF: High Frequency, HPTh: Heat Pain Threshold, HR: Heart Rate, HRV: Heart Rate Variability, LF: Low Frequency, MAP: Mean Arterial Pressure, PPTh: Pressure Pain Threshold, PPTo: Pressure Pain Tolerance, RRI: R-R Interval, SBP: Systolic Blood Pressure, SC: Skin Conductance, ∆: change in.
