## Supplementary 10 - Results for "The Stress-Pain Connection in Chronic Primary Pain: A Systematic Review and Meta-Analysis of Physiological Stress Markers in Relation to Experimental Pain Responses"

### **Supplementary File 10.** Results

### **Study characteristics of all 52 studies**

The main study characteristics of all 52 included studies are summarized below, and an overview of the relevant data of each individual study can be found in Supplementary File 6 - Table S3.

##### ***Study Design and Population***

Five of the 52 studies had a cross-sectional design, and 47 studies had a case-control design. A total of 2,657 participants were included across the studies (*n* = 1,160 pain-free controls; *n* = 802 individuals with chronic widespread pain; *n* = 695 individuals with chronic localized pain). Samples sizes ranged from ten to 59 in the pain-free controls, from eight to 81 in the chronic widespread pain groups, and from ten to 94 in the chronic localized pain groups. The chronic widespread pain groups only consisted of patients with fibromyalgia and were investigated in 30 studies, except for one study that investigated a mix of chronic widespread and localized back pain patients (Loffler, et al., 2023). Chronic localized pain groups were investigated in 22 studies and consisted of chronic (low) back pain (Bandeira, et al., 2021; Flor, et al., 2004; Miyachi, et al., 2025; Muhtz, et al., 2013; Nees, et al., 2019; Tan, et al., 2023; Venezia, et al., 2024), chronic neck pain (Rampazo, et al., 2024; Valera-Calero & Varol, 2022), chronic neck-shoulder pain (Larsson, et al., 2008), temporomandibular disorders (Maixner, et al., 1997; Mohn, et al., 2008; Quartana, et al., 2010), irritable bowel syndrome (Chalaye, et al., 2012; Jarrett, et al., 2016; Jarrett, et al., 2014; Murray, et al., 2004), ulcerative colitis (Galeazzi, et al., 2001), vulvar vestibulitis (Granot, et al., 2002), chronic pelvic pain (Poli-Neto, et al., 2020), functional chest pain (Farmer, et al., 2014) and chronic complex regional pain syndrome (Scheuren, et al., 2023). The mean age ± SD was 39.9 ± 8.6 (range: 25.4-53.3) years for the pain-free controls, 46.8 ± 8.5 (range: 37.3-57.1) years for the chronic widespread pain group, and 38.9 ± 10.4 (range: 27.1-67.9) years for the chronic localized pain groups. However, several studies did not report the mean or SD of the participants’ age (Farmer, et al., 2014; Galeazzi, et al., 2001; Kadetoff & Kosek, 2007, 2010; Murray, et al., 2004; Reshkova, et al., 2015; Thieme, et al., 2022). In 30 of the 52 studies, only female participants were investigated. All 52 studies used baseline assessments, 23 studies reported data concerning reactivity, and 15 studies reported data concerning recovery of the stress systems. Twelve studies used a non-painful stressor and 15 studies used a painful stressor. Thirty-eight studies only investigated the ANS, 10 studies focused solely on the HPA axis, and four studies investigated both systems.

##### ***Physiological stress outcome measures***

***Baseline – ANS.*** Parameters of HRV were assessed in 16 studies, pre-ejection period (PEP) in three studies, HR frequency in 23 studies, BP and cardiac baroreflex sensitivity (BRS) in 24 studies, SC in eight studies, respiration rate in one study (Jonsson, et al., 2025), and (nor)epinephrine in three studies (Geiss, et al., 2012; Kadetoff & Kosek, 2010; Reshkova, et al., 2015). Parameters of HRV consisted of high frequency (HF) HRV, low frequency (LF) HRV, very HF (VHF) HRV, very LF (VLF) HRV, LF/HF ratio, total power, root mean square of successive differences between normal heartbeats (RMSSD) and the mean of standard deviations of the NN intervals (SDNN). Furthermore, cardiovagal tone (CVT), R-R interval (RRI) and the sympathovagal balance (SVB), which are closely associated with HRV, were also measured. The duration of the physiological ANS measurements ranged from 60 seconds to 20 minutes, with a duration of five (Bandeira, et al., 2021; Bossenger, et al., 2023; de la Coba, et al., 2018; Farmer, et al., 2014; Galeazzi, et al., 2001; Jarrett, et al., 2016; Loffler, et al., 2023; Lopez-Lopez, et al., 2021; Venezia, et al., 2024) or ten minutes (Crettaz, et al., 2013; Kadetoff & Kosek, 2010; Reyes Del Paso, et al., 2022; Reyes del Paso, et al., 2011; Van Den Houte, et al., 2018) most frequently reported.

***Baseline – HPA axis.*** Cortisol levels were assessed in 13 studies. One study assessed cortisol in hair (Davydov, et al., 2024). Four studies assessed cortisol in serum either in the morning (i.e., 8:00-9:30) or in the early afternoon (i.e., 14:00 - 16:00) (De Abreu Freitas, et al., 2012; Farmer, et al., 2014; Kadetoff & Kosek, 2010; Pardo, et al., 2019), one study collected urine from the night before and during the experiment to examine cortisol (Larsson, et al., 2008), and eight studies assessed cortisol in saliva at various timepoints: i) throughout the day (Geiss, et al., 2012; Nees, et al., 2019; Wingenfeld, et al., 2010), ii) in the morning (Jarrett, et al., 2014; Quartana, et al., 2010; Valera-Calero & Varol, 2022), iii) in the afternoon (Geiss, et al., 2012), and iv) before the start of pain assessments (Meeus, et al., 2008). One study did not specify the timing of the day (Muhtz, et al., 2013). Dehydroepiandrosterone (DHEA) was measured in one study (De Abreu Freitas, et al., 2012).

***Reactivity – ANS.*** Psychophysiological stressors were used to induce changes in ANS and consisted of non-painful (i.e., passive visualization task; arithmetic task; Trier Social Stress Test (TSST); personal involvement task; listening test; cold room exposure; and isometric contraction of the quadriceps muscle) or painful (i.e., cold pressor test, CPT; hot water, mechanical, thermal, somatic, visceral and pinprick stimuli; and wind-up ratio protocol) stressors and stress tasks. Twelve studies used a non-painful stressor, and fifteen studies used a painful stressor. During the stressor, parameters of HRV were examined in nine studies, HR and BP in 15 studies, SC in three studies (Bossenger, et al., 2023; Farmer, et al., 2014; Scheuren, et al., 2023), PEP in two studies (Bossenger, et al., 2023; Reyes Del Paso, et al., 2022), and (nor)epinephrine in one study (Kadetoff & Kosek, 2010).

***Reactivity – HPA axis.*** Two studies measured cortisol at exhaustion of an isometric quadriceps muscle contraction or during a cold room exposure (Kadetoff & Kosek, 2010; Pardo, et al., 2019), and four studies measured cortisol immediately to two minutes after a somatic or visceral pain stimuli, after an assessment of mechanical and thermal pain thresholds and after a conditioned pain modulation (CPM) procedure with thermal stimuli (Farmer, et al., 2014; Geiss, et al., 2012; Jarrett, et al., 2014; Meeus, et al., 2008; Quartana, et al., 2010).

***Recovery – ANS*.** The recovery of HRV and PEP was measured over a five-minute interval immediately after a CPT (Bandeira, et al., 2021; Reyes Del Paso, et al., 2022) recovery of HR and BP (sensitivity) ranged from immediately after a stressor for five minutes to 30 minutes after a stressor across studies (Crettaz, et al., 2013; Kadetoff & Kosek, 2010; Lopez-Lopez, et al., 2021; Reyes del Paso, et al., 2011; Woda, et al., 2013), and one study measured (nor)epinephrine one, ten, 20, 30, 90, 150, 195, 240, 285, 330 and 375 minutes after evaluation of pressure PTh (PPTh) (Geiss, et al., 2012).

***Recovery – HPA axis.*** The assessment of recovery of cortisol values ranged from 20 to 375 minutes after a psychophysiological stressor (Geiss, et al., 2012; Kadetoff & Kosek, 2010; Muhtz, et al., 2013; Quartana, et al., 2010).

##### ***Experimental pain outcome measures***

The most reported pain outcome measure was the static QST measure pressure pain threshold (PPTh), assessed at various body sites (in 27/52 studies). Different other static QST methods were used to assess pain sensitivity at various locations across the included studies. Cold PThs (CPThs) were assessed in six studies, heat PThs (HPThs) in seven studies, electrical PThs (EPThs) in six studies, and oesophageal PThs in one study. Cold pain tolerances (CPTos) were assessed in two studies, heat PTos in one study, electrical PTos (EPTos) in five studies, mechanical PTos in six studies, somatic PTos in one study, and visceral PTos in one study. Pain intensity ratings were used in three studies for cold stimuli, in three studies for heat stimuli, and in two studies for mechanical stimuli. Functionality of central pain processing using dynamic QST was evaluated in 15 studies. More specifically, temporal summation of pain (TSP) was assessed in four studies to evaluate pain facilitation, sustained response to evoked pain (SREP) in three studies to assess pain facilitation, CPM in seven studies to evaluate descending pain inhibition, and the nociceptive flexion reflex threshold (NFRT) was assessed in one study to evaluate spinal cord responses to noxious input.

All studies measured an experimental pain outcome measure at baseline. In three studies, PPThs were assessed during a stressor (isometric contraction of quadriceps muscle and TSST) (Kadetoff & Kosek, 2007, 2010; Lopez-Lopez, et al., 2021). In seven studies, PPThs, PPTos, EPThs, EPTos, CPThs, HPThs, or WUR were assessed after a stressor (which ranged from immediately after a stressor to 70 minutes after a stressor) (Bandeira, et al., 2021; Crettaz, et al., 2013; Kadetoff & Kosek, 2007, 2010; Loffler, et al., 2023; Lopez-Lopez, et al., 2021; Mohn, et al., 2008).

### **Synthesis of results in pain-free controls**

#### ***Baseline stress – pain (Cluster 1)***

***ANS: HR – Pain*.** No significant associations between HR and pain scores during CPT, EPTh, PPTh, PPTo, EPTo, NFRT, or CPM were found across six studies (all *p* > .05).

***ANS: SBP – Pain*.** A trend towards a positive association of SBP with static pain measures was observed across nine studies, with significant associations with PPTh, Ischemic PTh, PPTo, EPTo, ischemic PTo and HPTo measured at baseline in three studies (*r* = .38-69, all *p* < .05) (de la Coba, et al., 2018; Loffler, et al., 2023; Maixner, et al., 1997).

***ANS: DBP – Pain*.** One study reported a significant positive association with PPTh measured at baseline (*r* = .28, *p* < .05) (de la Coba, et al., 2018), whereas seven studies did not find report significant associations with PPTh, CPTh, EPTh, PPTo, CPTo, EPTo, or SREP (all *p* > .05).

***ANS: MAP – Pain*.** No significant associations were found with PPTh, EPTh, EPTo or NFRT (all *p* > .05) across three studies.

***ANS: BRS – Pain*.** Two studies examining the association between cardiac BRS and CPTh and CPTo at baseline found no significant associations (all *p* > .05).

***ANS: HRV and PEP – Pain*.** Across six studies, no significant association were found between parameters of HRV and PPTh, CPTh, CPTo, EPTh, EPTo or CPM at baseline (all *p* > .05).

***ANS: EDA – Pain*.** No significant associations were found with PPTh, PPTo, SREP or CPM (all *p* > .05).

***HPA – Pain.*** Across seven studies, only one study reported significant negative associations between cortisol measured at 13:00, and cortisol AUCi, with HPTh measured at baseline and CPTh measured at baseline, respectively (both *r* = -.46, both *p* < .05). One study investigated DHEA and its association with PPTh and PPTo, and found significantly positive associations (*r* = .69, *r* = .54 respectively, all *p* < .02) (De Abreu Freitas, et al., 2012).

#### ***Stress during or after a stressor – pain (Cluster 2a & 2b)***

***ANS: HR during stressor – Pain*.** One of three studies reported a significant negative association between HR and PPTh measured after the stressor (*r* = -.56, *p* = .02) (Kadetoff & Kosek, 2007).

***ANS: SBP, DBP, BRS, HRV during stressor – Pain*.** Across five studies, only three significant associations were found. One study reported a positive association between DBP measured during an arithmetic stressor and CPTo at baseline (*r* = .48, *p* < .05) (Reyes Del Paso, et al., 2022), one study reported a negative association between DBP measured during an isometric contraction and PPTh measured during the stressor (*r* = -.25, *p* = .05) (Kadetoff & Kosek, 2010), and one study reported a significant negative association between HR measured during an isometric contraction and PPTh measured after the stressor (*r* = -.56, *p* = .02) (Kadetoff & Kosek, 2007).

***ANS: EDA during stressor – Pain*.** One study measured sympathetic skin responses (SSR) during heat stimuli and found a significant positive association with heat pain ratings (*r*  = .50, *p* = .004) (Scheuren, et al., 2023).

***ANS: HR, SBP, DBP, BRS and HRV during recovery – Pain*.** Across six studies, only two significant associations were found in one study. This study reported a positive association between DBP measured during recovery and CPTh at baseline (*r* = .38, *p*  = .04), and a negative association between RRI during recovery and CPTo at baseline (*r* = .38, *p* = .04) (Reyes Del Paso, et al., 2022).

***HPA during stressor/recovery – Pain.*** Only one study evaluated cortisol levels during recovery in relation to PPTh and reported a positive association between cortisol during recovery and PPTh after the stressor (*r* = .55, *p* = .05) (Kadetoff & Kosek, 2010). Three studies investigated cortisol levels during a stressor and found no significant associations (all *p* > .05).

#### ***Change in stress – pain (Cluster 3a & 3b)***

***ANS reactivity: HR, SBP and DBP – pain.*** Across five studies, only one study reported significant positive associations between the change in HR (from baseline to during stressor) and EPTh and EPTo at baseline (*r* = .45-.47, *p* < .05) (Mohn, et al., 2008).

***ANS recovery: HR, SBP and DBP – pain.*** Across two studies, no significant associations were found with PPTh an PPTo (all *p* > .05) (Bandeira, et al., 2021; Lopez-Lopez, et al., 2021).

***HPA reactivity – Pain.*** In one study, no significant association was found between changes in cortisol (from baseline to immediately after stressor) with heat pain rating at baseline (all *p* < .05) (Meeus, et al., 2008).

***HPA recovery – Pain.*** No data available.

#### ***Stress – change in pain (Cluster 4)***

***ANS: HR, SBP, DBP and HRV – pain.*** SBP at baseline was negatively associated with change in PPTh (from baseline to during stressor) (*r* = .40, *p* = .02) (Lopez-Lopez, et al., 2021) and with change in EPTo (from baseline to after stressor) (*r* = .51, *p* = .03) (Loffler, et al., 2023) in two studies. Other associations in these studies were not significant. One other study did not yield significant associations (all *p* > .05) (Bandeira, et al., 2021).

#### ***Change in stress and pain (Cluster 5)***

Across four studies, no significant associations were found between reactivity and recovery of HR, SBP, DBP, HRV, PEP and with changes in PPTh, PPTo, EPTh, or EPTo (all *p* > .05).

#### ***Summary of results in pain-free controls***

**Qualitative analyses**

In pain-free controls, several significant associations between stress and pain were identified.
**At baseline**, higher SBP was positively associated with various static pain measures, including PPTh, ischemic PTh, PPTo, EPTo, ischemic PTo, and HPTo across three studies. Additionally, one study reported a positive association between DBP and PPTh. One study reported that lower cortisol measured at 13h, and lower cortisol AUCi was associated with higher HPTh and CPTh, respectively. Another study found that higher DHEA levels were associated with higher PPTh and PPTo.

**During stress exposure**, one study reported that lower heart rate was associated with higher PPTh, one study reported that higher DBP was associated with higher CPTo, one study reported that higher lower DBP was associated with higher PPTh, one study observed a positive association between SSR and heat pain ratings. **Following the stressor**, one study reported an association between higher DBP and higher CPTh, and an association between lower RRI and higher CPTo. Cortisol levels showed a significant positive association with PPTh.

When looking at **reactivity of the stress systems (i.e., change in stress marker)**, one study reported that a higher change in HR was associated with higher EPTh and EPTo.

Regarding associations between stress and **changes in pain**, higher SBP was linked to reduced changes in PPTh and EPTo, as evidenced by significant negative associations in two studies.

All other associations did not yield significance. All associations are based on very low to low certainty of evidence.

**Quantitative analyses**

Only one meta-analysis found an inverse association between HR measured during a stressor, and PPThs measured after a stressor (*r* = -.28; 95% CI = [- 0.49 ; - 0.033]).

### **References**

Bandeira, P. M., Reis, F. J. J., Muniz, F. D. N., Chaves, A. C. S., Fernandes, O., Jr., & Arruda-Sanchez, T. (2021). Heart Rate Variability and Pain Sensitivity in Chronic Low Back Pain Patients Exposed to Passive Viewing of Photographs of Daily Activities. *The Clinical Journal of Pain 37*, 591-97.

Bossenger, N. R., Lewis, G. N., Rice, D. A., & Shepherd, D. (2023). The autonomic and nociceptive response to acute exercise is impaired in people with knee osteoarthritis. *Neurobiology of Pain 13*, 100118.

Chalaye, P., Goffaux, P., Bourgault, P., Lafrenaye, S., Devroede, G., Watier, A., & Marchand, S. (2012). Comparing pain modulation and autonomic responses in fibromyalgia and irritable bowel syndrome patients. *The Clinical Journal of Pain 28*, 519-26.

Crettaz, B., Marziniak, M., Willeke, P., Young, P., Hellhammer, D., Stumpf, A., & Burgmer, M. (2013). Stress-induced allodynia--evidence of increased pain sensitivity in healthy humans and patients with chronic pain after experimentally induced psychosocial stress. *PLoS One, 8*, e69460.

Davydov, D. M., de la Coba, P., Contreras-Merino, A. M., & Reyes Del Paso, G. A. (2024). Impact of homeostatic body hydration status, evaluated by hemodynamic measures, on different pain sensitization paths to a chronic pain syndrome. *Scientific Reports 14*, 1908.

De Abreu Freitas, R. P., Lemos, T. M. A. M., Spyrides, M. H. C., & De Sousa, M. B. C. (2012). Influence of cortisol and DHEA-S on pain and other symptoms in post menopausal women with fibromyalgia. *Journal of Back and Musculoskeletal Rehabilitation, 25*, 245-52.

de la Coba, P., Bruehl, S., Duschek, S., & Reyes Del Paso, G. A. (2018). Blood pressure-related pain modulation in fibromyalgia: Differentiating between static versus dynamic pain indicators. *International Journal of Psychophysiology 134*, 79-85.

Farmer, A. D., Coen, S. J., Kano, M., Naqvi, H., Paine, P. A., Scott, S. M., Furlong, P. L., Lightman, S. L., Knowles, C. H., & Aziz, Q. (2014). Psychophysiological responses to visceral and somatic pain in functional chest pain identify clinically relevant pain clusters. *Neurogastroenterology & Motility 26*, 139-48.

Flor, H., Diers, M., & Birbaumer, N. (2004). Peripheral and electrocortical responses to painful and non-painful stimulation in chronic pain patients, tension headache patients and healthy controls. *Neuroscience letters, 361*, 147-50.

Galeazzi, F., Lucà, M. G., Lanaro, D., D'Incà, R., D'Odorico, A., Sturniolo, G. C., & Mastropaolo, G. (2001). Esophageal hyperalgesia in patients with ulcerative colitis:: Role of experimental stress. *American Journal of Gastroenterology, 96*, 2590-95.

Geiss, A., Rohleder, N., & Anton, F. (2012). Evidence for an association between an enhanced reactivity of interleukin-6 levels and reduced glucocorticoid sensitivity in patients with fibromyalgia. *Psychoneuroendocrinology, 37*, 671-84.

Granot, M., Friedman, M., Yarnitsky, D., & Zimmer, E. Z. (2002). Enhancement of the perception of systemic pain in women with vulvar vestibulitis. *Bjog-an International Journal of Obstetrics and Gynaecology, 109*, 863-66.

Jarrett, M. E., Han, C. J., Cain, K. C., Burr, R. L., Shulman, R. J., Barney, P. G., Naliboff, B. D., Zia, J., & Heitkemper, M. M. (2016). Relationships of abdominal pain, reports to visceral and temperature pain sensitivity, conditioned pain modulation, and heart rate variability in irritable bowel syndrome. *Neurogastroenterology & Motility 28*, 1094-103.

Jarrett, M. E., Shulman, R. J., Cain, K. C., Deechakawan, W., Smith, L. T., Richebé, P., Eugenio, M., & Heitkemper, M. M. (2014). Conditioned Pain Modulation in Women With Irritable Bowel Syndrome. *Biological Research for Nursing, 16*, 368-77.

Jonsson, K., Pikwer, A., Olsson, E. M., & Peterson, M. (2025). Altered breathing pattern and thoracic mobility in women with fibromyalgia: A case-control study. *The Journal of Pain*, 105508.

Kadetoff, D., & Kosek, E. (2007). The effects of static muscular contraction on blood pressure, heart rate, pain ratings and pressure pain thresholds in healthy individuals and patients with fibromyalgia. *European Journal of Pain, 11*, 39-47.

Kadetoff, D., & Kosek, E. (2010). Evidence of reduced sympatho-adrenal and hypothalamic-pituitary activity during static muscular work in patients with fibromyalgia. *Journal of Rehabilitation Medicine 42*, 765-72.

Larsson, B., Rosendal, L., Kristiansen, J., Sjogaard, G., Sogaard, K., Ghafouri, B., Abdiu, A., Kjaer, M., & Gerdle, B. (2008). Responses of algesic and metabolic substances to 8 h of repetitive manual work in myalgic human trapezius muscle. *Pain, 140*, 479-90.

Loffler, M., Schneider, P., Schuh-Hofer, S., Kamping, S., Usai, K., Treede, R. D., Nees, F., & Flor, H. (2023). Stress-induced hyperalgesia instead of analgesia in patients with chronic musculoskeletal pain. *Neurobiology of Pain 13*, 100110.

Lopez-Lopez, A., Matias-Pompa, B., Fernandez-Carnero, J., Gil-Martinez, A., Alonso-Fernandez, M., Alonso Perez, J. L., & Gonzalez Gutierrez, J. L. (2021). Blunted Pain Modulation Response to Induced Stress in Women with Fibromyalgia with and without Posttraumatic Stress Disorder Comorbidity: New Evidence of Hypo-Reactivity to Stress in Fibromyalgia? *Behavioral Medicine 47*, 311-23.

Maixner, W., Fillingim, R., Kincaid, S., Sigurdsson, A., & Harris, M. B. (1997). Relationship between pain sensitivity and resting arterial blood pressure in patients with painful temporomandibular disorders. *Psychosomatic medicine, 59*, 503-11.

Meeus, M., Nijs, J., Van de Wauwer, N., Toeback, L., & Truijen, S. (2008). Diffuse noxious inhibitory control is delayed in chronic fatigue syndrome: an experimental study. *Pain, 139*, 439-48.

Miyachi, R., Nishimura, T., Noguchi, M., Goda, A., Takeda, H., Takeshima, E., Kanazawa, Y., Imai, T., & Tanaka, W. (2025). Subgroup Characteristics of Middle-Aged and Older Women with Chronic Low Back Pain by Multiple Factors: A Hierarchical Cluster Analysis. *Journal of Functional Morphology and Kinesiology, 10*, 30.

Mohn, C., Vassend, O., & Knardahl, S. (2008). Experimental pain sensitivity in women with temporomandibular disorders and pain-free controls: The relationship to orofacial muscular contraction and cardiovascular responses. *The Clinical Journal of Pain 24*, 343-52.

Muhtz, C., Rodriguez-Raecke, R., Hinkelmann, K., Moeller-Bertram, T., Kiefer, F., Wiedemann, K., May, A., & Otte, C. (2013). Cortisol response to experimental pain in patients with chronic low back pain and patients with major depression. *Pain Medicine 14*, 498-503.

Murray, C. D. R., Flynn, J., Ratcliffe, L., Jacyna, M. R., Kamm, M. A., & Emmanuel, A. V. (2004). Effect of acute physical and psychological stress on gut autonomic innervation in irritable bowel syndrome. *Gastroenterology, 127*, 1695-703.

Nees, F., Loffler, M., Usai, K., & Flor, H. (2019). Hypothalamic-pituitary-adrenal axis feedback sensitivity in different states of back pain. *Psychoneuroendocrinology, 101*, 60-66.

Pardo, J. V., Larson, R. C., Spencer, R. J., Lee, J. T., Pasley, J. D., Torkelson, C. J., & Larson, A. A. (2019). Exposure to Cold Unmasks Potential Biomarkers of Fibromyalgia Syndrome Reflecting Insufficient Sympathetic Responses to Stress. *The Clinical Journal of Pain 35*, 407-19.

Poli-Neto, O. B., Oliveira, A. M. Z., Salata, M. C., Cesar Rosa, E. S. J., Machado, D. R. L., Candido-Dos-Reis, F. J., & Nogueira, A. A. (2020). Strength Exercise Has Different Effects on Pressure Pain Thresholds in Women with Endometriosis-Related Symptoms and Healthy Controls: A Quasi-experimental Study. *Pain Medicine 21*, 2280-87.

Quartana, P. J., Buenaver, L. F., Edwards, R. R., Klick, B., Haythornthwaite, J. A., & Smith, M. T. (2010). Pain catastrophizing and salivary cortisol responses to laboratory pain testing in temporomandibular disorder and healthy participants. *The Journal of Pain 11*, 186-94.

Rampazo, E. P., Rehder-Santos, P., de Andrade, A. L. M., Catai, A. M., & Liebano, R. E. (2024). Cardiac autonomic response to acute painful stimulus in individuals with chronic neck pain: A case-control study. *Musculoskeletal Science and Practice, 73*, 103141.

Reshkova, V., Kalinova, D., & Milanov, I. (2015). Evaluation of antiviral antibodies against Epstein-Barr Virus and neurotransmitters in patients with fibromyalgia. *J Neurol Neurosci, 6*, 35.

Reyes Del Paso, G. A., Contreras-Merino, A. M., & Duschek, S. (2022). The Role of Depressive Disorders in Autonomic Cardiovascular Dysregulation in Fibromyalgia. *Psychosomatic Medicine 84*, 793-802.

Reyes del Paso, G. A., Garrido, S., Pulgar, A., & Duschek, S. (2011). Autonomic cardiovascular control and responses to experimental pain stimulation in fibromyalgia syndrome. *Journal of Psychosomatic Research 70*, 125-34.

Scheuren, P. S., De Schoenmacker, I., Rosner, J., Brunner, F., Curt, A., & Hubli, M. (2023). Pain-autonomic measures reveal nociceptive sensitization in complex regional pain syndrome. *European Journal of Pain 27*, 72-85.

Tan, H., Tumilty, S., Chapple, C., Liu, L., Othman, R., & Baxter, G. D. (2023). Acupoints sensitization in people with and without chronic low back pain: a matched-sample cross-sectional study. *Journal of Back and Musculoskeletal Rehabilitation, 36*, 137-46.

Thieme, K., Jung, K., Mathys, M. G., Gracely, R. H., & Turk, D. C. (2022). Cardiac-Gated Neuromodulation Increased Baroreflex Sensitivity and Reduced Pain Sensitivity in Female Fibromyalgia Patients. *Journal of Clinical Medicine, 11*.

Valera-Calero, J. A., & Varol, U. (2022). Correlation among Routinary Physical Activity, Salivary Cortisol, and Chronic Neck Pain Severity in Office Workers: A Cross-Sectional Study. *Biomedicines, 10*.

Van Den Houte, M., Van Oudenhove, L., Van Diest, I., Bogaerts, K., Persoons, P., De Bie, J., & Van den Bergh, O. (2018). Negative Affectivity, Depression, and Resting Heart Rate Variability (HRV) as Possible Moderators of Endogenous Pain Modulation in Functional Somatic Syndromes. *Frontiers in Psychology 9*, 275.

Venezia, A., Fawsitt‐Jones, H., Hohenschurz‐Schmidt, D., Mancini, M., Howard, M., & Makovac, E. (2024). Investigating the effects of artificial baroreflex stimulation on pain perception: A comparative study in no‐pain and chronic low back pain individuals. *The Journal of Physiology, 602*, 6941-57.

Wingenfeld, K., Nutzinger, D., Kauth, J., Hellhammer, D. H., & Lautenbacher, S. (2010). Salivary Cortisol Release and Hypothalamic Pituitary Adrenal Axis Feedback Sensitivity in Fibromyalgia Is Associated With Depression But Not With Pain. *The Journal of Pain, 11*, 1195-202.

Woda, A., L'Heveder, G., Ouchchane, L., & Bodéré, C. (2013). Effect of experimental stress in 2 different pain conditions affecting the facial muscles. *The Journal of Pain 14*, 455-66.
